# Aptamer Proteomics for Biomarker Discovery in Heart Failure with Reduced Ejection Fraction

**DOI:** 10.1101/2022.07.27.22276826

**Authors:** Luqing Zhang, Jonathan W. Cunningham, Brian L. Claggett, Jaison Jacob, Mike Mendelson, Pablo Serrano-Fernandez, Sergio Kaiser, Denise Yates, Margaret Healey, Chien-Wei Chen, Gordon Turner, Natasha Patel-Murray, Faye Zhao, Michael T. Beste, Jason M. Laramie, William T. Abraham, Pardeep S. Jhund, Lars Kober, Milton Packer, Jean Rouleau, Michael R. Zile, Margaret F. Prescott, Martin Lefkowitz, John J.V. McMurray, Scott D. Solomon, William Chutkow

## Abstract

**Background:** Systematically characterizing associations between circulating proteins and risk for subsequent clinical events may improve clinical risk prediction and shed light on unrecognized biological pathways in heart failure (HF). Large-scale assays measuring thousands of proteins now enable broad proteomic investigation in clinical trials.

**Methods:** Serum levels of 4076 proteins were measured at baseline in the ATMOSPHERE (n=1258, 487 events over 6 years) and PARADIGM-HF (n=1257, 287 events over 4 years) trials of chronic HF with reduced ejection fraction using a modified aptamer-based proteomics assay. Proteins associated with the primary endpoint of HF hospitalization or cardiovascular death were identified in the ATMOSPHERE discovery cohort by Cox regression adjusted for age, sex, treatment arm, and anticoagulant use (false discovery rate<0.05), and were replicated in PARADIGM-HF (Bonferroni-corrected p<0.05). A proteomic risk score was derived in ATMOSPHERE using Cox LASSO penalized regression and evaluated in PARADIGM-HF compared to the MAGGIC clinical risk score and N-terminal pro-B-type natriuretic peptide (NT-proBNP) immunoassay. For proteins that were associated with the primary endpoint, two-sample Mendelian randomization was performed using genetic and outcome data from both trials and protein quantitative trait loci from deCODE to infer causal associations.

**Results:** We identified 377 serum proteins that were associated with the primary endpoint in ATMOSPHERE and replicated 167 in PARADIGM-HF. Prognostic proteins included known HF biomarkers such as Growth Differentiation Factor 15, NT-BNP, and Angiopoietin-2, and also a previously unrecognized HF biomarker: Sushi, Von Willebrand Factor Type A, EGF and Pentraxin Domain Containing 1 (SVEP1, HR 1.60 [95% CI 1.44-1.79] per standard deviation [SD], p=2×10^−17^). A 64-protein risk score derived in ATMOSPHERE predicted the primary endpoint in PARADIGM-HF with greater discrimination (C-statistic 0.70) than the MAGGIC clinical score (C-statistic 0.61), NT-proBNP (C-statistic 0.65), or both (C-statistic 0.66). Genetically controlled levels of BNP, WISP2, FSTL1, and CTSS were associated with the primary endpoint by Mendelian randomization.

**Conclusions:** We identified SVEP1, an extracellular matrix protein known to cause inflammation in vascular smooth muscle cells, as a new HF biomarker associated with risk of hospitalization or death. A 64-protein score improved risk discrimination compared with NT-proBNP and may assist in identifying high-risk patients.

## Introduction

Despite advances in prevention and treatment, heart failure with reduced ejection fraction remains a common condition with high morbidity and mortality.^1,2^ Circulating natriuretic peptide levels have improved heart failure diagnosis and risk prediction and are now incorporated into the definition of heart failure and inclusion criteria for most clinical trials.^3,4^ Prior research has identified other biomarkers—including soluble ST2, growth differentiation factor 15 (GDF15), and collagen processing proteins—which are associated with heart failure severity and suggest key disease mechanisms.^5–7^ Broader proteomic investigation provides an opportunity to evaluate unrecognized pathways for disease progression and novel treatment targets as well as to improve prognostic scores for trial enrichment and clinical management.^8,9^ Large-scale assays measuring thousands of proteins from a single sample now enable this approach.^10,11^

We performed discovery proteomics to identify serum proteins associated with risk for subsequent heart failure events in two multi-center randomized trials in patients with chronic heart failure with reduced ejection fraction.^12,13^ First, we measured serum levels of 4076 unique proteins at baseline in over 2500 patients using the SomaScan assay and identified proteins and protein pathways that were associated with risk for the primary endpoint, a composite of cardiovascular death or heart failure hospitalization. ATMOSPHERE was used for discovery and PARADIGM-HF for external validation. Second, we derived a proteomic risk score in ATMOSPHERE and externally validated its risk discrimination in PARADIGM-HF. Third, for those proteins that were associated with the primary endpoint, we leveraged genetic data from the two trials and performed two-sample Mendelian randomization to infer which proteins may causally contribute to heart failure progression.

## Methods

### Patient Population

The design and primary results of the ATMOSPHERE and PARADIGM-HF trials have been reported previously.^12–15^ The ATMOSPHERE trial randomized 7064 patients with heart failure, left ventricular ejection fraction ≤ 35%, and elevated plasma B-type natriuretic peptide concentration to either enalapril, aliskiren, or both. The PARADIGM-HF trial randomized a similar population of 8442 patients to either sacubitril/valsartan or enalapril. The primary endpoint of both studies was a composite of time to cardiovascular death or first heart failure hospitalization. Endpoints were adjudicated by a central clinical events committee in both trials.

### Measurements of Circulating Proteins

Serum samples for proteomic assays were available at baseline for 1258 patients in ATMOSPHERE and 1257 patients in PARADIGM-HF. Baseline was defined as during the enalapril run-in in ATMOSPHERE and prior to the run-in period in PARADIGM-HF. Samples were stored centrally at -80°C. The SomaScan version 3 proteomics assay was performed to quantify levels of 5034 SOMAmers (slow off-rate modified aptamers) targeting circulating proteins in serum samples at baseline in both trials; since some proteins are measured by multiple aptamers, 4076 unique proteins were measured.^9,10,16^ **Supplemental File 2** lists the aptamers measured and their target proteins. Modified aptamers are DNA oligonucleotides that have been selected to bind target proteins with high affinity and slow dissociation rates.^17^ The specificity of this assay for the correct target protein has been confirmed by mass spectrometry for approximately 1000 aptamers ^8^ and by the discovery of *cis* genetic variants associated with protein levels for additional aptamers.^8,18^ Median intra-assay and inter-assay coefficients of variation are <6%.^19^ We applied a global median normalization to correct for technical batch effects between plates.

### Identifying Protein Levels Predicting Risk for Heart Failure Events

**Figure 1** outlines the analysis plan. Associations between log-transformed and standardized individual protein levels and risk for the primary outcome (time to cardiovascular death or first heart failure hospitalization) were assessed by Cox regression. Three key secondary outcomes endpoints (cardiovascular death, heart failure hospitalization alone, and all cause death) were also investigated. Two covariate models were used: a minimally adjusted model including age, sex, treatment, and anticoagulant usage (based on our observation that anticoagulants affect detection of protein levels in serum), and a second model further adjusted for the following covariates known to predict risk for heart failure events: prior myocardial infarction, diabetes mellitus, atrial fibrillation, current smoking, prior heart failure hospitalization, time since heart failure diagnosis, systolic blood pressure, body mass index, low-density lipoprotein level, high-density lipoprotein level, estimated glomerular filtration rate, New York Heart Association functional class, and left ventricular ejection fraction. Hazard ratios were expressed per 1 standard deviation (SD) increase in log-transformed batch-adjusted protein level. Proteins associated with clinical events meeting a Benjamini-Hochberg false discovery rate (FDR) <0.05 in ATMOSPHERE were validated in PARADIGM-HF using Bonferroni correction for the number of proteins carried forward, to rigorously exclude false-positive associations due to multiple hypothesis testing. Two sensitivity analyses were performed. First, hazard ratios were expressed per doubling of protein levels rather than per standard deviation given possible differences in standard deviations between the two trials. Second, rank-based inverse normal transformation was applied to the protein levels to minimize the effect of outliers or skewed distributions of protein levels. QIAGEN Ingenuity Pathway Analysis was used to identify groups of biologically related proteins that were over-represented in the set of proteins associated with the primary endpoint in the two trials combined. Analyses were conducted in R v.4.0.

**Figure 1.**
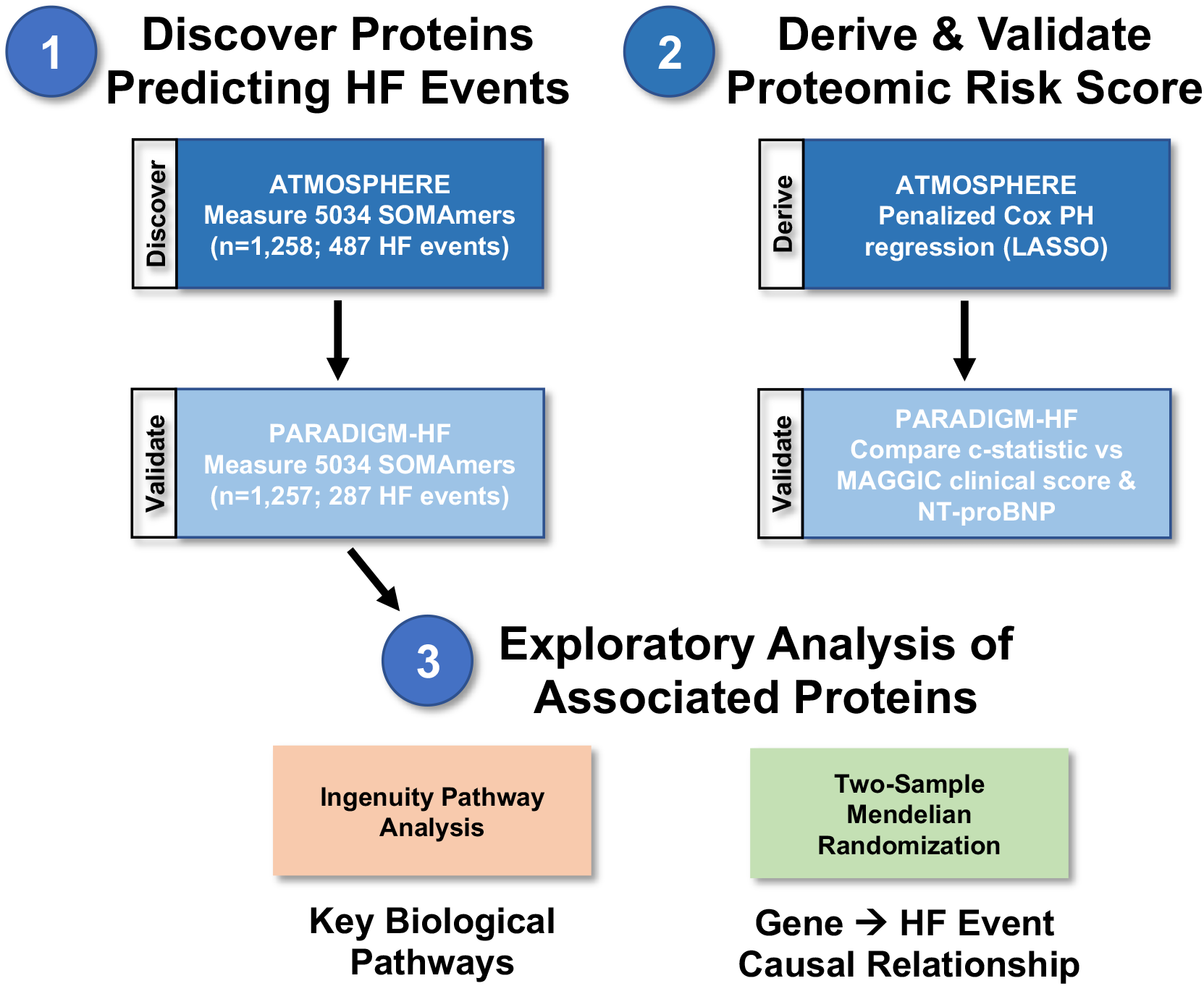
Schematic analysis overview.

### Derivation and Validation of a Proteomic Score Predicting Heart Failure Events

We developed a proteomic score for the primary endpoint and validated it in an independent cohort. In the ATMOSPHERE derivation cohort, penalized Cox LASSO regression was used to select variables and derive the proteomic risk score weights. The lambda parameter was optimized as the value with minimum mean cross-validated error in ATMOSPHERE. In PARADIGM-HF, we validated the proteomic risk score by comparing it to the Meta-analysis Global Group in Chronic Heart Failure (MAGGIC) risk score, a validated clinical risk score for mortality in patients with heart failure,^20^ NT-proBNP measured by traditional immunoassay, and linear combinations of MAGGIC and NT-proBNP, or MAGGIC and the proteomic score with coefficients derived strictly in the derivation set. Risk discrimination was measured using Harrell’s C-statistic. 95% confidence intervals for C-statistics and p-values for differences in C-statistics were estimated by the Somer’s D method using the somersd package in STATA version 16 and with bootstrapping.

### Two-Sample Mendelian Randomization

For proteins whose levels associated with the primary endpoint, we performed exploratory two sample Mendelian randomization analysis to assess their potential causal role in patient prognosis. First, protein quantitative trait loci (pQTLs) were defined in the deCODE cohort study, in which 4907 plasma proteins were measured in 33,559 Icelanders using the SomaScan assay.^21^ Second, associations between genetic variants and heart failure outcomes were assessed in a pooled sample of 3237 patients from PARADIGM-HF (n=1374) and ATMOSPHERE (n=1863) with available common variant genetic data, who experienced a total of 942 primary endpoint events. Samples with excess heterozygosity, sex mismatch, or cryptic relatedness were excluded. Imputation was performed with SHAPEIT2-IMPUTE2 using 1000 Genomes reference panel.^22,23^ Considering the small sample size of the outcome study, only high confidence common variants with minor allele frequency > 10% and imputation INFO score > 0.5 were included (a total of 4,863,989 SNPs). We performed a genome wide association study using Cox regression to identify associations between each SNP and the primary end point, adjusted for treatment, age, sex, trial and the first 10 principal components of ancestry.

We then performed two-sample Mendelian randomization using the pQTLs from the deCODE cohort and outcome data from the combined ATMOSPHERE-PARADIGM-HF cohort. We tested only proteins whose measured levels were significantly associated with the primary endpoint to increase the likelihood of biologically plausible findings. Proteins with significant *cis*-pQTLs in deCODE (p-value < 5 ×10^−8^ and F statistic > 10) were identified; SNPs within 10MB of the gene region were clumped by linkage disequilibrium (LD) coefficient r^2^<0.1; a sensitivity analysis using r^2^<0.01 was also performed. The R package TwoSampleMR was used to calculate two-sample Mendelian randomization results using IVW (inverse variance weighted) for proteins with multiple pQTLs or Wald ratio for proteins with a single pQTL.^24^ Horizontal pleiotropy was tested with MR-PRESSO.^25^

## Results

### Baseline Characteristics and Outcomes

Baseline characteristics of patients in the ATMOSPHERE and PARADIGM-HF trials with proteomic data are shown in **Table 1** and are compared to the full trial populations in **Supplemental Table 1**. ATMOSPHERE patients with available proteomic data (n=1,258) were older (median 67 years vs 64 years), less likely to be enrolled in Asia (0.4% vs 31%), and more likely to be White (96% vs 59%) and use an anticoagulant (39% vs 29%) than the fully enrolled ATMOSPHERE cohort. Ejection fraction (median 30% for both) and eGFR (median 72 vs 74 ml/min/1.73m^2^) were similar in the two groups. The rate of the primary endpoint, heart failure hospitalization or cardiovascular death, was similar in the two groups (11.5 events per 100 patient-years in those with proteomic data compared with 11.8 in those without, p=0.46). The replication cohort, patients with proteomic data in the PARADIGM-HF trial, had similar baseline characteristics to ATMOSPHERE.

**Table 1:** Baseline Characteristics of Patients with Available Proteomic Data.

| **Characteristic** | **ATMOSPHERE**<br>N = 1,258 | **PARADIGM-HF**<br>N = 1,257 |
| --- | --- | --- |
| **Age (years)** | 67 (59, 73) | 68 (61, 74) |
| **Female** | 240 (19%) | 233 (19%) |
| **Race** |  |  |
| Asian | 5 (0.4%) | 4 (0.3%) |
| Black | 12 (1.0%) | 14 (1.1%) |
| White | 1,205 (96%) | 1,213 (96%) |
| Other | 35 (2.8%) | 26 (2.1%) |
| Pacific Islander | 1 (<0.1%) | 0 (0%) |
| **Region** |  |  |
| Asia/Pacific and Other | 20 (1.6%) | 0 (0%) |
| Central/Eastern Europe | 506 (40%) | 584 (46%) |
| Latin America (including Central America) | 57 (4.5%) | 0 (0%) |
| North America | 54 (4.3%) | 56 (4.5%) |
| Western Europe | 621 (49%) | 617 (49%) |
| **Diabetes mellitus** | 389 (31%) | 476 (38%) |
| **Hypertension** | 893 (71%) | 963 (77%) |
| **Prior myocardial infarction** | 656 (52%) | 582 (46%) |
| **Ischemic cardiomyopathy** | 814 (65%) | 813 (65%) |
| **Prior stroke** | 97 (7.7%) | 137 (11%) |
| **Atrial fibrillation** | 518 (41%) | 606 (48%) |
| **NYHA functional class** |  |  |
| I / II | 789 (63%) | 857 (68%) |
| III / IV | 469 (37%) | 400 (32%) |
| **Left Ventricular Ejection fraction (%)** | 30.0 (25.0, 34.0) | 31.5 (27.5, 35.0) |
| **Medications at Baseline** |  |  |
| Anticoagulant | 493 (39%) | 560 (45%) |
| ACEi | 1,257 (100%) | 993 (79%) |
| ARBi | 35 (2.8%) | 266 (21%) |
| Diuretic | 1,028 (82%) | 1,058 (84%) |
| Digoxin | 312 (25%) | 280 (22%) |
| Beta-blocker | 1,187 (94%) | 1,201 (96%) |
| **Cardiac resynchronization therapy** | 107 (8.5%) | 115 (9.1%) |
| **Implantable cardioverter defibrillator** | 317 (25%) | 312 (25%) |
| **Body mass index (kg/m<sup>2</sup>)** | 28.1 (25.2, 31.6) | 28.5 (25.4, 32.1) |
| **Systolic blood pressure (mmHg)** | 130 (118, 140) | 130 (120, 142) |
| **eGFR (mL/min/1.73m<sup>2</sup>)** | 72 (59, 86) | 64 (53, 77) |
| **NT-proBNP (pg/mL)** | 1,342 (760, 2,390) | 1,492 (853, 2,935) |
| **NT-proBNP unknown** | 150 | 0 |

Values are n (%) for categorical variables or median (interquartile range) for continuous variables.
ACEi, angiotensin-converting enzyme inhibitor; ARB, angiotensin-receptor blocker; eGFR, estimated glomerular filtration rate; NT-proBNP, N-terminal pro-B-type natriuretic peptide; NYHA, New York Heart Association

### Individual Proteins Predicting Risk for Heart Failure Events

We identified 377 proteins significantly associated with the risk of heart failure hospitalization or cardiovascular death in ATMOSPHERE at a significance threshold of FDR <0.05 adjusting for age, sex, treatment, and anticoagulant use. Of these, 167 replicated in PARADIGM-HF after Bonferroni correction for the number of proteins carried forward (**Figure 2a, Table 2 and Supplemental Table 2**), and 302 replicated with at least nominal p-value <0.05 in the same direction. Biomarkers known to predict risk of heart failure hospitalization and cardiovascular death as well as and novel biomarkers were strongly predictive of the primary endpoint in both trials. The proteins most significantly associated with the primary endpoint included growth differentiation factor 15 (GDF15; replication cohort HR 1.66 [1.47-1.88] per standard deviation[SD], p=3.4×10^−16^), Angiopoietin 2 (ANGPT2; HR 1.73 [1.54-1.95] per SD, p=6.8×10^−20^), NT-proBNP (HR 1.63 [95% CI 1.44-1.86] per SD, p=7.2 × 10^−14^), thrombospondin 2 (THBS2; HR 1.66 [95% 1.49-1.85] per SD, p=3.4×10^−20^), and a novel heart failure biomarker: Sushi, Von Willebrand Factor Type A, EGF And Pentraxin Domain Containing 1 (SVEP1; HR 1.60 [95% CI 1.44-1.79] per SD, p=2.0 × 10^−17^). Sensitivity analyses in which hazard ratios were expressed per doubling of protein level (i.e. per unit increase after log2-transformation), or after rank-based inverse normal transformation of the protein levels to minimize outlier effects, demonstrated consistent results (**Supplemental Tables 3** and **4**). After adjustment for additional cardiovascular risk factors, 64 proteins remained significantly associated with the primary endpoint in both trials after the same adjustment for multiple hypothesis testing (**Figure 2b and Supplemental Table 5)**. Associations between protein levels and all-cause death, cardiovascular death, and heart failure hospitalization, are shown in **Supplemental Tables 6-11**.

**Figure 2:**
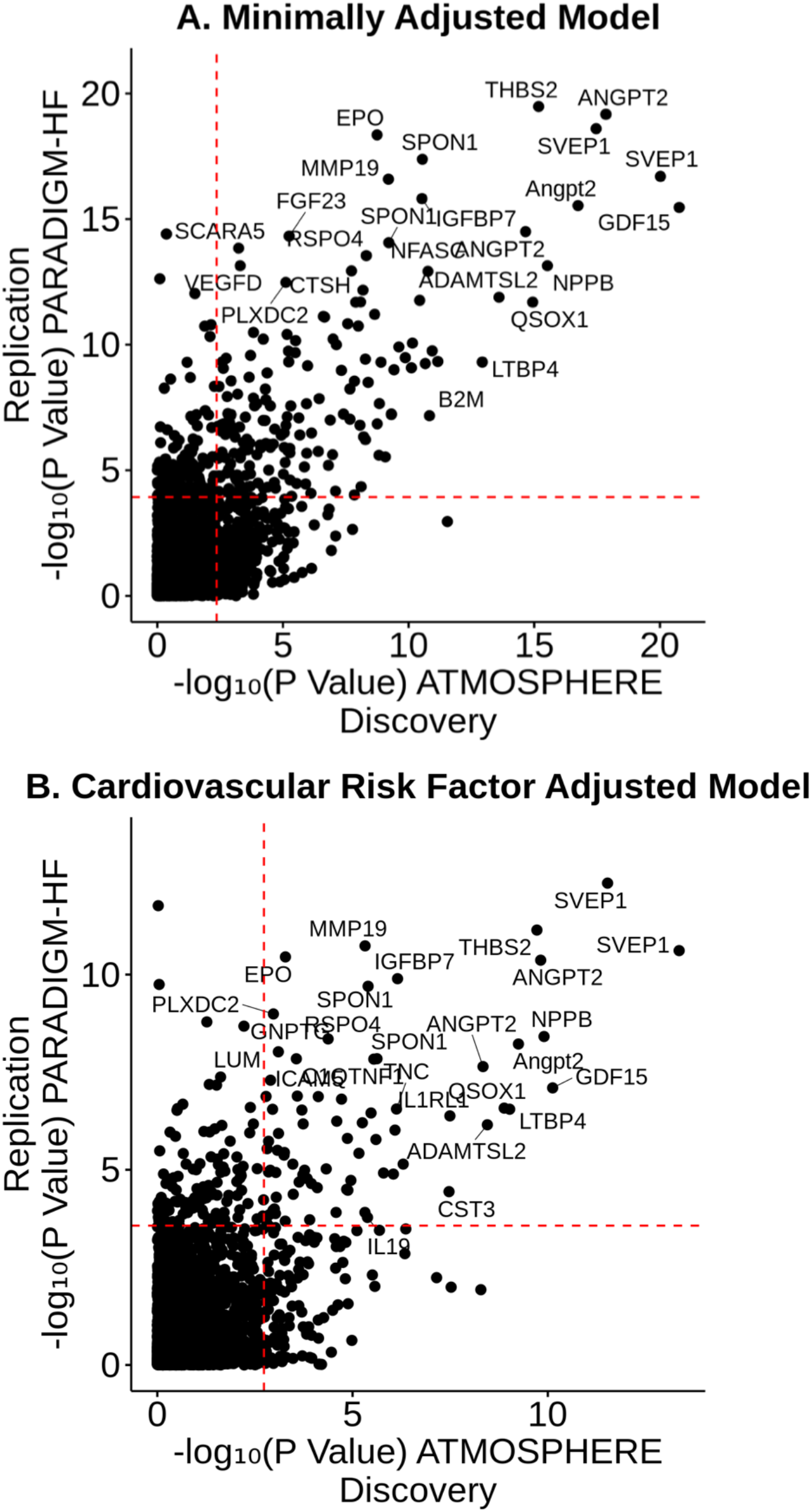
Protein Levels Associated with Risk for Heart Failure Hospitalization or Cardiovascular Death. Circulating proteins significantly associated with the time to first heart failure hospitalization or cardiovascular death. Associations were considered significant if the p-value was lower than false discovery rate threshold in ATMOSPHERE and Bonferroni-corrected threshold in PARADIGM-HF. Panel A: Minimally adjusted model includes age, sex, treatment and anticoagulant use as covariates. Panel B: Cardiovascular risk factor adjusted model is further adjusted for prior myocardial infarction, body mass index, diabetes mellitus, current smoking, time since heart failure diagnosis, prior heart failure hospitalization, systolic blood pressure, low-density and high-density lipoprotein concentrations, estimated glomerular filtration rate, atrial fibrillation, New York Heart Association functional class and left ventricular ejection fraction.

**Table 2:** Top 15 Proteins Most Strongly Associated with Risk of Heart Failure Hospitalization or Cardiovascular Death.

| Full protein name | Discovery (ATMOSPHERE) |  | Replication (PARADIGM) |  |
| --- | --- | --- | --- | --- |
|  | HR per SD (95% CI) | P Value | HR per SD (95% CI) | P Value |
| Growth/differentiation factor 15 (GDF15) | 1.50 (1.38-1.64) | $1.69 \times 10^{-21}$ | 1.66 (1.47-1.88) | $3.43 \times 10^{-16}$ |
| Sushi, von Willebrand factor type A, EGF and pentraxin domain-containing protein 1 (SVEP1) | 1.46 (1.35-1.58) | $9.42 \times 10^{-21}$ | 1.60 (1.44-1.79) | $2.01 \times 10^{-17}$ |
| Angiopoietin-2 (ANGPT2) | 1.48 (1.35-1.61) | $1.40 \times 10^{-18}$ | 1.73 (1.54-1.95) | $6.84 \times 10^{-20}$ |
| N-Terminal B-type Natriuretic Peptide (NT-proBNP) | 1.56 (1.40-1.73) | $2.97 \times 10^{-16}$ | 1.63 (1.44-1.86) | $7.17 \times 10^{-14}$ |
| Thrombospondin-2 (THBS2) | 1.42 (1.30-1.54) | $6.66 \times 10^{-16}$ | 1.66 (1.49-1.85) | $3.35 \times 10^{-20}$ |
| Sulfhydryl oxidase 1 (QSOX1) | 1.42 (1.31-1.55) | $1.14 \times 10^{-15}$ | 1.46 (1.31-1.62) | $2.05 \times 10^{-12}$ |
| ADAMTS-like protein 2 (ADAMTSL2) | 1.40 (1.28-1.52) | $2.51 \times 10^{-14}$ | 1.48 (1.33-1.65) | $1.31 \times 10^{-12}$ |
| Latent-transforming growth factor beta-binding protein 4 (LTBP4) | 1.43 (1.30-1.57) | $1.16 \times 10^{-13}$ | 1.35 (1.23-1.48) | $4.96 \times 10^{-10}$ |
| Cystatin-C (CST3) | 1.37 (1.25-1.50) | $6.85 \times 10^{-12}$ | 1.52 (1.33-1.73) | $4.69 \times 10^{-10}$ |
| Spondin-2 (SPON2) | 1.38 (1.26-1.52) | $1.14 \times 10^{-11}$ | 1.54 (1.35-1.76) | $1.77 \times 10^{-10}$ |
| Beta-2-microglobulin (B2M) | 1.37 (1.25-1.51) | $1.47 \times 10^{-11}$ | 1.45 (1.26-1.65) | $6.70 \times 10^{-8}$ |
| Neurofascin (NFASC) | 1.33 (1.22-1.44) | $1.70 \times 10^{-11}$ | 1.54 (1.38-1.73) | $1.21 \times 10^{-13}$ |
| Interleukin-1 receptor-like 1 (IL1RL1) | 1.39 (1.26-1.53) | $2.16 \times 10^{-11}$ | 1.48 (1.31-1.67) | $5.67 \times 10^{-10}$ |
| Spondin-1 (SPON1) | 1.33 (1.23-1.45) | $2.82 \times 10^{-11}$ | 1.52 (1.39-1.68) | $4.17 \times 10^{-18}$ |
| Insulin-like growth factor-binding protein 7 (IGFBP7) | 1.35 (1.24-1.48) | $2.93 \times 10^{-11}$ | 1.58 (1.41-1.76) | $1.53 \times 10^{-16}$ |

SVEP1 was among the strongest prognostic proteins in both trials. Target specific binding is supported by the measurement of 2 different aptamers for SVEP1 whose levels were highly correlated (r=0.976, p<2.2×10^−16^), and the presence of a strong SVEP1 *cis*-pQTL in deCODE (p<10^−250^) (**Supplemental Figure 1**). Baseline characteristics of patients by quartile of SVEP1 level are shown in **Supplemental Table 12**. Higher SVEP1 levels were associated with older age (median 71 years in highest quartile [Q4] vs 62 years in the lowest quartile [Q1]), greater prevalent atrial fibrillation (65% in Q4 vs 25% in Q1), and higher NT-proBNP levels measured by immunoassay (median 2823 pg/ml in Q4 vs 926 pg/ml in Q1). SVEP1 levels did not differ by sex, ejection fraction, or ischemic etiology of heart failure. SVEP1 was also among the most strongly prognostic proteins after adjustment for cardiovascular risk factors (validation cohort HR 1.49 [95% CI 1.32-1.67] per SD, p=2.4×10^−11^). SVEP1 remained associated with the primary endpoint after adjustment for NT-proBNP along with age, sex, treatment, and anti-coagulant use (HR 1.33 [95% CI 1.17-1.51] per SD, p=1.2×10^−5^).

Ingenuity Pathways Analysis identified 18 biological pathways significantly enriched with proteins that were associated with the primary endpoint after minimal adjustment (**Figure 3 and Supplemental Table 13**). Three of these were statistically significant at false discovery rate of <0.05: hepatic fibrosis and stellate cell activation (p=5.9×10^−5^), and agranulocyte (p=1.8×10^−4^) and granulocyte (p=1.9×10^−4^) adhesion and diapedesis.

**Figure 3:**
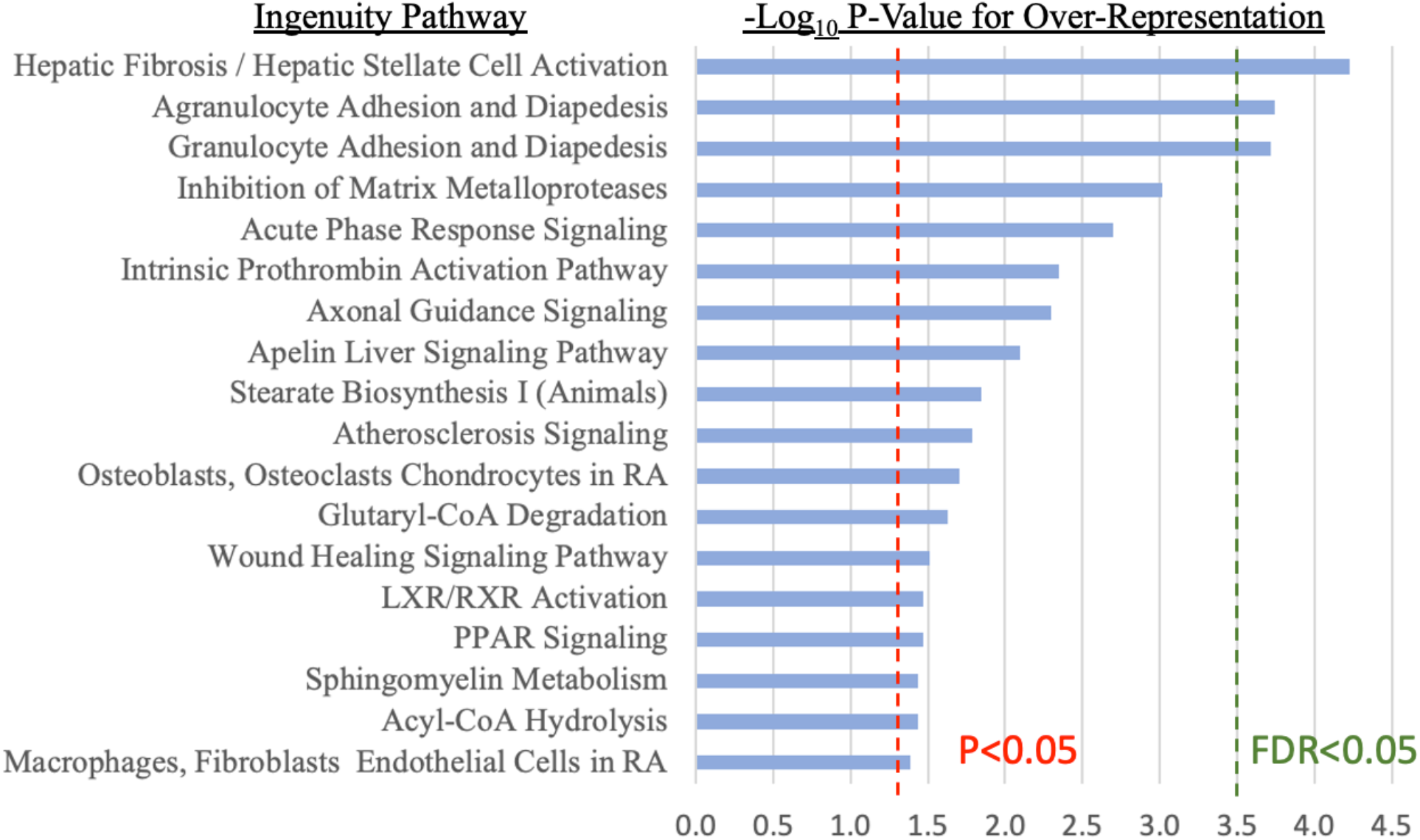
Pathways Significantly Associated with Risk for Heart Failure Hospitalization or Cardiovascular Death. Eighteen Ingenuity Canonical Pathways were significantly over-represented among proteins associated with risk for heart failure hospitalization or cardiovascular death. Red line indicates nominal p-value of 0.05. The top three pathways were statistically significant at a threshold of false discovery rate <0.05 (green line).

### Proteomic Risk Score

Using ATMOSPHERE as a derivation set, we developed a 64-protein weighted score reflecting risk for the primary endpoint using Cox LASSO regression. Beta coefficients for this score and the correlation between included proteins are shown in **Supplemental Figure 2 and Supplemental Table 14**. The proteins with the highest weights were NT-proBNP, basal cell adhesion molecule, cystatin C, and GDF15. NT-proBNP measurements by ELISA and aptamer-based methods were correlated (r=0.72, **Supplemental Figure 3**). In the PARADIGM-HF external validation set, the proteomic score (C-statistic 0.70 [95% CI 0.67-0.73]) provided superior discrimination compared with MAGGIC risk score (C-statistic 0.61 [95% CI 0.57-0.64], p<0.001 vs proteomic score) and NT-proBNP measured by immunoassay (C-statistic 0.65 [95% CI 0.62-0.69], p=0.001 vs proteomic score) (**Figure 4**). A linear combination of the MAGGIC score and proteomic score was superior to a linear combination of NT-proBNP and the MAGGIC score (C-statistic 0.70 [95% CI 0.67-0.73] vs 0.66 [95% CI 0.63-0.69], p=0.002). The addition of the MAGGIC risk score to the proteomic score did not improve the C-statistic (C-statistic 0.70 [95% CI 0.67-0.73] for both, p=0.31). C-statistic confidence intervals determined by bootstrapping provided similar results (**Supplemental Figure 4)**. Results were similar when follow-up was truncated at one or two years (**Supplemental Table 15**). The event rate in the highest quartile of the proteomic risk score was 6-fold greater than in the lowest quartile, compared with 2.6-fold greater for the MAGGIC risk score and 4.1-fold greater for the NT-proBNP immunoassay (**Figure 5**).

**Figure 4:**
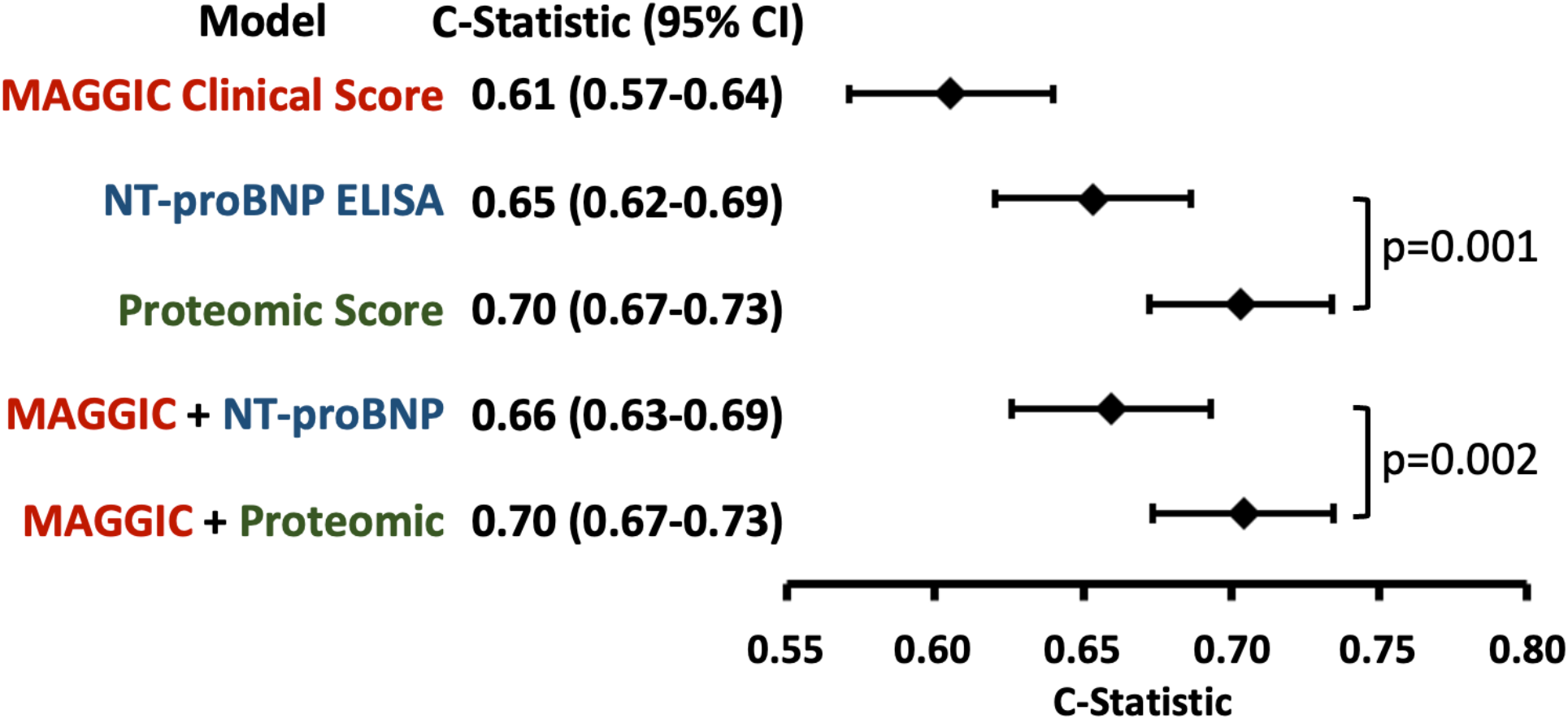
Proteomic Risk Score Discrimination Compared with Current Clinical Standards. Proteomic risk score derived in ATMOSPHERE provided superior risk discrimination compared with NT-proBNP and MAGGIC clinical risk score in the PARADIGM-HF validation cohort. Weights for combination models were derived strictly in ATMOSPHERE. C-statistics for each model and linear combinations of models. P-values for the difference in C-statistic between risk scores were calculated by the Somer’s D method.

**Figure 5:**
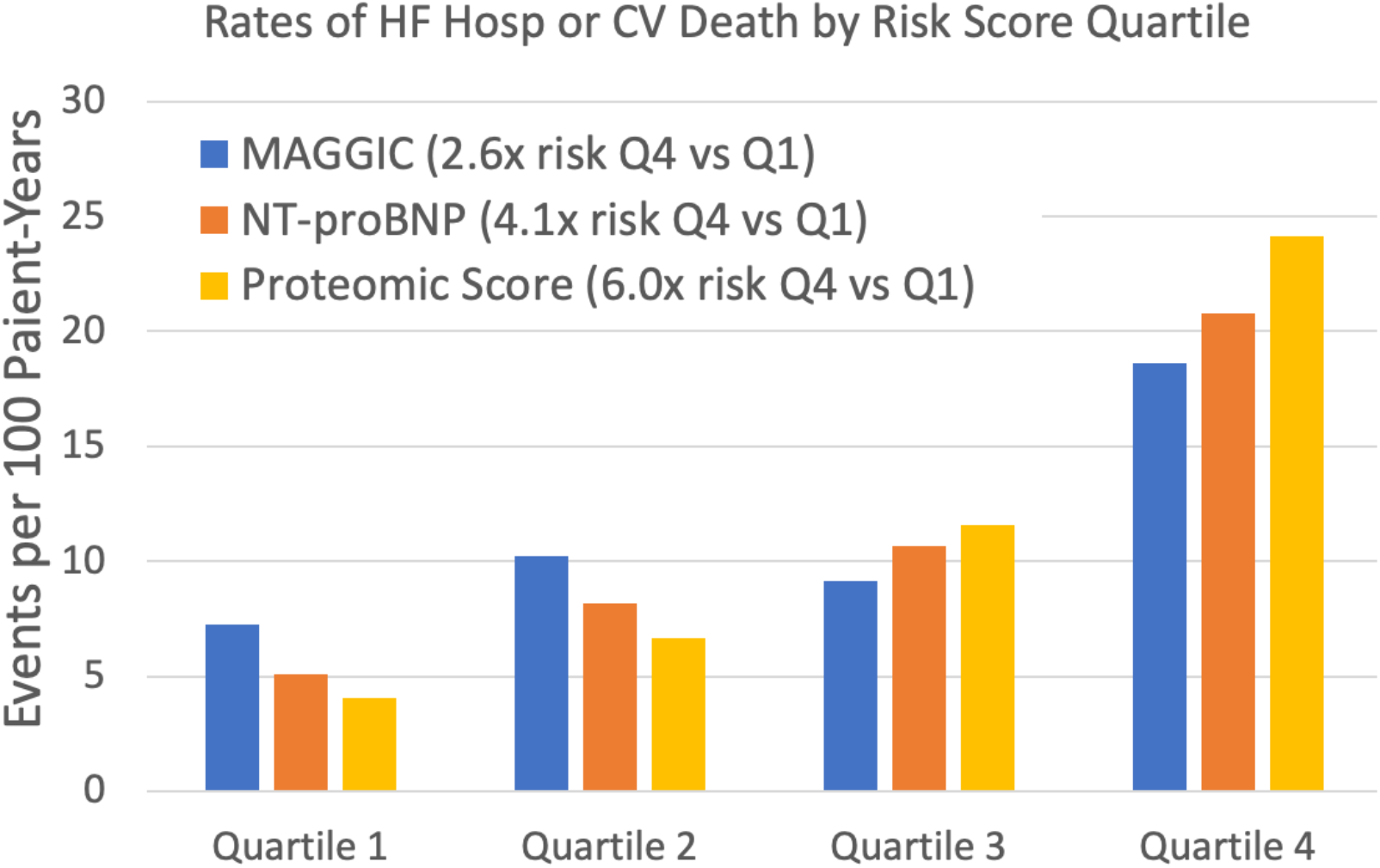
Primary Endpoint Event Rates by Risk Score Quartile in PARADIGM-HF Validation Cohort. In the PARADIGM-HF validation cohort, 1219 patients with complete data were divided into quartiles according to MAGGIC clinical risk score, NT-proBNP measured simultaneously to proteomic assay, or the proteomic risk score derived in ATMOSPHERE. The relative risk difference between the first and fourth quartiles, defined as observed event rate in the highest quartile divided by observed event rate in the lowest quartile, was greatest for the proteomic score.

### Mendelian Randomization

An exploratory Mendelian randomization analysis restricted to proteins whose levels were associated with the primary endpoint identified four genes with suggestive evidence as causally promoting heart failure progression. Significant *cis*-pQTLs were available for 506 of the 779 proteins whose levels were associated with the primary endpoint in combined ATMOSPHERE and PARADIGM-HF cohort. Genetically controlled lower levels of BNP (HR 0.23 per SD, p=8.5×10^−5^) and Cathepsin S (CTSS; HR 0.73 per SD, p=1.1×10^−4^), and genetically controlled higher levels of WNT1 Inducible Signaling Pathway Protein 2 (WISP2, also known as Cellular Communication Network Factor 5 or CCN5; HR 1.44 per SD, p=1.0×10^−4^) and Follistatin-Like 1 (FSTL1; HR 2.4 per SD, p=3.5×10^−4^) were significantly associated with risk for the primary endpoint after FDR correction (**Figure 6** and **Supplemental Table 16)**. At a more stringent linkage disequilibrium clumping threshold of r^2^<0.01, genetically controlled levels of the top four proteins were nominally associated with risk of heart failure events in the same direction as the main findings, but did not reach FDR <0.05.

**Figure 6:**
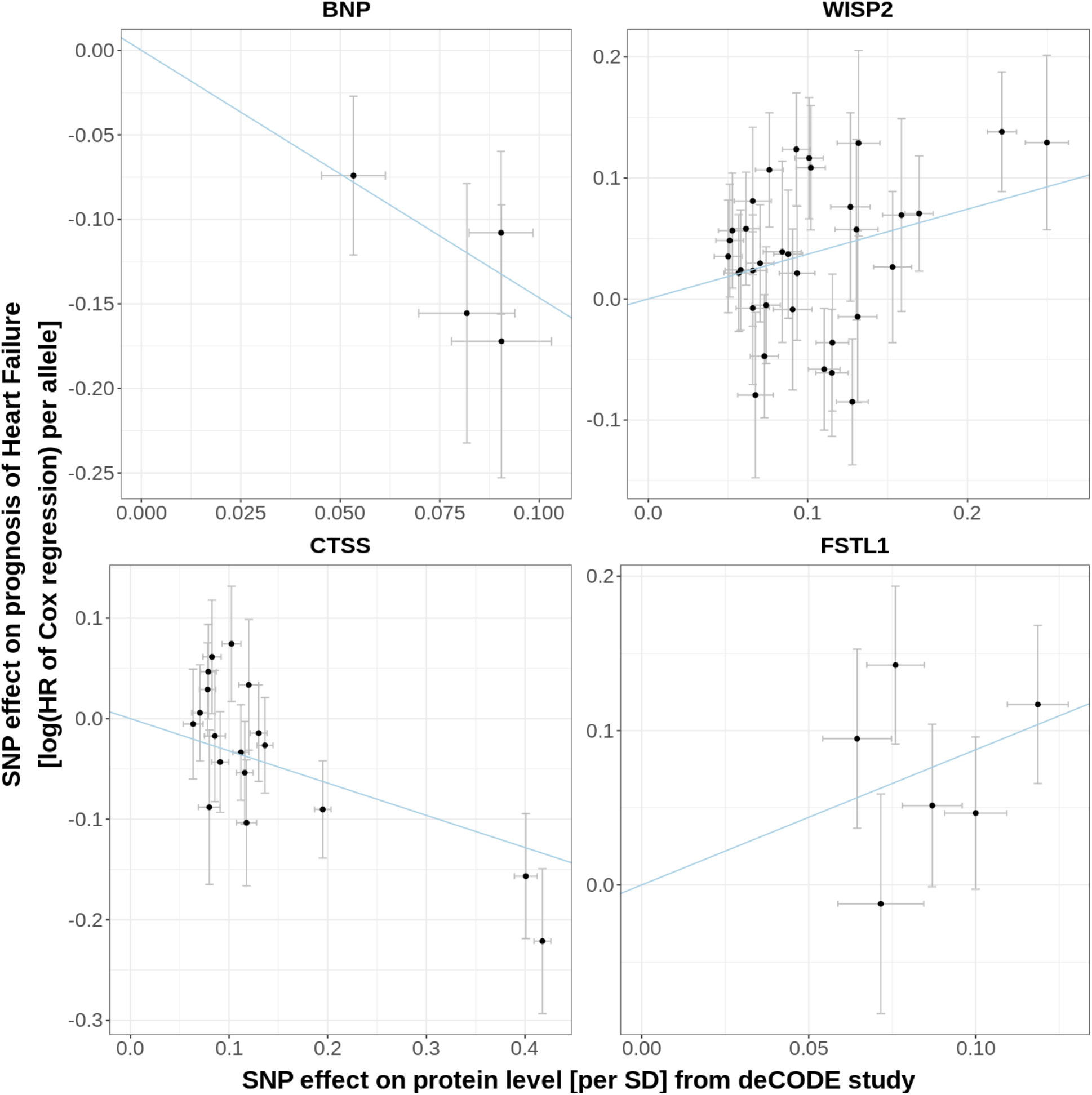
Comparison of Variant-Level Effects on Circulating BNP, WISP2, CTSS, FSLT1, and Risk for Heart Failure Hospitalization or Cardiovascular Death. Genetically controlled levels of four circulating proteins were significantly associated with risk for the primary endpoint by two sample Mendelian randomization analysis.

## Discussion

In two clinical trials in heart failure with reduced ejection fraction, we leveraged a 5034-aptamer proteomic assay to perform unbiased heart failure biomarker discovery. We identified 167 circulating proteins that predicted heart failure hospitalization or cardiovascular death in ATMOSPHERE and were replicated in PARADIGM-HF. SVEP1 is a novel heart failure biomarker whose prognostic value was comparable to NT-proBNP. We derived and externally validated a proteomic heart failure risk score whose performance exceeded an established clinical risk score and NT-proBNP. Exploratory Mendelian randomization analysis reproduced the known role of BNP in heart failure and nominated three additional proteins with potentially causal roles in the progression of heart failure. These results show the value of large-scale proteomics to identify biological mechanisms and clinically relevant biomarkers in heart failure.

Our results permit several conclusions. First, broad proteomic discovery identified numerous known and novel biomarkers for heart failure prognosis with high statistical confidence. We considered all aptamers available in the SomaScan platform rather than focusing on specific candidate proteins. Reassuringly, we reproduced associations for known biomarkers like NT-proBNPs,^7,26^ GDF15,^6^ angiopoetin-2,^27^ and thrombospondin-2.^28^ Several proteins were also identified in a recent single-center proteomics analysis of survival in heart failure with reduced ejection fraction survival conducted using plasma rather than serum specimens. Plasma samples may offer stronger proteomic signals but are less stable for storage. ^29^ Whereas most previous large-scale proteomic studies have investigated incident heart failure in healthy patients,^28,30^ we focused on predicting subsequent events in patients with established heart failure. Strengths of the present study include a large sample of patients with established heart failure, replication in a separate clinical trial, and rigorous clinical trial inclusion criteria, covariate ascertainment, and central endpoint adjudication. Our findings emphasize that circulating protein levels have strong prognostic value in heart failure.

Second, circulating SVEP1 is a novel heart failure biomarker with similar prognostic value to established biomarkers NT-proBNP and GDF15, which may suggest a role of vascular inflammation in heart failure. The presence of 2 different aptamers providing consistent measurements and a strong cis-pQTL support the accuracy of aptamer-based SVEP1 measurements. Plasma SVEP1 levels have been associated with risk of incident dementia, possibly with causal effect by Mendelian randomization,^31^ and with chronological age.^32^ SVEP1 is an extracellular matrix protein expressed in vascular smooth muscle cells. Human genetic and animal data nominate SVEP1 as a target in atherosclerosis. A rare missense variant is associated with coronary artery disease risk.^33^ Mendelian randomization analysis suggests that genetically controlled SVEP1 protein levels cause coronary artery disease, hypertension and diabetes.^34^ Murine knockout studies demonstrate the mechanism: SVEP1 promotes vascular inflammation and atherosclerosis through integrin, notch, and fibroblast growth factor receptor signaling.^34^ In our study, patients with ischemic heart disease did not have higher circulating SVEP1, indicating that this biomarker is more than just a marker of coronary disease.

Third, we derived a proteomic risk score that predicted cardiovascular death or heart failure hospitalization with better discrimination than the MAGGIC clinical risk score and NT-proBNP in an external validation cohort. The magnitude of improvement (C-statistic 0.05 higher than NT-proBNP, 0.10 higher than MAGGIC) was both statistically and clinically significant. Risk prediction in heart failure enables physicians to counsel patients about prognosis and identify high-risk patients for disease management programs or clinical trials. Computers can conveniently calculate proteomic scores from a single blood test, without requiring the clinician to enter parameters into a calculator. Proteomic scores may avoid biases specific to individual biomarkers. For example, NT-proBNP levels are lower in obesity and higher in atrial fibrillation for a given level of risk, while troponin levels are higher in patients with coronary ischemia. Moreover, proteomic risk scores may prove more reliable surrogate endpoints for phase II clinical trials, avoiding idiosyncratic effects of an experimental therapy on an individual biomarker that may be misleading.^35^ A 27-protein risk score for a composite of atherosclerotic and heart failure events was recently developed and validated for this clinical trial application.^36^ Notably, adding the MAGGIC risk score to the proteomic score did not improve discrimination; thus, the proteomic score incorporates the relevant information from a patient’s burden of clinical risk factors and may replace rather than complement clinical risk scores.

Fourth, Mendelian randomization suggested that some proteins identified in discovery proteomics may have causal roles in heart failure progression. The strongest result, BNP (whose precursors are encoded by the gene *NPPB)*, is biologically plausible. Variants in the *NPPB* promoter that cause greater levels of circulating natriuretic peptides have been associated with lower risk of hypertension and cardiovascular mortality.^37–39^ Our results extend these findings from healthy individuals to patients with established heart failure. Positive associations between genetically controlled levels of Follistatin Like 1 and WNT1-Inducible-Signaling Pathway Protein 2 and risk of heart failure hospitalization or cardiovascular death in our study should be considered in the context of preclinical models suggesting these proteins improve myocardial energetics, maladaptive remodeling, and myocardial fibrosis.^40,41^ Mendelian randomization findings should be considered exploratory given the small sample size of patients with available genetic and outcome data. The absence of a significant association between genetically controlled SVEP1 levels and the primary endpoint may reflect low statistical power. Ongoing collaborations to pool patients from multiple genotyped heart failure cohorts will provide greater power for pQTL-based genetic discovery.

These results should be considered in the context of the study design. The SomaScan modified aptamer proteomics platform has not been validated for all proteins in the panel. However, many have been validated by mass spectrometry, and *cis*-pQTLs have been identified for 552^8^ and 1046 proteins^18^ which strongly support aptamer specificity. Though this platform measures more proteins than any other proteomics assay available today, some serum proteins were not studied because a suitable aptamer is not available. Circulating serum protein levels do not necessarily reflect tissue expression or activity. Strict clinical trial inclusion criteria and exclusion of patients in decompensated heart failure may have homogenized protein levels, biasing results towards the null.

## Conclusion

Large-scale proteomic analysis in two clinical trials of patients with heart failure with reduced ejection fraction identified and replicated 167 circulating proteins significantly associated with cardiovascular death or heart failure hospitalization. This approach reproduced known associations and identified novel biomarkers including SVEP1, an extracellular matrix protein known to cause inflammation in vascular smooth muscle cells. A 64-protein risk score predicted heart failure events with greater discrimination than an established clinical risk score and NT-proBNP. Mendelian randomization extended previous evidence for a beneficial causal effect of NPPB expression to patients with established heart failure. These results show the value of large-scale proteomics to identify novel biological mechanisms and clinically relevant biomarkers in heart failure.

## Supporting information

Supplemental Material SOMAmer protein mapping

## Data Availability

The data, analytic methods, and study materials will not be made available to other researchers for purposes of reproducing the results

## Acknowledgements

None

## Notes

**Conflicts of Interest** Drs. Zhang, Mendelson, Serrano-Fernandez, Kaiser, Yates, Chen, Turner, Patel-Murray, Beste, Laramie, Prescott, Lefkowitz, and Chutkow, Ms. Healey, and Ms. Zhao are employees of Novartis. Dr. Cunningham reports no relationships with industry. Dr. Claggett reports consulting fees from Amgen, Boehringer-Ingelheim, Cardurion, Corvia, MyoKardia, and Novartis outside the submitted work. Dr. Jacob reports salary support from Novartis and Moderna. Dr. Abraham reports consulting fees from Abbott, ARCA biopharma, Boehringer Ingelheim, Cardionomic, CVRx, Edwards Lifesciences, Respicardia, Sensible Medical, and Vectorious, and salary support from V-Wave Medical. Dr. Jhund reports consulting fees, advisory board fees, and lecture fees from Novartis; advisory board fees from Cytokinetics; and grant support from Boehringer Ingelheim. Dr. Kober reports speakers honoriaria from Novo Nordisk, Novartis, AstraZeneca and Boehringer Ingleheim; support from AstraZeneca; and personal fees from Novartis and Bristol Myers Squibb as a speaker. Dr. Packer reports consulting fees from AbbVie, Akcea, Actavis, Amgen, AstraZeneca, Bayer, Boehringer Ingelheim, Cardiorentis, Daiichi Sankyo, Gilead, Johnson & Johnson, Novo Nordisk, Pfizer, Relypsa, Sanofi, Synthetic Biologics, and Theravance. Dr. Rouleau reports grants and consulting fees from Novartis and consulting fees from Abbott, AstraZeneca, MyoKardia, and Sanofi. Dr. Zile has received research funding from Novartis; and has been a consultant for Novartis, Abbott, Boston Scientific, CVRx, EBR, Endotronics, Ironwood, Merck, Medtronic, MyoKardia, and V Wave. Dr McMurray has received funding to his institution, Glasgow University, for his work on clinical trials, consulting, and other activities from Alnylam, Amgen, AstraZeneca, Bayer, Bristol Myers Squibb, Cardurion, Cytokinetics, GlaxoSmithKline, Novartis, Pfizer, and Theracos; and has received personal lecture fees from the Corpus, Abbott, Hickma, Sun Pharmaceuticals, and Medscape. Dr. Solomon reports Actelion, Alnylam, Amgen, AstraZeneca, Bellerophon, Bayer, Bristol Myers Squibb, Celladon, Cytokinetics, Eidos, Gilead, GSK, Ionis, Lilly, Mesoblast, MyoKardia, NIH/NHLBI, Neurotronik, Novartis, Novo Nordisk, Respicardia, Sanofi Pasteur, Theracos, and Us2.aI outside the submitted work; consulting fees from Abbott, Action, Akros, Alnylam, Amgen, Arena, AstraZeneca, Bayer, Boehringer-Ingelheim, Bristol Myers Squibb, Cardior, Cardurion, Corvia, Cytokinetics, DaiichiSankyo, GSK, Lilly, Merck, MyoKardia, Novartis, Roche, Theracos, Quantum Genomics, Cardurion, Janssen, Cardiac Dimensions, Tenaya, Sanofi-Pasteur, Dinaqor, Tremeau, CellProThera, Moderna, American Regent, and Sarepta; and participation on a Data Safety Monitoring Board or Advisory Board for Janssen.

### Competing Interest Statement

Drs. Zhang, Mendelson, Serrano-Fernandez, Kaiser, Yates, Chen, Turner, Patel-Murray, Prescott, Lefkowitz, and Chutkow, Ms. Healey, and Ms. Zhao are employees of Novartis. Dr. Cunningham reports no relationships with industry. Dr. Claggett reports consulting fees from Amgen, Boehringer-Ingelheim, Cardurion, Corvia, MyoKardia, and Novartis outside the submitted work. Dr. Jacob reports salary support from Novartis and Moderna. Dr. Abraham reports consulting fees from Abbott, ARCA biopharma, Boehringer Ingelheim, Cardionomic, CVRx, Edwards Lifesciences, Respicardia, Sensible Medical, and Vectorious, and salary support from V-Wave Medical. Dr. Jhund reports consulting fees, advisory board fees, and lecture fees from Novartis; advisory board fees from Cytokinetics; and grant support from Boehringer Ingelheim. Dr. Kober reports speakers honoriaria from Novo Nordisk, Novartis, AstraZeneca and Boehringer Ingleheim; support from AstraZeneca; and personal fees from Novartis and Bristol Myers Squibb as a speaker. Dr. Packer reports consulting fees from AbbVie, Akcea, Actavis, Amgen, AstraZeneca, Bayer, Boehringer Ingelheim, Cardiorentis, Daiichi Sankyo, Gilead, Johnson & Johnson, Novo Nordisk, Pfizer, Relypsa, Sanofi, Synthetic Biologics, and Theravance. Dr. Rouleau reports grants and consulting fees from Novartis and consulting fees from Abbott, AstraZeneca, MyoKardia, and Sanofi. Dr. Zile has received research funding from Novartis; and has been a consultant for Novartis, Abbott, Boston Scientific, CVRx, EBR, Endotronics, Ironwood, Merck, Medtronic, MyoKardia, and V Wave. Dr McMurray has received funding to his institution, Glasgow University, for his work on clinical trials, consulting, and other activities from Alnylam, Amgen, AstraZeneca, Bayer, Bristol Myers Squibb, Cardurion, Cytokinetics, GlaxoSmithKline, Novartis, Pfizer, and Theracos; and has received personal lecture fees from the Corpus, Abbott, Hickma, Sun Pharmaceuticals, and Medscape. Dr. Solomon reports Actelion, Alnylam, Amgen, AstraZeneca, Bellerophon, Bayer, Bristol Myers Squibb, Celladon, Cytokinetics, Eidos, Gilead, GSK, Ionis, Lilly, Mesoblast, MyoKardia, NIH/NHLBI, Neurotronik, Novartis, Novo Nordisk, Respicardia, Sanofi Pasteur, Theracos, and Us2.aI outside the submitted work; consulting fees from Abbott, Action, Akros, Alnylam, Amgen, Arena, AstraZeneca, Bayer, Boehringer-Ingelheim, Bristol Myers Squibb, Cardior, Cardurion, Corvia, Cytokinetics, DaiichiSankyo, GSK, Lilly, Merck, MyoKardia, Novartis, Roche, Theracos, Quantum Genomics, Cardurion, Janssen, Cardiac Dimensions, Tenaya, Sanofi-Pasteur, Dinaqor, Tremeau, CellProThera, Moderna, American Regent, and Sarepta; and participation on a Data Safety Monitoring Board or Advisory Board for Janssen.

### Funding Statement

The PARADIGM-HF and ATMOSPHERE trials, and this proteomic study, were funded by Novartis.

### Author Declarations

Secondary analysis of data from ATMOSPHERE and PARADIGM-HF was approved by the Mass General Brigham Institutional Review Board

