## Supplemental Material Figures and Tables for "Aptamer Proteomics for Biomarker Discovery in Heart Failure with Reduced Ejection Fraction"

**Supplemental Table 1: Baseline Characteristics of Patients With and Without Available Proteomic Data**

| | ATMOSPHERE | | PARADIGM-HF | |
|---|---|---|---|---|
| **Characteristic** | **Proteomic Data**, N = 1,258[1] | **No Proteomic Data**, N = 5,758[1] | **Proteomic Data**, N = 1,257[1] | **No Proteomic Data**, N = 7,185[1] |
| **Age (years)** | 67 (59, 73) | 64 (55, 72) | 68 (61, 74) | 64 (56, 72) |
| **Gender** | | | | |
| Female | 240 (19%) | 1,285 (22%) | 233 (19%) | 1,614 (22%) |
| Male | 1,018 (81%) | 4,473 (78%) | 1,024 (81%) | 5,571 (78%) |
| **Race** | | | | |
| Asian | 5 (0.4%) | 1,759 (31%) | 4 (0.3%) | 1,506 (21%) |
| Black | 12 (1.0%) | 97 (1.7%) | 14 (1.1%) | 414 (5.8%) |
| Caucasian | 1,205 (96%) | 3,387 (59%) | 1,213 (96%) | 4,366 (61%) |
| Native American | 0 (0%) | 42 (0.7%) | 0 (0%) | 172 (2.4%) |
| Other | 35 (2.8%) | 419 (7.3%) | 26 (2.1%) | 726 (10%) |
| Pacific Islander | 1 (<0.1%) | 0 (0%) | 0 (0%) | 1 (<0.1%) |
| Unknown | 0 | 54 | | |
| **Region** | | | | |
| Asia/Pacific and Other | 20 (1.6%) | 1,905 (33%) | 0 (0%) | 1,811 (25%) |
| Central/Eastern Europe | 506 (40%) | 1,438 (25%) | 584 (46%) | 2,219 (31%) |
| Latin America (including Central America) | 57 (4.5%) | 1,062 (18%) | 0 (0%) | 1,428 (20%) |
| North America | 54 (4.3%) | 123 (2.1%) | 56 (4.5%) | 546 (7.6%) |
| Western Europe | 621 (49%) | 1,230 (21%) | 617 (49%) | 1,181 (16%) |
| **Diabetes mellitus** | 389 (31%) | 1,547 (27%) | 476 (38%) | 2,440 (34%) |
| **Hypertension** | 893 (71%) | 3,439 (60%) | 963 (77%) | 5,007 (70%) |
| **Myocardial infarction** | 656 (52%) | 2,191 (38%) | 582 (46%) | 3,069 (43%) |
| **Ischemic cardiomyopathy** | 814 (65%) | 3,116 (54%) | 813 (65%) | 4,245 (59%) |
| **Stroke** | 97 (7.7%) | 395 (6.9%) | 137 (11%) | 592 (8.2%) |
| **Atrial fibrillation** | 518 (41%) | 1,872 (33%) | 606 (48%) | 2,505 (35%) |
| **NYHA function class** | | | | |
| I / II | 789 (63%) | 3,649 (63%) | 857 (68%) | 4,631 (64%) |
| III / IV | 469 (37%) | 2,109 (37%) | 400 (32%) | 2,554 (36%) |
| **Ejection fraction (%)** | 30.0 (25.0, 34.0) | 30.0 (25.0, 33.0) | 32 (28, 35) | 30 (25, 34) |
| Unknown | | | 0 | 1 |
| **Anticoagulant usage** | 493 (39%) | 1,685 (29%) | 560 (45%) | 2,139 (30%) |
| **ACEi** | 1,257 (100%) | 5,749 (100%) | 993 (79%) | 5,567 (77%) |
| **ARBi** | 35 (2.8%) | 110 (1.9%) | 266 (21%) | 1,641 (23%) |
| **Diuretic** | 1,028 (82%) | 4,458 (77%) | 1,058 (84%) | 5,913 (82%) |
| **Digoxin** | 312 (25%) | 2,002 (35%) | 280 (22%) | 2,279 (32%) |
| **Beta-blocker** | 1,187 (94%) | 5,392 (94%) | 1,201 (96%) | 6,758 (94%) |

| | | | | |
|---|---|---|---|---|
| **Cardiac resynchronization therapy** | 107 (8.5%) | 286 (5.0%) | 115 (9.1%) | 460 (6.4%) |
| **Implantable cardioverter defibrillator** | 317 (25%) | 668 (12%) | 312 (25%) | 934 (13%) |
| **BMI (kg/m²)** | 28.1 (25.2, 31.6) | 26.4 (23.3, 30.0) | 28.5 (25.4, 32.1) | 27.1 (24.1, 30.9) |
| Unknown | 3 | 14 | 0 | 10 |
| **Systolic blood pressure (mmHG)** | 130 (118, 140) | 125 (112, 140) | 130 (120, 142) | 126 (115, 140) |
| **eGFR (60mL/min/1.73m²)** | 72 (59, 86) | 74 (61, 88) | 64 (53, 77) | 67 (54, 80) |
| Unknown | 0 | 2 | | |
| **NT-proBNP (pg/mL)** | 1,342 (760, 2,390) | 1,390 (798, 2,512) | 1,492 (853, 2,935) | 1,642 (895, 3,315) |
| Unknown | 150 | 1,291 | 0 | 13 |

[1] Median (IQR); n (%)

Values are n (%) for categorical variables or median (interquartile range) for continuous variables. ACEi, angiotensin-converting enzyme inhibitor; ARB, angiotensin-receptor blocker; eGFR, estimated glomerular filtration rate; NT-proBNP, N-terminal pro–B-type natriuretic peptide; NYHA, New York Heart Association

**Supplemental Table 2: Proteins Significantly Associated With Heart Failure Hospitalization or Cardiovascular Death in Minimally Adjusted Model**

| | Discovery (ATMOSPHERE) | | Replication (PARADIGM) | |
|---|---|---|---|---|
| Protein | HR (95% CI) | *P* Value | HR (95% CI) | *P* Value |
| GDF15 | 1.50 (1.38-1.64) | $1.69 \times 10^{-21}$ | 1.66 (1.47-1.88) | $3.43 \times 10^{-16}$ |
| SVEP1 | 1.46 (1.35-1.58) | $9.42 \times 10^{-21}$ | 1.60 (1.44-1.79) | $2.01 \times 10^{-17}$ |
| ANGPT2 | 1.48 (1.35-1.61) | $1.40 \times 10^{-18}$ | 1.73 (1.54-1.95) | $6.84 \times 10^{-20}$ |
| NPPB | 1.56 (1.40-1.73) | $2.97 \times 10^{-16}$ | 1.63 (1.44-1.86) | $7.17 \times 10^{-14}$ |
| THBS2 | 1.42 (1.30-1.54) | $6.66 \times 10^{-16}$ | 1.66 (1.49-1.85) | $3.35 \times 10^{-20}$ |
| QSOX1 | 1.42 (1.31-1.55) | $1.14 \times 10^{-15}$ | 1.46 (1.31-1.62) | $2.05 \times 10^{-12}$ |
| ADAMTSL2 | 1.40 (1.28-1.52) | $2.51 \times 10^{-14}$ | 1.48 (1.33-1.65) | $1.31 \times 10^{-12}$ |
| LTBP4 | 1.43 (1.30-1.57) | $1.16 \times 10^{-13}$ | 1.35 (1.23-1.48) | $4.96 \times 10^{-10}$ |
| CST3 | 1.37 (1.25-1.50) | $6.85 \times 10^{-12}$ | 1.52 (1.33-1.73) | $4.69 \times 10^{-10}$ |
| SPON2 | 1.38 (1.26-1.52) | $1.14 \times 10^{-11}$ | 1.54 (1.35-1.76) | $1.77 \times 10^{-10}$ |
| B2M | 1.37 (1.25-1.51) | $1.47 \times 10^{-11}$ | 1.45 (1.26-1.65) | $6.70 \times 10^{-8}$ |
| NFASC | 1.33 (1.22-1.44) | $1.70 \times 10^{-11}$ | 1.54 (1.38-1.73) | $1.21 \times 10^{-13}$ |
| IL1RL1 | 1.39 (1.26-1.53) | $2.16 \times 10^{-11}$ | 1.48 (1.31-1.67) | $5.67 \times 10^{-10}$ |
| SPON1 | 1.33 (1.23-1.45) | $2.82 \times 10^{-11}$ | 1.52 (1.39-1.68) | $4.17 \times 10^{-18}$ |
| IGFBP7 | 1.35 (1.24-1.48) | $2.93 \times 10^{-11}$ | 1.58 (1.41-1.76) | $1.53 \times 10^{-16}$ |
| COL6A3 | 1.34 (1.23-1.47) | $3.62 \times 10^{-11}$ | 1.53 (1.36-1.72) | $1.74 \times 10^{-12}$ |
| FSTL3 | 1.39 (1.26-1.53) | $7.01 \times 10^{-11}$ | 1.56 (1.36-1.78) | $8.76 \times 10^{-11}$ |
| TNC | 1.35 (1.23-1.48) | $7.70 \times 10^{-11}$ | 1.37 (1.24-1.52) | $8.37 \times 10^{-10}$ |
| EFEMP1 | 1.33 (1.22-1.45) | $2.43 \times 10^{-10}$ | 1.32 (1.22-1.44) | $1.24 \times 10^{-10}$ |
| TNNI3 | 1.25 (1.17-1.34) | $3.78 \times 10^{-10}$ | 1.27 (1.18-1.38) | $9.99 \times 10^{-10}$ |
| PXDN | 1.39 (1.25-1.54) | $4.82 \times 10^{-10}$ | 1.45 (1.27-1.66) | $5.75 \times 10^{-8}$ |
| SEMA6B | 1.33 (1.21-1.45) | $4.83 \times 10^{-10}$ | 1.25 (1.15-1.36) | $5.97 \times 10^{-8}$ |
| MMP19 | 1.36 (1.23-1.49) | $6.31 \times 10^{-10}$ | 1.59 (1.43-1.77) | $2.59 \times 10^{-17}$ |
| ENG | 1.28 (1.18-1.38) | $8.36 \times 10^{-10}$ | 1.33 (1.18-1.50) | $2.94 \times 10^{-6}$ |
| ATP5PF | 1.26 (1.17-1.36) | $1.45 \times 10^{-9}$ | 1.30 (1.19-1.42) | $2.22 \times 10^{-8}$ |
| TREM1 | 1.34 (1.22-1.47) | $1.51 \times 10^{-9}$ | 1.36 (1.20-1.55) | $2.52 \times 10^{-6}$ |
| IL19 | 1.32 (1.21-1.45) | $1.77 \times 10^{-9}$ | 1.36 (1.21-1.52) | $1.40 \times 10^{-7}$ |
| EPO | 1.30 (1.19-1.41) | $1.79 \times 10^{-9}$ | 1.60 (1.45-1.78) | $4.50 \times 10^{-19}$ |
| COL28A1 | 1.33 (1.21-1.46) | $2.24 \times 10^{-9}$ | 1.57 (1.38-1.79) | $6.29 \times 10^{-12}$ |
| MFAP4 | 1.29 (1.18-1.40) | $4.07 \times 10^{-9}$ | 1.51 (1.32-1.73) | $3.17 \times 10^{-9}$ |
| RSPO4 | 1.20 (1.13-1.28) | $4.81 \times 10^{-9}$ | 1.37 (1.26-1.48) | $2.82 \times 10^{-14}$ |
| NBL1 | 1.31 (1.19-1.43) | $5.21 \times 10^{-9}$ | 1.37 (1.21-1.55) | $6.00 \times 10^{-7}$ |
| IGFBP1 | 1.32 (1.20-1.45) | $6.16 \times 10^{-9}$ | 1.38 (1.22-1.57) | $4.63 \times 10^{-7}$ |
| PTK7 | 1.31 (1.19-1.43) | $6.54 \times 10^{-9}$ | 1.56 (1.38-1.77) | $6.80 \times 10^{-13}$ |
| BOC | 1.31 (1.19-1.43) | $7.73 \times 10^{-9}$ | 1.28 (1.14-1.44) | $4.57 \times 10^{-5}$ |
| CD5 | 1.23 (1.15-1.32) | $8.16 \times 10^{-9}$ | 1.28 (1.19-1.37) | $2.02 \times 10^{-12}$ |
| FABP3 | 1.33 (1.21-1.47) | $8.59 \times 10^{-9}$ | 1.43 (1.25-1.64) | $1.60 \times 10^{-7}$ |
| PTPRU | 1.26 (1.17-1.37) | $9.98 \times 10^{-9}$ | 1.44 (1.29-1.60) | $1.84 \times 10^{-11}$ |

| | | | | |
|---|---|---|---|---|
| C1QTNF1 | 1.30 (1.18-1.42) | $1.26 \times 10^{-8}$ | 1.48 (1.33-1.65) | $2.07 \times 10^{-12}$ |
| KERA | 1.30 (1.18-1.42) | $1.43 \times 10^{-8}$ | 1.33 (1.21-1.46) | $2.83 \times 10^{-9}$ |
| ADM | 1.28 (1.17-1.39) | $1.43 \times 10^{-8}$ | 1.21 (1.10-1.33) | $9.73 \times 10^{-5}$ |
| CTSH | 1.25 (1.16-1.35) | $1.87 \times 10^{-8}$ | 1.58 (1.40-1.78) | $1.16 \times 10^{-13}$ |
| EPHA2 | 1.31 (1.19-1.44) | $2.15 \times 10^{-8}$ | 1.43 (1.25-1.63) | $9.24 \times 10^{-8}$ |
| RNASE1 | 1.32 (1.20-1.45) | $2.16 \times 10^{-8}$ | 1.48 (1.30-1.70) | $5.73 \times 10^{-9}$ |
| TFF3 | 1.28 (1.17-1.40) | $2.63 \times 10^{-8}$ | 1.54 (1.36-1.74) | $1.48 \times 10^{-11}$ |
| STC1 | 1.29 (1.18-1.42) | $4.70 \times 10^{-8}$ | 1.49 (1.31-1.70) | $1.06 \times 10^{-9}$ |
| TAGLN | 1.33 (1.20-1.48) | $7.43 \times 10^{-8}$ | 1.58 (1.37-1.81) | $1.01 \times 10^{-10}$ |
| CXCL13 | 1.25 (1.15-1.35) | $8.08 \times 10^{-8}$ | 1.23 (1.11-1.37) | $6.87 \times 10^{-5}$ |
| WFDC2 | 1.30 (1.18-1.43) | $1.01 \times 10^{-7}$ | 1.55 (1.36-1.76) | $6.03 \times 10^{-11}$ |
| DCN | 1.25 (1.15-1.36) | $1.06 \times 10^{-7}$ | 1.32 (1.18-1.48) | $2.39 \times 10^{-6}$ |
| RNASE6 | 1.28 (1.17-1.40) | $1.31 \times 10^{-7}$ | 1.31 (1.19-1.45) | $9.97 \times 10^{-8}$ |
| IGSF3 | 1.26 (1.16-1.38) | $1.55 \times 10^{-7}$ | 1.30 (1.16-1.45) | $6.42 \times 10^{-6}$ |
| ICAM5 | 1.27 (1.16-1.40) | $2.23 \times 10^{-7}$ | 1.55 (1.36-1.75) | $8.10 \times 10^{-12}$ |
| CHST15 | 1.28 (1.17-1.41) | $2.46 \times 10^{-7}$ | 1.52 (1.35-1.72) | $7.60 \times 10^{-12}$ |
| CILP2 | 0.79 (0.72-0.86) | $3.62 \times 10^{-7}$ | 0.70 (0.62-0.79) | $1.40 \times 10^{-8}$ |
| RELT | 1.26 (1.15-1.38) | $3.92 \times 10^{-7}$ | 1.27 (1.15-1.41) | $1.78 \times 10^{-6}$ |
| Igh | 1.25 (1.15-1.37) | $7.29 \times 10^{-7}$ | 1.33 (1.19-1.48) | $3.25 \times 10^{-7}$ |
| FABP4 | 1.27 (1.16-1.40) | $7.71 \times 10^{-7}$ | 1.28 (1.13-1.45) | $8.38 \times 10^{-5}$ |
| MMP7 | 1.27 (1.15-1.39) | $1.04 \times 10^{-6}$ | 1.42 (1.27-1.58) | $6.96 \times 10^{-10}$ |
| COLEC11 | 1.25 (1.14-1.37) | $1.12 \times 10^{-6}$ | 1.33 (1.18-1.50) | $2.11 \times 10^{-6}$ |
| CCL14 | 1.27 (1.16-1.41) | $1.15 \times 10^{-6}$ | 1.46 (1.28-1.67) | $2.25 \times 10^{-8}$ |
| B3GAT3 | 1.25 (1.14-1.37) | $1.24 \times 10^{-6}$ | 1.28 (1.14-1.45) | $3.57 \times 10^{-5}$ |
| TNFRSF1A | 1.25 (1.14-1.38) | $1.40 \times 10^{-6}$ | 1.32 (1.17-1.49) | $7.38 \times 10^{-6}$ |
| MMP2 | 1.26 (1.14-1.38) | $2.13 \times 10^{-6}$ | 1.38 (1.22-1.57) | $3.89 \times 10^{-7}$ |
| UMOD | 1.25 (1.14-1.37) | $2.34 \times 10^{-6}$ | 1.37 (1.22-1.53) | $8.18 \times 10^{-8}$ |
| FCN3 | 0.78 (0.71-0.87) | $2.86 \times 10^{-6}$ | 0.80 (0.72-0.89) | $3.38 \times 10^{-5}$ |
| TFRC | 1.24 (1.13-1.35) | $3.12 \times 10^{-6}$ | 1.46 (1.31-1.64) | $6.99 \times 10^{-11}$ |
| KLK10 | 1.24 (1.13-1.35) | $4.50 \times 10^{-6}$ | 1.27 (1.12-1.43) | $1.11 \times 10^{-4}$ |
| LAMC2 | 1.22 (1.12-1.33) | $4.86 \times 10^{-6}$ | 1.28 (1.17-1.39) | $2.74 \times 10^{-8}$ |
| KREMEN1 | 1.23 (1.12-1.34) | $5.18 \times 10^{-6}$ | 1.23 (1.12-1.36) | $2.45 \times 10^{-5}$ |
| CTHRC1 | 1.19 (1.11-1.29) | $5.29 \times 10^{-6}$ | 1.23 (1.13-1.34) | $2.03 \times 10^{-6}$ |
| PRSS22 | 1.22 (1.12-1.32) | $5.35 \times 10^{-6}$ | 1.35 (1.19-1.52) | $1.34 \times 10^{-6}$ |
| FGF23 | 1.15 (1.08-1.22) | $5.63 \times 10^{-6}$ | 1.41 (1.29-1.53) | $4.72 \times 10^{-15}$ |
| WFDC1 | 1.24 (1.13-1.35) | $5.86 \times 10^{-6}$ | 1.51 (1.33-1.73) | $4.84 \times 10^{-10}$ |
| LUM | 1.23 (1.13-1.35) | $6.00 \times 10^{-6}$ | 1.45 (1.30-1.63) | $1.81 \times 10^{-10}$ |
| GABARAPL1 | 1.21 (1.11-1.31) | $6.37 \times 10^{-6}$ | 1.33 (1.18-1.49) | $1.65 \times 10^{-6}$ |
| TNFRSF11B | 1.24 (1.13-1.36) | $6.79 \times 10^{-6}$ | 1.44 (1.26-1.64) | $7.39 \times 10^{-8}$ |
| RBL2 | 1.22 (1.12-1.33) | $6.89 \times 10^{-6}$ | 1.40 (1.27-1.55) | $3.94 \times 10^{-11}$ |
| WARS1 | 1.24 (1.13-1.36) | $7.58 \times 10^{-6}$ | 1.38 (1.22-1.55) | $1.60 \times 10^{-7}$ |
| PLXDC2 | 1.20 (1.11-1.30) | $7.68 \times 10^{-6}$ | 1.51 (1.35-1.69) | $3.33 \times 10^{-13}$ |

| | | | | |
|---|---|---|---|---|
| PARVA | 1.21 (1.12-1.32) | $8.14 \times 10^{-6}$ | 1.23 (1.13-1.35) | $4.93 \times 10^{-6}$ |
| CHRDL2 | 1.23 (1.12-1.35) | $8.78 \times 10^{-6}$ | 1.40 (1.23-1.59) | $3.18 \times 10^{-7}$ |
| DNAJB12 | 1.19 (1.10-1.29) | $8.90 \times 10^{-6}$ | 1.30 (1.17-1.44) | $1.21 \times 10^{-6}$ |
| CNDP1 | 0.81 (0.74-0.89) | $9.55 \times 10^{-6}$ | 0.77 (0.69-0.87) | $1.43 \times 10^{-5}$ |
| TST | 1.22 (1.12-1.33) | $1.17 \times 10^{-5}$ | 1.36 (1.21-1.54) | $3.98 \times 10^{-7}$ |
| GAS1 | 1.22 (1.11-1.33) | $1.99 \times 10^{-5}$ | 1.23 (1.11-1.35) | $5.26 \times 10^{-5}$ |
| AGRP | 1.17 (1.09-1.26) | $2.12 \times 10^{-5}$ | 1.31 (1.18-1.44) | $2.28 \times 10^{-7}$ |
| CD163 | 1.22 (1.11-1.33) | $2.20 \times 10^{-5}$ | 1.31 (1.15-1.48) | $3.05 \times 10^{-5}$ |
| TNFRSF1B | 1.23 (1.12-1.36) | $2.42 \times 10^{-5}$ | 1.41 (1.23-1.62) | $8.90 \times 10^{-7}$ |
| CHST9 | 1.17 (1.09-1.26) | $3.16 \times 10^{-5}$ | 1.20 (1.12-1.30) | $1.21 \times 10^{-6}$ |
| COL18A1 | 1.22 (1.11-1.34) | $3.17 \times 10^{-5}$ | 1.45 (1.27-1.65) | $2.74 \times 10^{-8}$ |
| FCGR3B | 1.21 (1.11-1.33) | $3.53 \times 10^{-5}$ | 1.37 (1.21-1.55) | $9.17 \times 10^{-7}$ |
| SCARF2 | 1.16 (1.08-1.25) | $3.66 \times 10^{-5}$ | 1.31 (1.16-1.48) | $9.84 \times 10^{-6}$ |
| PPP4R3A | 1.19 (1.09-1.29) | $3.96 \times 10^{-5}$ | 1.27 (1.13-1.42) | $4.76 \times 10^{-5}$ |
| GRN | 1.21 (1.10-1.33) | $4.25 \times 10^{-5}$ | 1.46 (1.29-1.65) | $1.33 \times 10^{-9}$ |
| CD93 | 1.21 (1.10-1.32) | $4.77 \times 10^{-5}$ | 1.43 (1.26-1.61) | $1.59 \times 10^{-8}$ |
| TFF2 | 1.19 (1.09-1.29) | $5.01 \times 10^{-5}$ | 1.29 (1.18-1.40) | $5.85 \times 10^{-9}$ |
| NRP1 | 1.20 (1.10-1.31) | $5.14 \times 10^{-5}$ | 1.37 (1.22-1.53) | $1.04 \times 10^{-7}$ |
| IGFBP2 | 1.24 (1.12-1.37) | $5.86 \times 10^{-5}$ | 1.33 (1.16-1.53) | $5.34 \times 10^{-5}$ |
| HTRA2 | 1.19 (1.09-1.29) | $6.07 \times 10^{-5}$ | 1.28 (1.19-1.38) | $6.04 \times 10^{-11}$ |
| CTSB | 1.21 (1.10-1.33) | $6.11 \times 10^{-5}$ | 1.42 (1.25-1.61) | $1.03 \times 10^{-7}$ |
| IBSP | 1.18 (1.09-1.29) | $7.51 \times 10^{-5}$ | 1.31 (1.17-1.46) | $1.27 \times 10^{-6}$ |
| RNASE4 | 1.20 (1.10-1.32) | $8.05 \times 10^{-5}$ | 1.41 (1.23-1.61) | $9.63 \times 10^{-7}$ |
| MRC2 | 1.20 (1.09-1.31) | $9.82 \times 10^{-5}$ | 1.30 (1.15-1.47) | $3.24 \times 10^{-5}$ |
| ROBO2 | 1.20 (1.10-1.32) | $1.10 \times 10^{-4}$ | 1.42 (1.26-1.60) | $2.23 \times 10^{-8}$ |
| BMP6 | 1.18 (1.08-1.28) | $1.34 \times 10^{-4}$ | 1.33 (1.20-1.46) | $2.72 \times 10^{-8}$ |
| GNPTG | 1.15 (1.07-1.23) | $1.49 \times 10^{-4}$ | 1.46 (1.30-1.63) | $3.29 \times 10^{-11}$ |
| SIGLEC7 | 1.19 (1.09-1.30) | $1.50 \times 10^{-4}$ | 1.36 (1.22-1.51) | $1.33 \times 10^{-8}$ |
| FSTL1 | 1.19 (1.09-1.31) | $1.95 \times 10^{-4}$ | 1.43 (1.28-1.60) | $2.71 \times 10^{-10}$ |
| MENT | 0.84 (0.77-0.92) | $1.97 \times 10^{-4}$ | 0.76 (0.67-0.86) | $1.40 \times 10^{-5}$ |
| FLRT3 | 1.18 (1.08-1.29) | $2.00 \times 10^{-4}$ | 1.26 (1.13-1.41) | $4.40 \times 10^{-5}$ |
| ADAM12 | 1.16 (1.07-1.25) | $2.04 \times 10^{-4}$ | 1.27 (1.15-1.39) | $8.31 \times 10^{-7}$ |
| IL6ST | 1.20 (1.09-1.31) | $2.14 \times 10^{-4}$ | 1.35 (1.18-1.54) | $6.70 \times 10^{-6}$ |
| IL18R1 | 1.16 (1.07-1.25) | $2.46 \times 10^{-4}$ | 1.27 (1.13-1.42) | $4.64 \times 10^{-5}$ |
| INHBB | 1.18 (1.08-1.30) | $2.72 \times 10^{-4}$ | 1.37 (1.21-1.55) | $5.77 \times 10^{-7}$ |
| AMIGO2 | 1.19 (1.08-1.30) | $2.82 \times 10^{-4}$ | 1.34 (1.19-1.50) | $6.16 \times 10^{-7}$ |
| SEMA6A | 1.18 (1.08-1.30) | $3.02 \times 10^{-4}$ | 1.30 (1.15-1.46) | $1.62 \times 10^{-5}$ |
| GREM2 | 1.17 (1.08-1.28) | $3.10 \times 10^{-4}$ | 1.31 (1.19-1.44) | $7.61 \times 10^{-8}$ |
| SFRP1 | 1.18 (1.08-1.29) | $3.54 \times 10^{-4}$ | 1.29 (1.16-1.43) | $1.63 \times 10^{-6}$ |
| FGF7 | 1.15 (1.07-1.25) | $3.73 \times 10^{-4}$ | 1.32 (1.18-1.48) | $7.82 \times 10^{-7}$ |
| NTF3 | 1.14 (1.06-1.22) | $3.79 \times 10^{-4}$ | 1.22 (1.11-1.35) | $7.85 \times 10^{-5}$ |
| CDH7 | 0.85 (0.78-0.93) | $4.33 \times 10^{-4}$ | 0.78 (0.69-0.88) | $5.31 \times 10^{-5}$ |

| | | | | |
|---|---|---|---|---|
| PIGR | 1.18 (1.07-1.29) | $4.41 \times 10^{-4}$ | 1.33 (1.18-1.49) | $1.98 \times 10^{-6}$ |
| IGF1R | 1.17 (1.07-1.28) | $4.95 \times 10^{-4}$ | 1.34 (1.18-1.51) | $3.21 \times 10^{-6}$ |
| VEGFD | 1.16 (1.07-1.26) | $5.02 \times 10^{-4}$ | 1.39 (1.27-1.51) | $7.18 \times 10^{-14}$ |
| NOTUM | 0.85 (0.78-0.93) | $5.30 \times 10^{-4}$ | 0.72 (0.64-0.82) | $1.89 \times 10^{-7}$ |
| IL6 | 1.12 (1.05-1.20) | $5.32 \times 10^{-4}$ | 1.17 (1.08-1.27) | $1.06 \times 10^{-4}$ |
| CD300C | 1.18 (1.08-1.30) | $5.51 \times 10^{-4}$ | 1.32 (1.16-1.50) | $3.59 \times 10^{-5}$ |
| CRELD1 | 1.18 (1.08-1.30) | $5.65 \times 10^{-4}$ | 1.29 (1.14-1.46) | $6.79 \times 10^{-5}$ |
| SCARA5 | 1.17 (1.07-1.28) | $5.82 \times 10^{-4}$ | 1.39 (1.28-1.52) | $1.43 \times 10^{-14}$ |
| PLA2G2A | 1.17 (1.07-1.29) | $6.37 \times 10^{-4}$ | 1.42 (1.26-1.60) | $9.31 \times 10^{-9}$ |
| PSMD7 | 1.14 (1.06-1.24) | $6.65 \times 10^{-4}$ | 0.70 (0.60-0.82) | $1.52 \times 10^{-5}$ |
| ANGPTL1 | 1.16 (1.06-1.27) | $7.23 \times 10^{-4}$ | 1.32 (1.16-1.50) | $2.44 \times 10^{-5}$ |
| NELL2 | 1.18 (1.07-1.30) | $7.89 \times 10^{-4}$ | 1.23 (1.12-1.35) | $1.95 \times 10^{-5}$ |
| OLFM1 | 1.13 (1.05-1.22) | $8.81 \times 10^{-4}$ | 1.24 (1.13-1.35) | $2.12 \times 10^{-6}$ |
| ESM1 | 1.15 (1.06-1.25) | $8.85 \times 10^{-4}$ | 1.16 (1.08-1.25) | $7.88 \times 10^{-5}$ |
| MMP12 | 1.18 (1.07-1.30) | $9.77 \times 10^{-4}$ | 1.38 (1.22-1.57) | $3.21 \times 10^{-7}$ |
| CHRDL1 | 1.18 (1.07-1.30) | $1.14 \times 10^{-3}$ | 1.47 (1.27-1.70) | $1.82 \times 10^{-7}$ |
| UNC5C | 1.18 (1.07-1.30) | $1.15 \times 10^{-3}$ | 1.47 (1.28-1.68) | $6.10 \times 10^{-8}$ |
| CCN1 | 1.13 (1.05-1.22) | $1.15 \times 10^{-3}$ | 1.26 (1.17-1.36) | $2.76 \times 10^{-9}$ |
| SCARF1 | 1.16 (1.06-1.26) | $1.21 \times 10^{-3}$ | 1.30 (1.16-1.46) | $5.51 \times 10^{-6}$ |
| CCDC80 | 1.18 (1.07-1.30) | $1.37 \times 10^{-3}$ | 1.43 (1.25-1.63) | $1.20 \times 10^{-7}$ |
| CILP | 1.14 (1.05-1.24) | $1.42 \times 10^{-3}$ | 1.23 (1.11-1.36) | $6.72 \times 10^{-5}$ |
| EPHA10 | 1.13 (1.05-1.22) | $1.43 \times 10^{-3}$ | 1.21 (1.10-1.32) | $5.48 \times 10^{-5}$ |
| BMP7 | 1.15 (1.05-1.25) | $1.46 \times 10^{-3}$ | 1.24 (1.11-1.37) | $7.17 \times 10^{-5}$ |
| TGFB3 | 1.14 (1.05-1.23) | $1.46 \times 10^{-3}$ | 1.22 (1.12-1.33) | $9.40 \times 10^{-6}$ |
| UNC5B | 1.17 (1.06-1.28) | $1.50 \times 10^{-3}$ | 1.29 (1.17-1.41) | $1.15 \times 10^{-7}$ |
| DNASE1L2 | 1.14 (1.05-1.24) | $1.56 \times 10^{-3}$ | 1.32 (1.20-1.46) | $5.56 \times 10^{-8}$ |
| EBI3 | 1.17 (1.06-1.28) | $1.66 \times 10^{-3}$ | 1.49 (1.30-1.71) | $1.16 \times 10^{-8}$ |
| SLPI | 1.16 (1.06-1.27) | $1.67 \times 10^{-3}$ | 1.28 (1.13-1.44) | $7.51 \times 10^{-5}$ |
| BCHE | 0.86 (0.79-0.95) | $1.76 \times 10^{-3}$ | 0.75 (0.66-0.85) | $1.32 \times 10^{-5}$ |
| PCDHGA10 | 1.12 (1.04-1.21) | $1.83 \times 10^{-3}$ | 1.28 (1.18-1.38) | $3.51 \times 10^{-10}$ |
| NRP2 | 1.16 (1.06-1.28) | $1.95 \times 10^{-3}$ | 1.34 (1.20-1.51) | $3.70 \times 10^{-7}$ |
| ITGA1\|ITGB1 | 1.15 (1.05-1.26) | $2.04 \times 10^{-3}$ | 1.28 (1.15-1.42) | $3.10 \times 10^{-6}$ |
| CCL21 | 1.15 (1.05-1.26) | $2.30 \times 10^{-3}$ | 1.37 (1.22-1.55) | $1.40 \times 10^{-7}$ |
| MINPP1 | 1.14 (1.05-1.25) | $2.37 \times 10^{-3}$ | 1.33 (1.21-1.46) | $8.80 \times 10^{-10}$ |
| PLA2R1 | 1.14 (1.05-1.24) | $2.57 \times 10^{-3}$ | 1.40 (1.26-1.55) | $4.85 \times 10^{-10}$ |
| CERT1 | 1.14 (1.05-1.24) | $2.67 \times 10^{-3}$ | 1.28 (1.16-1.42) | $1.90 \times 10^{-6}$ |
| HSPA1A | 1.14 (1.05-1.24) | $2.73 \times 10^{-3}$ | 1.33 (1.20-1.48) | $1.49 \times 10^{-7}$ |
| TXNDC5 | 1.15 (1.05-1.25) | $2.77 \times 10^{-3}$ | 1.41 (1.24-1.61) | $2.77 \times 10^{-7}$ |
| APOF | 1.17 (1.05-1.30) | $3.16 \times 10^{-3}$ | 1.37 (1.19-1.58) | $1.96 \times 10^{-5}$ |
| C1QL1 | 1.14 (1.05-1.25) | $3.44 \times 10^{-3}$ | 1.23 (1.12-1.36) | $1.68 \times 10^{-5}$ |
| TMED10 | 1.12 (1.04-1.22) | $3.50 \times 10^{-3}$ | 1.44 (1.27-1.63) | $4.58 \times 10^{-9}$ |
| MATN2 | 1.16 (1.05-1.27) | $3.62 \times 10^{-3}$ | 1.32 (1.15-1.52) | $6.98 \times 10^{-5}$ |

| | | | | |
|---|---|---|---|---|
| LRRC32 | 1.12 (1.04-1.20) | $3.75 \times 10^{-3}$ | 1.35 (1.19-1.54) | $2.59 \times 10^{-6}$ |
| EFNA4 | 1.12 (1.04-1.21) | $3.85 \times 10^{-3}$ | 1.25 (1.14-1.37) | $3.40 \times 10^{-6}$ |
| FJX1 | 1.14 (1.04-1.25) | $3.95 \times 10^{-3}$ | 1.26 (1.16-1.37) | $1.79 \times 10^{-7}$ |

**Supplemental Table 3: Proteins Significantly Associated With Heart Failure Hospitalization or Cardiovascular Death in Minimally Adjusted Model, With Hazard Ratios Expressed per Doubling in Protein Value**

| | Discovery (ATMOSPHERE) | | Replication (PARADIGM) | |
|---|---|---|---|---|
| **Protein** | **HR (95% CI)** | ***P* Value** | **HR (95% CI)** | ***P* Value** |
| GDF15 | 1.90 (1.67-2.17) | $1.69 \times 10^{-21}$ | 2.09 (1.75- 2.50) | $3.43 \times 10^{-16}$ |
| SVEP1 | 1.72 (1.54-1.93) | $9.42 \times 10^{-21}$ | 1.85 (1.61- 2.14) | $2.01 \times 10^{-17}$ |
| ANGPT2 | 1.63 (1.46-1.82) | $1.40 \times 10^{-18}$ | 2.09 (1.78- 2.45) | $6.84 \times 10^{-20}$ |
| NPPB | 1.56 (1.40-1.73) | $2.97 \times 10^{-16}$ | 1.60 (1.41- 1.81) | $7.17 \times 10^{-14}$ |
| THBS2 | 1.36 (1.26-1.46) | $6.66 \times 10^{-16}$ | 1.62 (1.46- 1.80) | $3.35 \times 10^{-20}$ |
| QSOX1 | 5.02 (3.38-7.45) | $1.14 \times 10^{-15}$ | 5.16 (3.27- 8.16) | $2.05 \times 10^{-12}$ |
| ADAMTSL2 | 1.97 (1.65-2.34) | $2.51 \times 10^{-14}$ | 2.26 (1.80- 2.83) | $1.31 \times 10^{-12}$ |
| LTBP4 | 2.25 (1.82-2.79) | $1.16 \times 10^{-13}$ | 1.95 (1.58- 2.40) | $4.96 \times 10^{-10}$ |
| CST3 | 2.50 (1.93-3.25) | $6.85 \times 10^{-12}$ | 3.00 (2.13- 4.25) | $4.69 \times 10^{-10}$ |
| SPON2 | 1.91 (1.59-2.31) | $1.14 \times 10^{-11}$ | 2.37 (1.82- 3.09) | $1.77 \times 10^{-10}$ |
| B2M | 2.08 (1.68-2.57) | $1.47 \times 10^{-11}$ | 2.19 (1.65- 2.92) | $6.70 \times 10^{-8}$ |
| NFASC | 2.36 (1.84-3.03) | $1.70 \times 10^{-11}$ | 2.97 (2.23- 3.96) | $1.21 \times 10^{-13}$ |
| IL1RL1 | 1.58 (1.38-1.81) | $2.16 \times 10^{-11}$ | 1.72 (1.45- 2.05) | $5.67 \times 10^{-10}$ |
| SPON1 | 1.91 (1.58-2.31) | $2.82 \times 10^{-11}$ | 2.42 (1.98- 2.95) | $4.17 \times 10^{-18}$ |
| IGFBP7 | 2.07 (1.67-2.56) | $2.93 \times 10^{-11}$ | 2.84 (2.22- 3.64) | $1.53 \times 10^{-16}$ |
| COL6A3 | 2.43 (1.87-3.17) | $3.62 \times 10^{-11}$ | 2.55 (1.97- 3.31) | $1.74 \times 10^{-12}$ |
| FSTL3 | 2.01 (1.63-2.49) | $7.01 \times 10^{-11}$ | 2.42 (1.85- 3.16) | $8.76 \times 10^{-11}$ |
| TNC | 2.44 (1.87-3.20) | $7.70 \times 10^{-11}$ | 2.39 (1.81- 3.15) | $8.37 \times 10^{-10}$ |
| EFEMP1 | 2.58 (1.92-3.46) | $2.43 \times 10^{-10}$ | 2.32 (1.80- 3.00) | $1.24 \times 10^{-10}$ |
| TNNI3 | 1.59 (1.38-1.84) | $3.78 \times 10^{-10}$ | 1.72 (1.44- 2.04) | $9.99 \times 10^{-10}$ |
| PXDN | 1.48 (1.31-1.68) | $4.82 \times 10^{-10}$ | 1.57 (1.33- 1.85) | $5.75 \times 10^{-8}$ |
| SEMA6B | 1.97 (1.59-2.44) | $4.83 \times 10^{-10}$ | 1.70 (1.40- 2.06) | $5.97 \times 10^{-8}$ |
| MMP19 | 2.12 (1.67-2.69) | $6.31 \times 10^{-10}$ | 3.00 (2.33- 3.87) | $2.59 \times 10^{-17}$ |
| ENG | 1.69 (1.43-2.00) | $8.36 \times 10^{-10}$ | 2.00 (1.50- 2.68) | $2.94 \times 10^{-6}$ |
| ATP5PF | 1.57 (1.36-1.82) | $1.45 \times 10^{-9}$ | 1.60 (1.36- 1.88) | $2.22 \times 10^{-8}$ |
| TREM1 | 1.88 (1.53-2.31) | $1.51 \times 10^{-9}$ | 1.91 (1.46- 2.50) | $2.52 \times 10^{-6}$ |
| IL19 | 1.83 (1.50-2.22) | $1.77 \times 10^{-9}$ | 1.90 (1.50- 2.41) | $1.40 \times 10^{-7}$ |
| EPO | 1.48 (1.30-1.69) | $1.79 \times 10^{-9}$ | 1.90 (1.65- 2.19) | $4.50 \times 10^{-19}$ |
| COL28A1 | 1.92 (1.55-2.38) | $2.24 \times 10^{-9}$ | 2.52 (1.93- 3.27) | $6.29 \times 10^{-12}$ |
| MFAP4 | 1.70 (1.42-2.02) | $4.07 \times 10^{-9}$ | 1.81 (1.49- 2.21) | $3.17 \times 10^{-9}$ |
| RSPO4 | 1.54 (1.33-1.78) | $4.81 \times 10^{-9}$ | 2.03 (1.69- 2.43) | $2.82 \times 10^{-14}$ |
| NBL1 | 1.81 (1.48-2.20) | $5.21 \times 10^{-9}$ | 1.68 (1.37- 2.07) | $6.00 \times 10^{-7}$ |
| IGFBP1 | 1.47 (1.29-1.68) | $6.16 \times 10^{-9}$ | 1.45 (1.25- 1.67) | $4.63 \times 10^{-7}$ |
| PTK7 | 2.03 (1.60-2.58) | $6.54 \times 10^{-9}$ | 3.11 (2.28- 4.23) | $6.80 \times 10^{-13}$ |
| BOC | 1.73 (1.43-2.08) | $7.73 \times 10^{-9}$ | 1.69 (1.31- 2.17) | $4.57 \times 10^{-5}$ |
| CD5 | 1.73 (1.44-2.09) | $8.16 \times 10^{-9}$ | 1.96 (1.62- 2.36) | $2.02 \times 10^{-12}$ |
| FABP3 | 1.55 (1.34-1.80) | $8.59 \times 10^{-9}$ | 1.69 (1.39- 2.05) | $1.60 \times 10^{-7}$ |

| | | | | |
|---|---|---|---|---|
| PTPRU | 1.67 (1.40-1.99) | $9.98 \times 10^{-9}$ | 2.30 (1.81- 2.94) | $1.84 \times 10^{-11}$ |
| C1QTNF1 | 3.04 (2.07-4.45) | $1.26 \times 10^{-8}$ | 5.10 (3.24- 8.03) | $2.07 \times 10^{-12}$ |
| KERA | 1.79 (1.46-2.19) | $1.43 \times 10^{-8}$ | 1.86 (1.52- 2.28) | $2.83 \times 10^{-9}$ |
| ADM | 1.76 (1.45-2.15) | $1.43 \times 10^{-8}$ | 1.53 (1.24- 1.90) | $9.73 \times 10^{-5}$ |
| CTSH | 2.46 (1.80-3.37) | $1.87 \times 10^{-8}$ | 4.79 (3.17- 7.24) | $1.16 \times 10^{-13}$ |
| EPHA2 | 1.84 (1.48-2.27) | $2.15 \times 10^{-8}$ | 2.08 (1.59- 2.73) | $9.24 \times 10^{-8}$ |
| RNASE1 | 1.44 (1.27-1.64) | $2.16 \times 10^{-8}$ | 1.62 (1.38- 1.91) | $5.73 \times 10^{-9}$ |
| TFF3 | 1.59 (1.35-1.87) | $2.63 \times 10^{-8}$ | 2.16 (1.73- 2.71) | $1.48 \times 10^{-11}$ |
| STC1 | 1.72 (1.42-2.09) | $4.70 \times 10^{-8}$ | 2.32 (1.77- 3.04) | $1.06 \times 10^{-9}$ |
| TAGLN | 1.61 (1.35-1.91) | $7.43 \times 10^{-8}$ | 2.08 (1.67- 2.60) | $1.01 \times 10^{-10}$ |
| CXCL13 | 1.40 (1.24-1.59) | $8.08 \times 10^{-8}$ | 1.40 (1.19- 1.66) | $6.87 \times 10^{-5}$ |
| WFDC2 | 1.75 (1.42-2.15) | $1.01 \times 10^{-7}$ | 2.39 (1.84- 3.11) | $6.03 \times 10^{-11}$ |
| DCN | 2.08 (1.59-2.73) | $1.06 \times 10^{-7}$ | 2.53 (1.72- 3.72) | $2.39 \times 10^{-6}$ |
| RNASE6 | 1.57 (1.33-1.85) | $1.31 \times 10^{-7}$ | 1.60 (1.34- 1.90) | $9.97 \times 10^{-8}$ |
| IGSF3 | 1.89 (1.49-2.40) | $1.55 \times 10^{-7}$ | 1.98 (1.47- 2.67) | $6.42 \times 10^{-6}$ |
| ICAM5 | 1.69 (1.39-2.07) | $2.23 \times 10^{-7}$ | 2.36 (1.84- 3.01) | $8.10 \times 10^{-12}$ |
| CHST15 | 2.03 (1.55-2.65) | $2.46 \times 10^{-7}$ | 2.97 (2.17- 4.05) | $7.60 \times 10^{-12}$ |
| CILP2 | 0.60 (0.50-0.73) | $3.62 \times 10^{-7}$ | 0.47 (0.36- 0.61) | $1.40 \times 10^{-8}$ |
| RELT | 1.85 (1.46-2.34) | $3.92 \times 10^{-7}$ | 1.74 (1.39- 2.19) | $1.78 \times 10^{-6}$ |
| Igh | 1.56 (1.31-1.85) | $7.29 \times 10^{-7}$ | 1.69 (1.38- 2.07) | $3.25 \times 10^{-7}$ |
| FABP4 | 1.43 (1.24-1.65) | $7.71 \times 10^{-7}$ | 1.39 (1.18- 1.63) | $8.38 \times 10^{-5}$ |
| MMP7 | 1.56 (1.30-1.86) | $1.04 \times 10^{-6}$ | 1.85 (1.52- 2.25) | $6.96 \times 10^{-10}$ |
| COLEC11 | 1.42 (1.23-1.64) | $1.12 \times 10^{-6}$ | 1.55 (1.29- 1.85) | $2.11 \times 10^{-6}$ |
| CCL14 | 1.75 (1.40-2.19) | $1.15 \times 10^{-6}$ | 2.22 (1.68- 2.93) | $2.25 \times 10^{-8}$ |
| B3GAT3 | 1.83 (1.44-2.34) | $1.24 \times 10^{-6}$ | 2.01 (1.44- 2.80) | $3.57 \times 10^{-5}$ |
| TNFRSF1A | 1.77 (1.40-2.24) | $1.40 \times 10^{-6}$ | 1.89 (1.43- 2.49) | $7.38 \times 10^{-6}$ |
| MMP2 | 1.71 (1.37-2.13) | $2.13 \times 10^{-6}$ | 2.25 (1.65- 3.08) | $3.89 \times 10^{-7}$ |
| UMOD | 2.19 (1.58-3.04) | $2.34 \times 10^{-6}$ | 2.62 (1.84- 3.73) | $8.18 \times 10^{-8}$ |
| FCN3 | 0.64 (0.53-0.77) | $2.86 \times 10^{-6}$ | 0.66 (0.54- 0.80) | $3.38 \times 10^{-5}$ |
| TFRC | 1.56 (1.29-1.87) | $3.12 \times 10^{-6}$ | 2.14 (1.70- 2.69) | $6.99 \times 10^{-11}$ |
| KLK10 | 1.51 (1.26-1.80) | $4.50 \times 10^{-6}$ | 1.60 (1.26- 2.04) | $1.11 \times 10^{-4}$ |
| LAMC2 | 1.52 (1.27-1.81) | $4.86 \times 10^{-6}$ | 1.64 (1.38- 1.95) | $2.74 \times 10^{-8}$ |
| KREMEN1 | 1.83 (1.41-2.38) | $5.18 \times 10^{-6}$ | 1.85 (1.39- 2.46) | $2.45 \times 10^{-5}$ |
| CTHRC1 | 1.85 (1.42-2.42) | $5.29 \times 10^{-6}$ | 2.27 (1.62- 3.19) | $2.03 \times 10^{-6}$ |
| PRSS22 | 1.56 (1.29-1.89) | $5.35 \times 10^{-6}$ | 1.98 (1.50- 2.61) | $1.34 \times 10^{-6}$ |
| FGF23 | 1.31 (1.16-1.47) | $5.63 \times 10^{-6}$ | 1.84 (1.58- 2.15) | $4.72 \times 10^{-15}$ |
| WFDC1 | 1.61 (1.31-1.98) | $5.86 \times 10^{-6}$ | 2.79 (2.02- 3.86) | $4.84 \times 10^{-10}$ |
| LUM | 2.12 (1.53-2.93) | $6.00 \times 10^{-6}$ | 3.30 (2.28- 4.76) | $1.81 \times 10^{-10}$ |
| GABARAPL1 | 1.55 (1.28-1.87) | $6.37 \times 10^{-6}$ | 1.94 (1.48- 2.54) | $1.65 \times 10^{-6}$ |
| TNFRSF11B | 1.61 (1.31-1.99) | $6.79 \times 10^{-6}$ | 2.26 (1.68- 3.04) | $7.39 \times 10^{-8}$ |
| RBL2 | 1.83 (1.41-2.39) | $6.89 \times 10^{-6}$ | 2.99 (2.16- 4.14) | $3.94 \times 10^{-11}$ |
| WARS1 | 1.53 (1.27-1.85) | $7.58 \times 10^{-6}$ | 1.73 (1.41- 2.13) | $1.60 \times 10^{-7}$ |

| | | | | |
|---|---|---|---|---|
| PLXDC2 | 1.67 (1.33-2.09) | $7.68 \times 10^{-6}$ | 3.83 (2.67- 5.50) | $3.33 \times 10^{-13}$ |
| PARVA | 1.68 (1.34-2.12) | $8.14 \times 10^{-6}$ | 1.62 (1.32- 1.99) | $4.93 \times 10^{-6}$ |
| CHRDL2 | 1.48 (1.25-1.76) | $8.78 \times 10^{-6}$ | 1.83 (1.45- 2.32) | $3.18 \times 10^{-7}$ |
| DNAJB12 | 1.55 (1.28-1.88) | $8.90 \times 10^{-6}$ | 1.80 (1.42- 2.29) | $1.21 \times 10^{-6}$ |
| CNDP1 | 0.67 (0.56-0.80) | $9.55 \times 10^{-6}$ | 0.63 (0.51- 0.78) | $1.43 \times 10^{-5}$ |
| TST | 1.39 (1.20-1.62) | $1.17 \times 10^{-5}$ | 1.73 (1.40- 2.14) | $3.98 \times 10^{-7}$ |
| GAS1 | 2.86 (1.77-4.64) | $1.99 \times 10^{-5}$ | 2.04 (1.45- 2.89) | $5.26 \times 10^{-5}$ |
| AGRP | 1.45 (1.22-1.71) | $2.12 \times 10^{-5}$ | 1.84 (1.46- 2.32) | $2.28 \times 10^{-7}$ |
| CD163 | 1.52 (1.25-1.85) | $2.20 \times 10^{-5}$ | 1.75 (1.35- 2.29) | $3.05 \times 10^{-5}$ |
| TNFRSF1B | 1.56 (1.27-1.91) | $2.42 \times 10^{-5}$ | 1.92 (1.48- 2.49) | $8.90 \times 10^{-7}$ |
| CHST9 | 1.41 (1.20-1.65) | $3.16 \times 10^{-5}$ | 1.47 (1.26- 1.71) | $1.21 \times 10^{-6}$ |
| COL18A1 | 1.72 (1.33-2.23) | $3.17 \times 10^{-5}$ | 2.62 (1.87- 3.68) | $2.74 \times 10^{-8}$ |
| FCGR3B | 1.55 (1.26-1.90) | $3.53 \times 10^{-5}$ | 2.07 (1.55- 2.76) | $9.17 \times 10^{-7}$ |
| SCARF2 | 1.76 (1.34-2.30) | $3.66 \times 10^{-5}$ | 2.36 (1.61- 3.45) | $9.84 \times 10^{-6}$ |
| PPP4R3A | 1.42 (1.20-1.69) | $3.96 \times 10^{-5}$ | 1.64 (1.29- 2.07) | $4.76 \times 10^{-5}$ |
| GRN | 1.78 (1.35-2.34) | $4.25 \times 10^{-5}$ | 3.16 (2.18- 4.58) | $1.33 \times 10^{-9}$ |
| CD93 | 1.68 (1.31-2.16) | $4.77 \times 10^{-5}$ | 2.32 (1.73- 3.10) | $1.59 \times 10^{-8}$ |
| TFF2 | 1.72 (1.33-2.24) | $5.01 \times 10^{-5}$ | 1.86 (1.51- 2.30) | $5.85 \times 10^{-9}$ |
| NRP1 | 1.64 (1.29-2.08) | $5.14 \times 10^{-5}$ | 2.57 (1.82- 3.64) | $1.04 \times 10^{-7}$ |
| IGFBP2 | 1.34 (1.16-1.55) | $5.86 \times 10^{-5}$ | 1.46 (1.22- 1.76) | $5.34 \times 10^{-5}$ |
| HTRA2 | 1.66 (1.29-2.12) | $6.07 \times 10^{-5}$ | 1.90 (1.57- 2.31) | $6.04 \times 10^{-11}$ |
| CTSB | 1.37 (1.18-1.61) | $6.11 \times 10^{-5}$ | 1.78 (1.44- 2.20) | $1.03 \times 10^{-7}$ |
| IBSP | 1.35 (1.16-1.56) | $7.51 \times 10^{-5}$ | 1.63 (1.34- 1.99) | $1.27 \times 10^{-6}$ |
| RNASE4 | 1.60 (1.27-2.01) | $8.05 \times 10^{-5}$ | 2.60 (1.77- 3.80) | $9.63 \times 10^{-7}$ |
| MRC2 | 1.52 (1.23-1.87) | $9.82 \times 10^{-5}$ | 1.84 (1.38- 2.45) | $3.24 \times 10^{-5}$ |
| ROBO2 | 1.70 (1.30-2.23) | $1.10 \times 10^{-4}$ | 2.69 (1.90- 3.80) | $2.23 \times 10^{-8}$ |
| BMP6 | 1.30 (1.13-1.48) | $1.34 \times 10^{-4}$ | 1.65 (1.38- 1.97) | $2.72 \times 10^{-8}$ |
| GNPTG | 1.64 (1.27-2.12) | $1.49 \times 10^{-4}$ | 6.02 (3.54-10.23) | $3.29 \times 10^{-11}$ |
| SIGLEC7 | 1.57 (1.24-1.97) | $1.50 \times 10^{-4}$ | 2.15 (1.65- 2.79) | $1.33 \times 10^{-8}$ |
| FSTL1 | 1.65 (1.27-2.15) | $1.95 \times 10^{-4}$ | 2.83 (2.05- 3.90) | $2.71 \times 10^{-10}$ |
| MENT | 0.70 (0.58-0.85) | $1.97 \times 10^{-4}$ | 0.50 (0.36- 0.68) | $1.40 \times 10^{-5}$ |
| FLRT3 | 1.53 (1.22-1.92) | $2.00 \times 10^{-4}$ | 1.76 (1.34- 2.31) | $4.40 \times 10^{-5}$ |
| ADAM12 | 1.59 (1.25-2.04) | $2.04 \times 10^{-4}$ | 2.04 (1.54- 2.72) | $8.31 \times 10^{-7}$ |
| IL6ST | 1.66 (1.27-2.17) | $2.14 \times 10^{-4}$ | 2.29 (1.60- 3.28) | $6.70 \times 10^{-6}$ |
| IL18R1 | 1.41 (1.17-1.69) | $2.46 \times 10^{-4}$ | 1.79 (1.35- 2.37) | $4.64 \times 10^{-5}$ |
| INHBB | 1.49 (1.20-1.86) | $2.72 \times 10^{-4}$ | 2.01 (1.53- 2.65) | $5.77 \times 10^{-7}$ |
| AMIGO2 | 1.81 (1.31-2.49) | $2.82 \times 10^{-4}$ | 2.90 (1.91- 4.41) | $6.16 \times 10^{-7}$ |
| SEMA6A | 2.58 (1.54-4.32) | $3.02 \times 10^{-4}$ | 2.83 (1.76- 4.54) | $1.62 \times 10^{-5}$ |
| GREM2 | 1.47 (1.19-1.81) | $3.10 \times 10^{-4}$ | 1.85 (1.48- 2.32) | $7.61 \times 10^{-8}$ |
| SFRP1 | 1.33 (1.14-1.56) | $3.54 \times 10^{-4}$ | 1.49 (1.26- 1.75) | $1.63 \times 10^{-6}$ |
| FGF7 | 1.47 (1.19-1.82) | $3.73 \times 10^{-4}$ | 1.97 (1.51- 2.58) | $7.82 \times 10^{-7}$ |
| NTF3 | 1.40 (1.16-1.68) | $3.79 \times 10^{-4}$ | 1.75 (1.33- 2.31) | $7.85 \times 10^{-5}$ |

| | | | | |
|---|---|---|---|---|
| CDH7 | 0.66 (0.53-0.83) | $4.33 \times 10^{-4}$ | 0.50 (0.35- 0.70) | $5.31 \times 10^{-5}$ |
| PIGR | 1.27 (1.11-1.45) | $4.41 \times 10^{-4}$ | 1.56 (1.30- 1.88) | $1.98 \times 10^{-6}$ |
| IGF1R | 1.51 (1.20-1.90) | $4.95 \times 10^{-4}$ | 2.23 (1.59- 3.12) | $3.21 \times 10^{-6}$ |
| VEGFD | 1.29 (1.12-1.48) | $5.02 \times 10^{-4}$ | 1.84 (1.57- 2.16) | $7.18 \times 10^{-14}$ |
| NOTUM | 0.77 (0.66-0.89) | $5.30 \times 10^{-4}$ | 0.56 (0.45- 0.69) | $1.89 \times 10^{-7}$ |
| IL6 | 1.35 (1.14-1.60) | $5.32 \times 10^{-4}$ | 1.44 (1.20- 1.74) | $1.06 \times 10^{-4}$ |
| CD300C | 1.46 (1.18-1.81) | $5.51 \times 10^{-4}$ | 1.86 (1.38- 2.49) | $3.59 \times 10^{-5}$ |
| CRELD1 | 1.46 (1.18-1.82) | $5.65 \times 10^{-4}$ | 1.75 (1.33- 2.31) | $6.79 \times 10^{-5}$ |
| SCARA5 | 1.52 (1.20-1.93) | $5.82 \times 10^{-4}$ | 2.19 (1.79- 2.68) | $1.43 \times 10^{-14}$ |
| PLA2G2A | 1.23 (1.09-1.38) | $6.37 \times 10^{-4}$ | 1.50 (1.31- 1.72) | $9.31 \times 10^{-9}$ |
| PSMD7 | 1.38 (1.15-1.66) | $6.65 \times 10^{-4}$ | 0.36 (0.22- 0.57) | $1.52 \times 10^{-5}$ |
| ANGPTL1 | 1.62 (1.22-2.14) | $7.23 \times 10^{-4}$ | 2.16 (1.51- 3.08) | $2.44 \times 10^{-5}$ |
| NELL2 | 1.67 (1.24-2.26) | $7.89 \times 10^{-4}$ | 1.76 (1.36- 2.27) | $1.95 \times 10^{-5}$ |
| OLFM1 | 1.35 (1.13-1.62) | $8.81 \times 10^{-4}$ | 1.66 (1.34- 2.04) | $2.12 \times 10^{-6}$ |
| ESM1 | 1.67 (1.23-2.26) | $8.85 \times 10^{-4}$ | 1.54 (1.24- 1.91) | $7.88 \times 10^{-5}$ |
| MMP12 | 1.25 (1.09-1.43) | $9.77 \times 10^{-4}$ | 1.55 (1.31- 1.83) | $3.21 \times 10^{-7}$ |
| CHRDL1 | 1.47 (1.16-1.85) | $1.14 \times 10^{-3}$ | 2.35 (1.71- 3.25) | $1.82 \times 10^{-7}$ |
| UNC5C | 1.50 (1.17-1.91) | $1.15 \times 10^{-3}$ | 2.41 (1.75- 3.31) | $6.10 \times 10^{-8}$ |
| CCN1 | 1.24 (1.09-1.42) | $1.15 \times 10^{-3}$ | 1.65 (1.40- 1.95) | $2.76 \times 10^{-9}$ |
| SCARF1 | 1.48 (1.17-1.89) | $1.21 \times 10^{-3}$ | 2.10 (1.52- 2.89) | $5.51 \times 10^{-6}$ |
| CCDC80 | 1.53 (1.18-1.99) | $1.37 \times 10^{-3}$ | 2.26 (1.67- 3.06) | $1.20 \times 10^{-7}$ |
| CILP | 1.27 (1.10-1.47) | $1.42 \times 10^{-3}$ | 1.43 (1.20- 1.71) | $6.72 \times 10^{-5}$ |
| EPHA10 | 1.29 (1.10-1.51) | $1.43 \times 10^{-3}$ | 1.52 (1.24- 1.86) | $5.48 \times 10^{-5}$ |
| BMP7 | 1.35 (1.12-1.63) | $1.46 \times 10^{-3}$ | 1.72 (1.31- 2.24) | $7.17 \times 10^{-5}$ |
| TGFB3 | 1.65 (1.21-2.24) | $1.46 \times 10^{-3}$ | 1.97 (1.46- 2.65) | $9.40 \times 10^{-6}$ |
| UNC5B | 1.69 (1.22-2.34) | $1.50 \times 10^{-3}$ | 1.93 (1.51- 2.46) | $1.15 \times 10^{-7}$ |
| DNASE1L2 | 1.28 (1.10-1.50) | $1.56 \times 10^{-3}$ | 1.86 (1.49- 2.32) | $5.56 \times 10^{-8}$ |
| EBI3 | 1.24 (1.09-1.43) | $1.66 \times 10^{-3}$ | 1.79 (1.46- 2.18) | $1.16 \times 10^{-8}$ |
| SLPI | 1.44 (1.15-1.81) | $1.67 \times 10^{-3}$ | 1.82 (1.35- 2.46) | $7.51 \times 10^{-5}$ |
| BCHE | 0.63 (0.47-0.84) | $1.76 \times 10^{-3}$ | 0.43 (0.30- 0.63) | $1.32 \times 10^{-5}$ |
| PCDHGA10 | 1.25 (1.09-1.45) | $1.83 \times 10^{-3}$ | 1.60 (1.38- 1.85) | $3.51 \times 10^{-10}$ |
| NRP2 | 1.48 (1.15-1.89) | $1.95 \times 10^{-3}$ | 2.24 (1.64- 3.06) | $3.70 \times 10^{-7}$ |
| ITGA1\|ITGB1 | 1.35 (1.12-1.64) | $2.04 \times 10^{-3}$ | 1.73 (1.38- 2.18) | $3.10 \times 10^{-6}$ |
| CCL21 | 1.33 (1.11-1.60) | $2.30 \times 10^{-3}$ | 1.88 (1.48- 2.37) | $1.40 \times 10^{-7}$ |
| MINPP1 | 1.67 (1.20-2.33) | $2.37 \times 10^{-3}$ | 3.21 (2.21- 4.66) | $8.80 \times 10^{-10}$ |
| PLA2R1 | 1.35 (1.11-1.64) | $2.57 \times 10^{-3}$ | 2.31 (1.78- 3.01) | $4.85 \times 10^{-10}$ |
| CERT1 | 1.23 (1.08-1.42) | $2.67 \times 10^{-3}$ | 1.51 (1.28- 1.80) | $1.90 \times 10^{-6}$ |
| HSPA1A | 1.28 (1.09-1.50) | $2.73 \times 10^{-3}$ | 1.95 (1.52- 2.51) | $1.49 \times 10^{-7}$ |
| TXNDC5 | 1.44 (1.13-1.82) | $2.77 \times 10^{-3}$ | 2.31 (1.68- 3.17) | $2.77 \times 10^{-7}$ |
| APOF | 1.31 (1.09-1.56) | $3.16 \times 10^{-3}$ | 1.66 (1.32- 2.10) | $1.96 \times 10^{-5}$ |
| C1QL1 | 1.65 (1.18-2.31) | $3.44 \times 10^{-3}$ | 2.22 (1.54- 3.19) | $1.68 \times 10^{-5}$ |
| TMED10 | 1.46 (1.13-1.89) | $3.50 \times 10^{-3}$ | 2.66 (1.92- 3.68) | $4.58 \times 10^{-9}$ |

| | | | | |
|---|---|---|---|---|
| MATN2 | 1.50 (1.14-1.97) | $3.62 \times 10^{-3}$ | 2.04 (1.43- 2.89) | $6.98 \times 10^{-5}$ |
| LRRC32 | 1.38 (1.11-1.71) | $3.75 \times 10^{-3}$ | 2.45 (1.69- 3.56) | $2.59 \times 10^{-6}$ |
| EFNA4 | 1.46 (1.13-1.89) | $3.85 \times 10^{-3}$ | 1.93 (1.46- 2.55) | $3.40 \times 10^{-6}$ |
| FJX1 | 1.46 (1.13-1.89) | $3.95 \times 10^{-3}$ | 1.73 (1.41- 2.12) | $1.79 \times 10^{-7}$ |

**Supplemental Table 4: Proteins Significantly Associated With Heart Failure Hospitalization or Cardiovascular Death in Minimally Adjusted Model After Rank-Based Inverse Normal Transformation**

| | Discovery (ATMOSPHERE) | | Replication (PARADIGM) | |
|---|---|---|---|---|
| Protein | HR per SD (95% CI) | *P* Value | HR per SD (95% CI) | *P* Value |
| GDF15 | 1.58 (1.43-1.75) | $5.76 \times 10^{-19}$ | 1.66 (1.44-1.90) | $5.06 \times 10^{-13}$ |
| SVEP1 | 1.54 (1.40-1.69) | $9.89 \times 10^{-19}$ | 1.64 (1.44-1.87) | $7.96 \times 10^{-14}$ |
| ANGPT2 | 1.51 (1.38-1.66) | $8.16 \times 10^{-18}$ | 1.77 (1.55-2.02) | $1.94 \times 10^{-17}$ |
| NPPB | 1.53 (1.39-1.69) | $1.83 \times 10^{-17}$ | 1.59 (1.40-1.79) | $7.65 \times 10^{-14}$ |
| QSOX1 | 1.46 (1.33-1.60) | $4.05 \times 10^{-15}$ | 1.52 (1.35-1.71) | $3.17 \times 10^{-12}$ |
| THBS2 | 1.44 (1.31-1.58) | $6.10 \times 10^{-14}$ | 1.70 (1.50-1.94) | $7.46 \times 10^{-16}$ |
| ADAMTSL2 | 1.41 (1.29-1.55) | $3.26 \times 10^{-13}$ | 1.54 (1.36-1.73) | $3.31 \times 10^{-12}$ |
| LTBP4 | 1.43 (1.29-1.58) | $2.67 \times 10^{-12}$ | 1.45 (1.28-1.64) | $4.40 \times 10^{-9}$ |
| COL6A3 | 1.39 (1.26-1.53) | $1.61 \times 10^{-11}$ | 1.52 (1.33-1.73) | $4.07 \times 10^{-10}$ |
| PXDN | 1.39 (1.26-1.53) | $1.66 \times 10^{-11}$ | 1.44 (1.26-1.65) | $9.89 \times 10^{-8}$ |
| CST3 | 1.39 (1.26-1.53) | $1.72 \times 10^{-11}$ | 1.51 (1.32-1.73) | $4.87 \times 10^{-9}$ |
| SPON2 | 1.39 (1.26-1.54) | $2.35 \times 10^{-11}$ | 1.50 (1.31-1.72) | $4.86 \times 10^{-9}$ |
| NFASC | 1.38 (1.25-1.51) | $4.20 \times 10^{-11}$ | 1.54 (1.36-1.76) | $5.55 \times 10^{-11}$ |
| SPON1 | 1.39 (1.26-1.53) | $6.11 \times 10^{-11}$ | 1.67 (1.46-1.91) | $1.07 \times 10^{-13}$ |
| RSPO4 | 1.37 (1.25-1.50) | $7.59 \times 10^{-11}$ | 1.61 (1.42-1.82) | $7.26 \times 10^{-14}$ |
| FSTL3 | 1.39 (1.26-1.54) | $9.85 \times 10^{-11}$ | 1.53 (1.33-1.76) | $2.91 \times 10^{-9}$ |
| IL1RL1 | 1.39 (1.26-1.53) | $1.08 \times 10^{-10}$ | 1.45 (1.28-1.65) | $1.37 \times 10^{-8}$ |
| B2M | 1.38 (1.25-1.52) | $1.10 \times 10^{-10}$ | 1.42 (1.23-1.63) | $9.70 \times 10^{-7}$ |
| TNC | 1.36 (1.24-1.50) | $2.19 \times 10^{-10}$ | 1.46 (1.29-1.65) | $6.62 \times 10^{-10}$ |
| IGFBP7 | 1.37 (1.24-1.50) | $2.42 \times 10^{-10}$ | 1.62 (1.43-1.84) | $5.87 \times 10^{-14}$ |
| MMP19 | 1.35 (1.23-1.49) | $7.68 \times 10^{-10}$ | 1.67 (1.47-1.90) | $1.81 \times 10^{-15}$ |
| EFEMP1 | 1.35 (1.23-1.49) | $8.04 \times 10^{-10}$ | 1.48 (1.30-1.67) | $8.22 \times 10^{-10}$ |
| ATP5PF | 1.33 (1.21-1.45) | $8.95 \times 10^{-10}$ | 1.40 (1.25-1.58) | $1.02 \times 10^{-8}$ |
| SEMA6B | 1.33 (1.21-1.45) | $2.80 \times 10^{-9}$ | 1.40 (1.24-1.58) | $6.83 \times 10^{-8}$ |
| IL19 | 1.33 (1.21-1.46) | $3.78 \times 10^{-9}$ | 1.38 (1.21-1.56) | $8.53 \times 10^{-7}$ |
| COL28A1 | 1.34 (1.21-1.47) | $3.80 \times 10^{-9}$ | 1.55 (1.35-1.78) | $3.24 \times 10^{-10}$ |
| ENG | 1.32 (1.20-1.44) | $4.14 \times 10^{-9}$ | 1.34 (1.18-1.52) | $4.45 \times 10^{-6}$ |
| TREM1 | 1.34 (1.21-1.47) | $4.48 \times 10^{-9}$ | 1.35 (1.19-1.55) | $8.05 \times 10^{-6}$ |
| NBL1 | 1.35 (1.22-1.49) | $6.33 \times 10^{-9}$ | 1.35 (1.17-1.55) | $2.52 \times 10^{-5}$ |
| PTK7 | 1.32 (1.20-1.45) | $8.34 \times 10^{-9}$ | 1.55 (1.36-1.77) | $3.15 \times 10^{-11}$ |
| MFAP4 | 1.31 (1.20-1.44) | $9.07 \times 10^{-9}$ | 1.48 (1.30-1.69) | $4.62 \times 10^{-9}$ |
| TNNI3 | 1.31 (1.19-1.43) | $9.44 \times 10^{-9}$ | 1.46 (1.30-1.64) | $1.73 \times 10^{-10}$ |
| CTSH | 1.32 (1.20-1.45) | $9.89 \times 10^{-9}$ | 1.60 (1.40-1.84) | $9.62 \times 10^{-12}$ |
| FABP3 | 1.33 (1.21-1.47) | $1.00 \times 10^{-8}$ | 1.44 (1.26-1.65) | $1.30 \times 10^{-7}$ |
| PTPRU | 1.30 (1.18-1.42) | $1.70 \times 10^{-8}$ | 1.46 (1.29-1.65) | $1.21 \times 10^{-9}$ |
| C1QTNF1 | 1.30 (1.19-1.43) | $1.75 \times 10^{-8}$ | 1.54 (1.36-1.74) | $7.42 \times 10^{-12}$ |
| EPO | 1.31 (1.19-1.43) | $1.78 \times 10^{-8}$ | 1.65 (1.45-1.87) | $1.70 \times 10^{-14}$ |

| | | | | |
|---|---|---|---|---|
| RNASE1 | 1.32 (1.20-1.45) | $2.18 \times 10^{-8}$ | 1.48 (1.29-1.69) | $2.77 \times 10^{-8}$ |
| EPHA2 | 1.34 (1.21-1.48) | $2.76 \times 10^{-8}$ | 1.39 (1.21-1.59) | $3.84 \times 10^{-6}$ |
| BOC | 1.30 (1.18-1.42) | $3.11 \times 10^{-8}$ | 1.28 (1.13-1.44) | $7.91 \times 10^{-5}$ |
| TFF3 | 1.31 (1.19-1.45) | $3.64 \times 10^{-8}$ | 1.55 (1.35-1.77) | $4.14 \times 10^{-10}$ |
| STC1 | 1.30 (1.18-1.43) | $4.63 \times 10^{-8}$ | 1.47 (1.28-1.69) | $2.47 \times 10^{-8}$ |
| KERA | 1.30 (1.18-1.43) | $5.32 \times 10^{-8}$ | 1.42 (1.25-1.60) | $1.88 \times 10^{-8}$ |
| ADM | 1.30 (1.18-1.43) | $6.33 \times 10^{-8}$ | 1.30 (1.15-1.47) | $2.16 \times 10^{-5}$ |
| CXCL13 | 1.29 (1.18-1.42) | $6.41 \times 10^{-8}$ | 1.30 (1.15-1.46) | $2.31 \times 10^{-5}$ |
| TAGLN | 1.33 (1.20-1.48) | $7.33 \times 10^{-8}$ | 1.55 (1.34-1.79) | $3.05 \times 10^{-9}$ |
| WFDC2 | 1.30 (1.18-1.43) | $1.02 \times 10^{-7}$ | 1.55 (1.35-1.77) | $1.54 \times 10^{-10}$ |
| RNASE6 | 1.29 (1.18-1.42) | $1.38 \times 10^{-7}$ | 1.41 (1.24-1.60) | $2.06 \times 10^{-7}$ |
| CHST9 | 1.27 (1.16-1.39) | $1.65 \times 10^{-7}$ | 1.34 (1.19-1.51) | $1.24 \times 10^{-6}$ |
| IGFBP1 | 1.30 (1.18-1.44) | $1.81 \times 10^{-7}$ | 1.37 (1.20-1.56) | $2.63 \times 10^{-6}$ |
| FGF23 | 1.27 (1.16-1.39) | $1.85 \times 10^{-7}$ | 1.53 (1.36-1.73) | $5.69 \times 10^{-12}$ |
| TIMP2 | 1.27 (1.16-1.39) | $2.04 \times 10^{-7}$ | 1.27 (1.13-1.42) | $8.24 \times 10^{-5}$ |
| RELT | 1.28 (1.17-1.41) | $2.32 \times 10^{-7}$ | 1.35 (1.19-1.54) | $5.61 \times 10^{-6}$ |
| DCN | 1.28 (1.17-1.41) | $2.69 \times 10^{-7}$ | 1.33 (1.18-1.50) | $4.64 \times 10^{-6}$ |
| CD5 | 1.28 (1.16-1.41) | $3.00 \times 10^{-7}$ | 1.54 (1.35-1.75) | $5.15 \times 10^{-11}$ |
| SOD3 | 1.28 (1.16-1.40) | $3.21 \times 10^{-7}$ | 1.37 (1.21-1.54) | $2.94 \times 10^{-7}$ |
| IGSF3 | 1.27 (1.16-1.39) | $4.02 \times 10^{-7}$ | 1.30 (1.15-1.47) | $2.46 \times 10^{-5}$ |
| CILP2 | 0.79 (0.72-0.86) | $4.06 \times 10^{-7}$ | 0.70 (0.62-0.79) | $1.23 \times 10^{-8}$ |
| ICAM5 | 1.27 (1.16-1.39) | $4.10 \times 10^{-7}$ | 1.55 (1.37-1.75) | $6.12 \times 10^{-12}$ |
| CHST15 | 1.29 (1.17-1.42) | $4.24 \times 10^{-7}$ | 1.52 (1.33-1.73) | $3.40 \times 10^{-10}$ |
| FABP4 | 1.28 (1.16-1.41) | $7.71 \times 10^{-7}$ | 1.30 (1.15-1.47) | $4.48 \times 10^{-5}$ |
| DNAJB12 | 1.26 (1.15-1.39) | $1.14 \times 10^{-6}$ | 1.37 (1.21-1.56) | $1.43 \times 10^{-6}$ |
| AGRP | 1.25 (1.14-1.37) | $1.24 \times 10^{-6}$ | 1.33 (1.18-1.50) | $3.13 \times 10^{-6}$ |
| CCL14 | 1.27 (1.15-1.40) | $1.33 \times 10^{-6}$ | 1.45 (1.27-1.65) | $2.94 \times 10^{-8}$ |
| SCARF2 | 1.27 (1.15-1.41) | $1.64 \times 10^{-6}$ | 1.31 (1.15-1.49) | $6.25 \times 10^{-5}$ |
| TFRC | 1.25 (1.14-1.37) | $1.86 \times 10^{-6}$ | 1.47 (1.30-1.67) | $3.03 \times 10^{-9}$ |
| GABARAPL1 | 1.27 (1.15-1.40) | $2.15 \times 10^{-6}$ | 1.38 (1.21-1.57) | $1.12 \times 10^{-6}$ |
| MMP2 | 1.26 (1.14-1.38) | $2.20 \times 10^{-6}$ | 1.39 (1.22-1.58) | $5.94 \times 10^{-7}$ |
| COLEC11 | 1.25 (1.14-1.38) | $2.34 \times 10^{-6}$ | 1.33 (1.17-1.51) | $8.87 \times 10^{-6}$ |
| PLXDC2 | 1.25 (1.14-1.37) | $2.62 \times 10^{-6}$ | 1.53 (1.35-1.73) | $4.35 \times 10^{-11}$ |
| CIRBP | 1.24 (1.14-1.36) | $2.85 \times 10^{-6}$ | 1.28 (1.14-1.45) | $3.77 \times 10^{-5}$ |
| PARVA | 1.25 (1.14-1.37) | $3.34 \times 10^{-6}$ | 1.34 (1.19-1.51) | $2.15 \times 10^{-6}$ |
| MMP7 | 1.26 (1.14-1.38) | $4.18 \times 10^{-6}$ | 1.45 (1.28-1.64) | $5.49 \times 10^{-9}$ |
| PCDHGA10 | 1.24 (1.13-1.36) | $5.00 \times 10^{-6}$ | 1.48 (1.31-1.67) | $4.07 \times 10^{-10}$ |
| EPHA10 | 1.23 (1.13-1.35) | $5.28 \times 10^{-6}$ | 1.37 (1.21-1.55) | $3.77 \times 10^{-7}$ |
| RBL2 | 1.23 (1.13-1.35) | $6.95 \times 10^{-6}$ | 1.47 (1.30-1.66) | $6.25 \times 10^{-10}$ |
| PPP4R3A | 1.24 (1.13-1.36) | $7.33 \times 10^{-6}$ | 1.31 (1.16-1.49) | $1.67 \times 10^{-5}$ |
| LAMC2 | 1.23 (1.12-1.35) | $7.48 \times 10^{-6}$ | 1.40 (1.24-1.58) | $5.44 \times 10^{-8}$ |
| WFDC1 | 1.23 (1.12-1.35) | $7.80 \times 10^{-6}$ | 1.50 (1.32-1.72) | $1.30 \times 10^{-9}$ |

| | | | | |
|---|---|---|---|---|
| TST | 1.23 (1.12-1.35) | $9.21 \times 10^{-6}$ | 1.37 (1.21-1.55) | $7.80 \times 10^{-7}$ |
| UMOD | 1.24 (1.13-1.36) | $1.14 \times 10^{-5}$ | 1.40 (1.24-1.59) | $1.13 \times 10^{-7}$ |
| Igh | 1.24 (1.13-1.37) | $1.15 \times 10^{-5}$ | 1.37 (1.21-1.54) | $4.62 \times 10^{-7}$ |
| B3GAT3 | 1.23 (1.12-1.34) | $1.18 \times 10^{-5}$ | 1.29 (1.14-1.45) | $3.53 \times 10^{-5}$ |
| PRSS22 | 1.23 (1.12-1.35) | $1.18 \times 10^{-5}$ | 1.35 (1.19-1.53) | $3.92 \times 10^{-6}$ |
| WARS1 | 1.24 (1.12-1.36) | $1.20 \times 10^{-5}$ | 1.38 (1.22-1.55) | $1.34 \times 10^{-7}$ |
| TNFRSF11B | 1.25 (1.13-1.38) | $1.22 \times 10^{-5}$ | 1.40 (1.22-1.61) | $1.82 \times 10^{-6}$ |
| PCDHGA12 | 1.22 (1.12-1.34) | $1.34 \times 10^{-5}$ | 1.56 (1.39-1.75) | $3.73 \times 10^{-14}$ |
| NOTUM | 0.82 (0.75-0.90) | $1.68 \times 10^{-5}$ | 0.72 (0.64-0.81) | $6.50 \times 10^{-8}$ |
| LUM | 1.23 (1.12-1.35) | $1.69 \times 10^{-5}$ | 1.45 (1.28-1.64) | $3.19 \times 10^{-9}$ |
| OLFM1 | 1.22 (1.11-1.33) | $1.81 \times 10^{-5}$ | 1.38 (1.23-1.56) | $1.14 \times 10^{-7}$ |
| CHRDL2 | 1.23 (1.12-1.35) | $2.25 \times 10^{-5}$ | 1.37 (1.20-1.56) | $3.71 \times 10^{-6}$ |
| COL18A1 | 1.23 (1.12-1.35) | $2.65 \times 10^{-5}$ | 1.44 (1.26-1.65) | $9.93 \times 10^{-8}$ |
| CCN1 | 1.21 (1.10-1.32) | $3.19 \times 10^{-5}$ | 1.50 (1.33-1.69) | $3.18 \times 10^{-11}$ |
| CD163 | 1.22 (1.11-1.33) | $3.59 \times 10^{-5}$ | 1.29 (1.14-1.47) | $7.64 \times 10^{-5}$ |
| TFF2 | 1.21 (1.10-1.32) | $3.80 \times 10^{-5}$ | 1.41 (1.25-1.60) | $3.21 \times 10^{-8}$ |
| IGFBP2 | 1.24 (1.12-1.37) | $3.87 \times 10^{-5}$ | 1.33 (1.16-1.53) | $5.30 \times 10^{-5}$ |
| TNFRSF1B | 1.23 (1.11-1.36) | $4.23 \times 10^{-5}$ | 1.36 (1.18-1.57) | $1.92 \times 10^{-5}$ |
| CD93 | 1.22 (1.11-1.33) | $4.24 \times 10^{-5}$ | 1.42 (1.25-1.62) | $4.37 \times 10^{-8}$ |
| HTRA2 | 1.21 (1.11-1.33) | $4.24 \times 10^{-5}$ | 1.50 (1.33-1.70) | $9.23 \times 10^{-11}$ |
| BID | 1.21 (1.10-1.32) | $4.55 \times 10^{-5}$ | 1.37 (1.21-1.54) | $3.43 \times 10^{-7}$ |
| CRP | 1.20 (1.10-1.32) | $6.08 \times 10^{-5}$ | 1.27 (1.13-1.43) | $9.26 \times 10^{-5}$ |
| GRN | 1.21 (1.10-1.33) | $6.09 \times 10^{-5}$ | 1.46 (1.28-1.66) | $1.03 \times 10^{-8}$ |
| MENT | 0.83 (0.76-0.91) | $6.28 \times 10^{-5}$ | 0.74 (0.66-0.84) | $4.79 \times 10^{-6}$ |
| RNASE4 | 1.21 (1.10-1.32) | $7.75 \times 10^{-5}$ | 1.39 (1.21-1.60) | $3.29 \times 10^{-6}$ |
| CLSTN3 | 1.20 (1.09-1.31) | $8.02 \times 10^{-5}$ | 1.51 (1.34-1.71) | $2.87 \times 10^{-11}$ |
| FCGR3B | 1.21 (1.10-1.32) | $8.08 \times 10^{-5}$ | 1.36 (1.19-1.54) | $3.29 \times 10^{-6}$ |
| FLRT3 | 1.20 (1.10-1.32) | $1.04 \times 10^{-4}$ | 1.30 (1.15-1.47) | $4.07 \times 10^{-5}$ |
| FGF7 | 1.19 (1.09-1.31) | $1.15 \times 10^{-4}$ | 1.36 (1.20-1.53) | $1.02 \times 10^{-6}$ |
| VEGFD | 1.21 (1.10-1.33) | $1.18 \times 10^{-4}$ | 1.61 (1.43-1.81) | $3.98 \times 10^{-15}$ |
| NRP1 | 1.20 (1.09-1.31) | $1.21 \times 10^{-4}$ | 1.37 (1.21-1.55) | $4.91 \times 10^{-7}$ |
| ROBO2 | 1.21 (1.10-1.33) | $1.26 \times 10^{-4}$ | 1.41 (1.24-1.60) | $1.63 \times 10^{-7}$ |
| CTSB | 1.21 (1.10-1.33) | $1.42 \times 10^{-4}$ | 1.40 (1.22-1.60) | $9.46 \times 10^{-7}$ |
| C1QL2 | 1.20 (1.09-1.31) | $1.43 \times 10^{-4}$ | 1.31 (1.16-1.48) | $1.39 \times 10^{-5}$ |
| BMP6 | 1.20 (1.09-1.32) | $1.45 \times 10^{-4}$ | 1.37 (1.21-1.55) | $7.49 \times 10^{-7}$ |
| MRC2 | 1.20 (1.09-1.31) | $1.52 \times 10^{-4}$ | 1.29 (1.14-1.46) | $5.26 \times 10^{-5}$ |
| SIGLEC7 | 1.20 (1.09-1.31) | $1.55 \times 10^{-4}$ | 1.42 (1.25-1.61) | $1.21 \times 10^{-7}$ |
| ENTPD1 | 1.19 (1.08-1.30) | $2.10 \times 10^{-4}$ | 1.30 (1.15-1.47) | $1.90 \times 10^{-5}$ |
| FSTL1 | 1.19 (1.09-1.31) | $2.37 \times 10^{-4}$ | 1.48 (1.30-1.68) | $2.95 \times 10^{-9}$ |
| TMED10 | 1.19 (1.08-1.31) | $2.58 \times 10^{-4}$ | 1.44 (1.26-1.64) | $1.10 \times 10^{-7}$ |
| GREM2 | 1.19 (1.08-1.30) | $2.58 \times 10^{-4}$ | 1.40 (1.23-1.59) | $3.62 \times 10^{-7}$ |
| CILP | 1.18 (1.08-1.29) | $2.66 \times 10^{-4}$ | 1.33 (1.18-1.50) | $2.93 \times 10^{-6}$ |

| | | | | |
|---|---|---|---|---|
| UBE2E1 | 1.18 (1.08-1.30) | $2.70 \times 10^{-4}$ | 1.29 (1.14-1.45) | $2.74 \times 10^{-5}$ |
| IBSP | 1.19 (1.08-1.31) | $2.73 \times 10^{-4}$ | 1.31 (1.15-1.49) | $4.33 \times 10^{-5}$ |
| EFNA4 | 1.19 (1.08-1.30) | $2.82 \times 10^{-4}$ | 1.31 (1.15-1.49) | $4.70 \times 10^{-5}$ |
| INHBB | 1.20 (1.09-1.32) | $2.86 \times 10^{-4}$ | 1.38 (1.20-1.57) | $2.97 \times 10^{-6}$ |
| PIGR | 1.19 (1.08-1.30) | $3.26 \times 10^{-4}$ | 1.33 (1.17-1.50) | $5.78 \times 10^{-6}$ |
| ALKAL2 | 1.17 (1.08-1.28) | $3.56 \times 10^{-4}$ | 1.31 (1.16-1.48) | $1.04 \times 10^{-5}$ |
| IL6ST | 1.19 (1.08-1.31) | $3.78 \times 10^{-4}$ | 1.34 (1.17-1.53) | $2.15 \times 10^{-5}$ |
| PAM | 1.19 (1.08-1.31) | $3.98 \times 10^{-4}$ | 1.35 (1.18-1.55) | $9.62 \times 10^{-6}$ |
| BCHE | 0.84 (0.77-0.93) | $4.14 \times 10^{-4}$ | 0.77 (0.68-0.87) | $2.60 \times 10^{-5}$ |
| CDH7 | 0.85 (0.78-0.93) | $4.46 \times 10^{-4}$ | 0.77 (0.68-0.87) | $3.67 \times 10^{-5}$ |
| IGF1R | 1.18 (1.08-1.29) | $4.63 \times 10^{-4}$ | 1.34 (1.18-1.52) | $4.76 \times 10^{-6}$ |
| CRELD1 | 1.18 (1.08-1.30) | $5.31 \times 10^{-4}$ | 1.30 (1.14-1.49) | $8.59 \times 10^{-5}$ |
| MYL6B | 1.19 (1.08-1.31) | $5.51 \times 10^{-4}$ | 1.30 (1.16-1.47) | $1.59 \times 10^{-5}$ |
| PSMD7 | 1.18 (1.07-1.29) | $5.68 \times 10^{-4}$ | 0.74 (0.65-0.85) | $2.43 \times 10^{-5}$ |
| TGFB3 | 1.17 (1.07-1.29) | $6.37 \times 10^{-4}$ | 1.44 (1.27-1.63) | $1.59 \times 10^{-8}$ |
| GNPTG | 1.17 (1.07-1.28) | $6.68 \times 10^{-4}$ | 1.45 (1.27-1.65) | $2.50 \times 10^{-8}$ |
| ASAH2 | 0.85 (0.78-0.94) | $7.11 \times 10^{-4}$ | 0.77 (0.68-0.87) | $3.59 \times 10^{-5}$ |
| ANGPTL1 | 1.18 (1.07-1.29) | $7.19 \times 10^{-4}$ | 1.31 (1.15-1.50) | $4.21 \times 10^{-5}$ |
| PLA2G2A | 1.18 (1.07-1.29) | $7.80 \times 10^{-4}$ | 1.40 (1.23-1.59) | $3.25 \times 10^{-7}$ |
| EBI3 | 1.17 (1.07-1.29) | $8.02 \times 10^{-4}$ | 1.48 (1.29-1.69) | $8.29 \times 10^{-9}$ |
| AMIGO2 | 1.18 (1.07-1.29) | $8.40 \times 10^{-4}$ | 1.36 (1.19-1.54) | $3.26 \times 10^{-6}$ |
| LRRC32 | 1.17 (1.07-1.29) | $8.74 \times 10^{-4}$ | 1.32 (1.16-1.51) | $4.15 \times 10^{-5}$ |
| SFRP1 | 1.18 (1.07-1.31) | $8.90 \times 10^{-4}$ | 1.30 (1.15-1.48) | $4.35 \times 10^{-5}$ |
| DNASE1L2 | 1.16 (1.06-1.27) | $9.22 \times 10^{-4}$ | 1.39 (1.23-1.57) | $8.75 \times 10^{-8}$ |
| CHRDL1 | 1.19 (1.07-1.32) | $1.06 \times 10^{-3}$ | 1.44 (1.23-1.67) | $3.45 \times 10^{-6}$ |
| CERT1 | 1.16 (1.06-1.27) | $1.08 \times 10^{-3}$ | 1.34 (1.19-1.52) | $1.79 \times 10^{-6}$ |
| AK2 | 1.16 (1.06-1.27) | $1.08 \times 10^{-3}$ | 1.33 (1.18-1.50) | $4.55 \times 10^{-6}$ |
| RRM2B | 1.16 (1.06-1.27) | $1.10 \times 10^{-3}$ | 1.32 (1.17-1.49) | $3.20 \times 10^{-6}$ |
| PSMB1 | 1.16 (1.06-1.27) | $1.14 \times 10^{-3}$ | 1.36 (1.21-1.53) | $6.25 \times 10^{-7}$ |
| SEMA6A | 1.17 (1.06-1.29) | $1.20 \times 10^{-3}$ | 1.31 (1.16-1.48) | $2.07 \times 10^{-5}$ |
| APOF | 1.18 (1.07-1.31) | $1.21 \times 10^{-3}$ | 1.37 (1.19-1.58) | $1.94 \times 10^{-5}$ |
| SCARF1 | 1.16 (1.06-1.27) | $1.22 \times 10^{-3}$ | 1.34 (1.18-1.52) | $3.53 \times 10^{-6}$ |
| MDM1 | 1.17 (1.06-1.28) | $1.27 \times 10^{-3}$ | 1.38 (1.22-1.56) | $1.48 \times 10^{-7}$ |
| PLA2R1 | 1.16 (1.06-1.27) | $1.29 \times 10^{-3}$ | 1.46 (1.29-1.64) | $1.07 \times 10^{-9}$ |
| GRPEL1 | 1.16 (1.06-1.28) | $1.30 \times 10^{-3}$ | 1.27 (1.13-1.43) | $6.09 \times 10^{-5}$ |
| IL18 | 1.15 (1.06-1.26) | $1.31 \times 10^{-3}$ | 1.40 (1.24-1.57) | $2.22 \times 10^{-8}$ |
| HSPA1A | 1.16 (1.06-1.26) | $1.35 \times 10^{-3}$ | 1.38 (1.22-1.56) | $1.79 \times 10^{-7}$ |
| MMP12 | 1.17 (1.06-1.29) | $1.43 \times 10^{-3}$ | 1.38 (1.22-1.57) | $4.93 \times 10^{-7}$ |
| RSPO3 | 1.17 (1.06-1.29) | $1.44 \times 10^{-3}$ | 1.30 (1.15-1.47) | $2.49 \times 10^{-5}$ |
| UNC5C | 1.18 (1.06-1.30) | $1.46 \times 10^{-3}$ | 1.44 (1.24-1.66) | $7.56 \times 10^{-7}$ |
| ARHGAP1 | 1.17 (1.06-1.28) | $1.46 \times 10^{-3}$ | 1.31 (1.15-1.48) | $2.54 \times 10^{-5}$ |
| IL6 | 1.16 (1.06-1.27) | $1.51 \times 10^{-3}$ | 1.28 (1.13-1.44) | $9.01 \times 10^{-5}$ |

| | | | | |
|---|---|---|---|---|
| CTHRC1 | 1.16 (1.06-1.27) | $1.52 \times 10^{-3}$ | 1.37 (1.22-1.54) | $1.99 \times 10^{-7}$ |
| SEPHS1 | 1.15 (1.06-1.26) | $1.64 \times 10^{-3}$ | 1.28 (1.13-1.44) | $6.12 \times 10^{-5}$ |
| FKBP7 | 1.16 (1.06-1.27) | $1.73 \times 10^{-3}$ | 1.38 (1.22-1.57) | $3.32 \times 10^{-7}$ |
| NRP2 | 1.17 (1.06-1.29) | $1.80 \times 10^{-3}$ | 1.41 (1.23-1.61) | $6.41 \times 10^{-7}$ |
| NPDC1 | 1.16 (1.06-1.27) | $1.83 \times 10^{-3}$ | 1.41 (1.23-1.60) | $3.77 \times 10^{-7}$ |
| OIT3 | 1.15 (1.05-1.26) | $1.86 \times 10^{-3}$ | 1.41 (1.25-1.60) | $2.17 \times 10^{-8}$ |
| TXNDC5 | 1.17 (1.06-1.29) | $1.87 \times 10^{-3}$ | 1.38 (1.20-1.59) | $6.40 \times 10^{-6}$ |
| MINPP1 | 1.15 (1.05-1.26) | $2.00 \times 10^{-3}$ | 1.46 (1.30-1.65) | $4.71 \times 10^{-10}$ |
| CCDC80 | 1.18 (1.06-1.31) | $2.11 \times 10^{-3}$ | 1.38 (1.20-1.59) | $8.72 \times 10^{-6}$ |
| CCL21 | 1.16 (1.05-1.27) | $2.15 \times 10^{-3}$ | 1.37 (1.22-1.56) | $3.93 \times 10^{-7}$ |
| ITGA1\|ITGB1 | 1.16 (1.05-1.27) | $2.17 \times 10^{-3}$ | 1.37 (1.20-1.55) | $1.33 \times 10^{-6}$ |
| C7 | 1.16 (1.05-1.27) | $2.25 \times 10^{-3}$ | 1.42 (1.25-1.62) | $1.06 \times 10^{-7}$ |
| ERLEC1 | 1.15 (1.05-1.26) | $2.63 \times 10^{-3}$ | 1.37 (1.21-1.54) | $3.87 \times 10^{-7}$ |
| UNC5B | 1.16 (1.05-1.29) | $2.69 \times 10^{-3}$ | 1.34 (1.17-1.54) | $2.38 \times 10^{-5}$ |
| CXCL8 | 1.15 (1.05-1.25) | $3.10 \times 10^{-3}$ | 1.31 (1.17-1.47) | $4.57 \times 10^{-6}$ |
| DAG1 | 1.15 (1.05-1.26) | $3.16 \times 10^{-3}$ | 1.28 (1.14-1.43) | $4.01 \times 10^{-5}$ |
| CD59 | 1.15 (1.05-1.26) | $3.51 \times 10^{-3}$ | 1.36 (1.19-1.56) | $7.59 \times 10^{-6}$ |
| RPN1 | 1.15 (1.05-1.26) | $3.54 \times 10^{-3}$ | 1.33 (1.18-1.49) | $1.87 \times 10^{-6}$ |
| MAZ | 1.14 (1.04-1.25) | $4.08 \times 10^{-3}$ | 1.27 (1.13-1.44) | $7.68 \times 10^{-5}$ |
| ESM1 | 1.15 (1.04-1.27) | $4.48 \times 10^{-3}$ | 1.33 (1.17-1.51) | $1.11 \times 10^{-5}$ |
| EDN1 | 1.15 (1.04-1.27) | $4.89 \times 10^{-3}$ | 1.47 (1.30-1.67) | $1.02 \times 10^{-9}$ |
| FJX1 | 1.14 (1.04-1.26) | $5.00 \times 10^{-3}$ | 1.35 (1.19-1.53) | $1.97 \times 10^{-6}$ |
| SCARA5 | 1.14 (1.04-1.26) | $5.15 \times 10^{-3}$ | 1.51 (1.33-1.71) | $9.56 \times 10^{-11}$ |
| SERPINA11 | 1.14 (1.04-1.24) | $5.23 \times 10^{-3}$ | 1.35 (1.20-1.52) | $8.74 \times 10^{-7}$ |

**Supplemental Table 5: Proteins Significantly Associated With Heart Failure Hospitalization or Cardiovascular Death in Cardiovascular Risk Factor Adjusted Model**

| | Discovery (ATMOSPHERE) | | Replication (PARADIGM) | |
|---|---|---|---|---|
| Protein | HR (95% CI) | *P* Value | HR (95% CI) | *P* Value |
| SVEP1 | 1.42 (1.30-1.55) | $4.34 \times 10^{-14}$ | 1.49 (1.32-1.67) | $2.44 \times 10^{-11}$ |
| GDF15 | 1.40 (1.26-1.54) | $7.52 \times 10^{-11}$ | 1.47 (1.28-1.69) | $8.00 \times 10^{-8}$ |
| NPPB | 1.46 (1.30-1.64) | $1.25 \times 10^{-10}$ | 1.50 (1.31-1.72) | $3.87 \times 10^{-9}$ |
| ANGPT2 | 1.40 (1.26-1.55) | $1.52 \times 10^{-10}$ | 1.53 (1.35-1.74) | $4.29 \times 10^{-11}$ |
| THBS2 | 1.37 (1.24-1.50) | $1.90 \times 10^{-10}$ | 1.50 (1.34-1.68) | $7.25 \times 10^{-12}$ |
| LTBP4 | 1.39 (1.25-1.55) | $9.49 \times 10^{-10}$ | 1.32 (1.19-1.47) | $2.81 \times 10^{-7}$ |
| QSOX1 | 1.34 (1.22-1.48) | $1.31 \times 10^{-9}$ | 1.34 (1.20-1.50) | $2.64 \times 10^{-7}$ |
| ADAMTSL2 | 1.33 (1.21-1.47) | $3.53 \times 10^{-9}$ | 1.35 (1.20-1.52) | $7.04 \times 10^{-7}$ |
| IL1RL1 | 1.35 (1.21-1.50) | $3.22 \times 10^{-8}$ | 1.38 (1.22-1.56) | $4.17 \times 10^{-7}$ |
| CST3 | 1.38 (1.23-1.54) | $3.39 \times 10^{-8}$ | 1.41 (1.20-1.66) | $3.64 \times 10^{-5}$ |
| SPON2 | 1.30 (1.17-1.44) | $5.05 \times 10^{-7}$ | 1.38 (1.20-1.59) | $7.16 \times 10^{-6}$ |
| IGFBP7 | 1.29 (1.16-1.42) | $7.04 \times 10^{-7}$ | 1.45 (1.30-1.63) | $1.28 \times 10^{-10}$ |
| TNC | 1.28 (1.16-1.41) | $7.50 \times 10^{-7}$ | 1.34 (1.20-1.50) | $2.78 \times 10^{-7}$ |
| EFEMP1 | 1.27 (1.15-1.39) | $9.01 \times 10^{-7}$ | 1.24 (1.12-1.36) | $1.29 \times 10^{-5}$ |
| ATP5PF | 1.23 (1.13-1.34) | $1.61 \times 10^{-6}$ | 1.25 (1.13-1.39) | $1.21 \times 10^{-5}$ |
| SPON1 | 1.28 (1.16-1.43) | $2.37 \times 10^{-6}$ | 1.47 (1.29-1.69) | $1.43 \times 10^{-8}$ |
| TNNI3 | 1.20 (1.11-1.30) | $2.52 \times 10^{-6}$ | 1.22 (1.13-1.33) | $1.68 \times 10^{-6}$ |
| C1QTNF1 | 1.25 (1.14-1.37) | $2.85 \times 10^{-6}$ | 1.42 (1.26-1.60) | $1.47 \times 10^{-8}$ |
| NFASC | 1.25 (1.14-1.37) | $3.36 \times 10^{-6}$ | 1.38 (1.22-1.57) | $3.52 \times 10^{-7}$ |
| IL19 | 1.27 (1.15-1.40) | $4.14 \times 10^{-6}$ | 1.27 (1.12-1.44) | $1.67 \times 10^{-4}$ |
| MMP19 | 1.27 (1.15-1.41) | $4.78 \times 10^{-6}$ | 1.48 (1.32-1.67) | $1.84 \times 10^{-11}$ |
| CNDP1 | 0.80 (0.73-0.88) | $4.82 \times 10^{-6}$ | 0.79 (0.70-0.89) | $1.24 \times 10^{-4}$ |
| COL6A3 | 1.26 (1.14-1.40) | $6.87 \times 10^{-6}$ | 1.36 (1.20-1.55) | $3.78 \times 10^{-6}$ |
| IGFBP1 | 1.27 (1.14-1.41) | $1.10 \times 10^{-5}$ | 1.34 (1.17-1.53) | $1.87 \times 10^{-5}$ |
| FSTL3 | 1.28 (1.15-1.44) | $1.42 \times 10^{-5}$ | 1.36 (1.18-1.58) | $3.17 \times 10^{-5}$ |
| MFAP4 | 1.22 (1.11-1.34) | $1.91 \times 10^{-5}$ | 1.43 (1.25-1.64) | $1.55 \times 10^{-7}$ |
| CD5 | 1.19 (1.10-1.29) | $2.54 \times 10^{-5}$ | 1.23 (1.13-1.33) | $5.71 \times 10^{-7}$ |
| CTHRC1 | 1.19 (1.10-1.29) | $2.64 \times 10^{-5}$ | 1.19 (1.09-1.29) | $1.24 \times 10^{-4}$ |
| RSPO4 | 1.16 (1.08-1.24) | $4.22 \times 10^{-5}$ | 1.32 (1.20-1.44) | $4.46 \times 10^{-9}$ |
| COL28A1 | 1.26 (1.13-1.40) | $4.72 \times 10^{-5}$ | 1.38 (1.20-1.60) | $9.50 \times 10^{-6}$ |
| PTK7 | 1.22 (1.10-1.34) | $7.55 \times 10^{-5}$ | 1.41 (1.24-1.60) | $1.34 \times 10^{-7}$ |
| TFF3 | 1.24 (1.11-1.38) | $8.01 \times 10^{-5}$ | 1.39 (1.19-1.62) | $2.88 \times 10^{-5}$ |

| | | | | |
|---|---|---|---|---|
| MMP2 | 1.22 (1.10-1.35) | $1.14 \times 10^{-4}$ | 1.31 (1.16-1.49) | $2.26 \times 10^{-5}$ |
| ANGPTL3 | 1.22 (1.10-1.34) | $1.25 \times 10^{-4}$ | 1.28 (1.13-1.46) | $1.91 \times 10^{-4}$ |
| TAGLN | 1.25 (1.12-1.41) | $1.46 \times 10^{-4}$ | 1.39 (1.20-1.62) | $1.68 \times 10^{-5}$ |
| KERA | 1.21 (1.09-1.33) | $1.69 \times 10^{-4}$ | 1.27 (1.14-1.41) | $1.03 \times 10^{-5}$ |
| PTPRU | 1.19 (1.08-1.30) | $1.84 \times 10^{-4}$ | 1.37 (1.21-1.55) | $6.69 \times 10^{-7}$ |
| CTSB | 1.22 (1.10-1.35) | $1.87 \times 10^{-4}$ | 1.34 (1.17-1.53) | $1.34 \times 10^{-5}$ |
| RBL2 | 1.19 (1.09-1.30) | $1.99 \times 10^{-4}$ | 1.32 (1.19-1.47) | $2.96 \times 10^{-7}$ |
| TNFRSF11B | 1.21 (1.09-1.33) | $2.34 \times 10^{-4}$ | 1.33 (1.17-1.52) | $2.06 \times 10^{-5}$ |
| CHST15 | 1.21 (1.09-1.33) | $2.60 \times 10^{-4}$ | 1.40 (1.24-1.59) | $1.30 \times 10^{-7}$ |
| ICAM5 | 1.20 (1.09-1.33) | $2.75 \times 10^{-4}$ | 1.44 (1.27-1.64) | $1.44 \times 10^{-8}$ |
| IBSP | 1.18 (1.08-1.28) | $3.30 \times 10^{-4}$ | 1.28 (1.14-1.44) | $4.25 \times 10^{-5}$ |
| TST | 1.19 (1.08-1.31) | $3.38 \times 10^{-4}$ | 1.33 (1.17-1.50) | $7.18 \times 10^{-6}$ |
| EPO | 1.17 (1.07-1.29) | $5.23 \times 10^{-4}$ | 1.43 (1.29-1.59) | $3.53 \times 10^{-11}$ |
| CCL14 | 1.21 (1.09-1.35) | $5.30 \times 10^{-4}$ | 1.30 (1.13-1.50) | $2.07 \times 10^{-4}$ |
| WFDC2 | 1.22 (1.09-1.36) | $5.55 \times 10^{-4}$ | 1.39 (1.21-1.60) | $3.61 \times 10^{-6}$ |
| STC1 | 1.19 (1.08-1.31) | $5.68 \times 10^{-4}$ | 1.37 (1.20-1.57) | $4.22 \times 10^{-6}$ |
| GNPTG | 1.14 (1.05-1.22) | $7.94 \times 10^{-4}$ | 1.41 (1.26-1.59) | $9.52 \times 10^{-9}$ |
| WARS1 | 1.19 (1.07-1.31) | $8.40 \times 10^{-4}$ | 1.33 (1.18-1.50) | $3.17 \times 10^{-6}$ |
| CCL28 | 1.18 (1.07-1.30) | $9.00 \times 10^{-4}$ | 1.28 (1.14-1.44) | $5.04 \times 10^{-5}$ |
| MMP7 | 1.19 (1.07-1.31) | $9.23 \times 10^{-4}$ | 1.32 (1.17-1.50) | $1.17 \times 10^{-5}$ |
| CHRDL2 | 1.18 (1.07-1.30) | $9.43 \times 10^{-4}$ | 1.29 (1.13-1.47) | $1.32 \times 10^{-4}$ |
| SHBG | 1.17 (1.06-1.28) | $1.01 \times 10^{-3}$ | 1.27 (1.13-1.43) | $1.01 \times 10^{-4}$ |
| PLXDC2 | 1.16 (1.06-1.26) | $1.06 \times 10^{-3}$ | 1.42 (1.27-1.59) | $1.02 \times 10^{-9}$ |
| CILP2 | 0.85 (0.77-0.94) | $1.10 \times 10^{-3}$ | 0.78 (0.68-0.88) | $1.10 \times 10^{-4}$ |
| FGF23 | 1.13 (1.05-1.22) | $1.12 \times 10^{-3}$ | 1.28 (1.17-1.41) | $2.82 \times 10^{-7}$ |
| LUM | 1.17 (1.06-1.29) | $1.26 \times 10^{-3}$ | 1.39 (1.24-1.57) | $5.09 \times 10^{-8}$ |
| AMIGO2 | 1.17 (1.06-1.29) | $1.37 \times 10^{-3}$ | 1.26 (1.12-1.42) | $1.43 \times 10^{-4}$ |
| WFDC1 | 1.17 (1.06-1.29) | $1.41 \times 10^{-3}$ | 1.40 (1.22-1.61) | $1.86 \times 10^{-6}$ |
| TFRC | 1.17 (1.06-1.28) | $1.42 \times 10^{-3}$ | 1.30 (1.16-1.46) | $1.14 \times 10^{-5}$ |
| TFF2 | 1.16 (1.06-1.27) | $1.54 \times 10^{-3}$ | 1.27 (1.15-1.40) | $2.89 \times 10^{-6}$ |
| ASAH2 | 0.84 (0.75-0.93) | $1.58 \times 10^{-3}$ | 0.77 (0.67-0.88) | $2.28 \times 10^{-4}$ |
| PLA2R1 | 1.16 (1.06-1.27) | $1.64 \times 10^{-3}$ | 1.33 (1.20-1.48) | $1.32 \times 10^{-7}$ |

**Supplemental Table 6: Proteins Significantly Associated With Cardiovascular Death in Minimally Adjusted Model**

| Protein | Discovery (ATMOSPHERE) | | Replication (PARADIGM) | |
|---|---|---|---|---|
| | HR (95% CI) | *P* Value | HR (95% CI) | *P* Value |
| SVEP1 | 1.54 (1.40-1.68) | $6.32 \times 10^{-20}$ | 1.58 (1.37-1.82) | $2.00 \times 10^{-10}$ |
| GDF15 | 1.54 (1.40-1.70) | $2.26 \times 10^{-18}$ | 1.47 (1.25-1.72) | $1.84 \times 10^{-6}$ |
| ANGPT2 | 1.56 (1.41-1.73) | $2.08 \times 10^{-17}$ | 1.73 (1.47-2.03) | $1.95 \times 10^{-11}$ |
| THBS2 | 1.48 (1.34-1.63) | $6.26 \times 10^{-15}$ | 1.65 (1.44-1.90) | $2.52 \times 10^{-12}$ |
| LTBP4 | 1.50 (1.35-1.66) | $3.65 \times 10^{-14}$ | 1.35 (1.19-1.53) | $1.71 \times 10^{-6}$ |
| IGFBP1 | 1.52 (1.36-1.70) | $6.87 \times 10^{-14}$ | 1.56 (1.32-1.84) | $1.21 \times 10^{-7}$ |
| SPON1 | 1.43 (1.30-1.58) | $5.08 \times 10^{-13}$ | 1.57 (1.39-1.78) | $5.09 \times 10^{-13}$ |
| ADAMTSL2 | 1.44 (1.30-1.60) | $1.31 \times 10^{-12}$ | 1.41 (1.22-1.63) | $4.83 \times 10^{-6}$ |
| RSPO4 | 1.27 (1.19-1.35) | $1.51 \times 10^{-12}$ | 1.42 (1.28-1.57) | $1.04 \times 10^{-11}$ |
| NPPB | 1.60 (1.40-1.82) | $1.84 \times 10^{-12}$ | 1.64 (1.37-1.96) | $5.00 \times 10^{-8}$ |
| QSOX1 | 1.45 (1.30-1.61) | $5.13 \times 10^{-12}$ | 1.34 (1.16-1.55) | $5.11 \times 10^{-5}$ |
| COL6A3 | 1.44 (1.30-1.59) | $5.90 \times 10^{-12}$ | 1.60 (1.38-1.87) | $1.25 \times 10^{-9}$ |
| B2M | 1.44 (1.29-1.60) | $2.32 \times 10^{-11}$ | 1.56 (1.31-1.86) | $4.36 \times 10^{-7}$ |
| SPON2 | 1.46 (1.31-1.64) | $3.64 \times 10^{-11}$ | 1.64 (1.38-1.95) | $1.90 \times 10^{-8}$ |
| TNNI3 | 1.30 (1.20-1.41) | $5.67 \times 10^{-11}$ | 1.28 (1.16-1.41) | $7.78 \times 10^{-7}$ |
| IGFBP2 | 1.32 (1.22-1.44) | $1.03 \times 10^{-10}$ | 1.34 (1.16-1.54) | $7.01 \times 10^{-5}$ |
| TNC | 1.46 (1.30-1.63) | $1.11 \times 10^{-10}$ | 1.57 (1.34-1.83) | $2.74 \times 10^{-8}$ |
| IGFBP7 | 1.41 (1.27-1.57) | $2.11 \times 10^{-10}$ | 1.64 (1.42-1.89) | $1.44 \times 10^{-11}$ |
| COL28A1 | 1.44 (1.28-1.61) | $2.44 \times 10^{-10}$ | 1.68 (1.42-1.99) | $1.31 \times 10^{-9}$ |
| NFASC | 1.34 (1.22-1.47) | $6.41 \times 10^{-10}$ | 1.54 (1.33-1.79) | $7.11 \times 10^{-9}$ |
| CST3 | 1.40 (1.26-1.56) | $7.94 \times 10^{-10}$ | 1.71 (1.44-2.03) | $9.74 \times 10^{-10}$ |
| C1QTNF1 | 1.40 (1.25-1.55) | $1.03 \times 10^{-9}$ | 1.52 (1.31-1.76) | $2.72 \times 10^{-8}$ |
| FSTL3 | 1.44 (1.28-1.62) | $1.47 \times 10^{-9}$ | 1.67 (1.40-1.99) | $1.22 \times 10^{-8}$ |
| PTK7 | 1.39 (1.25-1.55) | $1.55 \times 10^{-9}$ | 1.68 (1.43-1.97) | $3.25 \times 10^{-10}$ |
| CTSH | 1.31 (1.20-1.44) | $2.11 \times 10^{-9}$ | 1.73 (1.48-2.01) | $2.96 \times 10^{-12}$ |
| TAGLN | 1.46 (1.29-1.66) | $4.47 \times 10^{-9}$ | 1.66 (1.38-1.99) | $4.10 \times 10^{-8}$ |
| PXDN | 1.46 (1.29-1.66) | $4.48 \times 10^{-9}$ | 1.43 (1.20-1.71) | $8.49 \times 10^{-5}$ |
| RBL2 | 1.36 (1.22-1.50) | $7.80 \times 10^{-9}$ | 1.60 (1.42-1.80) | $2.33 \times 10^{-14}$ |
| WFDC2 | 1.41 (1.25-1.59) | $1.10 \times 10^{-8}$ | 1.65 (1.39-1.95) | $8.55 \times 10^{-9}$ |
| SEMA6B | 1.36 (1.22-1.51) | $1.25 \times 10^{-8}$ | 1.26 (1.14-1.40) | $4.95 \times 10^{-6}$ |
| EPO | 1.34 (1.21-1.48) | $1.31 \times 10^{-8}$ | 1.57 (1.37-1.80) | $1.20 \times 10^{-10}$ |
| TFF3 | 1.35 (1.21-1.49) | $1.33 \times 10^{-8}$ | 1.65 (1.39-1.95) | $8.23 \times 10^{-9}$ |
| EFEMP1 | 1.35 (1.21-1.49) | $1.96 \times 10^{-8}$ | 1.31 (1.17-1.47) | $1.98 \times 10^{-6}$ |
| RNASE1 | 1.39 (1.24-1.57) | $2.40 \times 10^{-8}$ | 1.60 (1.34-1.91) | $1.76 \times 10^{-7}$ |
| TREM1 | 1.36 (1.22-1.52) | $2.95 \times 10^{-8}$ | 1.44 (1.21-1.71) | $3.44 \times 10^{-5}$ |
| GABARAPL1 | 1.30 (1.19-1.43) | $3.46 \times 10^{-8}$ | 1.38 (1.18-1.61) | $4.93 \times 10^{-5}$ |
| IL1RL1 | 1.38 (1.23-1.55) | $5.02 \times 10^{-8}$ | 1.50 (1.27-1.77) | $2.04 \times 10^{-6}$ |
| GRN | 1.35 (1.21-1.51) | $7.01 \times 10^{-8}$ | 1.53 (1.29-1.80) | $4.97 \times 10^{-7}$ |

| | | | | |
|---|---|---|---|---|
| TFRC | 1.34 (1.21-1.50) | $7.95 \times 10^{-8}$ | 1.61 (1.39-1.87) | $3.25 \times 10^{-10}$ |
| RNASE6 | 1.34 (1.20-1.49) | $1.19 \times 10^{-7}$ | 1.44 (1.28-1.63) | $4.03 \times 10^{-9}$ |
| MMP19 | 1.36 (1.21-1.53) | $1.61 \times 10^{-7}$ | 1.64 (1.43-1.88) | $6.64 \times 10^{-13}$ |
| B3GAT3 | 1.33 (1.20-1.49) | $1.97 \times 10^{-7}$ | 1.44 (1.24-1.68) | $3.65 \times 10^{-6}$ |
| UMOD | 1.33 (1.19-1.48) | $2.18 \times 10^{-7}$ | 1.53 (1.32-1.77) | $1.22 \times 10^{-8}$ |
| Igh | 1.31 (1.18-1.45) | $2.46 \times 10^{-7}$ | 1.41 (1.22-1.62) | $1.65 \times 10^{-6}$ |
| TNFRSF1B | 1.36 (1.21-1.52) | $2.77 \times 10^{-7}$ | 1.54 (1.29-1.84) | $1.65 \times 10^{-6}$ |
| KERA | 1.32 (1.19-1.47) | $2.80 \times 10^{-7}$ | 1.31 (1.15-1.49) | $3.29 \times 10^{-5}$ |
| GREM2 | 1.30 (1.18-1.44) | $3.53 \times 10^{-7}$ | 1.38 (1.22-1.56) | $4.40 \times 10^{-7}$ |
| CCL14 | 1.36 (1.21-1.53) | $3.55 \times 10^{-7}$ | 1.60 (1.34-1.92) | $2.88 \times 10^{-7}$ |
| CD5 | 1.21 (1.13-1.31) | $3.56 \times 10^{-7}$ | 1.31 (1.21-1.42) | $1.48 \times 10^{-11}$ |
| TNFRSF1A | 1.32 (1.19-1.47) | $3.73 \times 10^{-7}$ | 1.47 (1.26-1.71) | $6.73 \times 10^{-7}$ |
| KLK10 | 1.33 (1.19-1.48) | $4.42 \times 10^{-7}$ | 1.45 (1.24-1.69) | $3.21 \times 10^{-6}$ |
| CD163 | 1.31 (1.18-1.46) | $4.52 \times 10^{-7}$ | 1.41 (1.19-1.67) | $5.27 \times 10^{-5}$ |
| CTSB | 1.33 (1.19-1.49) | $5.50 \times 10^{-7}$ | 1.53 (1.29-1.81) | $1.00 \times 10^{-6}$ |
| WARS1 | 1.33 (1.19-1.48) | $6.88 \times 10^{-7}$ | 1.54 (1.31-1.81) | $2.21 \times 10^{-7}$ |
| RELT | 1.31 (1.18-1.46) | $7.48 \times 10^{-7}$ | 1.32 (1.17-1.50) | $8.70 \times 10^{-6}$ |
| WFDC1 | 1.33 (1.19-1.48) | $7.99 \times 10^{-7}$ | 1.63 (1.37-1.93) | $3.45 \times 10^{-8}$ |
| ICAM5 | 1.32 (1.18-1.47) | $1.02 \times 10^{-6}$ | 1.66 (1.41-1.96) | $2.70 \times 10^{-9}$ |
| STC1 | 1.31 (1.17-1.46) | $1.35 \times 10^{-6}$ | 1.45 (1.22-1.72) | $2.92 \times 10^{-5}$ |
| MFAP4 | 1.28 (1.16-1.42) | $1.72 \times 10^{-6}$ | 2.05 (1.73-2.42) | $7.31 \times 10^{-17}$ |
| CHST15 | 1.31 (1.17-1.47) | $2.51 \times 10^{-6}$ | 1.57 (1.34-1.84) | $4.01 \times 10^{-8}$ |
| PTPRU | 1.26 (1.15-1.40) | $2.74 \times 10^{-6}$ | 1.53 (1.33-1.76) | $3.17 \times 10^{-9}$ |
| ROBO2 | 1.30 (1.16-1.45) | $3.94 \times 10^{-6}$ | 1.53 (1.32-1.77) | $9.80 \times 10^{-9}$ |
| TNFRSF11B | 1.30 (1.16-1.45) | $4.02 \times 10^{-6}$ | 1.55 (1.32-1.84) | $2.15 \times 10^{-7}$ |
| EPHA2 | 1.31 (1.17-1.47) | $4.42 \times 10^{-6}$ | 1.51 (1.29-1.77) | $5.34 \times 10^{-7}$ |
| MMP7 | 1.30 (1.16-1.45) | $5.63 \times 10^{-6}$ | 1.51 (1.31-1.74) | $2.18 \times 10^{-8}$ |
| CLPSL1 | 1.26 (1.14-1.40) | $5.65 \times 10^{-6}$ | 1.35 (1.19-1.53) | $3.19 \times 10^{-6}$ |
| LAMC2 | 1.26 (1.14-1.39) | $6.01 \times 10^{-6}$ | 1.30 (1.16-1.46) | $9.11 \times 10^{-6}$ |
| INHBB | 1.28 (1.15-1.43) | $6.14 \times 10^{-6}$ | 1.44 (1.23-1.70) | $7.88 \times 10^{-6}$ |
| PLXDC2 | 1.24 (1.13-1.36) | $6.36 \times 10^{-6}$ | 1.71 (1.49-1.98) | $9.00 \times 10^{-14}$ |
| LUM | 1.28 (1.15-1.42) | $7.04 \times 10^{-6}$ | 1.45 (1.25-1.69) | $1.18 \times 10^{-6}$ |
| OLFM1 | 1.20 (1.11-1.30) | $7.05 \times 10^{-6}$ | 1.26 (1.14-1.40) | $1.67 \times 10^{-5}$ |
| RSPO3 | 1.21 (1.11-1.32) | $7.16 \times 10^{-6}$ | 1.26 (1.15-1.37) | $2.27 \times 10^{-7}$ |
| NBL1 | 1.28 (1.15-1.42) | $7.49 \times 10^{-6}$ | 1.41 (1.21-1.65) | $1.01 \times 10^{-5}$ |
| FGF23 | 1.17 (1.09-1.25) | $7.87 \times 10^{-6}$ | 1.36 (1.22-1.53) | $5.88 \times 10^{-8}$ |
| CNDP1 | 0.78 (0.70-0.87) | $1.00 \times 10^{-5}$ | 0.73 (0.63-0.85) | $3.86 \times 10^{-5}$ |
| MMP2 | 1.30 (1.16-1.46) | $1.01 \times 10^{-5}$ | 1.56 (1.32-1.85) | $1.62 \times 10^{-7}$ |
| CHRDL2 | 1.28 (1.14-1.42) | $1.10 \times 10^{-5}$ | 1.51 (1.28-1.80) | $1.90 \times 10^{-6}$ |
| NRP2 | 1.27 (1.14-1.42) | $1.33 \times 10^{-5}$ | 1.46 (1.27-1.68) | $8.19 \times 10^{-8}$ |
| BMP6 | 1.24 (1.12-1.37) | $1.79 \times 10^{-5}$ | 1.40 (1.23-1.59) | $4.73 \times 10^{-7}$ |
| COL18A1 | 1.29 (1.15-1.44) | $1.80 \times 10^{-5}$ | 1.61 (1.35-1.91) | $9.04 \times 10^{-8}$ |

| | | | | |
|---|---|---|---|---|
| AGRP | 1.21 (1.11-1.32) | $2.15 \times 10^{-5}$ | 1.39 (1.22-1.58) | $6.80 \times 10^{-7}$ |
| PIGR | 1.26 (1.13-1.41) | $2.21 \times 10^{-5}$ | 1.47 (1.26-1.71) | $8.06 \times 10^{-7}$ |
| CD300C | 1.28 (1.14-1.43) | $2.58 \times 10^{-5}$ | 1.42 (1.20-1.68) | $4.56 \times 10^{-5}$ |
| DNAJB12 | 1.22 (1.11-1.34) | $3.08 \times 10^{-5}$ | 1.36 (1.18-1.56) | $1.36 \times 10^{-5}$ |
| RNASE4 | 1.26 (1.13-1.41) | $3.75 \times 10^{-5}$ | 1.59 (1.33-1.91) | $4.59 \times 10^{-7}$ |
| CXCL12 | 1.27 (1.13-1.42) | $4.13 \times 10^{-5}$ | 1.58 (1.31-1.89) | $9.02 \times 10^{-7}$ |
| CD93 | 1.26 (1.13-1.40) | $4.58 \times 10^{-5}$ | 1.59 (1.35-1.88) | $2.39 \times 10^{-8}$ |
| CHRDL1 | 1.27 (1.13-1.43) | $4.96 \times 10^{-5}$ | 1.66 (1.37-2.01) | $1.85 \times 10^{-7}$ |
| SIGLEC7 | 1.24 (1.12-1.38) | $5.21 \times 10^{-5}$ | 1.50 (1.32-1.72) | $1.38 \times 10^{-9}$ |
| CTHRC1 | 1.19 (1.09-1.29) | $6.01 \times 10^{-5}$ | 1.25 (1.12-1.40) | $6.91 \times 10^{-5}$ |
| CCDC80 | 1.27 (1.13-1.43) | $6.11 \times 10^{-5}$ | 1.70 (1.44-2.01) | $6.26 \times 10^{-10}$ |
| PLXNB2 | 1.24 (1.11-1.38) | $7.23 \times 10^{-5}$ | 1.43 (1.21-1.69) | $3.51 \times 10^{-5}$ |
| IL6ST | 1.25 (1.12-1.40) | $8.12 \times 10^{-5}$ | 1.50 (1.26-1.78) | $5.34 \times 10^{-6}$ |
| PPP4R3A | 1.22 (1.10-1.35) | $8.33 \times 10^{-5}$ | 1.36 (1.17-1.58) | $4.48 \times 10^{-5}$ |
| TGFB3 | 1.20 (1.09-1.31) | $8.50 \times 10^{-5}$ | 1.29 (1.16-1.43) | $2.79 \times 10^{-6}$ |
| H6PD | 1.25 (1.12-1.39) | $8.74 \times 10^{-5}$ | 1.42 (1.21-1.67) | $1.55 \times 10^{-5}$ |
| FSTL1 | 1.25 (1.12-1.40) | $9.65 \times 10^{-5}$ | 1.57 (1.36-1.82) | $5.70 \times 10^{-10}$ |
| HTRA2 | 1.22 (1.10-1.34) | $9.90 \times 10^{-5}$ | 1.36 (1.26-1.48) | $1.67 \times 10^{-13}$ |
| UNC5C | 1.26 (1.12-1.42) | $1.17 \times 10^{-4}$ | 1.61 (1.35-1.93) | $2.02 \times 10^{-7}$ |
| IGFLR1 | 1.19 (1.09-1.31) | $1.65 \times 10^{-4}$ | 1.35 (1.18-1.55) | $1.58 \times 10^{-5}$ |
| MMP12 | 1.25 (1.11-1.41) | $1.68 \times 10^{-4}$ | 1.53 (1.29-1.81) | $7.91 \times 10^{-7}$ |
| SCARA5 | 1.22 (1.10-1.35) | $1.73 \times 10^{-4}$ | 1.36 (1.21-1.52) | $1.05 \times 10^{-7}$ |
| CRELD1 | 1.25 (1.11-1.41) | $1.79 \times 10^{-4}$ | 1.43 (1.22-1.68) | $1.46 \times 10^{-5}$ |
| IGF1R | 1.23 (1.10-1.37) | $1.80 \times 10^{-4}$ | 1.51 (1.29-1.78) | $5.06 \times 10^{-7}$ |
| SLPI | 1.24 (1.10-1.38) | $2.21 \times 10^{-4}$ | 1.46 (1.25-1.71) | $1.71 \times 10^{-6}$ |
| APOF | 1.27 (1.12-1.45) | $2.32 \times 10^{-4}$ | 1.66 (1.36-2.01) | $4.05 \times 10^{-7}$ |
| CCN1 | 1.17 (1.07-1.27) | $2.57 \times 10^{-4}$ | 1.38 (1.26-1.51) | $5.50 \times 10^{-12}$ |
| PLA2G2A | 1.23 (1.10-1.37) | $3.04 \times 10^{-4}$ | 1.58 (1.35-1.85) | $1.04 \times 10^{-8}$ |
| AMIGO2 | 1.22 (1.10-1.36) | $3.04 \times 10^{-4}$ | 1.48 (1.28-1.71) | $1.84 \times 10^{-7}$ |
| VEGFD | 1.20 (1.08-1.32) | $3.75 \times 10^{-4}$ | 1.49 (1.34-1.67) | $1.03 \times 10^{-12}$ |
| PCDHGA10 | 1.16 (1.07-1.26) | $3.93 \times 10^{-4}$ | 1.26 (1.14-1.40) | $6.65 \times 10^{-6}$ |
| EPHB2 | 1.23 (1.10-1.38) | $3.94 \times 10^{-4}$ | 1.38 (1.18-1.62) | $8.20 \times 10^{-5}$ |
| IL15 | 1.19 (1.08-1.31) | $4.12 \times 10^{-4}$ | 1.39 (1.21-1.61) | $6.70 \times 10^{-6}$ |
| TFF2 | 1.19 (1.08-1.32) | $5.39 \times 10^{-4}$ | 1.33 (1.19-1.49) | $8.51 \times 10^{-7}$ |
| CCL21 | 1.21 (1.09-1.35) | $5.44 \times 10^{-4}$ | 1.48 (1.26-1.73) | $1.26 \times 10^{-6}$ |
| SFRP1 | 1.21 (1.09-1.35) | $6.00 \times 10^{-4}$ | 1.34 (1.17-1.52) | $1.23 \times 10^{-5}$ |
| IL15RA | 1.20 (1.08-1.34) | $7.47 \times 10^{-4}$ | 1.30 (1.17-1.44) | $1.03 \times 10^{-6}$ |
| TXNDC5 | 1.20 (1.08-1.33) | $7.67 \times 10^{-4}$ | 1.57 (1.33-1.87) | $1.68 \times 10^{-7}$ |
| LILRB2 | 1.21 (1.08-1.36) | $8.16 \times 10^{-4}$ | 1.46 (1.23-1.74) | $1.49 \times 10^{-5}$ |
| F8 | 1.21 (1.08-1.36) | $9.41 \times 10^{-4}$ | 1.54 (1.28-1.85) | $4.35 \times 10^{-6}$ |
| FJX1 | 1.20 (1.08-1.34) | $9.71 \times 10^{-4}$ | 1.30 (1.16-1.45) | $3.26 \times 10^{-6}$ |
| C9 | 1.21 (1.08-1.36) | $1.18 \times 10^{-3}$ | 1.58 (1.32-1.90) | $5.54 \times 10^{-7}$ |

| | | | | |
|---|---|---|---|---|
| GNPTG | 1.15 (1.06-1.26) | $1.21 \times 10^{-3}$ | 1.56 (1.36-1.79) | $1.45 \times 10^{-10}$ |
| CLSTN3 | 1.16 (1.06-1.27) | $1.24 \times 10^{-3}$ | 1.42 (1.27-1.59) | $1.53 \times 10^{-9}$ |
| UNC5B | 1.21 (1.08-1.35) | $1.31 \times 10^{-3}$ | 1.36 (1.22-1.53) | $1.01 \times 10^{-7}$ |
| AXIN2 | 1.20 (1.07-1.34) | $1.41 \times 10^{-3}$ | 1.65 (1.45-1.87) | $1.08 \times 10^{-14}$ |
| ADAM12 | 1.15 (1.05-1.25) | $1.63 \times 10^{-3}$ | 1.33 (1.18-1.49) | $2.74 \times 10^{-6}$ |
| EDN1 | 1.14 (1.05-1.24) | $1.66 \times 10^{-3}$ | 1.34 (1.19-1.52) | $1.72 \times 10^{-6}$ |
| MATN2 | 1.21 (1.07-1.36) | $1.68 \times 10^{-3}$ | 1.53 (1.28-1.84) | $3.80 \times 10^{-6}$ |
| NPDC1 | 1.18 (1.06-1.31) | $1.70 \times 10^{-3}$ | 1.45 (1.29-1.63) | $1.19 \times 10^{-9}$ |
| ANGPTL1 | 1.18 (1.06-1.31) | $1.70 \times 10^{-3}$ | 1.41 (1.19-1.67) | $6.87 \times 10^{-5}$ |
| SCARF1 | 1.18 (1.06-1.31) | $2.26 \times 10^{-3}$ | 1.43 (1.23-1.66) | $2.71 \times 10^{-6}$ |
| TMED10 | 1.15 (1.05-1.26) | $2.43 \times 10^{-3}$ | 1.65 (1.42-1.92) | $7.80 \times 10^{-11}$ |
| EBI3 | 1.20 (1.06-1.34) | $2.61 \times 10^{-3}$ | 1.67 (1.39-2.00) | $3.61 \times 10^{-8}$ |
| PAPPA | 1.19 (1.06-1.34) | $2.77 \times 10^{-3}$ | 1.53 (1.29-1.82) | $1.10 \times 10^{-6}$ |
| ITGA1\|ITGB1 | 1.18 (1.06-1.31) | $2.80 \times 10^{-3}$ | 1.34 (1.18-1.54) | $1.66 \times 10^{-5}$ |
| IBSP | 1.16 (1.05-1.28) | $3.07 \times 10^{-3}$ | 1.37 (1.18-1.58) | $2.66 \times 10^{-5}$ |
| TREM2 | 1.18 (1.06-1.32) | $3.49 \times 10^{-3}$ | 1.46 (1.23-1.72) | $1.07 \times 10^{-5}$ |
| MRC1 | 1.17 (1.05-1.31) | $4.50 \times 10^{-3}$ | 1.52 (1.30-1.78) | $2.56 \times 10^{-7}$ |
| GPNMB | 1.17 (1.05-1.31) | $4.72 \times 10^{-3}$ | 1.40 (1.24-1.59) | $1.33 \times 10^{-7}$ |
| MINPP1 | 1.16 (1.05-1.29) | $4.84 \times 10^{-3}$ | 1.51 (1.34-1.70) | $1.24 \times 10^{-11}$ |
| IGHG1\|IGHG2\|IGHG3\|IGHG4\|IGK\|IGL | 1.17 (1.05-1.31) | $5.12 \times 10^{-3}$ | 1.40 (1.18-1.65) | $7.69 \times 10^{-5}$ |
| CD59 | 1.12 (1.03-1.21) | $5.16 \times 10^{-3}$ | 1.39 (1.22-1.58) | $9.43 \times 10^{-7}$ |

**Supplemental Table 7: Proteins Significantly Associated With Cardiovascular Death in Cardiovascular Risk Factor Adjusted Model**

| | Discovery (ATMOSPHERE) | | Replication (PARADIGM) | |
|---|---|---|---|---|
| Protein | HR (95% CI) | *P* Value | HR (95% CI) | *P* Value |
| SVEP1 | 1.51 (1.36-1.68) | $1.78 \times 10^{-14}$ | 1.50 (1.28-1.76) | $3.89 \times 10^{-7}$ |
| ANGPT2 | 1.49 (1.32-1.69) | $9.81 \times 10^{-11}$ | 1.61 (1.35-1.92) | $1.01 \times 10^{-7}$ |
| LTBP4 | 1.49 (1.32-1.68) | $2.49 \times 10^{-10}$ | 1.36 (1.18-1.55) | $1.24 \times 10^{-5}$ |
| IGFBP1 | 1.49 (1.31-1.69) | $5.49 \times 10^{-10}$ | 1.60 (1.33-1.92) | $4.80 \times 10^{-7}$ |
| THBS2 | 1.42 (1.27-1.58) | $6.56 \times 10^{-10}$ | 1.57 (1.35-1.84) | $1.12 \times 10^{-8}$ |
| IGFBP2 | 1.33 (1.21-1.47) | $1.04 \times 10^{-8}$ | 1.37 (1.16-1.61) | $1.46 \times 10^{-4}$ |
| SPON1 | 1.39 (1.24-1.56) | $1.15 \times 10^{-8}$ | 1.55 (1.34-1.80) | $5.77 \times 10^{-9}$ |
| NPPB | 1.48 (1.28-1.70) | $6.60 \times 10^{-8}$ | 1.55 (1.28-1.88) | $6.64 \times 10^{-6}$ |
| SPON2 | 1.41 (1.24-1.59) | $6.67 \times 10^{-8}$ | 1.61 (1.33-1.94) | $8.15 \times 10^{-7}$ |
| COL6A3 | 1.39 (1.23-1.58) | $9.89 \times 10^{-8}$ | 1.57 (1.33-1.86) | $7.76 \times 10^{-8}$ |
| TNNI3 | 1.26 (1.16-1.38) | $1.21 \times 10^{-7}$ | 1.22 (1.10-1.35) | $1.46 \times 10^{-4}$ |
| B2M | 1.43 (1.25-1.63) | $1.38 \times 10^{-7}$ | 1.63 (1.33-2.00) | $3.61 \times 10^{-6}$ |
| TNC | 1.40 (1.23-1.58) | $1.53 \times 10^{-7}$ | 1.52 (1.28-1.80) | $1.65 \times 10^{-6}$ |
| CST3 | 1.44 (1.26-1.65) | $1.57 \times 10^{-7}$ | 1.97 (1.58-2.45) | $1.13 \times 10^{-9}$ |
| RBL2 | 1.34 (1.20-1.49) | $2.25 \times 10^{-7}$ | 1.52 (1.33-1.73) | $4.43 \times 10^{-10}$ |
| COL28A1 | 1.41 (1.24-1.61) | $2.96 \times 10^{-7}$ | 1.66 (1.38-2.00) | $1.17 \times 10^{-7}$ |
| RSPO4 | 1.22 (1.13-1.32) | $3.20 \times 10^{-7}$ | 1.37 (1.22-1.53) | $9.62 \times 10^{-8}$ |
| C1QTNF1 | 1.34 (1.20-1.50) | $3.39 \times 10^{-7}$ | 1.49 (1.26-1.75) | $1.84 \times 10^{-6}$ |
| IGFBP7 | 1.36 (1.21-1.54) | $4.37 \times 10^{-7}$ | 1.57 (1.35-1.82) | $5.16 \times 10^{-9}$ |
| Igh | 1.31 (1.17-1.45) | $1.17 \times 10^{-6}$ | 1.33 (1.15-1.54) | $1.17 \times 10^{-4}$ |
| PTK7 | 1.33 (1.18-1.50) | $1.93 \times 10^{-6}$ | 1.58 (1.34-1.88) | $1.39 \times 10^{-7}$ |
| NFASC | 1.31 (1.17-1.46) | $2.10 \times 10^{-6}$ | 1.46 (1.25-1.72) | $3.51 \times 10^{-6}$ |
| FSTL3 | 1.39 (1.21-1.60) | $2.42 \times 10^{-6}$ | 1.62 (1.33-1.97) | $1.33 \times 10^{-6}$ |
| RNASE1 | 1.43 (1.23-1.67) | $2.71 \times 10^{-6}$ | 1.74 (1.40-2.15) | $4.61 \times 10^{-7}$ |
| TAGLN | 1.40 (1.22-1.61) | $2.88 \times 10^{-6}$ | 1.61 (1.31-1.96) | $3.80 \times 10^{-6}$ |
| CTSB | 1.33 (1.18-1.50) | $4.56 \times 10^{-6}$ | 1.46 (1.23-1.74) | $1.93 \times 10^{-5}$ |
| IL1RL1 | 1.35 (1.19-1.53) | $4.60 \times 10^{-6}$ | 1.44 (1.21-1.71) | $3.01 \times 10^{-5}$ |
| B3GAT3 | 1.31 (1.16-1.48) | $7.92 \times 10^{-6}$ | 1.38 (1.18-1.61) | $5.09 \times 10^{-5}$ |
| TREM1 | 1.32 (1.17-1.50) | $8.70 \times 10^{-6}$ | 1.45 (1.20-1.75) | $9.37 \times 10^{-5}$ |
| GRN | 1.30 (1.15-1.47) | $1.63 \times 10^{-5}$ | 1.43 (1.20-1.69) | $4.21 \times 10^{-5}$ |
| CCL14 | 1.34 (1.17-1.53) | $2.00 \times 10^{-5}$ | 1.53 (1.26-1.86) | $1.32 \times 10^{-5}$ |
| WARS1 | 1.30 (1.15-1.47) | $2.10 \times 10^{-5}$ | 1.49 (1.26-1.75) | $1.68 \times 10^{-6}$ |
| TFRC | 1.28 (1.14-1.43) | $2.70 \times 10^{-5}$ | 1.45 (1.24-1.69) | $2.97 \times 10^{-6}$ |
| TFF3 | 1.31 (1.15-1.48) | $2.99 \times 10^{-5}$ | 1.71 (1.39-2.11) | $5.71 \times 10^{-7}$ |
| WFDC2 | 1.33 (1.16-1.53) | $3.70 \times 10^{-5}$ | 1.60 (1.33-1.93) | $5.87 \times 10^{-7}$ |
| MMP19 | 1.29 (1.14-1.46) | $4.19 \times 10^{-5}$ | 1.62 (1.39-1.88) | $3.21 \times 10^{-10}$ |

| | | | | |
|---|---|---|---|---|
| RNASE6 | 1.29 (1.14-1.46) | $4.77 \times 10^{-5}$ | 1.42 (1.24-1.63) | $4.99 \times 10^{-7}$ |
| TNFRSF1B | 1.31 (1.15-1.50) | $5.30 \times 10^{-5}$ | 1.50 (1.23-1.83) | $6.42 \times 10^{-5}$ |
| TNFRSF11B | 1.28 (1.13-1.44) | $6.20 \times 10^{-5}$ | 1.50 (1.26-1.79) | $4.66 \times 10^{-6}$ |
| RSPO3 | 1.21 (1.10-1.34) | $6.80 \times 10^{-5}$ | 1.28 (1.16-1.40) | $4.69 \times 10^{-7}$ |
| CLPSL1 | 1.24 (1.11-1.38) | $1.24 \times 10^{-4}$ | 1.32 (1.15-1.51) | $5.33 \times 10^{-5}$ |
| GREM2 | 1.26 (1.12-1.41) | $1.26 \times 10^{-4}$ | 1.36 (1.18-1.57) | $2.67 \times 10^{-5}$ |
| CD5 | 1.19 (1.09-1.30) | $1.59 \times 10^{-4}$ | 1.29 (1.18-1.42) | $3.15 \times 10^{-8}$ |
| KLK10 | 1.27 (1.12-1.43) | $1.61 \times 10^{-4}$ | 1.43 (1.21-1.69) | $3.36 \times 10^{-5}$ |
| LUM | 1.25 (1.11-1.40) | $1.63 \times 10^{-4}$ | 1.41 (1.20-1.65) | $2.29 \times 10^{-5}$ |
| ICAM5 | 1.26 (1.12-1.43) | $1.80 \times 10^{-4}$ | 1.59 (1.34-1.89) | $1.57 \times 10^{-7}$ |
| MMP2 | 1.26 (1.12-1.43) | $2.17 \times 10^{-4}$ | 1.50 (1.26-1.77) | $4.09 \times 10^{-6}$ |
| PLXDC2 | 1.22 (1.10-1.35) | $2.48 \times 10^{-4}$ | 1.62 (1.40-1.88) | $1.01 \times 10^{-10}$ |
| WFDC1 | 1.25 (1.11-1.41) | $2.54 \times 10^{-4}$ | 1.62 (1.34-1.96) | $7.11 \times 10^{-7}$ |
| RELT | 1.27 (1.11-1.44) | $2.70 \times 10^{-4}$ | 1.31 (1.14-1.50) | $1.19 \times 10^{-4}$ |
| TNFRSF1A | 1.26 (1.11-1.42) | $2.74 \times 10^{-4}$ | 1.42 (1.21-1.68) | $2.82 \times 10^{-5}$ |
| UMOD | 1.24 (1.10-1.40) | $2.89 \times 10^{-4}$ | 1.47 (1.25-1.72) | $2.08 \times 10^{-6}$ |
| NRP2 | 1.25 (1.11-1.40) | $2.97 \times 10^{-4}$ | 1.34 (1.17-1.53) | $3.36 \times 10^{-5}$ |
| CTSH | 1.22 (1.10-1.36) | $3.01 \times 10^{-4}$ | 1.81 (1.51-2.17) | $9.82 \times 10^{-11}$ |
| EPHA2 | 1.26 (1.11-1.44) | $3.68 \times 10^{-4}$ | 1.50 (1.25-1.80) | $1.41 \times 10^{-5}$ |
| CXCL12 | 1.25 (1.10-1.41) | $3.73 \times 10^{-4}$ | 1.55 (1.28-1.87) | $5.65 \times 10^{-6}$ |
| EPO | 1.22 (1.09-1.36) | $4.05 \times 10^{-4}$ | 1.42 (1.23-1.63) | $8.93 \times 10^{-7}$ |
| CHRDL2 | 1.24 (1.10-1.40) | $4.64 \times 10^{-4}$ | 1.43 (1.20-1.70) | $4.91 \times 10^{-5}$ |
| MMP7 | 1.24 (1.10-1.41) | $5.28 \times 10^{-4}$ | 1.53 (1.30-1.80) | $2.16 \times 10^{-7}$ |
| ROBO2 | 1.24 (1.10-1.40) | $5.60 \times 10^{-4}$ | 1.52 (1.29-1.78) | $2.55 \times 10^{-7}$ |
| MFAP4 | 1.21 (1.09-1.35) | $5.92 \times 10^{-4}$ | 1.97 (1.67-2.33) | $2.36 \times 10^{-15}$ |
| CHST15 | 1.24 (1.10-1.40) | $6.02 \times 10^{-4}$ | 1.49 (1.26-1.77) | $3.99 \times 10^{-6}$ |
| SCARA5 | 1.21 (1.08-1.36) | $7.02 \times 10^{-4}$ | 1.27 (1.13-1.43) | $8.31 \times 10^{-5}$ |
| IGF1R | 1.22 (1.09-1.38) | $7.66 \times 10^{-4}$ | 1.47 (1.25-1.74) | $5.01 \times 10^{-6}$ |
| FSTL1 | 1.23 (1.09-1.38) | $8.17 \times 10^{-4}$ | 1.57 (1.34-1.85) | $2.27 \times 10^{-8}$ |
| BMP6 | 1.20 (1.08-1.34) | $8.46 \times 10^{-4}$ | 1.30 (1.14-1.49) | $1.48 \times 10^{-4}$ |
| IL6ST | 1.23 (1.09-1.39) | $8.47 \times 10^{-4}$ | 1.41 (1.18-1.68) | $1.35 \times 10^{-4}$ |
| PTPRU | 1.20 (1.08-1.35) | $1.10 \times 10^{-3}$ | 1.46 (1.24-1.72) | $5.30 \times 10^{-6}$ |
| AXIN2 | 1.21 (1.08-1.36) | $1.13 \times 10^{-3}$ | 1.61 (1.41-1.84) | $2.82 \times 10^{-12}$ |
| AMIGO2 | 1.21 (1.08-1.36) | $1.34 \times 10^{-3}$ | 1.40 (1.20-1.64) | $2.29 \times 10^{-5}$ |
| INHBB | 1.21 (1.08-1.37) | $1.67 \times 10^{-3}$ | 1.43 (1.20-1.70) | $5.19 \times 10^{-5}$ |
| CD93 | 1.21 (1.07-1.37) | $1.76 \times 10^{-3}$ | 1.55 (1.31-1.84) | $5.06 \times 10^{-7}$ |
| CCN1 | 1.15 (1.05-1.25) | $1.83 \times 10^{-3}$ | 1.36 (1.23-1.50) | $2.57 \times 10^{-9}$ |
| RNASE4 | 1.21 (1.07-1.37) | $1.92 \times 10^{-3}$ | 1.53 (1.26-1.86) | $2.43 \times 10^{-5}$ |
| NBL1 | 1.23 (1.08-1.40) | $2.27 \times 10^{-3}$ | 1.49 (1.24-1.81) | $3.24 \times 10^{-5}$ |
| COL18A1 | 1.23 (1.08-1.40) | $2.33 \times 10^{-3}$ | 1.56 (1.29-1.89) | $3.88 \times 10^{-6}$ |

| | | | | |
|---|---|---|---|---|
| ADIPOQ | 1.23 (1.08-1.40) | $2.34 \times 10^{-3}$ | 1.46 (1.23-1.74) | $2.20 \times 10^{-5}$ |
| FGF23 | 1.14 (1.05-1.25) | $2.42 \times 10^{-3}$ | 1.28 (1.13-1.45) | $1.10 \times 10^{-4}$ |
| CCDC80 | 1.22 (1.07-1.40) | $2.47 \times 10^{-3}$ | 1.66 (1.39-1.97) | $1.22 \times 10^{-8}$ |
| LILRB2 | 1.20 (1.07-1.36) | $2.61 \times 10^{-3}$ | 1.42 (1.20-1.69) | $6.74 \times 10^{-5}$ |

**Supplemental Table 8: Proteins Significantly Associated With HF hospitalization in Minimally Adjusted Model**

| Protein | Discovery (ATMOSPHERE) | | Replication (PARADIGM) | |
|---|---|---|---|---|
| | HR (95% CI) | *P* Value | HR (95% CI) | *P* Value |
| GDF15 | 1.54 (1.38-1.71) | $4.38 \times 10^{-15}$ | 1.96 (1.69-2.27) | $4.09 \times 10^{-19}$ |
| SVEP1 | 1.52 (1.37-1.68) | $6.99 \times 10^{-15}$ | 1.70 (1.48-1.95) | $1.51 \times 10^{-14}$ |
| QSOX1 | 1.50 (1.34-1.67) | $9.10 \times 10^{-13}$ | 1.60 (1.40-1.82) | $1.92 \times 10^{-12}$ |
| NPPB | 1.68 (1.46-1.94) | $1.16 \times 10^{-12}$ | 1.72 (1.46-2.02) | $6.91 \times 10^{-11}$ |
| THBS2 | 1.48 (1.33-1.66) | $4.57 \times 10^{-12}$ | 1.75 (1.53-2.00) | $2.45 \times 10^{-16}$ |
| ANGPT2 | 1.48 (1.33-1.66) | $6.60 \times 10^{-12}$ | 1.79 (1.55-2.08) | $1.30 \times 10^{-14}$ |
| IGFBP7 | 1.47 (1.31-1.65) | $8.60 \times 10^{-11}$ | 1.61 (1.40-1.84) | $1.05 \times 10^{-11}$ |
| IL1RL1 | 1.50 (1.33-1.70) | $1.85 \times 10^{-10}$ | 1.50 (1.28-1.76) | $3.76 \times 10^{-7}$ |
| ENG | 1.41 (1.27-1.56) | $1.99 \times 10^{-10}$ | 1.47 (1.27-1.71) | $4.82 \times 10^{-7}$ |
| NFASC | 1.39 (1.25-1.55) | $6.85 \times 10^{-10}$ | 1.69 (1.47-1.94) | $1.28 \times 10^{-13}$ |
| EPO | 1.40 (1.26-1.56) | $1.55 \times 10^{-9}$ | 1.69 (1.48-1.92) | $1.46 \times 10^{-15}$ |
| LTBP4 | 1.46 (1.29-1.65) | $1.78 \times 10^{-9}$ | 1.43 (1.27-1.60) | $1.58 \times 10^{-9}$ |
| ADAMTSL2 | 1.41 (1.26-1.58) | $2.16 \times 10^{-9}$ | 1.60 (1.40-1.83) | $8.10 \times 10^{-12}$ |
| TNC | 1.43 (1.27-1.61) | $3.02 \times 10^{-9}$ | 1.40 (1.23-1.59) | $1.97 \times 10^{-7}$ |
| ATP5PF | 1.32 (1.20-1.45) | $6.26 \times 10^{-9}$ | 1.33 (1.19-1.49) | $8.12 \times 10^{-7}$ |
| SPON2 | 1.43 (1.26-1.61) | $1.26 \times 10^{-8}$ | 1.58 (1.34-1.88) | $1.21 \times 10^{-7}$ |
| FSTL3 | 1.45 (1.28-1.65) | $1.54 \times 10^{-8}$ | 1.59 (1.34-1.88) | $1.28 \times 10^{-7}$ |
| IGSF3 | 1.36 (1.22-1.52) | $5.17 \times 10^{-8}$ | 1.39 (1.21-1.60) | $3.15 \times 10^{-6}$ |
| EFEMP1 | 1.36 (1.22-1.52) | $8.07 \times 10^{-8}$ | 1.34 (1.21-1.50) | $4.92 \times 10^{-8}$ |
| PTPRU | 1.31 (1.19-1.44) | $8.20 \times 10^{-8}$ | 1.42 (1.24-1.62) | $5.35 \times 10^{-7}$ |
| MMP19 | 1.41 (1.24-1.60) | $1.31 \times 10^{-7}$ | 1.60 (1.39-1.83) | $3.10 \times 10^{-11}$ |
| FABP3 | 1.40 (1.24-1.60) | $2.19 \times 10^{-7}$ | 1.50 (1.26-1.78) | $3.88 \times 10^{-6}$ |
| B2M | 1.37 (1.21-1.54) | $3.46 \times 10^{-7}$ | 1.47 (1.24-1.75) | $8.40 \times 10^{-6}$ |
| PXDN | 1.43 (1.24-1.64) | $3.95 \times 10^{-7}$ | 1.54 (1.30-1.82) | $7.54 \times 10^{-7}$ |
| CST3 | 1.35 (1.20-1.52) | $6.90 \times 10^{-7}$ | 1.50 (1.27-1.78) | $2.38 \times 10^{-6}$ |
| SEMA6B | 1.33 (1.19-1.49) | $9.64 \times 10^{-7}$ | 1.29 (1.16-1.42) | $1.29 \times 10^{-6}$ |
| EPHA2 | 1.34 (1.19-1.51) | $1.14 \times 10^{-6}$ | 1.42 (1.20-1.68) | $3.87 \times 10^{-5}$ |
| CXCL13 | 1.29 (1.16-1.43) | $1.41 \times 10^{-6}$ | 1.31 (1.16-1.49) | $2.78 \times 10^{-5}$ |
| RSPO4 | 1.21 (1.12-1.32) | $2.59 \times 10^{-6}$ | 1.37 (1.23-1.52) | $3.89 \times 10^{-9}$ |
| NBL1 | 1.32 (1.17-1.48) | $2.64 \times 10^{-6}$ | 1.38 (1.18-1.62) | $8.64 \times 10^{-5}$ |
| STC1 | 1.33 (1.18-1.50) | $3.33 \times 10^{-6}$ | 1.57 (1.34-1.85) | $3.96 \times 10^{-8}$ |
| TNNI3 | 1.23 (1.12-1.34) | $4.36 \times 10^{-6}$ | 1.29 (1.17-1.42) | $4.64 \times 10^{-7}$ |
| COL28A1 | 1.33 (1.18-1.51) | $6.37 \times 10^{-6}$ | 1.58 (1.34-1.87) | $5.59 \times 10^{-8}$ |
| COL6A3 | 1.30 (1.16-1.46) | $7.05 \times 10^{-6}$ | 1.54 (1.32-1.79) | $1.92 \times 10^{-8}$ |
| PTK7 | 1.31 (1.16-1.47) | $7.16 \times 10^{-6}$ | 1.62 (1.39-1.90) | $8.98 \times 10^{-10}$ |
| CD5 | 1.22 (1.12-1.33) | $7.98 \times 10^{-6}$ | 1.27 (1.16-1.39) | $1.82 \times 10^{-7}$ |
| RNASE1 | 1.33 (1.18-1.52) | $8.63 \times 10^{-6}$ | 1.48 (1.25-1.76) | $6.46 \times 10^{-6}$ |

| | | | | |
|---|---|---|---|---|
| IL19 | 1.32 (1.17-1.48) | $9.34 \times 10^{-6}$ | 1.41 (1.22-1.63) | $2.20 \times 10^{-6}$ |
| SPON1 | 1.29 (1.15-1.45) | $9.98 \times 10^{-6}$ | 1.57 (1.39-1.77) | $3.31 \times 10^{-13}$ |
| CHST15 | 1.32 (1.17-1.49) | $1.09 \times 10^{-5}$ | 1.62 (1.39-1.88) | $3.21 \times 10^{-10}$ |
| DCN | 1.27 (1.14-1.41) | $1.34 \times 10^{-5}$ | 1.42 (1.23-1.65) | $2.50 \times 10^{-6}$ |
| KERA | 1.30 (1.15-1.46) | $1.49 \times 10^{-5}$ | 1.38 (1.23-1.55) | $7.47 \times 10^{-8}$ |
| CTSH | 1.26 (1.14-1.40) | $1.63 \times 10^{-5}$ | 1.56 (1.33-1.82) | $2.19 \times 10^{-8}$ |
| COLEC11 | 1.29 (1.15-1.45) | $2.17 \times 10^{-5}$ | 1.43 (1.23-1.66) | $2.28 \times 10^{-6}$ |
| C1QTNF1 | 1.29 (1.14-1.45) | $2.63 \times 10^{-5}$ | 1.49 (1.30-1.71) | $1.54 \times 10^{-8}$ |
| TNFRSF11B | 1.28 (1.14-1.45) | $4.82 \times 10^{-5}$ | 1.44 (1.22-1.71) | $2.51 \times 10^{-5}$ |
| MMP7 | 1.28 (1.14-1.45) | $5.51 \times 10^{-5}$ | 1.45 (1.26-1.67) | $2.42 \times 10^{-7}$ |
| ICAM5 | 1.28 (1.14-1.45) | $5.70 \times 10^{-5}$ | 1.49 (1.27-1.75) | $1.39 \times 10^{-6}$ |
| TAGLN | 1.33 (1.16-1.53) | $5.81 \times 10^{-5}$ | 1.68 (1.41-2.00) | $6.21 \times 10^{-9}$ |
| PLXDC2 | 1.23 (1.11-1.37) | $7.09 \times 10^{-5}$ | 1.46 (1.26-1.69) | $3.87 \times 10^{-7}$ |
| CILP2 | 0.78 (0.69-0.88) | $7.63 \times 10^{-5}$ | 0.62 (0.53-0.72) | $1.82 \times 10^{-9}$ |
| FGF23 | 1.17 (1.08-1.27) | $8.05 \times 10^{-5}$ | 1.48 (1.34-1.63) | $1.14 \times 10^{-14}$ |
| DNAJB12 | 1.23 (1.11-1.37) | $8.39 \times 10^{-5}$ | 1.30 (1.13-1.48) | $1.34 \times 10^{-4}$ |
| PARVA | 1.24 (1.11-1.39) | $1.17 \times 10^{-4}$ | 1.28 (1.15-1.42) | $4.17 \times 10^{-6}$ |
| CCL14 | 1.29 (1.13-1.47) | $1.26 \times 10^{-4}$ | 1.43 (1.21-1.70) | $3.29 \times 10^{-5}$ |
| FLRT3 | 1.25 (1.11-1.40) | $1.53 \times 10^{-4}$ | 1.32 (1.15-1.51) | $7.65 \times 10^{-5}$ |
| TST | 1.25 (1.11-1.40) | $1.66 \times 10^{-4}$ | 1.42 (1.22-1.65) | $5.68 \times 10^{-6}$ |
| FGF7 | 1.21 (1.09-1.34) | $1.90 \times 10^{-4}$ | 1.37 (1.19-1.58) | $1.06 \times 10^{-5}$ |
| WARS1 | 1.26 (1.12-1.43) | $1.99 \times 10^{-4}$ | 1.35 (1.16-1.58) | $1.18 \times 10^{-4}$ |
| VEGFD | 1.22 (1.10-1.36) | $2.05 \times 10^{-4}$ | 1.39 (1.25-1.55) | $2.60 \times 10^{-9}$ |
| CHRDL2 | 1.25 (1.11-1.41) | $2.36 \times 10^{-4}$ | 1.45 (1.24-1.71) | $5.97 \times 10^{-6}$ |
| GABARAPL1 | 1.22 (1.10-1.36) | $2.44 \times 10^{-4}$ | 1.40 (1.21-1.62) | $5.35 \times 10^{-6}$ |
| CHST9 | 1.19 (1.08-1.31) | $2.61 \times 10^{-4}$ | 1.29 (1.19-1.40) | $1.01 \times 10^{-9}$ |
| FSTL1 | 1.26 (1.11-1.42) | $2.68 \times 10^{-4}$ | 1.41 (1.22-1.63) | $3.86 \times 10^{-6}$ |
| SCARF2 | 1.17 (1.08-1.28) | $3.19 \times 10^{-4}$ | 1.36 (1.17-1.59) | $6.70 \times 10^{-5}$ |
| TFF3 | 1.24 (1.10-1.39) | $4.37 \times 10^{-4}$ | 1.58 (1.35-1.85) | $9.75 \times 10^{-9}$ |
| IGFBP1 | 1.25 (1.10-1.41) | $5.60 \times 10^{-4}$ | 1.40 (1.19-1.64) | $4.47 \times 10^{-5}$ |
| HSPA1A | 1.21 (1.09-1.35) | $5.95 \times 10^{-4}$ | 1.32 (1.14-1.51) | $1.14 \times 10^{-4}$ |
| ITGA1\|ITGB1 | 1.21 (1.08-1.35) | $6.86 \times 10^{-4}$ | 1.28 (1.13-1.46) | $1.09 \times 10^{-4}$ |
| HTRA2 | 1.20 (1.08-1.34) | $7.08 \times 10^{-4}$ | 1.23 (1.10-1.37) | $1.53 \times 10^{-4}$ |
| Igh | 1.22 (1.09-1.37) | $7.73 \times 10^{-4}$ | 1.35 (1.17-1.55) | $2.57 \times 10^{-5}$ |
| NRP1 | 1.22 (1.09-1.37) | $8.09 \times 10^{-4}$ | 1.53 (1.33-1.77) | $2.39 \times 10^{-9}$ |
| TFRC | 1.22 (1.09-1.38) | $9.15 \times 10^{-4}$ | 1.44 (1.24-1.67) | $1.56 \times 10^{-6}$ |
| ROBO2 | 1.23 (1.09-1.39) | $9.60 \times 10^{-4}$ | 1.41 (1.21-1.65) | $1.85 \times 10^{-5}$ |
| BMP6 | 1.20 (1.08-1.34) | $9.62 \times 10^{-4}$ | 1.35 (1.20-1.53) | $1.21 \times 10^{-6}$ |
| CD93 | 1.22 (1.08-1.37) | $9.69 \times 10^{-4}$ | 1.37 (1.17-1.61) | $9.43 \times 10^{-5}$ |
| UMOD | 1.23 (1.09-1.38) | $1.00 \times 10^{-3}$ | 1.34 (1.15-1.55) | $1.23 \times 10^{-4}$ |
| MENT | 0.82 (0.73-0.92) | $1.03 \times 10^{-3}$ | 0.70 (0.60-0.82) | $9.78 \times 10^{-6}$ |

| | | | | |
|---|---|---|---|---|
| CRK | 1.20 (1.08-1.34) | $1.08 \times 10^{-3}$ | 1.28 (1.13-1.45) | $1.11 \times 10^{-4}$ |
| COL18A1 | 1.23 (1.09-1.40) | $1.08 \times 10^{-3}$ | 1.41 (1.19-1.67) | $6.24 \times 10^{-5}$ |
| WFDC2 | 1.24 (1.09-1.41) | $1.10 \times 10^{-3}$ | 1.54 (1.30-1.82) | $3.76 \times 10^{-7}$ |
| CDH7 | 0.82 (0.73-0.92) | $1.18 \times 10^{-3}$ | 0.70 (0.60-0.81) | $1.83 \times 10^{-6}$ |
| MRC2 | 1.21 (1.08-1.37) | $1.32 \times 10^{-3}$ | 1.36 (1.16-1.58) | $1.39 \times 10^{-4}$ |
| WFDC1 | 1.22 (1.08-1.38) | $1.34 \times 10^{-3}$ | 1.48 (1.25-1.75) | $5.63 \times 10^{-6}$ |
| SCARF1 | 1.21 (1.08-1.35) | $1.39 \times 10^{-3}$ | 1.32 (1.15-1.53) | $1.30 \times 10^{-4}$ |
| FCN3 | 0.81 (0.70-0.92) | $1.56 \times 10^{-3}$ | 0.76 (0.67-0.87) | $3.61 \times 10^{-5}$ |
| CXCL8 | 1.17 (1.06-1.30) | $1.64 \times 10^{-3}$ | 1.28 (1.15-1.43) | $4.43 \times 10^{-6}$ |
| SIGLEC7 | 1.21 (1.07-1.36) | $1.66 \times 10^{-3}$ | 1.33 (1.16-1.52) | $5.27 \times 10^{-5}$ |
| PRSS22 | 1.19 (1.07-1.33) | $1.78 \times 10^{-3}$ | 1.39 (1.19-1.62) | $2.99 \times 10^{-5}$ |
| KREMEN1 | 1.20 (1.07-1.35) | $1.81 \times 10^{-3}$ | 1.27 (1.13-1.42) | $3.61 \times 10^{-5}$ |
| FJX1 | 1.20 (1.07-1.35) | $1.82 \times 10^{-3}$ | 1.26 (1.13-1.40) | $4.20 \times 10^{-5}$ |
| CTSB | 1.19 (1.07-1.34) | $1.85 \times 10^{-3}$ | 1.24 (1.11-1.38) | $1.36 \times 10^{-4}$ |
| AGRP | 1.17 (1.06-1.30) | $1.90 \times 10^{-3}$ | 1.34 (1.18-1.51) | $4.83 \times 10^{-6}$ |
| MINPP1 | 1.19 (1.07-1.33) | $2.13 \times 10^{-3}$ | 1.26 (1.12-1.42) | $1.39 \times 10^{-4}$ |
| PRSS2 | 1.21 (1.07-1.37) | $2.23 \times 10^{-3}$ | 1.40 (1.20-1.63) | $2.37 \times 10^{-5}$ |
| CBR1 | 1.19 (1.06-1.34) | $2.63 \times 10^{-3}$ | 1.35 (1.16-1.56) | $7.57 \times 10^{-5}$ |
| LAMC2 | 1.19 (1.06-1.34) | $2.89 \times 10^{-3}$ | 1.34 (1.21-1.47) | $8.06 \times 10^{-9}$ |

**Supplemental Table 9: Proteins Significantly Associated With HF hospitalization in Cardiovascular Risk Factor Adjusted Model**

| Protein | Discovery (ATMOSPHERE) | | Replication (PARADIGM) | |
|---|---|---|---|---|
| | HR (95% CI) | *P* Value | HR (95% CI) | *P* Value |
| SVEP1 | 1.45 (1.29-1.63) | $7.30 \times 10^{-10}$ | 1.56 (1.36-1.80) | $7.26 \times 10^{-10}$ |
| NPPB | 1.57 (1.34-1.84) | $1.49 \times 10^{-8}$ | 1.53 (1.29-1.82) | $1.14 \times 10^{-6}$ |
| QSOX1 | 1.40 (1.24-1.58) | $5.18 \times 10^{-8}$ | 1.45 (1.26-1.66) | $1.17 \times 10^{-7}$ |
| GDF15 | 1.41 (1.24-1.61) | $1.23 \times 10^{-7}$ | 1.67 (1.41-1.99) | $3.73 \times 10^{-9}$ |
| IL1RL1 | 1.43 (1.25-1.65) | $2.29 \times 10^{-7}$ | 1.37 (1.18-1.60) | $5.86 \times 10^{-5}$ |
| THBS2 | 1.39 (1.23-1.57) | $2.83 \times 10^{-7}$ | 1.52 (1.32-1.75) | $7.32 \times 10^{-9}$ |
| IGFBP7 | 1.37 (1.20-1.55) | $1.69 \times 10^{-6}$ | 1.44 (1.25-1.66) | $7.17 \times 10^{-7}$ |
| LTBP4 | 1.39 (1.21-1.60) | $2.27 \times 10^{-6}$ | 1.36 (1.20-1.55) | $3.83 \times 10^{-6}$ |
| ENG | 1.31 (1.17-1.46) | $3.13 \times 10^{-6}$ | 1.33 (1.14-1.55) | $2.68 \times 10^{-4}$ |
| ATP5PF | 1.28 (1.15-1.43) | $5.21 \times 10^{-6}$ | 1.27 (1.12-1.45) | $2.63 \times 10^{-4}$ |
| ANGPT2 | 1.35 (1.18-1.54) | $6.68 \times 10^{-6}$ | 1.53 (1.30-1.79) | $1.44 \times 10^{-7}$ |
| TNC | 1.33 (1.17-1.50) | $7.68 \times 10^{-6}$ | 1.34 (1.16-1.54) | $4.42 \times 10^{-5}$ |
| CXCL13 | 1.28 (1.15-1.44) | $1.49 \times 10^{-5}$ | 1.29 (1.13-1.47) | $1.94 \times 10^{-4}$ |
| NFASC | 1.31 (1.16-1.49) | $1.50 \times 10^{-5}$ | 1.46 (1.25-1.70) | $1.64 \times 10^{-6}$ |
| ADAMTSL2 | 1.32 (1.16-1.49) | $1.83 \times 10^{-5}$ | 1.43 (1.24-1.66) | $1.35 \times 10^{-6}$ |
| EPO | 1.26 (1.12-1.41) | $1.04 \times 10^{-4}$ | 1.47 (1.29-1.67) | $1.26 \times 10^{-8}$ |
| MMP19 | 1.31 (1.14-1.50) | $1.14 \times 10^{-4}$ | 1.43 (1.24-1.66) | $1.70 \times 10^{-6}$ |
| RSPO4 | 1.18 (1.08-1.29) | $3.05 \times 10^{-4}$ | 1.30 (1.15-1.46) | $1.54 \times 10^{-5}$ |
| FGF23 | 1.19 (1.08-1.31) | $3.91 \times 10^{-4}$ | 1.32 (1.18-1.48) | $6.65 \times 10^{-7}$ |
| PTPRU | 1.22 (1.09-1.36) | $4.49 \times 10^{-4}$ | 1.34 (1.14-1.57) | $3.00 \times 10^{-4}$ |

**Supplemental Table 10: Proteins Significantly Associated With All Cause Death in Minimally Adjusted Model**

| | Discovery (ATMOSPHERE) | | Replication (PARADIGM) | |
|---|---|---|---|---|
| **Protein** | **HR (95% CI)** | ***P* Value** | **HR (95% CI)** | ***P* Value** |
| SVEP1 | 1.53 (1.40-1.66) | $1.36 \times 10^{-22}$ | 1.45 (1.28-1.65) | $5.28 \times 10^{-9}$ |
| GDF15 | 1.55 (1.42-1.69) | $7.40 \times 10^{-22}$ | 1.49 (1.30-1.70) | $6.12 \times 10^{-9}$ |
| ANGPT2 | 1.51 (1.37-1.66) | $3.16 \times 10^{-17}$ | 1.62 (1.41-1.86) | $1.46 \times 10^{-11}$ |
| LTBP4 | 1.48 (1.34-1.63) | $2.42 \times 10^{-15}$ | 1.30 (1.17-1.45) | $2.04 \times 10^{-6}$ |
| THBS2 | 1.45 (1.32-1.59) | $2.71 \times 10^{-15}$ | 1.54 (1.36-1.74) | $9.75 \times 10^{-12}$ |
| IGFBP1 | 1.50 (1.36-1.66) | $5.32 \times 10^{-15}$ | 1.58 (1.37-1.81) | $1.52 \times 10^{-10}$ |
| ADAMTSL2 | 1.43 (1.31-1.57) | $4.89 \times 10^{-14}$ | 1.33 (1.17-1.51) | $1.28 \times 10^{-5}$ |
| B2M | 1.45 (1.32-1.60) | $9.87 \times 10^{-14}$ | 1.61 (1.39-1.86) | $1.49 \times 10^{-10}$ |
| NPPB | 1.57 (1.39-1.77) | $1.62 \times 10^{-13}$ | 1.48 (1.26-1.72) | $8.60 \times 10^{-7}$ |
| COL6A3 | 1.42 (1.30-1.57) | $3.34 \times 10^{-13}$ | 1.49 (1.31-1.70) | $3.81 \times 10^{-9}$ |
| SPON1 | 1.40 (1.28-1.53) | $3.91 \times 10^{-13}$ | 1.50 (1.34-1.68) | $3.96 \times 10^{-12}$ |
| RSPO4 | 1.25 (1.17-1.33) | $3.34 \times 10^{-12}$ | 1.37 (1.25-1.50) | $4.03 \times 10^{-11}$ |
| IGFBP2 | 1.32 (1.22-1.43) | $4.28 \times 10^{-12}$ | 1.39 (1.24-1.57) | $2.30 \times 10^{-8}$ |
| C1QTNF1 | 1.41 (1.28-1.56) | $7.23 \times 10^{-12}$ | 1.48 (1.31-1.68) | $6.35 \times 10^{-10}$ |
| SPON2 | 1.44 (1.30-1.60) | $7.84 \times 10^{-12}$ | 1.59 (1.37-1.84) | $6.88 \times 10^{-10}$ |
| EFEMP1 | 1.37 (1.25-1.51) | $4.02 \times 10^{-11}$ | 1.30 (1.18-1.42) | $6.71 \times 10^{-8}$ |
| NFASC | 1.33 (1.22-1.45) | $5.47 \times 10^{-11}$ | 1.43 (1.26-1.63) | $6.38 \times 10^{-8}$ |
| COL28A1 | 1.41 (1.27-1.57) | $5.63 \times 10^{-11}$ | 1.59 (1.37-1.83) | $3.87 \times 10^{-10}$ |
| FSTL3 | 1.44 (1.29-1.61) | $6.12 \times 10^{-11}$ | 1.53 (1.32-1.78) | $4.04 \times 10^{-8}$ |
| CST3 | 1.39 (1.26-1.54) | $7.02 \times 10^{-11}$ | 1.64 (1.42-1.90) | $3.24 \times 10^{-11}$ |
| TNC | 1.42 (1.28-1.58) | $1.06 \times 10^{-10}$ | 1.52 (1.33-1.74) | $1.61 \times 10^{-9}$ |
| WFDC2 | 1.43 (1.28-1.59) | $1.46 \times 10^{-10}$ | 1.58 (1.36-1.82) | $6.85 \times 10^{-10}$ |
| IGFBP7 | 1.38 (1.25-1.52) | $1.49 \times 10^{-10}$ | 1.47 (1.29-1.67) | $8.97 \times 10^{-9}$ |
| CTSH | 1.31 (1.20-1.42) | $2.06 \times 10^{-10}$ | 1.66 (1.46-1.89) | $2.29 \times 10^{-14}$ |
| IL1RL1 | 1.40 (1.26-1.56) | $3.65 \times 10^{-10}$ | 1.45 (1.26-1.68) | $2.76 \times 10^{-7}$ |
| PTK7 | 1.37 (1.24-1.51) | $6.11 \times 10^{-10}$ | 1.57 (1.37-1.81) | $2.19 \times 10^{-10}$ |
| TNNI3 | 1.27 (1.18-1.37) | $7.79 \times 10^{-10}$ | 1.22 (1.11-1.34) | $3.48 \times 10^{-5}$ |
| TAGLN | 1.44 (1.28-1.61) | $1.07 \times 10^{-9}$ | 1.53 (1.31-1.79) | $1.18 \times 10^{-7}$ |
| TFF3 | 1.33 (1.21-1.47) | $3.61 \times 10^{-9}$ | 1.61 (1.39-1.87) | $1.98 \times 10^{-10}$ |
| CNDP1 | 0.75 (0.68-0.82) | $7.47 \times 10^{-9}$ | 0.73 (0.64-0.82) | $5.18 \times 10^{-7}$ |
| TNFRSF1A | 1.34 (1.21-1.48) | $7.60 \times 10^{-9}$ | 1.50 (1.32-1.70) | $3.96 \times 10^{-10}$ |
| SEMA6B | 1.33 (1.21-1.47) | $7.96 \times 10^{-9}$ | 1.22 (1.12-1.34) | $1.45 \times 10^{-5}$ |
| RBL2 | 1.32 (1.20-1.45) | $1.33 \times 10^{-8}$ | 1.56 (1.40-1.74) | $2.51 \times 10^{-16}$ |
| MMP19 | 1.36 (1.22-1.51) | $2.26 \times 10^{-8}$ | 1.52 (1.35-1.72) | $1.73 \times 10^{-11}$ |
| RNASE1 | 1.36 (1.22-1.52) | $2.55 \times 10^{-8}$ | 1.41 (1.22-1.65) | $7.70 \times 10^{-6}$ |
| RNASE6 | 1.33 (1.20-1.47) | $2.78 \times 10^{-8}$ | 1.40 (1.25-1.56) | $1.86 \times 10^{-9}$ |
| MMP7 | 1.34 (1.21-1.48) | $2.97 \times 10^{-8}$ | 1.48 (1.31-1.68) | $3.21 \times 10^{-10}$ |

| | | | | |
|---|---|---|---|---|
| TNFRSF1B | 1.35 (1.21-1.50) | $4.96 \times 10^{-8}$ | 1.53 (1.32-1.78) | $2.51 \times 10^{-8}$ |
| GRN | 1.32 (1.20-1.47) | $5.20 \times 10^{-8}$ | 1.52 (1.32-1.75) | $6.57 \times 10^{-9}$ |
| LAMC2 | 1.28 (1.17-1.40) | $5.32 \times 10^{-8}$ | 1.27 (1.15-1.41) | $3.50 \times 10^{-6}$ |
| TNFRSF11B | 1.32 (1.19-1.46) | $6.49 \times 10^{-8}$ | 1.45 (1.25-1.67) | $5.52 \times 10^{-7}$ |
| EPO | 1.29 (1.18-1.42) | $7.65 \times 10^{-8}$ | 1.48 (1.32-1.67) | $9.86 \times 10^{-11}$ |
| CD5 | 1.21 (1.13-1.30) | $8.13 \times 10^{-8}$ | 1.28 (1.19-1.37) | $4.73 \times 10^{-12}$ |
| Igh | 1.30 (1.18-1.43) | $9.09 \times 10^{-8}$ | 1.42 (1.26-1.60) | $4.94 \times 10^{-9}$ |
| RELT | 1.31 (1.18-1.44) | $1.23 \times 10^{-7}$ | 1.27 (1.14-1.42) | $3.00 \times 10^{-5}$ |
| CD163 | 1.30 (1.18-1.43) | $1.26 \times 10^{-7}$ | 1.40 (1.21-1.61) | $3.54 \times 10^{-6}$ |
| EPHA2 | 1.32 (1.19-1.47) | $1.49 \times 10^{-7}$ | 1.43 (1.25-1.65) | $4.51 \times 10^{-7}$ |
| UMOD | 1.31 (1.18-1.44) | $1.51 \times 10^{-7}$ | 1.52 (1.34-1.72) | $5.24 \times 10^{-11}$ |
| NBL1 | 1.30 (1.18-1.43) | $1.79 \times 10^{-7}$ | 1.38 (1.21-1.57) | $1.07 \times 10^{-6}$ |
| FCGR3B | 1.30 (1.17-1.43) | $2.77 \times 10^{-7}$ | 1.36 (1.18-1.56) | $1.80 \times 10^{-5}$ |
| WARS1 | 1.31 (1.18-1.45) | $2.84 \times 10^{-7}$ | 1.38 (1.21-1.59) | $4.35 \times 10^{-6}$ |
| STC1 | 1.30 (1.17-1.44) | $4.60 \times 10^{-7}$ | 1.39 (1.20-1.61) | $1.41 \times 10^{-5}$ |
| ICAM5 | 1.30 (1.17-1.44) | $4.64 \times 10^{-7}$ | 1.57 (1.36-1.81) | $3.60 \times 10^{-10}$ |
| WFDC1 | 1.30 (1.17-1.44) | $5.06 \times 10^{-7}$ | 1.50 (1.30-1.74) | $5.16 \times 10^{-8}$ |
| TFRC | 1.29 (1.17-1.43) | $5.23 \times 10^{-7}$ | 1.54 (1.35-1.76) | $7.90 \times 10^{-11}$ |
| CKB\|CKM | 0.75 (0.67-0.84) | $5.57 \times 10^{-7}$ | 0.74 (0.63-0.85) | $5.59 \times 10^{-5}$ |
| PPP4R3A | 1.25 (1.15-1.37) | $5.75 \times 10^{-7}$ | 1.36 (1.20-1.55) | $1.61 \times 10^{-6}$ |
| CTSB | 1.30 (1.17-1.44) | $7.28 \times 10^{-7}$ | 1.46 (1.26-1.69) | $4.04 \times 10^{-7}$ |
| B3GAT3 | 1.29 (1.16-1.43) | $9.69 \times 10^{-7}$ | 1.34 (1.17-1.53) | $1.83 \times 10^{-5}$ |
| CD300C | 1.30 (1.17-1.44) | $1.26 \times 10^{-6}$ | 1.41 (1.23-1.63) | $1.53 \times 10^{-6}$ |
| HTRA2 | 1.25 (1.14-1.36) | $1.44 \times 10^{-6}$ | 1.36 (1.26-1.46) | $1.54 \times 10^{-16}$ |
| CLPSL1 | 1.26 (1.14-1.38) | $1.47 \times 10^{-6}$ | 1.35 (1.20-1.50) | $1.49 \times 10^{-7}$ |
| CLSTN3 | 1.21 (1.12-1.31) | $1.56 \times 10^{-6}$ | 1.41 (1.28-1.55) | $3.92 \times 10^{-12}$ |
| MFAP4 | 1.25 (1.14-1.38) | $2.06 \times 10^{-6}$ | 1.87 (1.62-2.17) | $3.78 \times 10^{-17}$ |
| INHBB | 1.27 (1.15-1.40) | $2.48 \times 10^{-6}$ | 1.38 (1.20-1.58) | $6.67 \times 10^{-6}$ |
| PIGR | 1.27 (1.15-1.40) | $2.78 \times 10^{-6}$ | 1.43 (1.25-1.62) | $1.14 \times 10^{-7}$ |
| CHST15 | 1.28 (1.16-1.43) | $2.79 \times 10^{-6}$ | 1.45 (1.26-1.67) | $2.72 \times 10^{-7}$ |
| CCL15 | 1.26 (1.14-1.38) | $2.87 \times 10^{-6}$ | 1.30 (1.14-1.48) | $6.24 \times 10^{-5}$ |
| CHRDL1 | 1.29 (1.16-1.44) | $3.05 \times 10^{-6}$ | 1.58 (1.35-1.86) | $2.35 \times 10^{-8}$ |
| CCL14 | 1.29 (1.16-1.44) | $3.88 \times 10^{-6}$ | 1.49 (1.28-1.74) | $3.41 \times 10^{-7}$ |
| KLK10 | 1.27 (1.15-1.41) | $4.48 \times 10^{-6}$ | 1.34 (1.17-1.53) | $2.26 \times 10^{-5}$ |
| PTPRU | 1.24 (1.13-1.36) | $4.94 \times 10^{-6}$ | 1.32 (1.16-1.51) | $2.42 \times 10^{-5}$ |
| PRSS22 | 1.24 (1.13-1.35) | $5.52 \times 10^{-6}$ | 1.33 (1.16-1.52) | $5.34 \times 10^{-5}$ |
| TFF2 | 1.23 (1.12-1.34) | $6.30 \times 10^{-6}$ | 1.32 (1.19-1.45) | $7.60 \times 10^{-8}$ |
| SLAMF6 | 1.20 (1.11-1.29) | $6.50 \times 10^{-6}$ | 1.24 (1.12-1.37) | $2.80 \times 10^{-5}$ |
| LUM | 1.26 (1.14-1.39) | $6.57 \times 10^{-6}$ | 1.38 (1.21-1.57) | $1.33 \times 10^{-6}$ |
| RSPO3 | 1.20 (1.11-1.30) | $6.98 \times 10^{-6}$ | 1.22 (1.12-1.32) | $1.93 \times 10^{-6}$ |
| NRP2 | 1.26 (1.14-1.40) | $7.15 \times 10^{-6}$ | 1.36 (1.19-1.54) | $2.95 \times 10^{-6}$ |
| PLXDC2 | 1.22 (1.12-1.33) | $7.15 \times 10^{-6}$ | 1.64 (1.45-1.86) | $7.65 \times 10^{-15}$ |

| | | | | |
|---|---|---|---|---|
| GAS1 | 1.25 (1.13-1.37) | $7.20 \times 10^{-6}$ | 1.27 (1.14-1.41) | $1.62 \times 10^{-5}$ |
| OLFM1 | 1.19 (1.10-1.28) | $9.27 \times 10^{-6}$ | 1.23 (1.11-1.36) | $6.00 \times 10^{-5}$ |
| ROBO2 | 1.26 (1.14-1.40) | $9.96 \times 10^{-6}$ | 1.46 (1.28-1.66) | $1.09 \times 10^{-8}$ |
| IGHA1\|IGHA2 | 1.25 (1.13-1.37) | $1.09 \times 10^{-5}$ | 1.34 (1.17-1.54) | $2.45 \times 10^{-5}$ |
| FJX1 | 1.24 (1.13-1.37) | $1.19 \times 10^{-5}$ | 1.25 (1.13-1.39) | $1.46 \times 10^{-5}$ |
| MMP2 | 1.27 (1.14-1.41) | $1.20 \times 10^{-5}$ | 1.40 (1.21-1.62) | $6.56 \times 10^{-6}$ |
| FGF23 | 1.16 (1.08-1.23) | $1.49 \times 10^{-5}$ | 1.35 (1.22-1.49) | $4.09 \times 10^{-9}$ |
| IL6ST | 1.25 (1.13-1.39) | $1.56 \times 10^{-5}$ | 1.42 (1.23-1.65) | $3.62 \times 10^{-6}$ |
| COL18A1 | 1.26 (1.14-1.41) | $1.59 \times 10^{-5}$ | 1.52 (1.31-1.77) | $3.36 \times 10^{-8}$ |
| CRP | 1.27 (1.14-1.42) | $1.65 \times 10^{-5}$ | 1.37 (1.18-1.58) | $2.95 \times 10^{-5}$ |
| CRELD1 | 1.27 (1.14-1.42) | $1.78 \times 10^{-5}$ | 1.35 (1.17-1.55) | $3.73 \times 10^{-5}$ |
| IL15RA | 1.24 (1.12-1.36) | $1.79 \times 10^{-5}$ | 1.32 (1.20-1.44) | $4.13 \times 10^{-9}$ |
| AGRP | 1.20 (1.10-1.30) | $2.12 \times 10^{-5}$ | 1.29 (1.14-1.45) | $3.05 \times 10^{-5}$ |
| JAML | 1.25 (1.13-1.38) | $2.47 \times 10^{-5}$ | 1.37 (1.19-1.57) | $1.20 \times 10^{-5}$ |
| SIGLEC7 | 1.23 (1.12-1.36) | $2.55 \times 10^{-5}$ | 1.48 (1.31-1.67) | $3.19 \times 10^{-10}$ |
| CXCL12 | 1.25 (1.13-1.39) | $2.67 \times 10^{-5}$ | 1.50 (1.28-1.75) | $3.51 \times 10^{-7}$ |
| SCARA5 | 1.23 (1.11-1.35) | $2.85 \times 10^{-5}$ | 1.37 (1.24-1.50) | $1.47 \times 10^{-10}$ |
| CD48 | 1.19 (1.10-1.29) | $2.88 \times 10^{-5}$ | 1.46 (1.28-1.66) | $2.51 \times 10^{-8}$ |
| CCN1 | 1.17 (1.09-1.27) | $3.27 \times 10^{-5}$ | 1.39 (1.28-1.50) | $8.04 \times 10^{-16}$ |
| BMP6 | 1.22 (1.11-1.33) | $3.41 \times 10^{-5}$ | 1.33 (1.18-1.50) | $2.77 \times 10^{-6}$ |
| HPSE | 1.23 (1.12-1.36) | $3.75 \times 10^{-5}$ | 1.42 (1.23-1.64) | $1.63 \times 10^{-6}$ |
| PLXNB2 | 1.23 (1.11-1.36) | $3.97 \times 10^{-5}$ | 1.35 (1.17-1.57) | $4.58 \times 10^{-5}$ |
| CKM | 0.79 (0.71-0.89) | $4.07 \times 10^{-5}$ | 0.70 (0.59-0.83) | $3.84 \times 10^{-5}$ |
| CHGB | 1.24 (1.12-1.37) | $4.26 \times 10^{-5}$ | 1.30 (1.15-1.47) | $3.60 \times 10^{-5}$ |
| NRP1 | 1.22 (1.11-1.35) | $4.30 \times 10^{-5}$ | 1.33 (1.17-1.52) | $1.71 \times 10^{-5}$ |
| CHRDL2 | 1.24 (1.12-1.37) | $4.44 \times 10^{-5}$ | 1.40 (1.21-1.62) | $8.33 \times 10^{-6}$ |
| RNASE4 | 1.24 (1.12-1.37) | $5.49 \times 10^{-5}$ | 1.53 (1.31-1.78) | $6.98 \times 10^{-8}$ |
| TXNDC5 | 1.21 (1.10-1.33) | $5.70 \times 10^{-5}$ | 1.57 (1.36-1.81) | $1.07 \times 10^{-9}$ |
| BID | 1.18 (1.09-1.28) | $6.92 \times 10^{-5}$ | 1.23 (1.12-1.36) | $2.13 \times 10^{-5}$ |
| EPHB2 | 1.24 (1.11-1.38) | $7.72 \times 10^{-5}$ | 1.34 (1.17-1.54) | $2.61 \times 10^{-5}$ |
| CD93 | 1.23 (1.11-1.36) | $8.85 \times 10^{-5}$ | 1.47 (1.28-1.70) | $9.13 \times 10^{-8}$ |
| VEGFD | 1.20 (1.09-1.31) | $9.49 \times 10^{-5}$ | 1.47 (1.33-1.63) | $1.32 \times 10^{-14}$ |
| H6PD | 1.22 (1.10-1.35) | $1.14 \times 10^{-4}$ | 1.39 (1.21-1.60) | $2.01 \times 10^{-6}$ |
| LMAN2 | 1.24 (1.11-1.38) | $1.17 \times 10^{-4}$ | 1.38 (1.19-1.61) | $2.63 \times 10^{-5}$ |
| MMP12 | 1.24 (1.11-1.38) | $1.22 \times 10^{-4}$ | 1.44 (1.25-1.67) | $5.65 \times 10^{-7}$ |
| IGFLR1 | 1.19 (1.09-1.29) | $1.24 \times 10^{-4}$ | 1.32 (1.17-1.48) | $4.91 \times 10^{-6}$ |
| CCDC80 | 1.24 (1.11-1.38) | $1.35 \times 10^{-4}$ | 1.61 (1.39-1.86) | $1.27 \times 10^{-10}$ |
| TGFB3 | 1.18 (1.08-1.28) | $1.36 \times 10^{-4}$ | 1.31 (1.20-1.43) | $6.82 \times 10^{-10}$ |
| ITGA1\|ITGB1 | 1.20 (1.09-1.33) | $1.59 \times 10^{-4}$ | 1.30 (1.16-1.47) | $1.62 \times 10^{-5}$ |
| UNC5C | 1.24 (1.11-1.38) | $1.62 \times 10^{-4}$ | 1.54 (1.32-1.79) | $4.42 \times 10^{-8}$ |
| ANGPTL1 | 1.20 (1.09-1.31) | $1.81 \times 10^{-4}$ | 1.41 (1.22-1.63) | $2.69 \times 10^{-6}$ |
| GNPTG | 1.16 (1.07-1.25) | $1.88 \times 10^{-4}$ | 1.52 (1.35-1.71) | $3.41 \times 10^{-12}$ |

| | | | | |
|---|---|---|---|---|
| SLPI | 1.22 (1.10-1.35) | $2.28 \times 10^{-4}$ | 1.39 (1.21-1.59) | $2.37 \times 10^{-6}$ |
| MRC1 | 1.21 (1.09-1.34) | $2.34 \times 10^{-4}$ | 1.52 (1.33-1.74) | $1.08 \times 10^{-9}$ |
| OIT3 | 1.16 (1.07-1.25) | $2.36 \times 10^{-4}$ | 1.23 (1.14-1.32) | $3.87 \times 10^{-8}$ |
| PLA2G2A | 1.21 (1.09-1.34) | $2.71 \times 10^{-4}$ | 1.54 (1.35-1.76) | $2.57 \times 10^{-10}$ |
| LILRB2 | 1.21 (1.09-1.34) | $2.83 \times 10^{-4}$ | 1.38 (1.19-1.60) | $1.94 \times 10^{-5}$ |
| NELL2 | 1.22 (1.10-1.36) | $2.88 \times 10^{-4}$ | 1.21 (1.10-1.33) | $6.13 \times 10^{-5}$ |
| EPHA10 | 1.16 (1.07-1.25) | $3.05 \times 10^{-4}$ | 1.24 (1.12-1.38) | $4.20 \times 10^{-5}$ |
| APOF | 1.24 (1.10-1.40) | $3.27 \times 10^{-4}$ | 1.59 (1.35-1.88) | $3.94 \times 10^{-8}$ |
| CRIP2 | 1.17 (1.07-1.27) | $3.75 \times 10^{-4}$ | 1.22 (1.14-1.32) | $8.79 \times 10^{-8}$ |
| UNC5B | 1.21 (1.09-1.34) | $3.83 \times 10^{-4}$ | 1.36 (1.23-1.51) | $1.50 \times 10^{-9}$ |
| IL15 | 1.18 (1.08-1.29) | $3.91 \times 10^{-4}$ | 1.34 (1.19-1.52) | $2.30 \times 10^{-6}$ |
| IGF1R | 1.20 (1.08-1.32) | $4.27 \times 10^{-4}$ | 1.35 (1.17-1.55) | $2.48 \times 10^{-5}$ |
| AMIGO2 | 1.20 (1.08-1.33) | $4.41 \times 10^{-4}$ | 1.40 (1.23-1.60) | $4.41 \times 10^{-7}$ |
| FSTL1 | 1.20 (1.09-1.34) | $4.44 \times 10^{-4}$ | 1.52 (1.34-1.72) | $1.28 \times 10^{-10}$ |
| LRTM2 | 0.83 (0.75-0.92) | $5.88 \times 10^{-4}$ | 1.26 (1.14-1.40) | $7.63 \times 10^{-6}$ |
| TEK | 1.18 (1.07-1.29) | $6.23 \times 10^{-4}$ | 0.65 (0.55-0.77) | $3.29 \times 10^{-7}$ |
| C9 | 1.20 (1.08-1.34) | $6.77 \times 10^{-4}$ | 1.52 (1.31-1.77) | $5.52 \times 10^{-8}$ |
| MATN2 | 1.21 (1.08-1.34) | $7.10 \times 10^{-4}$ | 1.41 (1.21-1.65) | $1.24 \times 10^{-5}$ |
| LILRA5 | 1.20 (1.08-1.33) | $7.61 \times 10^{-4}$ | 1.33 (1.17-1.52) | $2.03 \times 10^{-5}$ |
| CA11 | 1.14 (1.05-1.24) | $1.07 \times 10^{-3}$ | 1.32 (1.16-1.50) | $1.80 \times 10^{-5}$ |
| MSMB | 1.19 (1.07-1.32) | $1.09 \times 10^{-3}$ | 1.38 (1.19-1.59) | $1.09 \times 10^{-5}$ |
| F8 | 1.19 (1.07-1.33) | $1.17 \times 10^{-3}$ | 1.43 (1.22-1.67) | $7.68 \times 10^{-6}$ |
| MENT | 0.84 (0.76-0.93) | $1.22 \times 10^{-3}$ | 0.68 (0.58-0.79) | $3.24 \times 10^{-7}$ |
| C7 | 1.20 (1.07-1.34) | $1.25 \times 10^{-3}$ | 1.38 (1.18-1.60) | $3.07 \times 10^{-5}$ |
| CCL21 | 1.18 (1.06-1.30) | $1.71 \times 10^{-3}$ | 1.37 (1.20-1.58) | $6.20 \times 10^{-6}$ |
| ADAM12 | 1.14 (1.05-1.23) | $1.89 \times 10^{-3}$ | 1.30 (1.17-1.45) | $1.82 \times 10^{-6}$ |
| PCDHGA12 | 1.13 (1.05-1.22) | $1.92 \times 10^{-3}$ | 1.32 (1.24-1.42) | $1.71 \times 10^{-16}$ |
| ADIPOQ | 1.18 (1.06-1.31) | $1.97 \times 10^{-3}$ | 1.37 (1.20-1.56) | $4.22 \times 10^{-6}$ |
| TMED10 | 1.15 (1.05-1.25) | $2.03 \times 10^{-3}$ | 1.60 (1.41-1.82) | $5.90 \times 10^{-13}$ |
| IGHG1\|IGHG2\|IGHG3\|IGHG4\|IGK\|IGL | 1.18 (1.06-1.30) | $2.04 \times 10^{-3}$ | 1.39 (1.21-1.60) | $3.55 \times 10^{-6}$ |
| S100A13 | 1.17 (1.06-1.29) | $2.08 \times 10^{-3}$ | 1.36 (1.18-1.57) | $1.65 \times 10^{-5}$ |
| GPNMB | 1.17 (1.06-1.29) | $2.22 \times 10^{-3}$ | 1.45 (1.27-1.67) | $8.76 \times 10^{-8}$ |
| TREM2 | 1.18 (1.06-1.30) | $2.31 \times 10^{-3}$ | 1.40 (1.22-1.62) | $3.16 \times 10^{-6}$ |
| ADSS2 | 1.15 (1.05-1.25) | $2.47 \times 10^{-3}$ | 1.27 (1.15-1.40) | $4.81 \times 10^{-6}$ |
| ESM1 | 1.15 (1.05-1.26) | $2.59 \times 10^{-3}$ | 1.18 (1.09-1.28) | $4.15 \times 10^{-5}$ |
| TNFRSF21 | 1.17 (1.06-1.30) | $2.77 \times 10^{-3}$ | 1.39 (1.20-1.62) | $1.27 \times 10^{-5}$ |
| CD59 | 1.12 (1.04-1.20) | $2.85 \times 10^{-3}$ | 1.39 (1.24-1.54) | $4.55 \times 10^{-9}$ |
| IL18R1 | 1.14 (1.05-1.25) | $2.88 \times 10^{-3}$ | 1.31 (1.15-1.50) | $5.97 \times 10^{-5}$ |
| PAPPA | 1.17 (1.06-1.31) | $2.95 \times 10^{-3}$ | 1.50 (1.30-1.74) | $4.82 \times 10^{-8}$ |
| NPDC1 | 1.16 (1.05-1.28) | $3.02 \times 10^{-3}$ | 1.41 (1.26-1.57) | $6.01 \times 10^{-10}$ |

| | | | | |
|---|---|---|---|---|
| C1QL1 | 1.16 (1.05-1.28) | $3.22 \times 10^{-3}$ | 1.26 (1.14-1.39) | $1.07 \times 10^{-5}$ |
| SCARF1 | 1.16 (1.05-1.28) | $3.70 \times 10^{-3}$ | 1.43 (1.26-1.63) | $3.56 \times 10^{-8}$ |
| EDN1 | 1.12 (1.04-1.22) | $3.88 \times 10^{-3}$ | 1.32 (1.18-1.47) | $6.63 \times 10^{-7}$ |
| ST6GAL1 | 1.13 (1.04-1.22) | $5.18 \times 10^{-3}$ | 1.27 (1.14-1.41) | $6.99 \times 10^{-6}$ |
| EFNA4 | 1.13 (1.04-1.23) | $5.61 \times 10^{-3}$ | 1.32 (1.19-1.46) | $1.68 \times 10^{-7}$ |
| TMX3 | 1.14 (1.04-1.26) | $7.28 \times 10^{-3}$ | 1.35 (1.18-1.55) | $1.42 \times 10^{-5}$ |
| MINPP1 | 1.14 (1.04-1.26) | $7.30 \times 10^{-3}$ | 1.46 (1.30-1.63) | $3.76 \times 10^{-11}$ |
| SDF2L1 | 1.13 (1.03-1.23) | $7.58 \times 10^{-3}$ | 1.32 (1.19-1.45) | $6.22 \times 10^{-8}$ |
| TWSG1 | 1.15 (1.04-1.27) | $7.59 \times 10^{-3}$ | 1.29 (1.17-1.42) | $5.79 \times 10^{-7}$ |
| PLA2R1 | 1.14 (1.03-1.25) | $7.60 \times 10^{-3}$ | 1.36 (1.19-1.54) | $2.87 \times 10^{-6}$ |

**Supplemental Table 11: Proteins Significantly Associated With All Cause Death in Cardiovascular Risk Factor Adjusted Model**

| | Discovery (ATMOSPHERE) | | Replication (PARADIGM) | |
|---|---|---|---|---|
| **Protein** | **HR (95% CI)** | ***P* Value** | **HR (95% CI)** | ***P* Value** |
| SVEP1 | 1.54 (1.40-1.70) | $1.24 \times 10^{-18}$ | 1.34 (1.17-1.54) | $3.21 \times 10^{-5}$ |
| GDF15 | 1.52 (1.36-1.69) | $2.78 \times 10^{-14}$ | 1.41 (1.21-1.64) | $1.62 \times 10^{-5}$ |
| LTBP4 | 1.52 (1.36-1.70) | $3.32 \times 10^{-13}$ | 1.30 (1.15-1.46) | $2.59 \times 10^{-5}$ |
| ANGPT2 | 1.49 (1.33-1.67) | $3.91 \times 10^{-12}$ | 1.51 (1.30-1.76) | $1.10 \times 10^{-7}$ |
| THBS2 | 1.42 (1.28-1.57) | $1.61 \times 10^{-11}$ | 1.46 (1.27-1.67) | $4.97 \times 10^{-8}$ |
| IGFBP1 | 1.49 (1.32-1.67) | $1.93 \times 10^{-11}$ | 1.59 (1.36-1.86) | $3.41 \times 10^{-9}$ |
| B2M | 1.47 (1.30-1.66) | $5.94 \times 10^{-10}$ | 1.76 (1.48-2.08) | $1.26 \times 10^{-10}$ |
| IGFBP2 | 1.32 (1.21-1.44) | $1.03 \times 10^{-9}$ | 1.42 (1.24-1.62) | $2.54 \times 10^{-7}$ |
| SPON1 | 1.38 (1.25-1.54) | $1.26 \times 10^{-9}$ | 1.42 (1.25-1.62) | $1.50 \times 10^{-7}$ |
| COL6A3 | 1.41 (1.26-1.58) | $1.42 \times 10^{-9}$ | 1.48 (1.28-1.71) | $1.33 \times 10^{-7}$ |
| SPON2 | 1.42 (1.27-1.59) | $1.66 \times 10^{-9}$ | 1.57 (1.34-1.85) | $3.05 \times 10^{-8}$ |
| C1QTNF1 | 1.37 (1.23-1.51) | $3.12 \times 10^{-9}$ | 1.46 (1.28-1.68) | $5.24 \times 10^{-8}$ |
| CST3 | 1.45 (1.28-1.65) | $5.96 \times 10^{-9}$ | 1.94 (1.61-2.33) | $1.82 \times 10^{-12}$ |
| IL1RL1 | 1.41 (1.26-1.59) | $7.85 \times 10^{-9}$ | 1.41 (1.21-1.63) | $6.13 \times 10^{-6}$ |
| FSTL3 | 1.44 (1.27-1.64) | $1.05 \times 10^{-8}$ | 1.49 (1.26-1.77) | $2.60 \times 10^{-6}$ |
| TNC | 1.40 (1.25-1.57) | $1.23 \times 10^{-8}$ | 1.47 (1.27-1.69) | $1.60 \times 10^{-7}$ |
| COL28A1 | 1.42 (1.26-1.60) | $1.46 \times 10^{-8}$ | 1.58 (1.35-1.85) | $1.90 \times 10^{-8}$ |
| CNDP1 | 0.74 (0.67-0.82) | $1.48 \times 10^{-8}$ | 0.75 (0.66-0.86) | $3.11 \times 10^{-5}$ |
| NFASC | 1.33 (1.21-1.47) | $1.94 \times 10^{-8}$ | 1.34 (1.17-1.55) | $4.38 \times 10^{-5}$ |
| IGFBP7 | 1.37 (1.23-1.53) | $1.95 \times 10^{-8}$ | 1.39 (1.21-1.59) | $2.76 \times 10^{-6}$ |
| EFEMP1 | 1.34 (1.21-1.48) | $2.33 \times 10^{-8}$ | 1.24 (1.11-1.37) | $6.12 \times 10^{-5}$ |
| TAGLN | 1.42 (1.25-1.61) | $1.14 \times 10^{-7}$ | 1.48 (1.25-1.76) | $7.88 \times 10^{-6}$ |
| Igh | 1.31 (1.18-1.44) | $1.41 \times 10^{-7}$ | 1.37 (1.21-1.55) | $5.33 \times 10^{-7}$ |
| RBL2 | 1.31 (1.18-1.45) | $2.22 \times 10^{-7}$ | 1.47 (1.31-1.64) | $4.76 \times 10^{-11}$ |
| RNASE1 | 1.44 (1.25-1.65) | $2.74 \times 10^{-7}$ | 1.52 (1.27-1.83) | $7.17 \times 10^{-6}$ |
| PTK7 | 1.33 (1.19-1.48) | $2.78 \times 10^{-7}$ | 1.47 (1.27-1.70) | $3.00 \times 10^{-7}$ |
| TNFRSF11B | 1.33 (1.19-1.48) | $3.07 \times 10^{-7}$ | 1.39 (1.19-1.61) | $2.01 \times 10^{-5}$ |
| WFDC2 | 1.39 (1.22-1.57) | $3.09 \times 10^{-7}$ | 1.55 (1.32-1.81) | $5.51 \times 10^{-8}$ |
| RSPO4 | 1.21 (1.12-1.30) | $3.43 \times 10^{-7}$ | 1.31 (1.18-1.45) | $5.94 \times 10^{-7}$ |
| CD163 | 1.31 (1.18-1.46) | $6.24 \times 10^{-7}$ | 1.36 (1.18-1.57) | $2.90 \times 10^{-5}$ |
| CTSB | 1.32 (1.18-1.48) | $1.60 \times 10^{-6}$ | 1.39 (1.19-1.62) | $1.97 \times 10^{-5}$ |
| MMP19 | 1.32 (1.18-1.48) | $1.77 \times 10^{-6}$ | 1.51 (1.32-1.72) | $1.60 \times 10^{-9}$ |
| TNFRSF1B | 1.34 (1.19-1.51) | $2.42 \times 10^{-6}$ | 1.53 (1.29-1.81) | $6.54 \times 10^{-7}$ |
| GRN | 1.30 (1.17-1.45) | $2.48 \times 10^{-6}$ | 1.43 (1.24-1.65) | $1.27 \times 10^{-6}$ |
| TFF3 | 1.32 (1.17-1.48) | $2.58 \times 10^{-6}$ | 1.70 (1.42-2.03) | $6.14 \times 10^{-9}$ |
| MMP7 | 1.31 (1.17-1.46) | $3.16 \times 10^{-6}$ | 1.54 (1.34-1.77) | $7.15 \times 10^{-10}$ |
| WARS1 | 1.30 (1.16-1.46) | $3.95 \times 10^{-6}$ | 1.34 (1.17-1.54) | $3.48 \times 10^{-5}$ |

| | | | | |
|---|---|---|---|---|
| TNFRSF1A | 1.30 (1.16-1.45) | $4.72 \times 10^{-6}$ | 1.49 (1.30-1.70) | $7.38 \times 10^{-9}$ |
| EPHA2 | 1.31 (1.17-1.48) | $4.95 \times 10^{-6}$ | 1.42 (1.22-1.66) | $1.05 \times 10^{-5}$ |
| RNASE6 | 1.30 (1.16-1.46) | $5.52 \times 10^{-6}$ | 1.39 (1.24-1.57) | $6.74 \times 10^{-8}$ |
| CLPSL1 | 1.25 (1.13-1.39) | $1.02 \times 10^{-5}$ | 1.32 (1.17-1.49) | $4.51 \times 10^{-6}$ |
| CD300C | 1.29 (1.15-1.45) | $1.43 \times 10^{-5}$ | 1.44 (1.24-1.68) | $2.43 \times 10^{-6}$ |
| CD5 | 1.19 (1.10-1.29) | $1.67 \times 10^{-5}$ | 1.28 (1.18-1.39) | $5.27 \times 10^{-9}$ |
| RSPO3 | 1.21 (1.11-1.33) | $1.84 \times 10^{-5}$ | 1.23 (1.12-1.34) | $7.66 \times 10^{-6}$ |
| CTSH | 1.24 (1.12-1.37) | $2.01 \times 10^{-5}$ | 1.78 (1.53-2.07) | $7.11 \times 10^{-14}$ |
| CLSTN3 | 1.21 (1.11-1.31) | $2.04 \times 10^{-5}$ | 1.36 (1.22-1.51) | $7.86 \times 10^{-9}$ |
| TFRC | 1.26 (1.13-1.40) | $2.18 \times 10^{-5}$ | 1.41 (1.23-1.61) | $8.33 \times 10^{-7}$ |
| PPP4R3A | 1.23 (1.12-1.36) | $2.45 \times 10^{-5}$ | 1.33 (1.17-1.52) | $2.49 \times 10^{-5}$ |
| ICAM5 | 1.26 (1.13-1.42) | $4.43 \times 10^{-5}$ | 1.48 (1.28-1.71) | $1.53 \times 10^{-7}$ |
| NBL1 | 1.28 (1.14-1.44) | $4.82 \times 10^{-5}$ | 1.47 (1.26-1.72) | $8.89 \times 10^{-7}$ |
| WFDC1 | 1.26 (1.12-1.40) | $5.60 \times 10^{-5}$ | 1.49 (1.27-1.74) | $1.07 \times 10^{-6}$ |
| HTRA2 | 1.21 (1.10-1.33) | $6.06 \times 10^{-5}$ | 1.34 (1.24-1.46) | $2.23 \times 10^{-12}$ |
| SCARA5 | 1.23 (1.11-1.37) | $6.06 \times 10^{-5}$ | 1.28 (1.15-1.42) | $2.38 \times 10^{-6}$ |
| UMOD | 1.24 (1.12-1.39) | $7.50 \times 10^{-5}$ | 1.45 (1.27-1.66) | $4.04 \times 10^{-8}$ |
| CXCL12 | 1.25 (1.12-1.40) | $8.52 \times 10^{-5}$ | 1.46 (1.24-1.72) | $3.75 \times 10^{-6}$ |
| PLXDC2 | 1.21 (1.10-1.34) | $9.36 \times 10^{-5}$ | 1.53 (1.35-1.74) | $4.94 \times 10^{-11}$ |
| CHST15 | 1.25 (1.12-1.40) | $9.71 \times 10^{-5}$ | 1.36 (1.18-1.58) | $3.74 \times 10^{-5}$ |
| HPSE | 1.23 (1.11-1.36) | $1.02 \times 10^{-4}$ | 1.40 (1.21-1.61) | $4.52 \times 10^{-6}$ |
| PIGR | 1.24 (1.11-1.38) | $1.13 \times 10^{-4}$ | 1.35 (1.18-1.54) | $1.16 \times 10^{-5}$ |
| CCL14 | 1.28 (1.13-1.44) | $1.14 \times 10^{-4}$ | 1.45 (1.23-1.71) | $8.10 \times 10^{-6}$ |
| TFF2 | 1.21 (1.10-1.34) | $1.59 \times 10^{-4}$ | 1.34 (1.19-1.50) | $5.58 \times 10^{-7}$ |
| MFAP4 | 1.21 (1.10-1.34) | $1.93 \times 10^{-4}$ | 1.83 (1.58-2.12) | $5.45 \times 10^{-16}$ |
| IL15RA | 1.22 (1.10-1.36) | $2.15 \times 10^{-4}$ | 1.29 (1.17-1.42) | $3.52 \times 10^{-7}$ |
| CCN1 | 1.16 (1.07-1.25) | $2.48 \times 10^{-4}$ | 1.37 (1.26-1.50) | $1.53 \times 10^{-12}$ |
| GNPTG | 1.16 (1.07-1.26) | $2.48 \times 10^{-4}$ | 1.49 (1.32-1.70) | $4.81 \times 10^{-10}$ |
| TXNDC5 | 1.21 (1.09-1.34) | $2.65 \times 10^{-4}$ | 1.58 (1.35-1.85) | $8.77 \times 10^{-9}$ |
| CD48 | 1.17 (1.07-1.27) | $2.70 \times 10^{-4}$ | 1.45 (1.25-1.67) | $3.75 \times 10^{-7}$ |
| MRC1 | 1.22 (1.10-1.35) | $2.78 \times 10^{-4}$ | 1.46 (1.28-1.68) | $5.75 \times 10^{-8}$ |
| EPO | 1.20 (1.09-1.33) | $3.46 \times 10^{-4}$ | 1.36 (1.20-1.53) | $7.42 \times 10^{-7}$ |
| CHRDL1 | 1.25 (1.11-1.41) | $3.47 \times 10^{-4}$ | 1.60 (1.34-1.91) | $1.85 \times 10^{-7}$ |
| S100A13 | 1.22 (1.10-1.37) | $3.55 \times 10^{-4}$ | 1.36 (1.17-1.58) | $5.11 \times 10^{-5}$ |
| OIT3 | 1.17 (1.07-1.28) | $3.55 \times 10^{-4}$ | 1.27 (1.16-1.38) | $6.61 \times 10^{-8}$ |
| CRIP2 | 1.17 (1.07-1.28) | $3.85 \times 10^{-4}$ | 1.18 (1.10-1.28) | $1.19 \times 10^{-5}$ |
| ADIPOQ | 1.24 (1.10-1.41) | $3.91 \times 10^{-4}$ | 1.48 (1.28-1.72) | $1.62 \times 10^{-7}$ |
| LILRB2 | 1.22 (1.09-1.36) | $4.07 \times 10^{-4}$ | 1.35 (1.16-1.56) | $7.96 \times 10^{-5}$ |
| INHBB | 1.22 (1.09-1.36) | $4.32 \times 10^{-4}$ | 1.36 (1.17-1.57) | $5.03 \times 10^{-5}$ |
| LRTM2 | 0.82 (0.74-0.92) | $4.93 \times 10^{-4}$ | 1.27 (1.14-1.42) | $1.63 \times 10^{-5}$ |
| ROBO2 | 1.22 (1.09-1.36) | $6.09 \times 10^{-4}$ | 1.42 (1.24-1.63) | $5.92 \times 10^{-7}$ |

| | | | | |
|---|---|---|---|---|
| COL18A1 | 1.23 (1.09-1.39) | $7.24 \times 10^{-4}$ | 1.51 (1.28-1.77) | $6.44 \times 10^{-7}$ |
| FSTL1 | 1.21 (1.08-1.35) | $7.58 \times 10^{-4}$ | 1.50 (1.30-1.72) | $8.74 \times 10^{-9}$ |
| RNASE4 | 1.21 (1.08-1.36) | $9.37 \times 10^{-4}$ | 1.51 (1.28-1.78) | $1.55 \times 10^{-6}$ |
| ITGA1\|ITGB1 | 1.18 (1.07-1.31) | $1.28 \times 10^{-3}$ | 1.31 (1.15-1.48) | $3.52 \times 10^{-5}$ |
| CD93 | 1.20 (1.07-1.34) | $1.29 \times 10^{-3}$ | 1.42 (1.23-1.64) | $2.53 \times 10^{-6}$ |
| SIGLEC7 | 1.19 (1.07-1.32) | $1.34 \times 10^{-3}$ | 1.44 (1.26-1.64) | $3.24 \times 10^{-8}$ |
| PLA2G2A | 1.20 (1.07-1.34) | $1.40 \times 10^{-3}$ | 1.48 (1.29-1.70) | $4.14 \times 10^{-8}$ |
| C1QL1 | 1.19 (1.07-1.32) | $1.64 \times 10^{-3}$ | 1.30 (1.16-1.46) | $4.05 \times 10^{-6}$ |
| VEGFD | 1.17 (1.06-1.30) | $1.68 \times 10^{-3}$ | 1.39 (1.25-1.55) | $1.10 \times 10^{-9}$ |
| IGHG1\|IGHG2\|IGHG3\|IGHG4\|IGK\|IGL | 1.19 (1.07-1.32) | $1.74 \times 10^{-3}$ | 1.34 (1.16-1.55) | $6.10 \times 10^{-5}$ |
| FGF23 | 1.13 (1.05-1.23) | $2.50 \times 10^{-3}$ | 1.27 (1.14-1.42) | $1.84 \times 10^{-5}$ |
| AXIN2 | 1.18 (1.06-1.31) | $2.69 \times 10^{-3}$ | 1.66 (1.48-1.85) | $1.12 \times 10^{-18}$ |
| CCDC80 | 1.20 (1.07-1.36) | $2.84 \times 10^{-3}$ | 1.59 (1.37-1.85) | $1.40 \times 10^{-9}$ |
| ANGPTL1 | 1.17 (1.06-1.30) | $2.89 \times 10^{-3}$ | 1.38 (1.19-1.61) | $3.28 \times 10^{-5}$ |
| TGFB3 | 1.15 (1.05-1.26) | $3.15 \times 10^{-3}$ | 1.37 (1.24-1.51) | $3.69 \times 10^{-10}$ |
| UNC5C | 1.21 (1.06-1.37) | $3.25 \times 10^{-3}$ | 1.55 (1.31-1.84) | $5.37 \times 10^{-7}$ |
| TEK | 1.16 (1.05-1.28) | $3.39 \times 10^{-3}$ | 0.63 (0.53-0.74) | $5.27 \times 10^{-8}$ |
| CD59 | 1.18 (1.06-1.33) | $3.55 \times 10^{-3}$ | 1.47 (1.30-1.66) | $4.66 \times 10^{-10}$ |
| MATN2 | 1.20 (1.06-1.35) | $3.57 \times 10^{-3}$ | 1.43 (1.21-1.69) | $2.58 \times 10^{-5}$ |
| ZHX3 | 0.87 (0.79-0.96) | $3.98 \times 10^{-3}$ | 0.75 (0.66-0.85) | $4.71 \times 10^{-6}$ |
| ADSS2 | 1.15 (1.04-1.26) | $4.04 \times 10^{-3}$ | 1.26 (1.13-1.40) | $3.15 \times 10^{-5}$ |
| SCARF1 | 1.17 (1.05-1.29) | $4.21 \times 10^{-3}$ | 1.44 (1.26-1.65) | $1.35 \times 10^{-7}$ |
| IGFLR1 | 1.15 (1.04-1.26) | $4.62 \times 10^{-3}$ | 1.36 (1.19-1.55) | $4.64 \times 10^{-6}$ |
| IL15 | 1.16 (1.05-1.28) | $4.79 \times 10^{-3}$ | 1.30 (1.14-1.48) | $5.87 \times 10^{-5}$ |
| UNC5B | 1.18 (1.05-1.33) | $4.80 \times 10^{-3}$ | 1.35 (1.21-1.50) | $2.91 \times 10^{-8}$ |
| TMED10 | 1.16 (1.04-1.28) | $4.91 \times 10^{-3}$ | 1.79 (1.54-2.09) | $1.00 \times 10^{-13}$ |
| SDF2L1 | 1.14 (1.04-1.26) | $5.26 \times 10^{-3}$ | 1.29 (1.17-1.44) | $1.42 \times 10^{-6}$ |

**Supplemental Table 12: Baseline Characteristics by SVEP1 Quartile**

| Characteristic | Quartile 1, N = 630[1] | Quartile 2, N = 630[1] | Quartile 3, N = 629[1] | Quartile 4, N = 629[1] | p-value[2] |
|---|---|---|---|---|---|
| **Age (years)** | 62 (56, 69) | 66 (60, 73) | 69 (62, 75) | 71 (63, 76) | <0.001 |
| **Gender** | | | | | 0.6 |
| Female | 112 (18%) | 114 (18%) | 129 (21%) | 118 (19%) | |
| Male | 518 (82%) | 516 (82%) | 500 (79%) | 511 (81%) | |
| **Race** | | | | | |
| Asian | 5 (0.8%) | 2 (0.3%) | 1 (0.2%) | 1 (0.2%) | |
| Black | 11 (1.7%) | 4 (0.6%) | 4 (0.6%) | 7 (1.1%) | |
| Caucasian | 597 (95%) | 611 (97%) | 604 (96%) | 609 (97%) | |
| Other | 17 (2.7%) | 13 (2.1%) | 19 (3.0%) | 12 (1.9%) | |
| Pacific Islander | 0 (0%) | 0 (0%) | 1 (0.2%) | 0 (0%) | |
| **Region** | | | | | |
| Asia/Pacific and Other | 8 (1.3%) | 5 (0.8%) | 7 (1.1%) | 0 (0%) | |
| Central/Eastern Europe | 277 (44%) | 264 (42%) | 275 (44%) | 274 (44%) | |
| Latin America (including Central America) | 22 (3.5%) | 15 (2.4%) | 7 (1.1%) | 13 (2.1%) | |
| North America | 27 (4.3%) | 24 (3.8%) | 28 (4.5%) | 31 (4.9%) | |
| Western Europe | 296 (47%) | 322 (51%) | 312 (50%) | 311 (49%) | |
| **Diabetes mellitus** | 173 (27%) | 209 (33%) | 244 (39%) | 242 (38%) | <0.001 |
| **Hypertension** | 446 (71%) | 457 (73%) | 466 (74%) | 487 (77%) | 0.052 |
| **Myocardial infarction** | 316 (50%) | 345 (55%) | 286 (45%) | 293 (47%) | 0.004 |
| **Ischemic cardiomyopathy** | 399 (63%) | 429 (68%) | 405 (64%) | 396 (63%) | 0.2 |
| **Stroke** | 47 (7.5%) | 66 (10%) | 57 (9.1%) | 65 (10%) | 0.2 |
| **Atrial fibrillation** | 157 (25%) | 237 (38%) | 324 (52%) | 408 (65%) | <0.001 |
| **NYHA function class** | | | | | <0.001 |
| I / II | 458 (73%) | 437 (69%) | 412 (66%) | 341 (54%) | |
| III / IV | 172 (27%) | 193 (31%) | 217 (34%) | 288 (46%) | |
| **Ejection fraction (%)** | 31.0 (27.0, 34.0) | 30.0 (27.0, 35.0) | 31.7 (28.0, 35.0) | 30.0 (25.0, 34.0) | <0.001 |
| **Anticoagulant usage** | 141 (22%) | 234 (37%) | 315 (50%) | 365 (58%) | <0.001 |
| **ACEi** | 572 (91%) | 565 (90%) | 558 (89%) | 558 (89%) | 0.6 |
| **ARBi** | 66 (10%) | 72 (11%) | 79 (13%) | 84 (13%) | 0.4 |
| **Diuretic** | 504 (80%) | 515 (82%) | 525 (83%) | 545 (87%) | 0.013 |
| **Digoxin** | 112 (18%) | 114 (18%) | 171 (27%) | 197 (31%) | <0.001 |
| **Beta-blocker** | 607 (96%) | 604 (96%) | 594 (94%) | 586 (93%) | 0.041 |
| **Cardiac resynchronization therapy** | 59 (9.4%) | 50 (7.9%) | 48 (7.6%) | 65 (10%) | 0.3 |

| | | | | | |
|---|---|---|---|---|---|
| **Implantable cardioverter defibrillator** | 182 (29%) | 168 (27%) | 134 (21%) | 145 (23%) | 0.008 |
| **BMI (kg/m²)** | 28.6 (25.5, 32.0) | 28.6 (25.7, 32.0) | 28.4 (25.4, 32.1) | 27.6 (24.7, 31.3) | 0.011 |
| Unknown | 2 | 0 | 1 | 0 | |
| **Systolic blood pressure (mmHG)** | 130 (118, 140) | 130 (120, 140) | 130 (120, 141) | 130 (120, 140) | 0.031 |
| **eGFR (60mL/min/1.73m²)** | 72 (59, 84) | 69 (58, 83) | 66 (55, 80) | 64 (52, 78) | <0.001 |
| **NT-proBNP (pg/mL)** | 926 (611, 1,484) | 1,249 (763, 2,075) | 1,542 (892, 2,462) | 2,823 (1,426, 5,165) | <0.001 |
| Unknown | 39 | 29 | 37 | 46 | |

[1] Median (IQR); n (%)
[2] Kruskal-Wallis rank sum test; Pearson's Chi-squared test

**Supplemental Table 13: Pathways Significantly Associated with Risk of Heart Failure Hospitalization or Cardiovascular Death**

| Ingenuity Canonical Pathways | $-\log_{10}$(p-value) | % of Proteins Over-Represented | Over-Represented Proteins Included in Pathway |
|---|---|---|---|
| Hepatic Fibrosis / Hepatic Stellate Cell Activation | 4.23 | 35% | AGT,BAMBI,CCL21,CERT1,COL11A2,COL13A1,COL18A1,COL1A1,COL28A1,COL6A1,COL6A3,CSF1,CXCL8,EDN1,FAS,FGFR1,FGFR2,FLT4,FN1,HGF,IFNAR1,IFNGR1,IGF1R,IL1R1,IL1R2,IL1RL1,IL6,MMP1,MMP2,MYL6B,STAT1,TGFB1,TGFB3,TIMP2,TNFRSF11B,TNFRSF1A,TNFRSF1B,VCAM1,VEGFD |
| Agranulocyte Adhesion and Diapedesis | 3.74 | 35% | C5,CCL14,CCL15,CCL16,CCL18,CCL19,CCL21,CCL23,CCL28,CDH5,CXCL10,CXCL11,CXCL12,CXCL13,CXCL16,CXCL6,CXCL8,FN1,IL18,IL1R1,ITGB7,MMP1,MMP12,MMP19,MMP2,MMP3,MMP7,MMP8,MYL6B,SELE,TNFRSF1A,VCAM1 |
| Granulocyte Adhesion and Diapedesis | 3.72 | 35% | C5,CCL14,CCL15,CCL16,CCL18,CCL19,CCL21,CCL23,CCL28,CDH5,CXCL10,CXCL11,CXCL12,CXCL13,CXCL16,CXCL6,CXCL8,IL18,IL1R1,IL1R2,IL1RL1,MMP1,MMP12,MMP19,MMP2,MMP3,MMP7,MMP8,SELE,TNFRSF11B,TNFRSF1A,TNFRSF1B,VCAM1 |
| Inhibition of Matrix Metalloproteases | 3.02 | 48% | ADAM12,MMP1,MMP12,MMP19,MMP2,MMP3,MMP7,MMP8,TFPI2,THBS2,TIMP2,TIMP4 |
| Acute Phase Response Signaling | 2.70 | 31% | AGT,ALB,APCS,APOA2,C2,C3,C5,C9,CRP,F8,FN1,HAMP,IL18,IL1R1,IL6,IL6ST,ITIH2,ITIH3,JUN,KLKB1,KRAS,MAP2K2,MBL2,PLG,PTPN11,RALA,SERPINA3,SERPINF2,SOD2,TF,TNFRSF11B,TNFRSF1A,TNFRSF1B,VWF |
| Intrinsic Prothrombin Activation Pathway | 2.35 | 41% | COL11A2,COL18A1,COL1A1,F11,F5,F8,KLK10,KLK11,KLK13,KLK14,KLK8,KLKB1 |
| Axonal Guidance Signaling | 2.30 | 26% | ADAM11,ADAM12,ADAM9,ADAMTS3,ADAMTS5,ADAMTS6,BDNF,BMP1,BMP6,BMP7,CFL1,CRK,CXCL12,EFNA4,EFNA5,EFNB2,EPHA10,EPHA2,EPHA7,EPHB2,EPHB6,GSK3B,ITGB7,KRAS,MAP2K2,MMP1,MMP12,MMP2,MMP3,MMP7,MMP8,MYL6B,NOTUM,NRP1,NRP2,NTF3,NTN1,NTN4,PAPPA,PLXNA1,PLXNB2,PLXNC1,PTPN11,RALA,ROBO1,ROBO2,SEMA3E,SEMA4C,SEMA6A,SEMA6B,SEMA7A,UNC5B,UNC5C,VEGFD,WNT5A |
| Apelin Liver Signaling Pathway | 2.10 | 47% | AGT,COL11A2,COL18A1,COL1A1,EDN1,EDN2,FAS,GSK3B |

| | | | |
|---|---|---|---|
| Stearate Biosynthesis I (Animals) | 1.85 | 67% | HNF4A,PPT1,PTGR1,TBXAS1 |
| Atherosclerosis Signaling | 1.79 | 30% | ALB,APOA2,APOF,APOL1,APOM,COL11A2,COL18A1,COL1A1,CSF1,CXCL12,CXCL8,IL18,IL6,LYZ,MMP1,MMP3,PCYOX1,PLA2G12B,PLA2G2A,PLA2R1,SELE,TGFB1,VCAM1 |
| Role of Osteoblasts, Osteoclasts and Chondrocytes in Rheumatoid Arthritis | 1.70 | 27% | ADAMTS5,BGLAP,BMP1,BMP6,BMP7,COL1A1,CSF1,CSF1R,DKK2,DKK3,FRZB,GSK3B,GSN,IL18,IL18R1,IL1R1,IL1R2,IL1RL1,IL6,JUN,MMP1,MMP3,MMP8,PTH,SFRP1,SPP1,TGFB1,TNFRSF11B,TNFRSF1A,TNFRSF1B,WNT5A |
| Glutaryl-CoA Degradation | 1.63 | 75% | HSD17B10,PARK7,TBXAS1 |
| Wound Healing Signaling Pathway | 1.51 | 26% | CERT1,COL11A2,COL13A1,COL18A1,COL1A1,COL28A1,COL6A1,COL6A3,CXCL8,FGF7,FGFR2,FN1,IFNGR1,IL15,IL18,IL1R1,IL1R2,IL1RL1,IL6,JUN,KRAS,LAMC2,MAP2K2,MMP1,MMP8,RALA,STAT1,TGFB1,TGFB3,TNFRSF11B,TNFRSF1A,TNFRSF1B,TNFSF13B,TNFSF15,TNFSF8,TYK2,VEGFD |
| PPAR Signaling | 1.47 | 30% | HSP90AA1,HSP90AB1,HSP90B1,IL18,IL1R1,IL1R2,IL1RL1,INS,JUN,KRAS,MAP2K2,MED1,RALA,TNFRSF11B,TNFRSF1A,TNFRSF1B |
| LXR/RXR Activation | 1.47 | 29% | AGT,ALB,APOA2,APOF,APOL1,APOM,C3,C9,IL18,IL1R1,IL1R2,IL1RL1,IL6,LYZ,PCYOX1,SERPINF2,TF,TNFRSF11B,TNFRSF1A,TNFRSF1B |
| Acyl-CoA Hydrolysis | 1.44 | 100% | HNF4A,PPT1 |
| Sphingomyelin Metabolism | 1.44 | 100% | ENPP7,SMPD1 |
| Role of Macrophages, Fibroblasts and Endothelial Cells in Rheumatoid Arthritis | 1.39 | 25% | C5,CREB3L4,CSF1,CXCL12,CXCL8,DKK2,DKK3,FCGR3A/FCGR3B,FN1,FRZB,GSK3B,IL15,IL18,IL18R1,IL1R1,IL1R2,IL1RL1,IL6,IL6ST,JUN,KRAS,MAP2K2,MIF,MMP1,MMP3,NOTUM,PRSS2,RALA,ROR2,SELE,SFRP1,TGFB1,TNFRSF11B,TNFRSF1A,TNFRSF1B,TNFSF13B,VCAM1,VEGFD,WNT5A |

Legend: Levels of proteins in 18 Ingenuity Canonical Pathways were significantly associated with risk of heart failure hospitalization or cardiovascular death.

**Supplemental Table 14: Proteomic Risk Score Weights**

| Protein | beta | Entrez Gene ID | Organism | SOMAmer Number |
|---|---|---|---|---|
| NPPB | 0.151900519 | 4879 | Human | anti_7655_11 |
| BCAM | 0.099557992 | 4059 | Human | anti_2816_50 |
| CST3 | 0.096290139 | 1471 | Human | anti_2609_59 |
| GDF15 | 0.093131244 | 9518 | Human | anti_4374_45 |
| Fgfr2 | -0.076260252 | 14183 | Mouse | anti_5497_29 |
| TNNI3 | 0.073567616 | 7137 | Human | anti_5441_67 |
| C1orf185 | 0.067910998 | 284546 | Human | anti_10667_78 |
| ESRRA | 0.057829637 | 2101 | Human | anti_9873_17 |
| INSL5 | 0.054448048 | 10022 | Human | anti_10462_14 |
| CLCA1 | 0.051919038 | 1179 | Human | anti_10496_11 |
| FSTL5 | 0.047394114 | 56884 | Human | anti_7099_33 |
| MMP9 | 0.046719495 | 4318 | Human | anti_2579_17 |
| QSOX1 | 0.045569277 | 5768 | Human | anti_6217_23 |
| IGLL1 | -0.045537151 | 3543 | Human | anti_6485_59 |
| ANGPT2 | 0.043148638 | 285 | Human | anti_2602_2 |
| MET | -0.042997844 | 4233 | Human | anti_11814_29 |
| SVEP1 | 0.042725615 | 79987 | Human | anti_11109_56 |
| CILP2 | -0.036637629 | 148113 | Human | anti_8841_65 |
| KIT | -0.034634831 | 3815 | Human | anti_2475_1 |
| NECTIN1 | -0.034293141 | 5818 | Human | anti_9300_13 |
| rev | 0.032671704 | 1724716 | HIV-2 | anti_2769_3 |
| TREM1 | 0.031906206 | 54210 | Human | anti_9266_1 |
| SLITRK3 | -0.031719216 | 22865 | Human | anti_10565_19 |
| REN | 0.025955348 | 5972 | Human | anti_3396_54 |
| ATG4B | 0.024020235 | 23192 | Human | anti_13629_25 |
| GKN2 | 0.023870432 | 200504 | Human | anti_6416_8 |
| BOC | 0.022225270 | 91653 | Human | anti_4328_2 |
| CNDP1 | -0.021457755 | 84735 | Human | anti_7870_8 |
| STC1 | 0.021289948 | 6781 | Human | anti_4930_21 |
| ARFIP2 | -0.021032689 | 23647 | Human | anti_12630_8 |

| | | | | |
|---|---|---|---|---|
| RAP1GDS1 | -0.020439342 | 5910 | Human | anti_14106_46 |
| TNNI3 | 0.019594699 | 7137 | Human | anti_5930_54 |
| COL11A2 | -0.017355220 | 1302 | Human | anti_11278_4 |
| ZNRF3 | 0.016569734 | 84133 | Human | anti_13428_52 |
| VWA2 | -0.016260130 | 340706 | Human | anti_7128_9 |
| ADSS2 | 0.015421496 | 159 | Human | anti_12644_63 |
| CSF1R | 0.015268194 | 1436 | Human | anti_2638_12 |
| HPGDS | -0.014279470 | 27306 | Human | anti_12549_33 |
| BET1L | -0.013804708 | 51272 | Human | anti_10959_125 |
| PILRA | 0.012673244 | 29992 | Human | anti_10816_150 |
| CXCL13 | 0.012600662 | 10563 | Human | anti_13701_2 |
| ADAMTS6 | 0.012599351 | 11174 | Human | anti_6441_62 |
| PFDN5 | 0.012567242 | 5204 | Human | anti_4271_75 |
| GRP | -0.011316279 | 2922 | Human | anti_8400_74 |
| MFAP1 | 0.011093630 | 4236 | Human | anti_5606_24 |
| IFNL3 | -0.010413780 | 282617 | Human | anti_5713_9 |
| ZNF134 | -0.010264565 | 7693 | Human | anti_12787_47 |
| NLGN4X | -0.009895443 | 57502 | Human | anti_5357_60 |
| IFNGR2 | -0.009639248 | 3460 | Human | anti_9305_89 |
| MMP14 | -0.009295171 | 4323 | Human | anti_5002_76 |
| Igh | 0.009010786 | | Mouse | anti_12906_137 |
| ZNF41 | 0.007835859 | 7592 | Human | anti_10003_15 |
| ULK3 | -0.007560956 | 25989 | Human | anti_12437_18 |
| NTS | 0.007250630 | 4922 | Human | anti_7857_22 |
| SPN | -0.006919377 | 6693 | Human | anti_12873_11 |
| FRS2 | -0.006625050 | 10818 | Human | anti_13025_4 |
| PTPRU | 0.006039151 | 10076 | Human | anti_8337_65 |
| ADAMTSL2 | 0.005597400 | 9719 | Human | anti_6379_62 |
| CDK2AP1 | -0.005184494 | 8099 | Human | anti_9450_18 |
| TOR1AIP1 | -0.004835733 | 26092 | Human | anti_9039_47 |
| HAMP | -0.003780840 | 57817 | Human | anti_3504_58 |
| TNFRSF10B | -0.002504706 | 8795 | Human | anti_10878_1 |
| PARK7 | 0.002448034 | 11315 | Human | anti_5016_61 |

| | | | | |
|---|---|---|---|---|
| fbpC | -0.002346403 | 886885 | Mycobacterium tuberculosis | anti_5575_1 |

**Supplemental Table 15: Risk Model C-Statistics at Truncated Follow-up**

Derivation Set: ATMOSPHERE

| model | One year | Two years | Four years | Overall |
|---|---|---|---|---|
| **MAGGIC Score** | 0.582<br>(0.527 - 0.638) | 0.592<br>(0.553 - 0.631) | 0.601<br>(0.571 - 0.630) | 0.598<br>(0.570 - 0.627) |
| **NT-proBNP** | 0.605<br>(0.549 - 0.661) | 0.619<br>(0.580 - 0.658) | 0.613<br>(0.583 - 0.643) | 0.611<br>(0.582 - 0.640) |
| **Proteomic Score** | 0.728<br>(0.680 - 0.775) | 0.721<br>(0.687 - 0.754) | 0.729<br>(0.704 - 0.755) | 0.728<br>(0.704 - 0.753) |
| **MAGGIC Score + NTPBNP** | 0.615<br>(0.558 - 0.671) | 0.636<br>(0.597 - 0.676) | 0.636<br>(0.606 - 0.666) | 0.633<br>(0.604 - 0.662) |
| **MAGGIC + Proteomics** | 0.727<br>(0.679 - 0.774) | 0.720<br>(0.686 - 0.754) | 0.729<br>(0.704 - 0.755) | 0.728<br>(0.703 - 0.752) |

Validation Set: PARADIGM-HF

| model | One year | Two years | Overall |
|---|---|---|---|
| **MAGGIC Score** | 0.627<br>(0.580 - 0.673) | 0.606<br>(0.568 - 0.643) | 0.605<br>(0.571 - 0.640) |
| **NT-proBNP** | 0.686<br>(0.642 - 0.730) | 0.665<br>(0.629 - 0.701) | 0.653<br>(0.620 - 0.686) |
| **Proteomic Score** | 0.734<br>(0.694 - 0.774) | 0.713<br>(0.680 - 0.746) | 0.703<br>(0.673 - 0.734) |
| **MAGGIC Score + NTPBNP** | 0.692<br>(0.647 - 0.737) | 0.668<br>(0.631 - 0.704) | 0.659<br>(0.626 - 0.693) |
| **MAGGIC + Proteomic Score** | 0.735<br>(0.695 - 0.775) | 0.714<br>(0.681 - 0.747) | 0.704<br>(0.673 - 0.735) |

C-statistics with 95% confidence intervals are shown. One-year and two-year time periods refer to truncated follow-up with the identical risk score.

**Supplemental Table 16: Causal Associations Between Protein Levels and Heart Failure Hospitalization or Cardiovascular Death by Mendelian Randomization**

| Protein | number of IVs | method | β | Nominal P-value | Horizontal Pleiotropy Test (MR_PRESSO Pvalue) |
|---|---|---|---|---|---|
| NPPB[1] | 4 | IVW | −1.46 | $8.50 \times 10^{-5}$ | 0.85 |
| WISP2[1] | 34 | IVW | 0.37 | $1.05 \times 10^{-4}$ | 0.21 |
| CTSS[1] | 18 | IVW | −0.32 | $1.12 \times 10^{-4}$ | 0.56 |
| FSTL1[1] | 6 | IVW | 0.88 | $3.48 \times 10^{-4}$ | 0.52 |
| ADAMTS5 | 11 | IVW | 0.34 | 0.0011 | 0.66 |
| OAF | 30 | IVW | 0.20 | 0.0020 | 0.63 |
| HAVCR1 | 22 | IVW | −0.28 | 0.0032 | 0.54 |
| B3GNT2 | 17 | IVW | 0.52 | 0.0040 | 0.14 |
| PIANP | 5 | IVW | 0.95 | 0.0062 | 0.57 |
| CNDP1 | 21 | IVW | −0.25 | 0.0070 | 0.37 |
| ATP1B2 | 16 | IVW | 0.24 | 0.0076 | 0.89 |
| ENPP7 | 37 | IVW | 0.13 | 0.0080 | 0.47 |
| P4HB | 3 | IVW | 0.87 | 0.0082 | NA |
| PLXNB2 | 23 | IVW | 0.23 | 0.0090 | 0.22 |
| FLRT2 | 23 | IVW | 0.27 | 0.0091 | 0.47 |
| COL18A1 | 11 | IVW | −0.51 | 0.0098 | 0.48 |
| PRSS57 | 18 | IVW | −0.20 | 0.012 | 0.90 |
| BCHE | 17 | IVW | −0.19 | 0.013 | 0.77 |
| WISP1 | 34 | IVW | −0.16 | 0.014 | 0.13 |
| INSL5 | 1 | Wald ratio | 2.06 | 0.014 | NA |
| FMOD | 5 | IVW | −0.72 | 0.016 | 0.76 |
| COL6A3 | 3 | IVW | 1.16 | 0.022 | NA |
| HSPA1A | 12 | IVW | −0.51 | 0.022 | 0.54 |
| HLA-DQA2 | 41 | IVW | −0.15 | 0.024 | 0.80 |
| IGFLR1 | 18 | IVW | −0.18 | 0.028 | 0.40 |
| MMP7 | 17 | IVW | −0.29 | 0.029 | 0.72 |
| NPPA | 1 | Wald ratio | −1.98 | 0.030 | NA |
| PCYOX1 | 29 | IVW | −0.16 | 0.032 | 0.73 |
| F11 | 12 | IVW | 0.20 | 0.033 | 0.73 |
| SCARF2 | 4 | IVW | 0.56 | 0.036 | 0.90 |
| TNFRSF21 | 5 | IVW | −0.62 | 0.038 | 0.88 |
| VASN | 1 | Wald ratio | −1.43 | 0.039 | NA |
| CSF1R | 4 | IVW | −0.60 | 0.040 | 0.64 |
| SFRP1 | 12 | IVW | 0.31 | 0.042 | 0.98 |

| | | | | | |
|---|---|---|---|---|---|
| KLK14 | 13 | IVW | 0.29 | 0.042 | 0.95 |
| DNAJB12 | 1 | Wald ratio | 1.17 | 0.042 | NA |
| IGFBP7 | 20 | IVW | 0.25 | 0.045 | 0.18 |
| ADM2 | 2 | IVW | 0.80 | 0.046 | NA |
| ASPN | 21 | IVW | –0.18 | 0.048 | 0.34 |
| AGT | 5 | IVW | 0.39 | 0.050 | 0.94 |

**Supplemental Figure 1: SVEP1 cis-pQTL Locuszoom plot in deCODE**

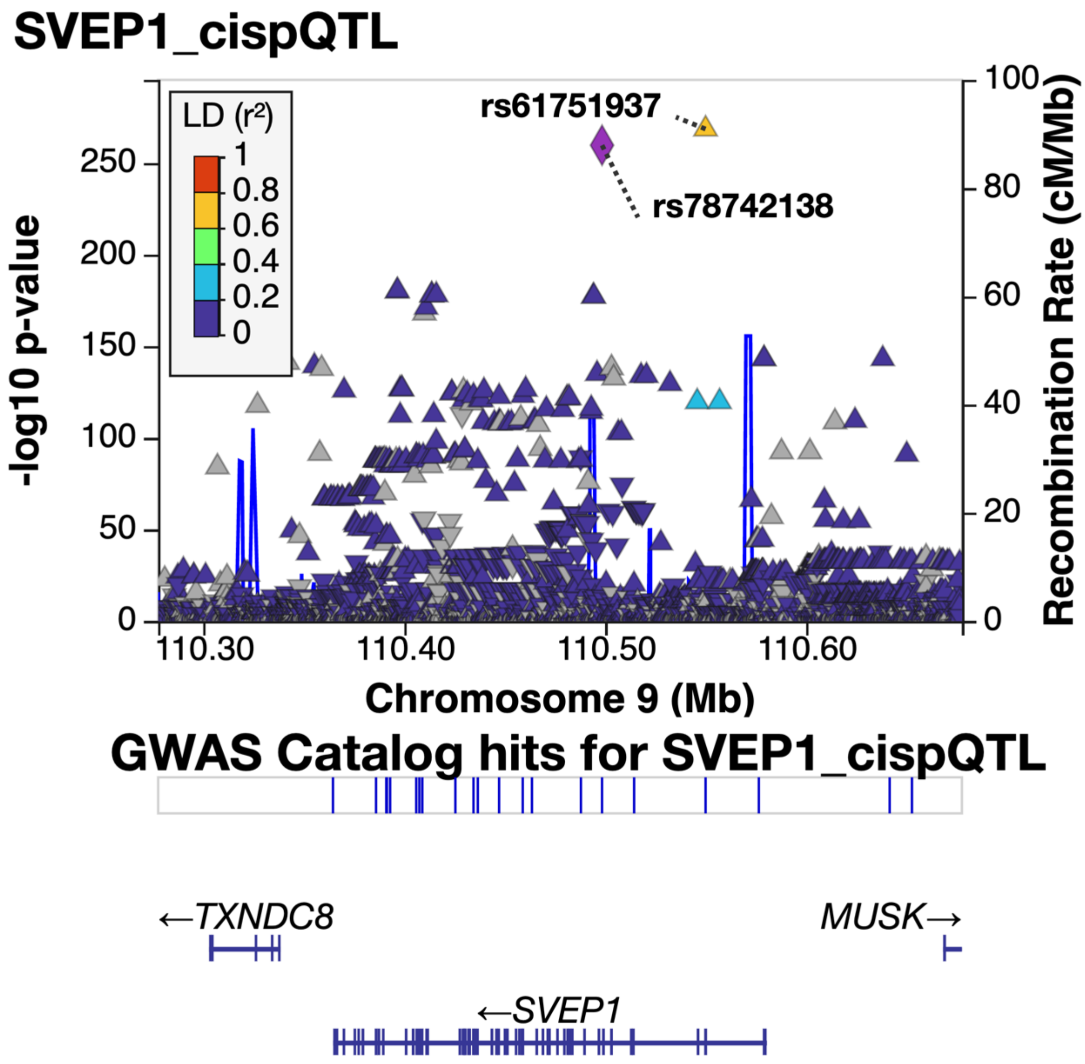


This locus zoom plot shows the strong association between genetic variants near the SVEP1 gene and plasma levels of SVEP1 protein measured by modified aptamer proteomics assay in the deCODE cohort. Observation of this strong cis-pQTL supports the specificity of the modified aptamer for SVEP1.

### Supplemental Figure 2: Correlation between Proteins Included in Proteomic Risk Score

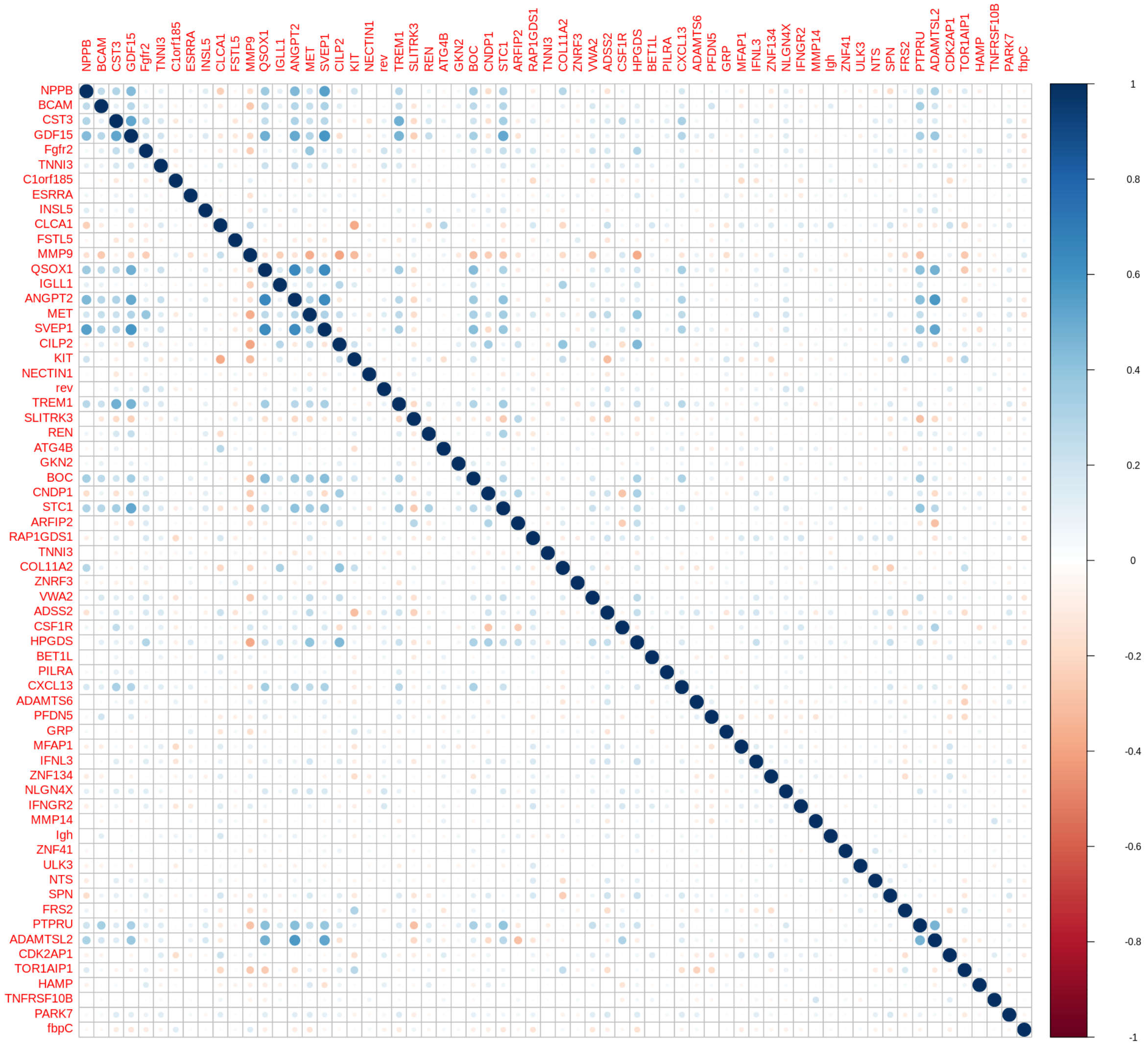


Correlation coefficients between the 64 proteins included in the proteomic risk score were calculated in the ATMOSPHERE derivation cohort.

**Supplementary Figure 3: Correlation between NT-proBNP Levels Measured by ELISA and Aptamer-Based Assays**

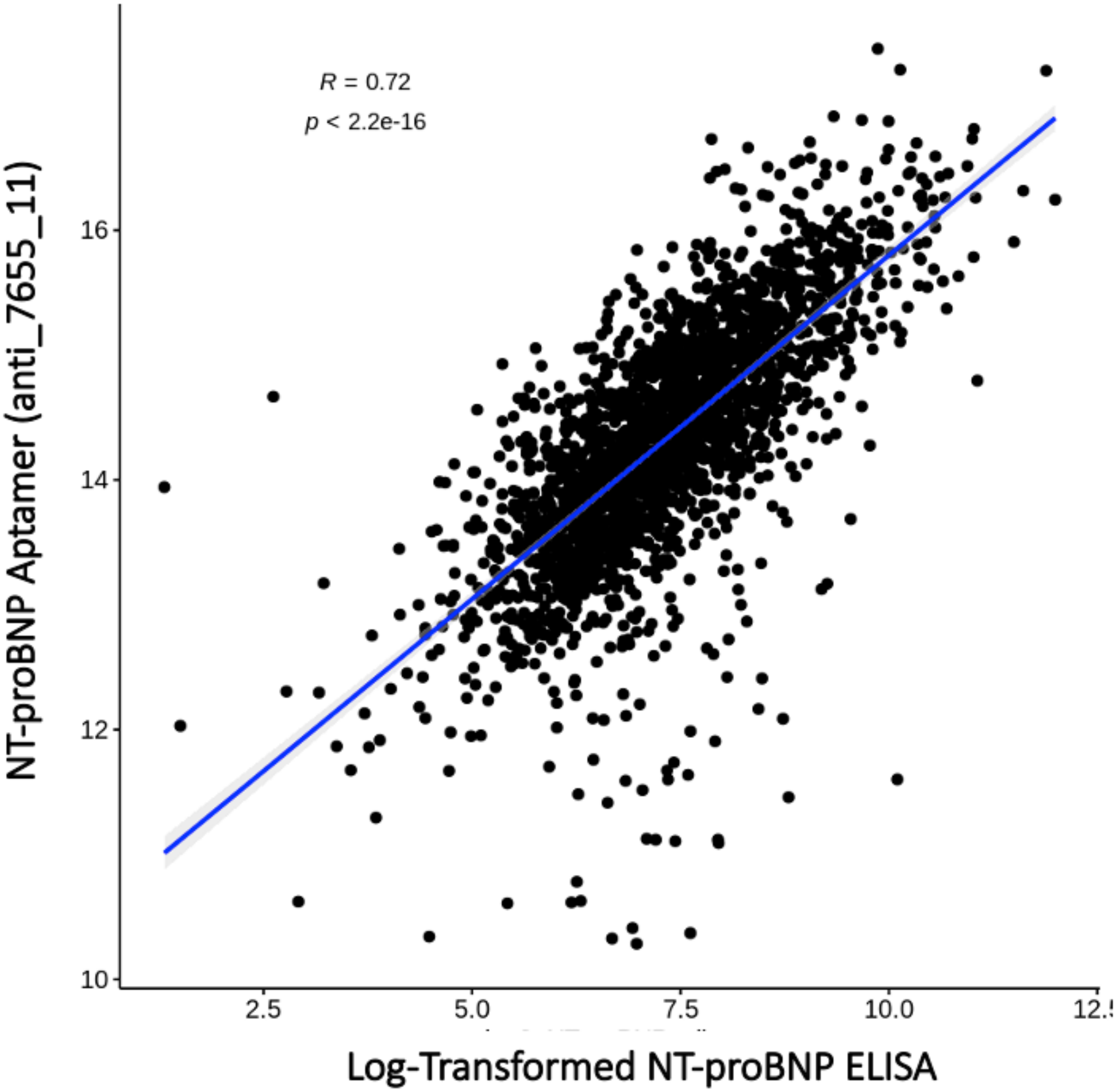

**Supplementary Figure 4: Confidence Intervals for C-Statistic Differences Determined by Bootstrapping**

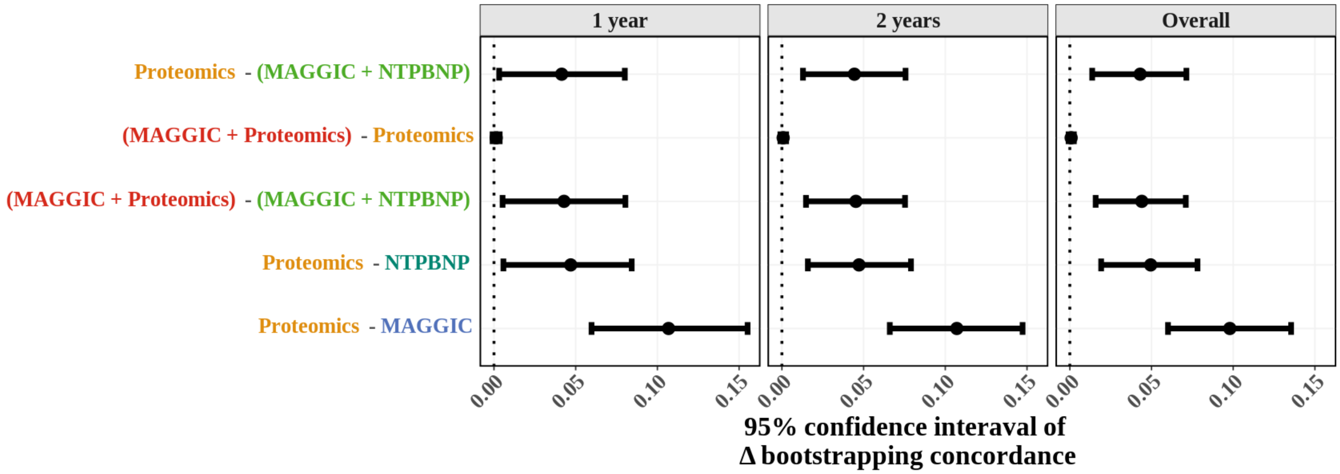


95% confidence intervals for the difference in C-statistic between models were calculated by bootstrapping rather than the Somer's D method as a sensitivity analysis. Results were consistent with both statistical approaches.
