## Supplemental Material SOMAmer protein mapping for "Aptamer Proteomics for Biomarker Discovery in Heart Failure with Reduced Ejection Fraction"

| SOMAmer | Target Full Name | Entrez Gene Symbol | Organism |
|---|---|---|---|
| anti_10041_3 | Hepatocyte nuclear factor 4-alpha | HNF4A | Human |
| anti_10391_1 | Angiopoietin-related protein 3 | ANGPTL3 | Human |
| anti_10445_20 | Apolipoprotein M | APOM | Human |
| anti_10492_51 | Glycoprotein Xg | XG | Human |
| anti_10514_5 | Prostaglandin-H2 D-isomerase | PTGDS | Human |
| anti_10666_7 | N-acetylglucosamine-1-phosphotransferase subunit gamma | GNPTG | Human |
| anti_10819_108 | Fibulin-1 | FBLN1 | Human |
| anti_10902_53 | Retroviral-like aspartic protease 1 | ASPRV1 | Human |
| anti_11089_7 | Immunoglobulin A | IGHA1\|IGHA2 | Human |
| anti_11237_49 | Procollagen C-endopeptidase enhancer 1 | PCOLCE | Human |
| anti_11510_31 | Apolipoprotein L1 | APOL1 | Human |
| anti_11510_51 | Apolipoprotein L1 | APOL1 | Human |
| anti_13095_51 | Lithostathine-1-alpha | REG1A | Human |
| anti_13114_50 | Lumican | LUM | Human |
| anti_13671_40 | Neutrophil elastase | ELANE | Human |
| anti_13678_169 | Complement factor D | CFD | Human |
| anti_13682_47 | Macrophage colony-stimulating factor 1 receptor | CSF1R | Human |
| anti_13710_6 | Plasma protease C1 inhibitor | SERPING1 | Human |
| anti_13722_105 | Complement component C9 | C9 | Human |
| anti_13731_14 | Complement component C7 | C7 | Human |
| anti_14048_7 | Interleukin-1 Receptor accessory protein | IL1RAP | Human |
| anti_14073_31 | Hemopexin | HPX | Human |
| anti_14088_38 | Insulin-like growth factor-binding protein 6 | IGFBP6 | Human |
| anti_14105_5 | Kallistatin | SERPINA4 | Human |
| anti_14237_1 | Heat shock 70 kDa protein 1A | HSPA1A | Human |
| anti_14708_59 | Complement component C8 gamma chain | C8G | Human |
| anti_2182_54 | Complement C4b | C4A\|C4B | Human |
| anti_2190_55 | Coagulation Factor XI | F11 | Human |
| anti_2211_9 | Metalloproteinase inhibitor 1 | TIMP1 | Human |
| anti_2278_61 | Metalloproteinase inhibitor 2 | TIMP2 | Human |
| anti_2292_17 | Complement component C9 | C9 | Human |
| anti_2381_52 | Complement C5 | C5 | Human |
| anti_2418_55 | Apolipoprotein E | APOE | Human |
| anti_2474_54 | Serum amyloid P-component | APCS | Human |
| anti_2523_31 | C-C motif chemokine 5 | CCL5 | Human |
| anti_2567_5 | Complement factor I | CFI | Human |
| anti_2579_17 | Matrix metalloproteinase-9 | MMP9 | Human |
| anti_2609_59 | Cystatin-C | CST3 | Human |
| anti_2638_12 | Macrophage colony-stimulating factor 1 receptor | CSF1R | Human |
| anti_2683_1 | Complement C3b, inactivated | C3 | Human |
| anti_2685_21 | Insulin-like growth factor-binding protein 5 | IGFBP5 | Human |
| anti_2697_7 | Platelet factor 4 | PF4 | Human |
| anti_2700_56 | Vitamin K-dependent protein S | PROS1 | Human |
| anti_2706_69 | Thyroxine-binding globulin | SERPINA7 | Human |
| anti_2744_57 | Immunoglobulin G | IGHG1\|IGHG2\|IGHG3\|IGHG4\|IGK\|IGL | Human |
| anti_2750_3 | Apolipoprotein A-I | APOA1 | Human |
| anti_2753_2 | Complement C1q subcomponent | C1QA\|C1QB\|C1QC | Human |
| anti_2754_50 | Complement C3 | C3 | Human |
| anti_2755_8 | C3a anaphylatoxin des Arginine | C3 | Human |
| anti_2768_56 | Hemopexin | HPX | Human |
| anti_2780_35 | Lactotransferrin | LTF | Human |
| anti_2790_54 | Neutrophil-activating peptide 2 | PPBP | Human |
| anti_2795_23 | Lactotransferrin | LTF | Human |
| anti_2836_68 | Neutrophil gelatinase-associated lipocalin | LCN2 | Human |
| anti_2879_9 | Alpha-1-antichymotrypsin | SERPINA3 | Human |
| anti_2888_49 | Complement component C7 | C7 | Human |
| anti_2914_65 | Neutrophil-activating peptide 2 | PPBP | Human |
| anti_2937_10 | Apolipoprotein E (isoform E3) | APOE | Human |
| anti_2938_55 | Apolipoprotein E (isoform E4) | APOE | Human |
| anti_2943_5 | Cytochrome P450 3A4 | CYP3A4 | Human |
| anti_2950_57 | Insulin-like growth factor-binding protein 4 | IGFBP4 | Human |
| anti_2952_75 | Insulin-like growth factor I | IGF1 | Human |
| anti_2960_66 | Properdin | CFP | Human |

| SOMAmer | Target Full Name | Entrez Gene Symbol | Organism |
|---|---|---|---|
| anti_2961_1 | Vitamin K-dependent protein C | PROC | Human |
| anti_3024_18 | Alpha-2-antiplasmin | SERPINF2 | Human |
| anti_3031_66 | Ferritin | FTH1\|FTL | Human |
| anti_3043_49 | SPARC | SPARC | Human |
| anti_3044_3 | C-C motif chemokine 18 | CCL18 | Human |
| anti_3050_7 | von Willebrand factor | VWF | Human |
| anti_3054_3 | Haptoglobin | HP | Human |
| anti_3057_55 | WNT1-inducible-signaling pathway protein 1 | CCN4 | Human |
| anti_3060_43 | Complement component C9 | C9 | Human |
| anti_3069_52 | Immunoglobulin M | IGHM\|JCHAIN\|IGK\|IGL | Human |
| anti_3074_6 | Lipopolysaccharide-binding protein | LBP | Human |
| anti_3077_66 | Coagulation factor Xa | F10 | Human |
| anti_3079_62 | Retinoic acid receptor responder protein 2 | RARRES2 | Human |
| anti_3171_57 | Amyloid beta A4 protein | APP | Human |
| anti_3184_25 | Coagulation factor VII | F7 | Human |
| anti_3186_2 | Complement C2 | C2 | Human |
| anti_3206_4 | Lymphatic vessel endothelial hyaluronic acid receptor 1 | LYVE1 | Human |
| anti_3293_2 | CD5 antigen-like | CD5L | Human |
| anti_3311_27 | Low affinity immunoglobulin gamma Fc region receptor III-B | FCGR3B | Human |
| anti_3316_58 | Heparin cofactor 2 | SERPIND1 | Human |
| anti_3344_60 | Antithrombin-III | SERPINC1 | Human |
| anti_3366_51 | Extracellular matrix protein 1 | ECM1 | Human |
| anti_3367_8 | Fetuin-B | FETUB | Human |
| anti_3389_7 | Plasma serine protease inhibitor | SERPINA5 | Human |
| anti_3434_34 | Fibronectin Fragment 3 | FN1 | Human |
| anti_3435_53 | Fibronectin Fragment 4 | FN1 | Human |
| anti_3449_58 | Kallistatin | SERPINA4 | Human |
| anti_3474_19 | Thrombospondin-1 | THBS1 | Human |
| anti_3484_60 | Angiotensinogen | AGT | Human |
| anti_3485_28 | Beta-2-microglobulin | B2M | Human |
| anti_3518_54 | Carboxypeptidase B2 | CPB2 | Human |
| anti_3554_24 | Adiponectin | ADIPOQ | Human |
| anti_3580_25 | Alpha-1-antitrypsin | SERPINA1 | Human |
| anti_3581_53 | Alpha-2-HS-glycoprotein | AHSG | Human |
| anti_3590_8 | Complement factor H | CFH | Human |
| anti_3617_80 | Hepatocyte growth factor activator | HGFAC | Human |
| anti_3700_15 | Immunoglobulin G | IGHG1\|IGHG2\|IGHG3\|IGHG4\|IGK\|IGL | Human |
| anti_3707_12 | Serum albumin | ALB | Human |
| anti_3890_8 | L-lactate dehydrogenase B chain | LDHB | Human |
| anti_4127_75 | Complement component C6 | C6 | Human |
| anti_4129_72 | Complement factor B | CFB | Human |
| anti_4131_72 | Fibronectin | FN1 | Human |
| anti_4151_6 | Plasminogen | PLG | Human |
| anti_4152_58 | Plasma kallikrein | KLKB1 | Human |
| anti_4153_11 | Alpha-1-antichymotrypsin complex | SERPINA3 | Human |
| anti_4159_130 | Complement factor H | CFH | Human |
| anti_4162_54 | Serotransferrin | TF | Human |
| anti_4407_10 | Hepatocyte growth factor-like protein | MST1 | Human |
| anti_4481_34 | Complement C4 | C4A\|C4B | Human |
| anti_4482_66 | Complement C5b-C6 complex | C5\|C6 | Human |
| anti_4495_33 | Kininogen-1 | KNG1 | Human |
| anti_4544_4 | Connective tissue-activating peptide III | PPBP | Human |
| anti_4763_31 | Afamin | AFM | Human |
| anti_4775_34 | Gelsolin | GSN | Human |
| anti_4811_33 | Inter-alpha-trypsin inhibitor heavy chain H4 | ITIH4 | Human |
| anti_4831_4 | L-Selectin | SELL | Human |
| anti_4874_3 | Angiogenin | ANG | Human |
| anti_4876_32 | Coagulation factor IX | F9 | Human |
| anti_4878_3 | Coagulation Factor X | F10 | Human |
| anti_4900_8 | C3a anaphylatoxin | C3 | Human |
| anti_4906_35 | Coagulation Factor V | F5 | Human |
| anti_4918_21 | Kininogen-1 | KNG1 | Human |

| SOMAmer | Target Full Name | Entrez Gene Symbol | Organism |
|---|---|---|---|
| anti_4920_10 | Lysozyme C | LYZ | Human |
| anti_4929_55 | Sex hormone-binding globulin | SHBG | Human |
| anti_4983_6 | Endoplasmic reticulum resident protein 29 | ERP29 | Human |
| anti_4996_66 | Histidine-rich glycoprotein | HRG | Human |
| anti_4998_50 | Tyrosine-protein kinase JAK2 | JAK2 | Human |
| anti_5059_8 | Immunoglobulin G | IGHG1\|IGHG2\|IGHG3\|IGHG4\|IGK\|IGL | Human |
| anti_5071_3 | Immunoglobulin G | IGHG1\|IGHG2\|IGHG3\|IGHG4\|IGK\|IGL | Human |
| anti_5073_30 | Immunoglobulin G | IGHG1\|IGHG2\|IGHG3\|IGHG4\|IGK\|IGL | Human |
| anti_5108_72 | Neurogenic locus notch homolog protein 3 | NOTCH3 | Human |
| anti_5118_74 | Immunoglobulin G | IGHG1\|IGHG2\|IGHG3\|IGHG4\|IGK\|IGL | Human |
| anti_5231_79 | Proprotein convertase subtilisin/kexin type 9 | PCSK9 | Human |
| anti_5307_12 | Coagulation factor IXab | F9 | Human |
| anti_5312_49 | Apolipoprotein E (isoform E2) | APOE | Human |
| anti_5316_54 | Prothrombin | F2 | Human |
| anti_5412_53 | CD27 antigen | CD27 | Human |
| anti_5456_59 | Beta-Ala-His dipeptidase | CNDP1 | Human |
| anti_5462_62 | Ficolin-3 | FCN3 | Human |
| anti_5482_61 | Retinol-binding protein 4 | RBP4 | Human |
| anti_5508_62 | Cathepsin D | CTSD | Human |
| anti_5532_53 | Fibroblast growth factor receptor 1 | FGFR1 | Human |
| anti_5542_22 | Neuropilin-1 | NRP1 | Human |
| anti_5586_66 | Multiple inositol polyphosphate phosphatase 1 | MINPP1 | Human |
| anti_5601_2 | N-acetylmuramoyl-L-alanine amidase | PGLYRP2 | Human |
| anti_5632_6 | Cartilage acidic protein 1 | CRTAC1 | Human |
| anti_5658_64 | Coagulation factor XIII B chain | F13B | Human |
| anti_5682_13 | Vasorin | VASN | Human |
| anti_5701_81 | Tetranectin | CLEC3B | Human |
| anti_5803_24 | Complement C3d fragment | C3 | Human |
| anti_5982_50 | Complement factor H-related protein 1 | CFHR1 | Human |
| anti_6075_61 | Beta-hexosaminidase subunit beta | HEXB | Human |
| anti_6217_23 | Sulfhydryl oxidase 1 | QSOX1 | Human |
| anti_6225_3 | Prostasin | PRSS8 | Human |
| anti_6259_60 | Tenascin | TNC | Human |
| anti_6277_55 | Fc_MOUSE | Igh | Mouse |
| anti_6289_78 | Rho GTPase-activating protein 36 | ARHGAP36 | Human |
| anti_6334_9 | Inactive gamma-glutamyltranspeptidase 2 | GGT2 | Human |
| anti_6340_10 | Fc_MOUSE | Igh | Mouse |
| anti_6379_62 | ADAMTS-like protein 2 | ADAMTSL2 | Human |
| anti_6380_23 | Peptidyl-tRNA hydrolase ICT1, mitochondrial | MRPL58 | Human |
| anti_6390_18 | Neuropeptide S | NPS | Human |
| anti_6391_52 | Leukocyte immunoglobulin-like receptor subfamily A member 3 | LILRA3 | Human |
| anti_6415_90 | Carboxypeptidase N subunit 2 | CPN2 | Human |
| anti_6431_68 | Prenylcysteine oxidase 1 | PCYOX1 | Human |
| anti_6439_59 | Uncharacterized protein C14orf93 | C14orf93 | Human |
| anti_6442_6 | B melanoma antigen 3 | BAGE3 | Human |
| anti_6580_29 | Pregnancy zone protein | PZP | Human |
| anti_6583_67 | Protein Z-dependent protease inhibitor | SERPINA10 | Human |
| anti_6605_17 | Insulin-like growth factor-binding protein complex acid labile subunit | IGFALS | Human |
| anti_6606_61 | Metastasis-suppressor KiSS-1 | KISS1 | Human |
| anti_7163_26 | Extracellular glycoprotein lacritin | LACRT | Human |
| anti_7185_29 | Platelet glycoprotein V | GP5 | Human |
| anti_7638_30 | Vesicular integral-membrane protein VIP36 | LMAN2 | Human |
| anti_7640_29 | Low-density lipoprotein receptor-related protein 1B | LRP1B | Human |
| anti_7735_17 | Pigment epithelium-derived factor | SERPINF1 | Human |
| anti_7784_1 | Kininogen-1 | KNG1 | Human |
| anti_7831_39 | Retinol-binding protein 4 | RBP4 | Human |
| anti_7885_17 | Complement factor H-related protein 5 | CFHR5 | Human |
| anti_7905_30 | Haptoglobin isoform 2 | HP | Human |

| SOMAmer | Target Full Name | Entrez Gene Symbol | Organism |
|---|---|---|---|
| anti_7909_37 | Sex hormone-binding globulin | SHBG | Human |
| anti_7918_114 | Alpha-amylase 1 | AMY1A | Human |
| anti_7955_195 | Inter-alpha-trypsin inhibitor heavy chain H1 | ITIH1 | Human |
| anti_8043_153 | Cartilage oligomeric matrix protein | COMP | Human |
| anti_8239_223 | Cholinesterase | BCHE | Human |
| anti_8262_20 | Apolipoprotein D | APOD | Human |
| anti_8280_238 | Vitronectin | VTN | Human |
| anti_8288_27 | Beta-2-glycoprotein 1 | APOH | Human |
| anti_8289_8 | Transmembrane glycoprotein NMB | GPNMB | Human |
| anti_8310_6 | Zymogen granule protein 16 homolog B | ZG16B | Human |
| anti_8463_2 | Extracellular superoxide dismutase [Cu-Zn] | SOD3 | Human |
| anti_8469_41 | Insulin-like growth factor-binding protein 2 | IGFBP2 | Human |
| anti_8480_29 | EGF-containing fibulin-like extracellular matrix protein 1 | EFEMP1 | Human |
| anti_8487_62 | Immunoglobulin A | IGHA1\|IGHA2 | Human |
| anti_8840_61 | Complement C1s subcomponent | C1S | Human |
| anti_8905_20 | Calcium-activated potassium channel subunit beta-3 | KCNMB3 | Human |
| anti_8958_51 | Neural cell adhesion molecule L1-like protein | CHL1 | Human |
| anti_8969_49 | Monocyte differentiation antigen CD14 | CD14 | Human |
| anti_9002_36 | Serpin A11 | SERPINA11 | Human |
| anti_9211_19 | Pigment epithelium-derived factor | SERPINF1 | Human |
| anti_9269_7 | Biotinidase | BTD | Human |
| anti_9312_8 | Zinc-alpha-2-glycoprotein | AZGP1 | Human |
| anti_9326_33 | Inter-alpha-trypsin inhibitor heavy chain H2 | ITIH2 | Human |
| anti_9348_1 | Complement C1r subcomponent-like protein | C1RL | Human |
| anti_9449_150 | C4b-binding protein alpha chain | C4BPA | Human |
| anti_9969_8 | Solute carrier family 22 member 16 | SLC22A16 | Human |
| anti_10011_65 | Inositol polyphosphate 5-phosphatase OCRL-1 | OCRL | Human |
| anti_10025_1 | Dihydrolipoyl dehydrogenase, mitochondrial | DLD | Human |
| anti_10034_16 | Fc_MOUSE | Igh | Mouse |
| anti_10036_201 | Zinc fingers and homeoboxes protein 3 | ZHX3 | Human |
| anti_10039_32 | Purine nucleoside phosphorylase | PNP | Human |
| anti_10046_55 | Baculoviral IAP repeat-containing protein 2 | BIRC2 | Human |
| anti_10047_12 | Neutrophil cytosol factor 2 | NCF2 | Human |
| anti_10048_7 | Core-binding factor subunit beta | CBFB | Human |
| anti_10058_1 | UV excision repair protein RAD23 homolog A | RAD23A | Human |
| anti_10064_12 | Putative hydrolase RBBP9 | RBBP9 | Human |
| anti_10082_251 | Neurofilament light polypeptide | NEFL | Human |
| anti_10085_25 | Steroidogenic acute regulatory protein, mitochondrial | STAR | Human |
| anti_10086_39 | Cystathionine beta-synthase | CBS | Human |
| anti_10339_48 | Gamma-enolase | ENO2 | Human |
| anti_10344_334 | Interleukin-10 receptor subunit alpha | IL10RA | Human |
| anti_10346_5 | Signal transducer and activator of transcription 3 | STAT3 | Human |
| anti_10351_51 | Interferon regulatory factor 1 | IRF1 | Human |
| anti_10354_57 | Signal transducer and activator of transcription 3 | STAT3 | Human |
| anti_10363_13 | Mothers against decapentaplegic homolog 3 | SMAD3 | Human |
| anti_10372_18 | Signal transducer and activator of transcription 6 | STAT6 | Human |
| anti_10378_6 | Angiopoietin-2_MOUSE | Angpt2 | Mouse |
| anti_10379_19 | A disintegrin and metalloproteinase with thrombospondin motifs 1_MOUSE | Adamts1 | Mouse |
| anti_10382_1 | Angiopoietin-related protein 3 | ANGPTL3 | Human |
| anti_10387_1 | Natriuretic peptides B | NPPB | Human |
| anti_10418_36 | Syntaxin-12 | STX12 | Human |
| anti_10419_1 | Scavenger receptor class A member 5 | SCARA5 | Human |
| anti_10428_1 | Killer cell immunoglobulin-like receptor 2DS2 | KIR2DS2 | Human |
| anti_10438_19 | Granulocyte-macrophage colony-stimulating factor receptor subunit alpha | CSF2RA | Human |
| anti_10451_11 | Nucleobindin-1 | NUCB1 | Human |
| anti_10464_6 | Anthrax toxin receptor 1 | ANTXR1 | Human |
| anti_10476_23 | Fc_MOUSE | Igh | Mouse |
| anti_10480_33 | CD59 glycoprotein | CD59 | Human |
| anti_10496_11 | Calcium-activated chloride channel regulator 1 | CLCA1 | Human |
| anti_10511_10 | Collagen alpha-3(VI) chain | COL6A3 | Human |

| SOMAmer | Target Full Name | Entrez Gene Symbol | Organism |
|---|---|---|---|
| anti_10521_10 | Matrix-remodeling-associated protein 8 | MXRA8 | Human |
| anti_10522_167 | Fc_MOUSE | Igh | Mouse |
| anti_10531_18 | GTPase NRas | NRAS | Human |
| anti_10551_7 | Linker for activation of T-cells family member 1 | LAT | Human |
| anti_10553_8 | Torsin-1A-interacting protein 2 | TOR1AIP2 | Human |
| anti_10557_6 | Testis-expressed sequence 29 protein | TEX29 | Human |
| anti_10565_19 | SLIT and NTRK-like protein 3 | SLITRK3 | Human |
| anti_10569_28 | Microfibrillar-associated protein 2 | MFAP2 | Human |
| anti_10572_65 | Cystatin-8 | CST8 | Human |
| anti_10575_31 | Poly(U)-binding-splicing factor PUF60 | PUF60 | Human |
| anti_10576_7 | Plexin domain-containing protein 2 | PLXDC2 | Human |
| anti_10580_14 | Surfactant-associated protein 2 | SFTA2 | Human |
| anti_10589_7 | Retinoic acid early transcript 1L protein | RAET1L | Human |
| anti_10600_24 | Calmegin | CLGN | Human |
| anti_10606_34 | Torsin-1A-interacting protein 1 | TOR1AIP1 | Human |
| anti_10612_18 | Procollagen-lysine,2-oxoglutarate 5-dioxygenase 3 | PLOD3 | Human |
| anti_10615_18 | Myelin protein P0 | MPZ | Human |
| anti_10616_67 | Podocalyxin-like protein 2 | PODXL2 | Human |
| anti_10620_21 | Beta-microseminoprotein | MSMB | Human |
| anti_10626_116 | Alpha-N-acetylgalactosaminide alpha-2,6-sialyltransferase 3 | ST6GALNAC3 | Human |
| anti_10627_87 | Amyloid-like protein 2 | APLP2 | Human |
| anti_10635_33 | Membrane protein FAM159B | FAM159B | Human |
| anti_10637_50 | UPF0577 protein KIAA1324 | KIAA1324 | Human |
| anti_10639_1 | Fc_MOUSE | Igh | Mouse |
| anti_10643_16 | Noelin-3 | OLFM3 | Human |
| anti_10645_72 | Fc_MOUSE | Igh | Mouse |
| anti_10647_18 | Activating signal cointegrator 1 complex subunit 1 | ASCC1 | Human |
| anti_10677_9 | NADH dehydrogenase [ubiquinone] 1 beta subcomplex subunit 4 | NDUFB4 | Human |
| anti_10692_48 | Fc_MOUSE | Igh | Mouse |
| anti_10701_30 | Fc_MOUSE | Igh | Mouse |
| anti_10702_1 | Collagen alpha-1(XXVIII) chain | COL28A1 | Human |
| anti_10714_7 | Angiotensin-converting enzyme | ACE | Human |
| anti_10715_5 | Fc_MOUSE | Igh | Mouse |
| anti_10721_76 | Heat shock 70 kDa protein 1A | HSPA1A | Human |
| anti_10722_13 | Tyrosine-protein kinase SYK | SYK | Human |
| anti_10723_41 | Fc_MOUSE | Igh | Mouse |
| anti_10731_10 | Heparan sulfate glucosamine 3-O-sulfotransferase 5 | HS3ST5 | Human |
| anti_10737_96 | Leukocyte elastase inhibitor | SERPINB1 | Human |
| anti_10743_13 | SLIT and NTRK-like protein 1 | SLITRK1 | Human |
| anti_10749_18 | Heat shock 70 kDa protein 1A | HSPA1A | Human |
| anti_10753_31 | Disintegrin and metalloproteinase domain-containing protein 7 | ADAM7 | Human |
| anti_10760_107 | Glutamate receptor 4 | GRIA4 | Human |
| anti_10767_52 | G-protein coupled receptor 64 | ADGRG2 | Human |
| anti_10781_19 | C-type lectin domain family 4 member G | CLEC4G | Human |
| anti_10795_32 | Interleukin-15 | IL15 | Human |
| anti_10798_4 | CMRF35-like molecule 2 | CD300E | Human |
| anti_10801_11 | Ephrin-A2 | EFNA2 | Human |
| anti_10817_26 | TOMM20-like protein 1 | TOMM20L | Human |
| anti_10818_36 | Sphingomyelin phosphodiesterase | SMPD1 | Human |
| anti_10823_19 | Fc_MOUSE | Igh | Mouse |
| anti_10833_64 | Hedgehog-interacting protein | HHIP | Human |
| anti_10835_25 | Alpha-1,4-N-acetylglucosaminyltransferase | A4GNT | Human |
| anti_10837_131 | Fc_MOUSE | Igh | Mouse |
| anti_10842_7 | Beta-1,3-galactosyl-O-glycosyl-glycoprotein beta-1,6-N-acetylglucosaminyltransferase 4 | GCNT4 | Human |
| anti_10851_77 | Interleukin-27 subunit beta | EBI3 | Human |
| anti_10854_15 | Beta-1,3-galactosyl-O-glycosyl-glycoprotein beta-1,6-N-acetylglucosaminyltransferase 4 | GCNT4 | Human |
| anti_10855_55 | Plexin-B2 | PLXNB2 | Human |
| anti_10866_60 | Serine/threonine-protein phosphatase 4 regulatory subunit 3A | PPP4R3A | Human |

| SOMAmer | Target Full Name | Entrez Gene Symbol | Organism |
|---|---|---|---|
| anti_10870_32 | Spastin | SPAST | Human |
| anti_10876_300 | BRCA1-associated ATM activator 1 | BRAT1 | Human |
| anti_10880_38 | Protein FAM163B | FAM163B | Human |
| anti_10882_12 | Desmocollin-1 | DSC1 | Human |
| anti_10885_36 | Bombesin receptor-activated protein C6orf89 | C6orf89 | Human |
| anti_10889_2 | Melanocortin-2 receptor accessory protein 2 | MRAP2 | Human |
| anti_10892_8 | Oncostatin-M-specific receptor subunit beta | OSMR | Human |
| anti_10894_25 | Secretagogin | SCGN | Human |
| anti_10907_116 | Neurotrimin | NTM | Human |
| anti_10916_44 | Secretory phospholipase A2 receptor | PLA2R1 | Human |
| anti_10945_11 | Syntaxin-6 | STX6 | Human |
| anti_10956_82 | Glycine cleavage system H protein, mitochondrial | GCSH | Human |
| anti_10961_15 | Retinoic acid receptor responder protein 3 | PLAAT4 | Human |
| anti_10966_1 | Alpha-2-HS-glycoprotein | AHSG | Human |
| anti_10967_12 | Prolactin | PRL | Human |
| anti_10970_3 | Ecto-ADP-ribosyltransferase 3 | ART3 | Human |
| anti_10977_55 | Unique cartilage matrix-associated protein | UCMA | Human |
| anti_10990_21 | Leucine-rich repeat serine/threonine-protein kinase 2 | LRRK2 | Human |
| anti_11067_13 | Osteocalcin | BGLAP | Human |
| anti_11075_59 | Interleukin-6_MOUSE | non-human | Mouse |
| anti_11076_135 | N-acylneuraminate cytidylyltransferase_NEIME | neuA | Neisseria meningitidis |
| anti_11096_57 | HemK methyltransferase family member 2 | N6AMT1 | Human |
| anti_11102_22 | Regenerating islet-derived protein 4 | REG4 | Human |
| anti_11104_13 | Chitinase-3-like protein 1 | CHI3L1 | Human |
| anti_11105_171 | Alpha-enolase | ENO1 | Human |
| anti_11109_56 | Sushi, von Willebrand factor type A, EGF and pentraxin domain-containing protein 1 | SVEP1 | Human |
| anti_11116_16 | Uncharacterized protein C11orf87 | C11orf87 | Human |
| anti_11118_107 | Ras-related protein Rab-17 | RAB17 | Human |
| anti_11120_49 | N-acetyltransferase 14 | NAT14 | Human |
| anti_11121_56 | Brorin | VWC2 | Human |
| anti_11140_56 | Collagen alpha-1(I) chain | COL1A1 | Human |
| anti_11142_11 | Angiopoietin-related protein 1 | ANGPTL1 | Human |
| anti_11143_32 | Noelin-3 | OLFM3 | Human |
| anti_11152_46 | Kallikrein-13 | KLK13 | Human |
| anti_11157_35 | Heat shock 70 kDa protein 1A | HSPA1A | Human |
| anti_11160_56 | RING finger protein 122 | RNF122 | Human |
| anti_11171_25 | Filamin-A | FLNA | Human |
| anti_11174_8 | Thrombospondin type-1 domain-containing protein 7A | THSD7A | Human |
| anti_11177_16 | Keratin, type II cytoskeletal 5 | KRT5 | Human |
| anti_11178_21 | Sushi, von Willebrand factor type A, EGF and pentraxin domain-containing protein 1 | SVEP1 | Human |
| anti_11184_51 | Chromogranin-A | CHGA | Human |
| anti_11196_31 | Collagen alpha-3(VI) chain | COL6A3 | Human |
| anti_11200_52 | Complement component C1q receptor | CD93 | Human |
| anti_11203_97 | Pyruvate kinase PKLR | PKLR | Human |
| anti_11205_10 | Integrin beta-7 | ITGB7 | Human |
| anti_11208_15 | N-acetylglucosamine-1-phosphodiester alpha-N-acetylglucosaminidase | NAGPA | Human |
| anti_11212_7 | Thioredoxin domain-containing protein 5 | TXNDC5 | Human |
| anti_11214_40 | DnaJ homolog subfamily B member 9 | DNAJB9 | Human |
| anti_11215_6 | Cadherin-15 | CDH15 | Human |
| anti_11218_84 | Thiopurine S-methyltransferase | TPMT | Human |
| anti_11241_8 | Argininosuccinate lyase | ASL | Human |
| anti_11243_90 | Adhesion G-protein coupled receptor F1 | ADGRF1 | Human |
| anti_11245_43 | Filamin-A | FLNA | Human |
| anti_11246_3 | BPI fold-containing family B member 1 | BPIFB1 | Human |
| anti_11257_1 | Dihydropteridine reductase | QDPR | Human |
| anti_11258_41 | Mucosal addressin cell adhesion molecule 1 | MADCAM1 | Human |
| anti_11263_57 | Calsequestrin-1 | CASQ1 | Human |
| anti_11265_8 | Retinal dehydrogenase 1 | ALDH1A1 | Human |

| SOMAmer | Target Full Name | Entrez Gene Symbol | Organism |
|---|---|---|---|
| anti_11277_23 | Cyclic AMP-dependent transcription factor ATF-6 alpha | ATF6 | Human |
| anti_11278_4 | Collagen alpha-2(XI) chain | COL11A2 | Human |
| anti_11286_78 | Selenoprotein S | SELENOS | Human |
| anti_11294_7 | Transmembrane and coiled-coil domains protein 3 | TMCC3 | Human |
| anti_11320_29 | E3 ubiquitin-protein ligase CHFR | CHFR | Human |
| anti_11330_15 | Casein kinase II subunit beta | CSNK2B | Human |
| anti_11333_82 | Rho GTPase-activating protein 25 | ARHGAP25 | Human |
| anti_11347_9 | Transaldolase | TALDO1 | Human |
| anti_11348_132 | Prolyl 4-hydroxylase subunit alpha-2 | P4HA2 | Human |
| anti_11354_21 | 1-phosphatidylinositol 4,5-bisphosphate phosphodiesterase beta-1 | PLCB1 | Human |
| anti_11355_10 | Eukaryotic translation initiation factor 5A-2 | EIF5A2 | Human |
| anti_11361_73 | Thymidine phosphorylase | TYMP | Human |
| anti_11364_18 | Mitofusin-1 | MFN1 | Human |
| anti_11369_23 | Alcohol dehydrogenase class-3 | ADH5 | Human |
| anti_11372_2 | Zinc finger protein 18 | ZNF18 | Human |
| anti_11382_5 | Biliverdin reductase A | BLVRA | Human |
| anti_11387_3 | Cyclic AMP-dependent transcription factor ATF-6 beta | ATF6B | Human |
| anti_11391_69 | Mevalonate kinase | MVK | Human |
| anti_11401_181 | E3 ubiquitin-protein ligase RNF146 | RNF146 | Human |
| anti_11416_23 | F-box/LRR-repeat protein 4 | FBXL4 | Human |
| anti_11421_10 | EH domain-containing protein 4 | EHD4 | Human |
| anti_11424_4 | Fumarylacetoacetase | FAH | Human |
| anti_11425_31 | RUN and FYVE domain-containing protein 1 | RUFY1 | Human |
| anti_11430_49 | E3 ubiquitin-protein ligase DTX1 | DTX1 | Human |
| anti_11440_58 | Suppressor of cytokine signaling 3 | SOCS3 | Human |
| anti_11441_11 | Glycogen phosphorylase, liver form | PYGL | Human |
| anti_11448_34 | Galactokinase | GALK1 | Human |
| anti_11450_110 | Protein disulfide-isomerase | P4HB | Human |
| anti_11454_87 | Eukaryotic translation initiation factor 3 subunit G | EIF3G | Human |
| anti_11457_53 | UDP-glucose 4-epimerase | GALE | Human |
| anti_11458_30 | Poly(rC)-binding protein 1 | PCBP1 | Human |
| anti_11476_43 | Vacuolar protein sorting-associated protein 4A | VPS4A | Human |
| anti_11481_25 | Hepatitis A virus cellular receptor 2 | HAVCR2 | Human |
| anti_11486_26 | Zinc finger protein 174 | ZNF174 | Human |
| anti_11493_169 | Dynein light chain 2, cytoplasmic | DYNLL2 | Human |
| anti_11494_4 | RNA polymerase II elongation factor ELL2 | ELL2 | Human |
| anti_11514_196 | CD59 glycoprotein | CD59 | Human |
| anti_11530_37 | Porphobilinogen deaminase | HMBS | Human |
| anti_11536_9 | Histone H2A deubiquitinase MYSM1 | MYSM1 | Human |
| anti_11539_4 | ATP synthase subunit f, mitochondrial | ATP5MF | Human |
| anti_11542_11 | Transmembrane protein 230 | TMEM230 | Human |
| anti_11545_9 | Tumor necrosis factor receptor superfamily member 14 | TNFRSF14 | Human |
| anti_11547_84 | Muscle, skeletal receptor tyrosine-protein kinase | MUSK | Human |
| anti_11556_19 | ADP-ribosylation factor GTPase-activating protein 1 | ARFGAP1 | Human |
| anti_11563_51 | Synaptotagmin-like protein 4 | SYTL4 | Human |
| anti_11567_23 | Zinc finger protein 10 | ZNF10 | Human |
| anti_11571_75 | NKG2-E type II integral membrane protein | KLRC3 | Human |
| anti_11586_2 | Leucine-rich repeat neuronal protein 1 | LRRN1 | Human |
| anti_11608_5 | Microtubule-associated proteins 1A/1B light chain 3B | MAP1LC3B | Human |
| anti_11614_29 | Enhancer of rudimentary homolog | ERH | Human |
| anti_11615_16 | Dual adapter for phosphotyrosine and 3-phosphotyrosine and 3-phosphoinositide | DAPP1 | Human |
| anti_11626_7 | Ubiquitin-conjugating enzyme E2 variant 1 | UBE2V1 | Human |
| anti_11633_89 | Activator of 90 kDa heat shock protein ATPase homolog 1 | AHSA1 | Human |
| anti_11634_32 | Regulator of G-protein signaling 10 | RGS10 | Human |
| anti_11636_33 | Transgelin-2 | TAGLN2 | Human |
| anti_11649_3 | Stromal membrane-associated protein 1 | SMAP1 | Human |
| anti_11659_31 | Clathrin interactor 1 | CLINT1 | Human |

| SOMAmer | Target Full Name | Entrez Gene Symbol | Organism |
|---|---|---|---|
| anti_11666_72 | Regulator of G-protein signaling 8 | RGS8 | Human |
| anti_11672_17 | Kinesin-like protein KIF16B | KIF16B | Human |
| anti_11682_7 | Thioredoxin-interacting protein | TXNIP | Human |
| anti_11696_7 | Cellular retinoic acid-binding protein 2 | CRABP2 | Human |
| anti_11814_29 | Hepatocyte growth factor receptor | MET | Human |
| anti_11816_84 | Tyrosine-protein kinase JAK2 | JAK2 | Human |
| anti_11830_48 | Tyrosine-protein phosphatase non-receptor type 11 | PTPN11 | Human |
| anti_11836_144 | Epithelial discoidin domain-containing receptor 1 | DDR1 | Human |
| anti_11851_21 | Triggering receptor expressed on myeloid cells 2 | TREM2 | Human |
| anti_11895_21 | Fc_MOUSE | Igh | Mouse |
| anti_11918_54 | Fc_MOUSE | Igh | Mouse |
| anti_11926_23 | Alpha- and gamma-adaptin-binding protein p34 | AAGAB | Human |
| anti_12020_39 | Bisphosphoglycerate mutase | BPGM | Human |
| anti_12034_28 | Adenylyl cyclase-associated protein 1 | CAP1 | Human |
| anti_12077_32 | Growth/differentiation factor 8 | MSTN | Human |
| anti_12332_7 | Eukaryotic elongation factor 2 kinase | EEF2K | Human |
| anti_12333_87 | Ribose-5-phosphate isomerase | RPIA | Human |
| anti_12341_8 | Dual specificity protein phosphatase 6 | DUSP6 | Human |
| anti_12347_29 | Cerebral cavernous malformations 2 protein | CCM2 | Human |
| anti_12355_223 | Ribosome-recycling factor, mitochondrial | MRRF | Human |
| anti_12365_108 | RuvB-like 1 | RUVBL1 | Human |
| anti_12370_30 | Apolipoprotein F | APOF | Human |
| anti_12384_92 | COP9 signalosome complex subunit 7b | COPS7B | Human |
| anti_12385_4 | Calpain-3 | CAPN3 | Human |
| anti_12386_11 | Aminopeptidase B | RNPEP | Human |
| anti_12389_4 | Origin recognition complex subunit 6 | ORC6 | Human |
| anti_12392_30 | ADP-ribosylation factor-like protein 1 | ARL 1 | Human |
| anti_12406_119 | GTP-binding protein Di-Ras3 | DIRAS3 | Human |
| anti_12414_31 | 14-3-3 protein beta/alpha | YWHAB | Human |
| anti_12415_122 | Endothelial differentiation-related factor 1 | EDF1 | Human |
| anti_12417_46 | EKC/KEOPS complex subunit TPRKB | TPRKB | Human |
| anti_12420_10 | Glycerol-3-phosphate dehydrogenase 1-like protein | GPD1L | Human |
| anti_12426_19 | MOB kinase activator 1A | MOB1A | Human |
| anti_12427_8 | M-phase inducer phosphatase 2 | CDC25B | Human |
| anti_12431_13 | Protein pelota homolog | PELO | Human |
| anti_12433_8 | ADP-ribosylation factor-like protein 11 | ARL 11 | Human |
| anti_12436_84 | Glutathione S-transferase omega-1 | GSTO1 | Human |
| anti_12446_49 | Glutathione S-transferase A1 | GSTA1 | Human |
| anti_12456_5 | 6-phosphofructo-2-kinase/fructose-2,6-bisphosphatase 3 | PFKFB3 | Human |
| anti_12457_10 | 1,2-dihydroxy-3-keto-5-methylthiopentene dioxygenase | ADI1 | Human |
| anti_12458_79 | Calcineurin B homologous protein 1 | CHP1 | Human |
| anti_12461_8 | NAD-dependent protein deacylase sirtuin-5, mitochondrial | SIRT5 | Human |
| anti_12463_7 | Kelch-like protein 13 | KLHL13 | Human |
| anti_12466_7 | Heterogeneous nuclear ribonucleoprotein A1 | HNRNPA1 | Human |
| anti_12469_19 | Microtubule-associated protein RP/EB family member 1 | MAPRE1 | Human |
| anti_12476_50 | Fructose-2,6-bisphosphatase TIGAR | TIGAR | Human |
| anti_12480_9 | OTU domain-containing protein 5 | OTUD5 | Human |
| anti_12484_67 | Phosphatidylinositol transfer protein beta isoform | PITPNB | Human |
| anti_12488_9 | Malignant T-cell-amplified sequence 1 | MCTS1 | Human |
| anti_12490_92 | Ragulator complex protein LAMTOR3 | LAMTOR3 | Human |
| anti_12491_23 | Chloride intracellular channel protein 4 | CLIC4 | Human |
| anti_12494_99 | Gamma-aminobutyric acid receptor-associated protein-like 2 | GABARAPL2 | Human |
| anti_12500_88 | SUMO-activating enzyme subunit 2 | UBA2 | Human |
| anti_12501_10 | Tubulin-specific chaperone A | TBCA | Human |
| anti_12503_5 | Ovarian cancer G-protein coupled receptor 1 | GPR68 | Human |
| anti_12510_3 | Signal-transducing adaptor protein 1 | STAP1 | Human |
| anti_12517_52 | Programmed cell death protein 5 | PDCD5 | Human |
| anti_12524_18 | Diamine acetyltransferase 2 | SAT2 | Human |

| SOMAmer | Target Full Name | Entrez Gene Symbol | Organism |
|---|---|---|---|
| anti_12529_32 | Inactive peptidyl-prolyl cis-trans isomerase FKBP6 | FKBP6 | Human |
| anti_12530_14 | Cyclin-dependent kinases regulatory subunit 1 | CKS1B | Human |
| anti_12532_28 | Ubiquitin-conjugating enzyme E2 R1 | CDC34 | Human |
| anti_12540_25 | Rho-related GTP-binding protein RhoG | RHOG | Human |
| anti_12549_33 | Hematopoietic prostaglandin D synthase | HPGDS | Human |
| anti_12551_3 | E3 ubiquitin-protein ligase Itchy homolog | ITCH | Human |
| anti_12554_10 | DNA repair protein RAD51 homolog 4 | RAD51D | Human |
| anti_12562_1 | Serine/threonine-protein kinase N1 | PKN1 | Human |
| anti_12563_2 | Tumor necrosis factor alpha-induced protein 8 | TNFAIP8 | Human |
| anti_12564_9 | 60S ribosome subunit biogenesis protein NIP7 homolog | NIP7 | Human |
| anti_12572_236 | Embryonal Fyn-associated substrate | EFS | Human |
| anti_12574_36 | Endothelin-2 | EDN2 | Human |
| anti_12575_30 | C-1-tetrahydrofolate synthase, cytoplasmic | MTHFD1 | Human |
| anti_12599_10 | Fermitin family homolog 3 | FERMT3 | Human |
| anti_12603_87 | Polyadenylate-binding protein 4 | PABPC4 | Human |
| anti_12604_16 | Polycomb protein SCMH1 | SCMH1 | Human |
| anti_12619_14 | Protein phosphatase 1A | PPM1A | Human |
| anti_12620_3 | Septin-11 | SEPTIN11 | Human |
| anti_12621_55 | Serine/threonine-protein phosphatase 2A 65 kDa regulatory subunit A alpha isoform | PPP2R1A | Human |
| anti_12627_97 | Ubiquitin-like modifier-activating enzyme ATG7 | ATG7 | Human |
| anti_12630_8 | Arfaptin-2 | ARFIP2 | Human |
| anti_12632_14 | Arylamine N-acetyltransferase 1 | NAT1 | Human |
| anti_12643_4 | Beta-arrestin-1 | ARRB1 | Human |
| anti_12649_80 | Malate dehydrogenase, mitochondrial | MDH2 | Human |
| anti_12650_43 | Guanine nucleotide-binding protein G(k) subunit alpha | GNAI3 | Human |
| anti_12655_30 | Beta-soluble NSF attachment protein | NAPB | Human |
| anti_12658_72 | Pantothenate kinase 3 | PANK3 | Human |
| anti_12659_13 | Obg-like ATPase 1 | OLA1 | Human |
| anti_12661_44 | Gamma-aminobutyric acid receptor-associated protein-like 1 | GABARAPL1 | Human |
| anti_12665_16 | Interleukin enhancer-binding factor 2 | ILF2 | Human |
| anti_12667_2 | Guanine deaminase | GDA | Human |
| anti_12668_7 | Vacuolar protein sorting-associated protein 4B | VPS4B | Human |
| anti_12675_14 | Aspartate--tRNA ligase, cytoplasmic | DARS1 | Human |
| anti_12681_63 | Ubiquitin carboxyl-terminal hydrolase 21 | USP21 | Human |
| anti_12684_5 | Adseverin | SCIN | Human |
| anti_12689_56 | Actin-related protein 2/3 complex subunit 1B | ARPC1B | Human |
| anti_12690_33 | Septin-10 | SEPTIN10 | Human |
| anti_12704_26 | SH2 domain-containing protein 3C | SH2D3C | Human |
| anti_12706_2 | Serine/threonine-protein kinase MRCK alpha | CDC42BPA | Human |
| anti_12713_365 | Regulator of G-protein signaling 19 | RGS19 | Human |
| anti_12720_71 | Ubiquilin-4 | UBQLN4 | Human |
| anti_12727_7 | Prostaglandin F2 receptor negative regulator | PTGFRN | Human |
| anti_12730_3 | Kinetochore protein NDC80 homolog | NDC80 | Human |
| anti_12732_13 | MAGUK p55 subfamily member 7 | MPP7 | Human |
| anti_12737_12 | Plastin-1 | PLS1 | Human |
| anti_12742_160 | Transmembrane protein 8B | TMEM8B | Human |
| anti_12746_4 | Cytohesin-4 | CYTH4 | Human |
| anti_12747_89 | RNA-binding protein 3 | RBM3 | Human |
| anti_12754_14 | Cold shock domain-containing protein C2 | CSDC2 | Human |
| anti_12756_3 | Transcription regulator protein BACH2 | BACH2 | Human |
| anti_12758_47 | Glutamate receptor ionotropic, delta-2 | GRID2 | Human |
| anti_12768_3 | Protein phosphatase 1 regulatory subunit 3B | PPP1R3B | Human |
| anti_12775_6 | High mobility group protein B3 | HMGB3 | Human |
| anti_12777_11 | Splicing factor 1 | SF1 | Human |
| anti_12779_30 | PR domain zinc finger protein 4 | PRDM4 | Human |
| anti_12783_29 | Heterogeneous nuclear ribonucleoprotein M | HNRNPM | Human |
| anti_12784_10 | Amyloid beta A4 precursor protein-binding family B member 3 | APBB3 | Human |
| anti_12800_5 | Tubulin polymerization-promoting protein family member 2 | TPPP2 | Human |

| SOMAmer | Target Full Name | Entrez Gene Symbol | Organism |
|---|---|---|---|
| anti_12807_89 | Rho GTPase-activating protein 30 | ARHGAP30 | Human |
| anti_12817_1 | GTP-binding protein GEM | GEM | Human |
| anti_12822_34 | Amyloid beta A4 precursor protein-binding family B member 1 | APBB1 | Human |
| anti_12826_5 | Sodium/iodide cotransporter | SLC5A5 | Human |
| anti_12831_21 | Calcineurin B homologous protein 3 | TESC | Human |
| anti_12842_43 | Syntaxin-10 | STX10 | Human |
| anti_12845_18 | Sorting nexin-17 | SNX17 | Human |
| anti_12861_13 | Tropomodulin-3 | TMOD3 | Human |
| anti_12867_40 | Dynein light chain Tctex-type 3 | DYNLT3 | Human |
| anti_12881_17 | Retinol dehydrogenase 16 | RDH16 | Human |
| anti_12891_1 | MICAL-like protein 2 | MICALL2 | Human |
| anti_12893_159 | Gamma-interferon-inducible protein 16 | IFI16 | Human |
| anti_12894_3 | PAX-interacting protein 1 | PAXIP1 | Human |
| anti_12895_28 | Diacylglycerol kinase beta | DGKB | Human |
| anti_12906_137 | Fc_MOUSE | Igh | Mouse |
| anti_12912_25 | Fc_MOUSE | Igh | Mouse |
| anti_12923_51 | Keratin, type I cytoskeletal 17 | KRT17 | Human |
| anti_12934_1 | E3 ISG15--protein ligase HERC5 | HERC5 | Human |
| anti_12936_38 | Retinaldehyde-binding protein 1 | RLBP1 | Human |
| anti_12945_33 | Ras-related protein Rab-18 | RAB18 | Human |
| anti_12954_71 | Peroxisome proliferator-activated receptor alpha | PPARA | Human |
| anti_12960_9 | Glucokinase | GCK | Human |
| anti_13022_20 | Tumor protein p53-inducible protein 11 | TP53I11 | Human |
| anti_13028_2 | Fc_MOUSE | Igh | Mouse |
| anti_13042_7 | Potassium voltage-gated channel subfamily A member 10 | KCNA10 | Human |
| anti_13053_6 | Sodium- and chloride-dependent neutral and basic amino acid transporter B(0+) | SLC6A14 | Human |
| anti_13054_87 | Profilin-2 | PFN2 | Human |
| anti_13066_42 | Enhancer of mRNA-decapping protein 4 | EDC4 | Human |
| anti_13067_5 | cGMP-dependent protein kinase 1, beta isozyme | PRKG1 | Human |
| anti_13073_14 | Acidic leucine-rich nuclear phosphoprotein 32 family member A | ANP32A | Human |
| anti_13078_3 | Centrin-2 | CETN2 | Human |
| anti_13089_6 | Hypoxia-inducible factor 1-alpha | HIF1A | Human |
| anti_13090_17 | Protein S100-A6 | S100A6 | Human |
| anti_13093_6 | Secreted and transmembrane protein 1 | SECTM1 | Human |
| anti_13098_93 | Vascular endothelial growth factor D | VEGFD | Human |
| anti_13102_1 | Protein FAM3D | FAM3D | Human |
| anti_13103_125 | Chorionic somatomammotropin hormone | CSH1\|CSH2 | Human |
| anti_13104_32 | Ephrin-B1 | EFNB1 | Human |
| anti_13105_7 | Synaptosomal-associated protein 25 | SNAP25 | Human |
| anti_13107_9 | Ly6/PLAUR domain-containing protein 3 | LYPD3 | Human |
| anti_13109_82 | Neuronal growth regulator 1 | NEGR1 | Human |
| anti_13111_79 | B-cell lymphoma 6 protein | BCL6 | Human |
| anti_13112_179 | Follistatin-related protein 1 | FSTL1 | Human |
| anti_13113_7 | Osteopontin | SPP1 | Human |
| anti_13116_25 | CD177 antigen | CD177 | Human |
| anti_13117_232 | Choline/ethanolamine kinase | CHKB | Human |
| anti_13118_5 | SPARC-related modular calcium-binding protein 1 | SMOC1 | Human |
| anti_13119_26 | Protein Z-dependent protease inhibitor | SERPINA10 | Human |
| anti_13124_20 | Immunoglobulin superfamily containing leucine-rich repeat protein 2 | ISLR2 | Human |
| anti_13125_45 | Vitronectin | VTN | Human |
| anti_13126_52 | Desmocollin-2 | DSC2 | Human |
| anti_13131_5 | Hexokinase-1 | HK1 | Human |
| anti_13132_14 | Semaphorin-5A | SEMA5A | Human |
| anti_13133_73 | Latent-transforming growth factor beta-binding protein 4 | LTBP4 | Human |
| anti_13231_90 | Ig gamma-4, Kappa | IGHG4 | Human |
| anti_13236_25 | Protein Wnt-3a | WNT3A | Human |
| anti_13374_4 | Beta-defensin 113 | DEFB113 | Human |
| anti_13377_3 | Cytochrome b561 domain-containing protein 1 | CYB561D1 | Human |
| anti_13378_80 | Glutaredoxin-like protein C5orf63 | C5orf63 | Human |

| SOMAmer | Target Full Name | Entrez Gene Symbol | Organism |
| --- | --- | --- | --- |
| anti_13381_49 | Beta-1,4-galactosyltransferase 1 | B4GALT1 | Human |
| anti_13389_8 | Fc_MOUSE | Igh | Mouse |
| anti_13392_13 | Sodium/potassium-transporting ATPase subunit beta-1 | ATP1B1 | Human |
| anti_13393_46 | Derlin-1 | DERL1 | Human |
| anti_13397_88 | Beta-defensin 4A | DEFB4A | Human |
| anti_13400_13 | Inactive tyrosine-protein kinase transmembrane receptor ROR1 | ROR1 | Human |
| anti_13408_23 | WAP, Kazal, immunoglobulin, Kunitz and NTR domain-containing protein 2 | WFIKKN2 | Human |
| anti_13416_8 | Transmembrane protein 132D | TMEM132D | Human |
| anti_13422_66 | Ecto-NOX disulfide-thiol exchanger 2 | ENOX2 | Human |
| anti_13424_51 | Fc_MOUSE | Igh | Mouse |
| anti_13427_66 | Mannosyl-oligosaccharide 1,2-alpha-mannosidase IC | MAN1C1 | Human |
| anti_13430_50 | Urea transporter 1 | SLC14A1 | Human |
| anti_13438_115 | Chordin | CHRD | Human |
| anti_13447_42 | Shadow of prion protein | SPRN | Human |
| anti_13455_10 | Fc_MOUSE | Igh | Mouse |
| anti_13460_4 | Chondroadherin | CHAD | Human |
| anti_13464_8 | Nutritionally-regulated adipose and cardiac enriched protein homolog | NRAC | Human |
| anti_13465_5 | Calcipressin-1 | RCAN1 | Human |
| anti_13468_5 | Neuralized-like protein 4 | NEURL4 | Human |
| anti_13472_35 | Haloacid dehalogenase-like hydrolase domain-containing protein 2 | HDHD2 | Human |
| anti_13474_40 | Glutathione S-transferase kappa 1 | GSTK1 | Human |
| anti_13475_10 | Ubiquitin-conjugating enzyme E2 D4 | UBE2D4 | Human |
| anti_13479_8 | Protein FAM171A2 | FAM171A2 | Human |
| anti_13485_20 | Transmembrane protein 87B | TMEM87B | Human |
| anti_13486_9 | Polycystin-2 | PKD2 | Human |
| anti_13488_3 | Arfaptin-1 | ARFIP1 | Human |
| anti_13491_40 | Retinal rod rhodopsin-sensitive cGMP 3',5'-cyclic phosphodiesterase subunit delta | PDE6D | Human |
| anti_13495_48 | Hydroxycarboxylic acid receptor 2 | HCAR2 | Human |
| anti_13496_19 | Hydroxymethylglutaryl-CoA synthase, cytoplasmic | HMGCS1 | Human |
| anti_13498_1 | Inositol oxygenase | MIOX | Human |
| anti_13499_30 | Coagulation Factor VIII | F8 | Human |
| anti_13500_9 | Fc_MOUSE | Igh | Mouse |
| anti_13501_10 | Solute carrier family 35 member G2 | SLC35G2 | Human |
| anti_13507_51 | Tumor necrosis factor receptor type 1-associated DEATH domain protein | TRADD | Human |
| anti_13510_7 | Sarcoplasmic/endoplasmic reticulum calcium ATPase 3 | ATP2A3 | Human |
| anti_13516_46 | ER membrane protein complex subunit 4 | EMC4 | Human |
| anti_13518_5 | Arf-GAP with SH3 domain, ANK repeat and PH domain-containing protein 2 | ASAP2 | Human |
| anti_13522_20 | Visinin-like protein 1 | VSNL1 | Human |
| anti_13525_17 | Potassium voltage-gated channel subfamily G member 4 | KCNG4 | Human |
| anti_13529_39 | Nucleosome assembly protein 1-like 2 | NAP1L2 | Human |
| anti_13540_1 | G-protein coupled receptor 26 | GPR26 | Human |
| anti_13547_5 | 5-hydroxytryptamine receptor 7 | HTR7 | Human |
| anti_13561_5 | 5-hydroxytryptamine receptor 6 | HTR6 | Human |
| anti_13565_2 | Retinoblastoma-like protein 2 | RBL2 | Human |
| anti_13567_1 | Dihydropyrimidinase-related protein 2 | DPYSL2 | Human |
| anti_13572_43 | 26S proteasome non-ATPase regulatory subunit 11 | PSMD11 | Human |
| anti_13573_5 | Actin-related protein 2/3 complex subunit 3 | ARPC3 | Human |
| anti_13575_40 | Segment polarity protein dishevelled homolog DVL-2 | DVL2 | Human |
| anti_13576_15 | Glutathione S-transferase P | GSTP1 | Human |
| anti_13583_19 | Apoptotic protease-activating factor 1 | APAF1 | Human |
| anti_13589_10 | Amyloid beta A4 precursor protein-binding family B member 3 | APBB3 | Human |

| SOMAmer | Target Full Name | Entrez Gene Symbol | Organism |
|---|---|---|---|
| anti_13590_1 | Oligoribonuclease, mitochondrial | REXO2 | Human |
| anti_13596_3 | Ras-related protein Rab-27B | RAB27B | Human |
| anti_13603_7 | Rieske domain-containing protein | RFESD | Human |
| anti_13605_16 | Annexin A10 | ANXA10 | Human |
| anti_13610_9 | Melanoma-associated antigen 10 | MAGEA10 | Human |
| anti_13614_6 | CREB-binding protein | CREBBP | Human |
| anti_13615_60 | Cytoplasmic protein NCK2 | NCK2 | Human |
| anti_13622_16 | Serine/threonine-protein phosphatase 2A 56 kDa regulatory subunit alpha isoform | PPP2R5A | Human |
| anti_13628_58 | Growth factor receptor-bound protein 14 | GRB14 | Human |
| anti_13629_25 | Cysteine protease ATG4B | ATG4B | Human |
| anti_13632_10 | Zyxin | ZYX | Human |
| anti_13636_20 | Nucleosome assembly protein 1-like 1 | NAP1L1 | Human |
| anti_13652_2 | TOM1-like protein 1 | TOM1L1 | Human |
| anti_13659_36 | Manganese-transporting ATPase 13A1 | ATP13A1 | Human |
| anti_13669_6 | Fibroblast growth factor receptor 3 | FGFR3 | Human |
| anti_13670_81 | Group IIE secretory phospholipase A2 | PLA2G2E | Human |
| anti_13672_3 | Heat shock 70 kDa protein 6 | HSPA6 | Human |
| anti_13673_21 | T-complex protein 1 subunit eta | CCT7 | Human |
| anti_13676_46 | Inhibin beta B chain | INHBB | Human |
| anti_13683_18 | Chloride channel protein ClC-Kb | CLCNKB | Human |
| anti_13691_10 | Sodium-coupled monocarboxylate transporter 1 | SLC5A8 | Human |
| anti_13697_51 | Glycerol-3-phosphate dehydrogenase [NAD(+)], cytoplasmic | GPD1 | Human |
| anti_13700_10 | Annexin A2 | ANXA2 | Human |
| anti_13707_27 | SPARC-like protein 1 | SPARCL1 | Human |
| anti_13712_104 | Granzyme A | GZMA | Human |
| anti_13716_259 | Prothrombin | F2 | Human |
| anti_13717_15 | Ficolin-2 | FCN2 | Human |
| anti_13720_95 | Myeloblastin | PRTN3 | Human |
| anti_13728_19 | Endoplasmic reticulum resident protein 29 | ERP29 | Human |
| anti_13730_18 | Dipeptidyl peptidase 1 | CTSC | Human |
| anti_13732_79 | Cardiotrophin-1 | CTF1 | Human |
| anti_13740_51 | Secreted frizzled-related protein 3 | FRZB | Human |
| anti_13741_36 | Insulin-like growth factor-binding protein 1 | IGFBP1 | Human |
| anti_13742_66 | Interleukin-22 | IL22 | Human |
| anti_13744_37 | Interleukin-3 receptor subunit alpha | IL3RA | Human |
| anti_13747_9 | Carbonic anhydrase 6 | CA6 | Human |
| anti_13931_22 | 26S proteasome non-ATPase regulatory subunit 9 | PSMD9 | Human |
| anti_13932_45 | Rho guanine nucleotide exchange factor 7 | ARHGEF7 | Human |
| anti_13933_276 | NGFI-A-binding protein 1 | NAB1 | Human |
| anti_13937_75 | Cullin-associated NEDD8-dissociated protein 1 | CAND1 | Human |
| anti_13941_82 | Phenylalanine--tRNA ligase, mitochondrial | FARS2 | Human |
| anti_13946_8 | Probable ATP-dependent RNA helicase DDX23 | DDX23 | Human |
| anti_13948_50 | Gamma-aminobutyric acid type B receptor subunit 2 | GABBR2 | Human |
| anti_13959_7 | Cytosol aminopeptidase | LAP3 | Human |
| anti_13961_18 | Kinesin-like protein KIF3A | KIF3A | Human |
| anti_13963_7 | Toll-interacting protein | TOLLIP | Human |
| anti_13966_30 | Myomesin-3 | MYOM3 | Human |
| anti_13967_14 | Thioredoxin reductase 1, cytoplasmic | TXNRD1 | Human |
| anti_13983_27 | Quinone oxidoreductase | CRYZ | Human |
| anti_13988_67 | NmrA-like family domain-containing protein 1 | NMRAL1 | Human |
| anti_13991_47 | Eukaryotic translation initiation factor 4 gamma 3 | EIF4G3 | Human |
| anti_13992_12 | Vesicle-fusing ATPase | NSF | Human |
| anti_13993_20 | Band 4.1-like protein 1 | EPB41L1 | Human |
| anti_13994_1 | rRNA 2'-O-methyltransferase fibrillarin | FBL | Human |
| anti_14008_22 | mRNA-decapping enzyme 1A | DCP1A | Human |
| anti_14009_65 | Tumor necrosis factor alpha-induced protein 3 | TNFAIP3 | Human |
| anti_14011_17 | Protein S100-A11 | S100A11 | Human |
| anti_14023_84 | Bone sialoprotein 2 | IBSP | Human |
| anti_14028_22 | Phosphatidylinositol 4-phosphate 3-kinase C2 domain-containing subunit alpha | PIK3C2A | Human |
| anti_14029_42 | COP9 signalosome complex subunit 2 | COPS2 | Human |
| anti_14038_130 | Cystatin-F | CST7 | Human |

| SOMAmer | Target Full Name | Entrez Gene Symbol | Organism |
|---|---|---|---|
| anti_14047_78 | Brain-derived neurotrophic factor | BDNF | Human |
| anti_14054_17 | Interleukin-15 receptor subunit alpha | IL15RA | Human |
| anti_14057_68 | Tumor necrosis factor ligand superfamily member 15 | TNFSF15 | Human |
| anti_14068_29 | C-C motif chemokine 25 | CCL25 | Human |
| anti_14072_9 | Translationally-controlled tumor protein | TPT1 | Human |
| anti_14077_6 | Ficolin-3 | FCN3 | Human |
| anti_14078_69 | Lymphotactin | XCL1 | Human |
| anti_14084_191 | Fc_MOUSE | Igh | Mouse |
| anti_14085_28 | Interleukin-13 | IL13 | Human |
| anti_14086_11 | Acid sphingomyelinase-like phosphodiesterase 3a | SMPDL3A | Human |
| anti_14089_41 | Ephrin type-A receptor 5 | EPHA5 | Human |
| anti_14094_29 | Heparin-binding EGF-like growth factor | HBEGF | Human |
| anti_14097_86 | Carbohydrate sulfotransferase 15 | CHST15 | Human |
| anti_14101_2 | Ciliary neurotrophic factor receptor subunit alpha | CNTFR | Human |
| anti_14102_6 | Granulysin | GNLY | Human |
| anti_14107_1 | 5-formyltetrahydrofolate cyclo-ligase | MTHFS | Human |
| anti_14109_15 | C-C motif chemokine 15 | CCL15 | Human |
| anti_14110_200 | SPARC | SPARC | Human |
| anti_14115_34 | Adrenomedullin | ADM | Human |
| anti_14116_129 | Protein S100-A4 | S100A4 | Human |
| anti_14120_2 | E3 ubiquitin-protein ligase RNF43 | RNF43 | Human |
| anti_14123_34 | V-type immunoglobulin domain-containing suppressor of T-cell activation | VSIR | Human |
| anti_14124_6 | Ephrin-A2 | EFNA2 | Human |
| anti_14132_21 | HERV-H LTR-associating protein 2 | HHLA2 | Human |
| anti_14133_93 | Interleukin-1 receptor type 2 | IL1R2 | Human |
| anti_14136_234 | Complement component C1q receptor | CD93 | Human |
| anti_14144_3 | Histone H2A type 3 | HIST3H2A | Human |
| anti_14148_2 | Ubiquitin-like protein ISG15 | ISG15 | Human |
| anti_14150_7 | Interleukin-36 alpha | IL36A | Human |
| anti_14156_33 | 14-3-3 protein beta/alpha | YWHAB | Human |
| anti_14186_13 | E3 ubiquitin-protein ligase rififylin | RFFL | Human |
| anti_14192_31 | Phosducin-like protein 2 | PDCL2 | Human |
| anti_14204_55 | Forkhead box protein J2 | FOXJ2 | Human |
| anti_14208_3 | Retinoid-binding protein 7 | RBP7 | Human |
| anti_14222_68 | Protein S100-A5 | S100A5 | Human |
| anti_14226_120 | S-methylmethionine--homocysteine S-methyltransferase BHMT2 | BHMT2 | Human |
| anti_14229_5 | Adenylyltransferase and sulfurtransferase MOCS3 | MOCS3 | Human |
| anti_14254_27 | Tyrosine-protein phosphatase non-receptor type 4 | PTPN4 | Human |
| anti_14268_4 | Sulfiredoxin-1 | SRXN1 | Human |
| anti_14283_12 | Ras-related protein Rab-14 | RAB14 | Human |
| anti_14287_6 | Ras-related protein Rab-5C | RAB5C | Human |
| anti_14294_61 | Methyl-CpG-binding domain protein 1 | MBD1 | Human |
| anti_14309_8 | Heterogeneous nuclear ribonucleoprotein H | HNRNPH1 | Human |
| anti_14318_1 | Vacuolar protein sorting-associated protein 29 | VPS29 | Human |
| anti_14332_3 | Ras-related protein Ral-A | RALA | Human |
| anti_14337_1 | Trafficking protein particle complex subunit 3 | TRAPPC3 | Human |
| anti_14341_8 | Protein FAM69C | DIPK1C | Human |
| anti_14582_57 | Growth/differentiation factor 11 | GDF11 | Human |
| anti_14583_49 | Growth/differentiation factor 8 | MSTN | Human |
| anti_14593_152 | Protein FAM210A | FAM210A | Human |
| anti_14614_41 | Noncompact myelin-associated protein | NCMAP | Human |
| anti_14633_26 | Transmembrane protein 185A | TMEM185A | Human |
| anti_14643_27 | DNA/RNA-binding protein KIN17 | KIN | Human |
| anti_14663_44 | E3 ubiquitin-protein ligase RNF8 | RNF8 | Human |
| anti_14670_1 | Ski-like protein | SKIL | Human |
| anti_14675_20 | Eukaryotic translation initiation factor 4B | EIF4B | Human |
| anti_14684_17 | Calpain-2 catalytic subunit | CAPN2 | Human |
| anti_14685_17 | RAC-beta serine/threonine-protein kinase | AKT2 | Human |
| anti_14689_3 | Protein SEC13 homolog | SEC13 | Human |
| anti_14692_3 | Zinc finger protein 276 | ZNF276 | Human |
| anti_14705_1 | Vascular endothelial growth factor D | VEGFD | Human |
| anti_14711_27 | Cystatin-M | CST6 | Human |

| SOMAmer | Target Full Name | Entrez Gene Symbol | Organism |
|---|---|---|---|
| anti_14756_29 | Intercellular adhesion molecule 2 | ICAM2 | Human |
| anti_14759_149 | Cadherin-1 | CDH1 | Human |
| anti_2201_17 | Endostatin | COL18A1 | Human |
| anti_2212_69 | Tissue-type plasminogen activator | PLAT | Human |
| anti_2358_19 | Vascular endothelial growth factor receptor 3 | FLT4 | Human |
| anti_2421_7 | Brain-derived neurotrophic factor | BDNF | Human |
| anti_2429_27 | Complement component C8 | C8A\|C8B\|C8G | Human |
| anti_2436_49 | C-X-C motif chemokine 16 | CXCL16 | Human |
| anti_2475_1 | Mast/stem cell growth factor receptor Kit | KIT | Human |
| anti_2480_58 | Metalloproteinase inhibitor 3 | TIMP3 | Human |
| anti_2501_51 | Cadherin-1 | CDH1 | Human |
| anti_2515_14 | GDNF family receptor alpha-2 | GFRA2 | Human |
| anti_2516_57 | C-C motif chemokine 21 | CCL21 | Human |
| anti_2524_56 | High mobility group protein B1 | HMGB1 | Human |
| anti_2570_72 | Insulin-like growth factor-binding protein 2 | IGFBP2 | Human |
| anti_2571_12 | Insulin-like growth factor-binding protein 3 | IGFBP3 | Human |
| anti_2575_5 | Leptin | LEP | Human |
| anti_2580_83 | Myeloperoxidase | MPO | Human |
| anti_2585_2 | Prolactin | PRL | Human |
| anti_2590_69 | Inactive tyrosine-protein kinase transmembrane receptor ROR1 | ROR1 | Human |
| anti_2597_8 | Vascular endothelial growth factor A | VEGFA | Human |
| anti_2602_2 | Angiopoietin-2 | ANGPT2 | Human |
| anti_2605_49 | Tumor necrosis factor receptor superfamily member 8 | TNFRSF8 | Human |
| anti_2617_56 | Receptor tyrosine-protein kinase erbB-3 | ERBB3 | Human |
| anti_2620_4 | Interleukin-6 receptor subunit beta | IL6ST | Human |
| anti_2625_53 | Hsp90alpha | HSP90AA1 | Human |
| anti_2630_12 | Interleukin-1 Receptor accessory protein | IL1RAP | Human |
| anti_2633_52 | Interleukin-13 receptor subunit alpha-1 | IL13RA1 | Human |
| anti_2636_10 | Tumor necrosis factor receptor superfamily member 3 | LTBR | Human |
| anti_2637_77 | Macrophage mannose receptor 1 | MRC1 | Human |
| anti_2643_57 | Cadherin-3 | CDH3 | Human |
| anti_2647_66 | Rab GDP dissociation inhibitor beta | GDI2 | Human |
| anti_2652_15 | Urokinase plasminogen activator surface receptor | PLAUR | Human |
| anti_2654_19 | Tumor necrosis factor receptor superfamily member 1A | TNFRSF1A | Human |
| anti_2665_26 | Tumor necrosis factor receptor superfamily member 17 | TNFRSF17 | Human |
| anti_2668_70 | Calpain I | CAPN1\|CAPNS1 | Human |
| anti_2670_67 | Creatine kinase M-type | CKM | Human |
| anti_2677_1 | Epidermal growth factor receptor | EGFR | Human |
| anti_2682_68 | 60 kDa heat shock protein, mitochondrial | HSPD1 | Human |
| anti_2686_67 | Insulin-like growth factor-binding protein 6 | IGFBP6 | Human |
| anti_2687_2 | Melanoma-derived growth regulatory protein | MIA | Human |
| anti_2692_74 | Phospholipase A2, membrane associated | PLA2G2A | Human |
| anti_2693_20 | Oncostatin-M | OSM | Human |
| anti_2714_78 | Endothelial monocyte-activating polypeptide 2 | AIMP1 | Human |
| anti_2742_68 | Sialic acid-binding Ig-like lectin 7 | SIGLEC7 | Human |
| anti_2762_30 | Fibroblast growth factor 19 | FGF19 | Human |
| anti_2765_4 | Growth/differentiation factor 11/8 | GDF11\|MSTN | Human |
| anti_2770_51 | C-C motif chemokine 1 | CCL1 | Human |
| anti_2771_35 | Insulin-like growth factor-binding protein 1 | IGFBP1 | Human |
| anti_2774_10 | Interleukin-16 | IL16 | Human |
| anti_2778_10 | Interleukin-22 | IL22 | Human |
| anti_2783_18 | C-C motif chemokine 3-like 1 | CCL3L1 | Human |
| anti_2788_55 | Stromelysin-1 | MMP3 | Human |
| anti_2789_26 | Matrilysin | MMP7 | Human |
| anti_2796_62 | Fibrinogen | FGA\|FGB\|FGG | Human |
| anti_2811_27 | Angiopoietin-1 | ANGPT1 | Human |
| anti_2819_23 | Cadherin-5 | CDH5 | Human |
| anti_2822_56 | CD97 antigen | CD97 | Human |
| anti_2837_3 | Hepatocyte growth factor receptor | MET | Human |
| anti_2841_13 | Secreted frizzled-related protein 3 | FRZB | Human |

| SOMAmer | Target Full Name | Entrez Gene Symbol | Organism |
|---|---|---|---|
| anti_2844_53 | Tyrosine-protein kinase receptor Tie-1, soluble | TIE1 | Human |
| anti_2846_24 | Ubiquitin+1, truncated mutation for UbB | RPS27A | Human |
| anti_2849_49 | Allograft inflammatory factor 1 | AIF1 | Human |
| anti_2851_63 | C5a anaphylatoxin | C5 | Human |
| anti_2855_49 | Mitogen-activated protein kinase 3 | MAPK3 | Human |
| anti_2857_70 | Glucocorticoid receptor | NR3C1 | Human |
| anti_2859_69 | Histone deacetylase 8 | HDAC8 | Human |
| anti_2860_19 | Importin subunit alpha-1 | KPNA2 | Human |
| anti_2869_68 | Protein kinase C delta type | PRKCD | Human |
| anti_2870_29 | Ras-related C3 botulinum toxin substrate 1 | RAC1 | Human |
| anti_2871_73 | DNA repair protein RAD51 homolog 1 | RAD51 | Human |
| anti_2877_3 | SUMO-conjugating enzyme UBC9 | UBE2I | Human |
| anti_2900_53 | C-C motif chemokine 14 | CCL14 | Human |
| anti_2913_1 | C-C motif chemokine 23 | CCL23 | Human |
| anti_2917_3 | Tumor necrosis factor ligand superfamily member 11 | TNFSF11 | Human |
| anti_2925_9 | Plasminogen activator inhibitor 1 | SERPINE1 | Human |
| anti_2942_50 | Cytochrome c | CYCS | Human |
| anti_2946_52 | Complement factor D | CFD | Human |
| anti_2948_58 | Growth hormone receptor | GHR | Human |
| anti_2953_31 | Luteinizing hormone | CGA\|LHB | Human |
| anti_2954_56 | Neutrophil collagenase | MMP8 | Human |
| anti_2966_65 | Stem cell growth factor-beta | CLEC11A | Human |
| anti_2967_8 | Vascular cell adhesion protein 1 | VCAM1 | Human |
| anti_2974_61 | Contactin-1 | CNTN1 | Human |
| anti_2976_58 | Desmoglein-1 | DSG1 | Human |
| anti_2981_9 | Endothelial cell-selective adhesion molecule | ESAM | Human |
| anti_2985_35 | Growth-regulated alpha protein | CXCL1 | Human |
| anti_2986_49 | Gro-gamma | CXCL3 | Human |
| anti_2992_59 | Interleukin-17 receptor A | IL17RA | Human |
| anti_2997_8 | Junctional adhesion molecule B | JAM2 | Human |
| anti_2999_6 | Limbic system-associated membrane protein | LSAMP | Human |
| anti_3000_66 | Mannose-binding protein C | MBL2 | Human |
| anti_3005_5 | Tyrosine-protein phosphatase non-receptor type 1 | PTPN1 | Human |
| anti_3007_7 | Sialic acid-binding Ig-like lectin 9 | SIGLEC9 | Human |
| anti_3009_3 | Transforming growth factor beta receptor type 3 | TGFBR3 | Human |
| anti_3026_5 | Calpastatin | CAST | Human |
| anti_3029_52 | CD209 antigen | CD209 | Human |
| anti_3030_3 | C-type lectin domain family 4 member M | CLEC4M | Human |
| anti_3032_11 | Follicle stimulating hormone | CGA\|FSHB | Human |
| anti_3035_80 | Interleukin-19 | IL19 | Human |
| anti_3037_62 | Interleukin-1 beta | IL1B | Human |
| anti_3041_55 | C-type mannose receptor 2 | MRC2 | Human |
| anti_3042_7 | Myoglobin | MB | Human |
| anti_3045_72 | Pleiotrophin | PTN | Human |
| anti_3046_31 | Resistin | RETN | Human |
| anti_3049_61 | Trypsin-1 | PRSS1 | Human |
| anti_3061_61 | Cathepsin B | CTSB | Human |
| anti_3066_12 | Galectin-3 | LGALS3 | Human |
| anti_3067_67 | Growth/differentiation factor 9 | GDF9 | Human |
| anti_3122_6 | Diablo homolog, mitochondrial | DIABLO | Human |
| anti_3148_49 | Gro-beta | CXCL2 | Human |
| anti_3152_57 | Tumor necrosis factor receptor superfamily member 1B | TNFRSF1B | Human |
| anti_3169_70 | Alpha-L-iduronidase | IDUA | Human |
| anti_3172_28 | Arylsulfatase B | ARSB | Human |
| anti_3175_51 | A disintegrin and metalloproteinase with thrombospondin motifs 13 | ADAMTS13 | Human |
| anti_3179_51 | Lysosomal protective protein | CTSA | Human |
| anti_3181_50 | Cathepsin S | CTSS | Human |
| anti_3182_38 | Ectonucleoside triphosphate diphosphohydrolase 1 | ENTPD1 | Human |
| anti_3194_36 | Platelet glycoprotein VI | GP6 | Human |
| anti_3195_50 | Granulysin | GNLY | Human |
| anti_3197_70 | Insulin-degrading enzyme | IDE | Human |
| anti_3198_4 | Iduronate 2-sulfatase | IDS | Human |

| SOMAmer | Target Full Name | Entrez Gene Symbol | Organism |
|---|---|---|---|
| anti_3199_54 | Kallikrein-12 | KLK12 | Human |
| anti_3213_65 | Nidogen-1 | NID1 | Human |
| anti_3214_3 | Neuropilin-1 | NRP1 | Human |
| anti_3216_2 | Polymeric immunoglobulin receptor | PIGR | Human |
| anti_3217_74 | Glia-derived nexin | SERPINE2 | Human |
| anti_3218_8 | Glutamate carboxypeptidase 2 | FOLH1 | Human |
| anti_3220_40 | Proto-oncogene tyrosine-protein kinase receptor Ret | RET | Human |
| anti_3232_28 | Tartrate-resistant acid phosphatase type 5 | ACP5 | Human |
| anti_3234_23 | Coiled-coil domain-containing protein 80 | CCDC80 | Human |
| anti_3283_21 | Transforming growth factor-beta-induced protein ig-h3 | TGFBI | Human |
| anti_3285_23 | Complement C1r subcomponent | C1R | Human |
| anti_3291_30 | Low affinity immunoglobulin epsilon Fc receptor | FCER2 | Human |
| anti_3292_75 | CD48 antigen | CD48 | Human |
| anti_3298_52 | Contactin-4 | CNTN4 | Human |
| anti_3302_58 | Cystatin-F | CST7 | Human |
| anti_3303_23 | Cystatin-M | CST6 | Human |
| anti_3305_6 | Delta-like protein 4 | DLL4 | Human |
| anti_3309_2 | Low affinity immunoglobulin gamma Fc region receptor II-a | FCGR2A | Human |
| anti_3310_62 | Low affinity immunoglobulin gamma Fc region receptor II-b | FCGR2B | Human |
| anti_3313_21 | Ficolin-2 | FCN2 | Human |
| anti_3320_49 | Insulin-like growth factor-binding protein 7 | IGFBP7 | Human |
| anti_3325_2 | Matrilin-2 | MATN2 | Human |
| anti_3327_27 | Netrin-4 | NTN4 | Human |
| anti_3329_14 | Peptidoglycan recognition protein 1 | PGLYRP1 | Human |
| anti_3331_8 | RGM domain family member B | RGMB | Human |
| anti_3332_57 | Hemojuvelin | HJV | Human |
| anti_3336_50 | Tissue factor pathway inhibitor | TFPI | Human |
| anti_3339_33 | Thrombospondin-2 | THBS2 | Human |
| anti_3340_53 | Thrombospondin-4 | THBS4 | Human |
| anti_3343_1 | Aminoacylase-1 | ACY1 | Human |
| anti_3348_49 | Bone morphogenetic protein 1 | BMP1 | Human |
| anti_3352_80 | Carbonic anhydrase 6 | CA6 | Human |
| anti_3362_61 | Chordin-like protein 1 | CHRDL1 | Human |
| anti_3363_31 | Tyrosine-protein kinase CSK | CSK | Human |
| anti_3365_7 | Dickkopf-related protein 4 | DKK 4 | Human |
| anti_3378_49 | Kallikrein-7 | KLK7 | Human |
| anti_3381_24 | Tyrosine-protein kinase Lyn, isoform B | LYN | Human |
| anti_3391_10 | Phosphatidylinositol 4,5-bisphosphate 3-kinase catalytic subunit gamma isoform | PIK3CG | Human |
| anti_3392_68 | Protein kinase B gamma | AKT3 | Human |
| anti_3396_54 | Renin | REN | Human |
| anti_3399_31 | Stabilin-2 | STAB2 | Human |
| anti_3403_1 | Tryptase beta-2 | TPSB2 | Human |
| anti_3413_50 | Bcl-2-related protein A1 | BCL2A1 | Human |
| anti_3416_2 | Tyrosine-protein kinase BTK | BTK | Human |
| anti_3418_12 | Calcium/calmodulin-dependent protein kinase type 1D | CAMK1D | Human |
| anti_3419_49 | Calcium/calmodulin-dependent protein kinase type II subunit delta | CAMK2D | Human |
| anti_3431_54 | Ephrin type-A receptor 1 | EPHA1 | Human |
| anti_3437_80 | Receptor-type tyrosine-protein kinase FLT3 | FLT3 | Human |
| anti_3438_10 | Follistatin-related protein 3 | FSTL3 | Human |
| anti_3446_7 | Interleukin-18 receptor 1 | IL18R1 | Human |
| anti_3450_4 | Kallikrein-6 | KLK6 | Human |
| anti_3470_1 | E-selectin | SELE | Human |
| anti_3471_49 | Serine/threonine-protein kinase 16 | STK16 | Human |
| anti_3477_63 | High affinity nerve growth factor receptor | NTRK1 | Human |
| anti_3488_64 | Catalase | CAT | Human |
| anti_3503_4 | Integrin alpha-I: beta-1 complex | ITGA1\|ITGB1 | Human |
| anti_3504_58 | Hepcidin | HAMP | Human |
| anti_3506_49 | Lymphotoxin alpha2:beta1 | LTA\|LTB | Human |

| SOMAmer | Target Full Name | Entrez Gene Symbol | Organism |
|---|---|---|---|
| anti_3508_78 | C-C motif chemokine 22 | CCL22 | Human |
| anti_3509_1 | C-C motif chemokine 15 | CCL15 | Human |
| anti_3514_49 | Myeloblastin | PRTN3 | Human |
| anti_3516_60 | Stromal cell-derived factor 1 | CXCL12 | Human |
| anti_3519_3 | C-C motif chemokine 17 | CCL17 | Human |
| anti_3535_84 | Dickkopf-related protein 1 | DKK 1 | Human |
| anti_3583_54 | Arylsulfatase A | ARSA | Human |
| anti_3585_54 | Basigin | BSG | Human |
| anti_3587_53 | Bone morphogenetic protein 10 | BMP10 | Human |
| anti_3591_51 | Cadherin-6 | CDH6 | Human |
| anti_3592_4 | Calcium/calmodulin-dependent protein kinase type 1 | CAMK1 | Human |
| anti_3600_2 | Chitotriosidase-1 | CHIT1 | Human |
| anti_3601_54 | Neural cell adhesion molecule L1-like protein | CHL1 | Human |
| anti_3603_60 | C-type lectin domain family 7 member A | CLEC7A | Human |
| anti_3605_77 | Mannan-binding lectin serine protease 1 | MASP1 | Human |
| anti_3606_2 | Discoidin domain-containing receptor 2 | DDR2 | Human |
| anti_3607_71 | Dickkopf-related protein 3 | DKK 3 | Human |
| anti_3608_12 | Dipeptidyl peptidase 2 | DPP7 | Human |
| anti_3611_70 | Endothelin-converting enzyme 1 | ECE1 | Human |
| anti_3612_6 | Ephrin type-B receptor 4 | EPHB4 | Human |
| anti_3613_62 | Ficolin-1 | FCN1 | Human |
| anti_3622_33 | Legumain | LGMN | Human |
| anti_3623_84 | Lymphocyte antigen 86 | LY86 | Human |
| anti_3624_3 | Serine protease 27 | PRSS27 | Human |
| anti_3627_71 | Membrane metallo-endopeptidase-like 1 | MMEL1 | Human |
| anti_3628_3 | Dual specificity mitogen-activated protein kinase kinase 2 | MAP2K2 | Human |
| anti_3629_60 | Serine/threonine-protein kinase MRCK beta | CDC42BPB | Human |
| anti_3630_27 | Cell adhesion molecule 3 | CADM3 | Human |
| anti_3633_70 | Nidogen-2 | NID2 | Human |
| anti_3634_5 | Opioid-binding protein/cell adhesion molecule | OPCML | Human |
| anti_3635_76 | OCIA domain-containing protein 1 | OCIAD1 | Human |
| anti_3636_37 | Oxidized low-density lipoprotein receptor 1 | OLR1 | Human |
| anti_3640_14 | alpha-2-macroglobulin receptor-associated protein | LRPAP1 | Human |
| anti_3642_4 | SLAM family member 5 | CD84 | Human |
| anti_3643_90 | SLIT and NTRK-like protein 1 | SLITRK1 | Human |
| anti_3644_5 | Dickkopf-like protein 1 | DKKL1 | Human |
| anti_3646_7 | Tyrosine-protein kinase Tec | TEC | Human |
| anti_3647_49 | Toll-like receptor 4:Lymphocyte antigen 96 complex | TLR4ǀLY96 | Human |
| anti_3650_8 | Green fluorescent protein_AEQVI | GFP | Jellyfish |
| anti_3651_50 | Vascular endothelial growth factor receptor 2 | KDR | Human |
| anti_3657_74 | Calcineurin subunit B type 1 | PPP3R1 | Human |
| anti_3666_17 | Complement factor H-related protein 5 | CFHR5 | Human |
| anti_3676_15 | Cation-independent mannose-6-phosphate receptor | IGF2R | Human |
| anti_3681_87 | Kallikrein-14 | KLK14 | Human |
| anti_3685_53 | Membrane frizzled-related protein | MFRP | Human |
| anti_3708_62 | Alpha-2-macroglobulin | A2M | Human |
| anti_3709_4 | Alanine aminotransferase 1 | GPT | Human |
| anti_3710_49 | Angiostatin | PLG | Human |
| anti_3714_49 | Creatine kinase M-type:Creatine kinase B-type heterodimer | CKBǀCKM | Human |
| anti_3720_67 | Alpha-hemolysin_STAAU | hly | Staphylococcus aureus |
| anti_3736_60 | Bone morphogenetic protein 6 | BMP6 | Human |
| anti_3737_6 | Cathepsin H | CTSH | Human |
| anti_3739_72 | Integrin alpha-IIb: beta-3 complex | ITGA2BǀITGB3 | Human |
| anti_3741_4 | Interleukin-5 | IL5 | Human |
| anti_3748_74 | Apolipoprotein A-I_MOUSE | Apoa1 | Mouse |
| anti_3758_63 | Activated Protein C | PROC | Human |
| anti_3758_68 | Activated Protein C | PROC | Human |
| anti_3796_79 | Angiopoietin-related protein 4 | ANGPTL4 | Human |
| anti_3799_11 | Carbonic anhydrase 3 | CA3 | Human |
| anti_3805_16 | Endothelial cell-specific molecule 1 | ESM1 | Human |

| SOMAmer | Target Full Name | Entrez Gene Symbol | Organism |
|---|---|---|---|
| anti_3820_68 | MAP kinase-activated protein kinase 2 | MAPKAPK2 | Human |
| anti_3822_54 | MAP kinase-activated protein kinase 3 | MAPKAPK3 | Human |
| anti_3844_2 | Peptidyl-prolyl cis-trans isomerase A | PPIA | Human |
| anti_3847_56 | Persulfide dioxygenase ETHE1, mitochondrial | ETHE1 | Human |
| anti_3848_14 | Glyceraldehyde-3-phosphate dehydrogenase | GAPDH | Human |
| anti_3853_56 | Malate dehydrogenase, cytoplasmic | MDH1 | Human |
| anti_3855_56 | Peroxiredoxin-1 | PRDX1 | Human |
| anti_3872_2 | Translationally-controlled tumor protein | TPT1 | Human |
| anti_3877_67 | Calcium/calmodulin-dependent protein kinase kinase 1 | CAMKK1 | Human |
| anti_3888_8 | Inhibitor of growth protein 1 | ING1 | Human |
| anti_3894_15 | N-acetyl-D-glucosamine kinase | NAGK | Human |
| anti_3902_21 | S-phase kinase-associated protein 1 | SKP1 | Human |
| anti_3905_62 | Ubiquitin-conjugating enzyme E2 N | UBE2N | Human |
| anti_4125_52 | Advanced glycosylation end product-specific receptor, soluble | AGER | Human |
| anti_4126_22 | Bactericidal permeability-increasing protein | BPI | Human |
| anti_4135_84 | Immunoglobulin E | IGHE\|IGK\|IGL | Human |
| anti_4139_71 | Interleukin-6 receptor subunit alpha | IL6R | Human |
| anti_4148_49 | Pappalysin-1 | PAPPA | Human |
| anti_4149_8 | Platelet-derived growth factor subunit B | PDGFB | Human |
| anti_4154_57 | P-selectin | SELP | Human |
| anti_4155_3 | Tenascin | TNC | Human |
| anti_4157_2 | Thrombin | F2 | Human |
| anti_4160_49 | 72 kDa type IV collagenase | MMP2 | Human |
| anti_4179_57 | 14-3-3 protein gamma | YWHAG | Human |
| anti_4187_49 | 6-phosphogluconate dehydrogenase, decarboxylating | PGD | Human |
| anti_4192_10 | Alcohol dehydrogenase [NADP(+)] | AKR1A1 | Human |
| anti_4194_26 | Acidic leucine-rich nuclear phosphoprotein 32 family member B | ANP32B | Human |
| anti_4203_50 | Cofilin-1 | CFL1 | Human |
| anti_4209_60 | Vacuolar protein sorting-associated protein VTA1 homolog | VTA1 | Human |
| anti_4234_8 | Interleukin-1 receptor-like 1 | IL1RL1 | Human |
| anti_4237_70 | Leucine carboxyl methyltransferase 1 | LCMT1 | Human |
| anti_4240_31 | Pyruvate kinase PKM | PKM | Human |
| anti_4246_40 | Neural cell adhesion molecule L1 | L1CAM | Human |
| anti_4258_15 | Proliferation-associated protein 2G4 | PA2G4 | Human |
| anti_4272_46 | Glucose-6-phosphate isomerase | GPI | Human |
| anti_4276_10 | Phosphatidylethanolamine-binding protein 1 | PEBP1 | Human |
| anti_4282_3 | GTP-binding nuclear protein Ran | RAN | Human |
| anti_4292_5 | Alpha-soluble NSF attachment protein | NAPA | Human |
| anti_4297_62 | Spondin-1 | SPON1 | Human |
| anti_4306_4 | Transketolase | TKT | Human |
| anti_4309_59 | Triosephosphate isomerase | TPI1 | Human |
| anti_4314_12 | dCTP pyrophosphatase 1 | DCTPP1 | Human |
| anti_4332_6 | C-type lectin domain family 1 member B | CLEC1B | Human |
| anti_4336_2 | Serum amyloid A-1 protein | SAA1 | Human |
| anti_4337_49 | C-reactive protein | CRP | Human |
| anti_4342_10 | Intercellular adhesion molecule 1 | ICAM1 | Human |
| anti_4355_13 | Death-associated protein kinase 2 | DAPK2 | Human |
| anti_4359_87 | Dual specificity tyrosine-phosphorylation-regulated kinase 3 | DYRK3 | Human |
| anti_4374_45 | Growth/differentiation factor 15 | GDF15 | Human |
| anti_4413_3 | Antileukoproteinase | SLPI | Human |
| anti_4414_69 | Pulmonary surfactant-associated protein D | SFTPD | Human |
| anti_4430_44 | Collectin-11 | COLEC11 | Human |
| anti_4437_56 | Ectonucleoside triphosphate diphosphohydrolase 5 | ENTPD5 | Human |
| anti_4450_26 | Heterogeneous nuclear ribonucleoprotein A/B | HNRNPAB | Human |
| anti_4467_49 | SPARC-like protein 1 | SPARCL1 | Human |
| anti_4469_78 | Carbohydrate sulfotransferase 15 | CHST15 | Human |
| anti_4474_19 | Ubiquitin | RPS27A | Human |
| anti_4476_22 | Tyrosine-protein kinase ZAP-70 | ZAP70 | Human |
| anti_4479_14 | Plasma protease C1 inhibitor | SERPING1 | Human |

| SOMAmer | Target Full Name | Entrez Gene Symbol | Organism |
|---|---|---|---|
| anti_4498_62 | Neural cell adhesion molecule 1, 120 kDa isoform | NCAM1 | Human |
| anti_4499_21 | Platelet-derived growth factor subunit A | PDGFA | Human |
| anti_4500_50 | Stem cell growth factor-alpha | CLEC11A | Human |
| anti_4541_49 | Cell adhesion molecule-related/down-regulated by oncogenes | CDON | Human |
| anti_4542_24 | Clusterin | CLU | Human |
| anti_4546_27 | Adhesion G protein-coupled receptor E2 | ADGRE2 | Human |
| anti_4562_1 | Neurexophilin-1 | NXPH1 | Human |
| anti_4564_2 | Plexin-C1 | PLXNC1 | Human |
| anti_4567_82 | SH2 domain-containing protein 1A | SH2D1A | Human |
| anti_4593_11 | Pituitary adenylate cyclase-activating polypeptide 27 | ADCYAP1 | Human |
| anti_4706_17 | Protein 4.1 | EPB41 | Human |
| anti_4707_50 | 14-3-3 protein eta | YWHAH | Human |
| anti_4708_3 | Estradiol 17-beta-dehydrogenase 1 | HSD17B1 | Human |
| anti_4712_28 | Apolipoprotein D | APOD | Human |
| anti_4718_5 | Peptidyl-prolyl cis-trans isomerase B | PPIB | Human |
| anti_4719_58 | Protein disulfide-isomerase A3 | PDIA3 | Human |
| anti_4721_54 | Trefoil factor 3 | TFF3 | Human |
| anti_4771_10 | Acid sphingomyelinase-like phosphodiesterase 3a | SMPDL3A | Human |
| anti_4786_58 | UMP-CMP kinase | CMPK1 | Human |
| anti_4792_51 | gp41 C34 peptide, HIV | env | HIV-1 |
| anti_4815_25 | Thioredoxin domain-containing protein 12 | TXNDC12 | Human |
| anti_4829_43 | 14-3-3 protein sigma | SFN | Human |
| anti_4851_25 | Interleukin-1 alpha | IL1A | Human |
| anti_4866_59 | BDNF/NT-3 growth factors receptor | NTRK2 | Human |
| anti_4903_72 | Calcineurin | PPP3CA\|PPP3R1 | Human |
| anti_4905_63 | Coactosin-like protein | COTL1 | Human |
| anti_4907_56 | D-dimer | FGA\|FGB\|FGG | Human |
| anti_4911_49 | Glutathione S-transferase P | GSTP1 | Human |
| anti_4912_17 | Aspartate aminotransferase, cytoplasmic | GOT1 | Human |
| anti_4913_78 | C-C motif chemokine 16 | CCL16 | Human |
| anti_4915_64 | Hemoglobin | HBA1\|HBB | Human |
| anti_4916_2 | Immunoglobulin D | IGHD\|IGK\|IGL | Human |
| anti_4923_79 | Muellerian-inhibiting factor | AMH | Human |
| anti_4924_32 | Interstitial collagenase | MMP1 | Human |
| anti_4930_21 | Stanniocalcin-1 | STC1 | Human |
| anti_4957_1 | 40S ribosomal protein SA | RPSA | Human |
| anti_4959_2 | Anterior gradient protein 2 homolog | AGR2 | Human |
| anti_4960_72 | Annexin A1 | ANXA1 | Human |
| anti_4964_67 | Endoplasmic reticulum aminopeptidase 1 | ERAP1 | Human |
| anti_4965_27 | ATP synthase subunit beta, mitochondrial | ATP5F1B | Human |
| anti_4967_1 | Complement component 1 Q subcomponent-binding protein, mitochondrial | C1QBP | Human |
| anti_4969_2 | Carbonic anhydrase 1 | CA1 | Human |
| anti_4971_1 | Cathepsin Z | CTSZ | Human |
| anti_4978_54 | Drebrin-like protein | DBNL | Human |
| anti_4979_34 | Dermatopontin | DPT | Human |
| anti_4982_54 | Elafin | PI3 | Human |
| anti_4984_83 | S-formylglutathione hydrolase | ESD | Human |
| anti_4985_11 | Fatty acid-binding protein, epidermal | FABP5 | Human |
| anti_4988_49 | Fibroblast growth factor receptor 4 | FGFR4 | Human |
| anti_4989_7 | Fibrinogen gamma chain | FGG | Human |
| anti_4990_87 | Platelet glycoprotein Ib alpha chain | GP1BA | Human |
| anti_4992_49 | Granulins | GRN | Human |
| anti_4995_16 | 15-hydroxyprostaglandin dehydrogenase [NAD(+)] | HPGD | Human |
| anti_5000_52 | Galectin-3-binding protein | LGALS3BP | Human |
| anti_5001_6 | Mammaglobin-B | SCGB2A1 | Human |
| anti_5007_1 | Mitogen-activated protein kinase 14 | MAPK14 | Human |
| anti_5008_51 | Superoxide dismutase [Mn], mitochondrial | SOD2 | Human |
| anti_5012_67 | Adenylate kinase isoenzyme 1 | AK1 | Human |
| anti_5013_2 | Chloride intracellular channel protein 1 | CLIC1 | Human |
| anti_5015_15 | Platelet-activating factor acetylhydrolase | PLA2G7 | Human |
| anti_5018_68 | Peroxiredoxin-6 | PRDX6 | Human |

| SOMAmer | Target Full Name | Entrez Gene Symbol | Organism |
|---|---|---|---|
| anti_5019_16 | Ubiquitin carboxyl-terminal hydrolase isozyme L1 | UCHL1 | Human |
| anti_5020_50 | Phosphoglycerate kinase 1 | PGK 1 | Human |
| anti_5021_13 | Inorganic pyrophosphatase | PPA1 | Human |
| anti_5023_23 | Adenylosuccinate lyase | ADSL | Human |
| anti_5028_59 | Scavenger receptor cysteine-rich type 1 protein M130 | CD163 | Human |
| anti_5029_3 | Prolyl endopeptidase FAP | FAP | Human |
| anti_5030_52 | NAD-dependent protein deacetylase sirtuin-2 | SIRT2 | Human |
| anti_5034_79 | Trypsin-2 | PRSS2 | Human |
| anti_5061_27 | ICOS ligand | ICOSLG | Human |
| anti_5066_134 | CMRF35-like molecule 6 | CD300C | Human |
| anti_5069_9 | Complement decay-accelerating factor | CD55 | Human |
| anti_5077_28 | Ephrin type-B receptor 2 | EPHB2 | Human |
| anti_5078_82 | Ephrin type-B receptor 6 | EPHB6 | Human |
| anti_5080_131 | Transmembrane glycoprotein NMB | GPNMB | Human |
| anti_5082_51 | X-linked interleukin-1 receptor accessory protein-like 2 | IL1RAPL2 | Human |
| anti_5084_154 | Interleukin-17 receptor B | IL17RB | Human |
| anti_5090_49 | Leukocyte immunoglobulin-like receptor subfamily B member 1 | LILRB1 | Human |
| anti_5091_28 | Leukocyte immunoglobulin-like receptor subfamily B member 2 | LILRB2 | Human |
| anti_5092_51 | Protein jagged-1 | JAG1 | Human |
| anti_5104_57 | Natural cytotoxicity triggering receptor 1 | NCR1 | Human |
| anti_5105_2 | Reticulon-4 receptor | RTN4R | Human |
| anti_5107_7 | Neurogenic locus notch homolog protein 1 | NOTCH1 | Human |
| anti_5111_15 | Neurexin-3-beta | NRXN3 | Human |
| anti_5114_65 | Prolactin receptor | PRLR | Human |
| anti_5116_62 | Roundabout homolog 2 | ROBO2 | Human |
| anti_5125_6 | Sialic acid-binding Ig-like lectin 14 | SIGLEC14 | Human |
| anti_5129_12 | Scavenger receptor class F member 1 | SCARF1 | Human |
| anti_5132_71 | Interleukin-27 receptor subunit alpha | IL27RA | Human |
| anti_5139_32 | Netrin receptor UNC5C | UNC5C | Human |
| anti_5227_60 | [Pyruvate dehydrogenase (acetyl-transferring)] kinase isozyme 1, mitochondrial | PDK1 | Human |
| anti_5242_37 | Dual specificity mitogen-activated protein kinase kinase 4 | MAP2K4 | Human |
| anti_5246_64 | cGMP-dependent 3',5'-cyclic phosphodiesterase | PDE2A | Human |
| anti_5247_17 | Peptidyl-prolyl cis-trans isomerase A | PPIA | Human |
| anti_5249_31 | Serine/threonine-protein kinase 17B | STK17B | Human |
| anti_5250_53 | Inosine-5'-monophosphate dehydrogenase 2 | IMPDH2 | Human |
| anti_5252_33 | Dual 3',5'-cyclic-AMP and -GMP phosphodiesterase 11A | PDE11A | Human |
| anti_5254_69 | cGMP-inhibited 3',5'-cyclic phosphodiesterase A | PDE3A | Human |
| anti_5255_22 | cAMP-specific 3',5'-cyclic phosphodiesterase 4D | PDE4D | Human |
| anti_5256_86 | cGMP-specific 3',5'-cyclic phosphodiesterase | PDE5A | Human |
| anti_5259_2 | Mitogen-activated protein kinase kinase kinase 7:TGF-beta-activated kinase 1 and MAP3K7-binding protein 1 fusion | MAP3K7\|TAB1 | Human |
| anti_5260_80 | Non-receptor tyrosine-protein kinase TYK2 | TYK2 | Human |
| anti_5261_13 | Abelson tyrosine-protein kinase 2 | ABL2 | Human |
| anti_5262_57 | Breast cancer anti-estrogen resistance protein 3 | BCAR3 | Human |
| anti_5264_65 | Calreticulin | CALR | Human |
| anti_5272_55 | SHC-transforming protein 1 | SHC1 | Human |
| anti_5301_7 | Eotaxin | CCL11 | Human |
| anti_5308_89 | Neutrophil elastase | ELANE | Human |
| anti_5315_22 | Troponin T, cardiac muscle | TNNT2 | Human |
| anti_5335_73 | Annexin A6 | ANXA6 | Human |
| anti_5340_24 | Caspase-10 | CASP10 | Human |
| anti_5345_51 | Cytoskeleton-associated protein 2 | CKAP2 | Human |
| anti_5346_24 | Copine-1 | CPNE1 | Human |
| anti_5349_69 | Delta-like protein 1 | DLL1 | Human |
| anti_5350_14 | Glypican-6 | GPC6 | Human |
| anti_5351_52 | Heterogeneous nuclear ribonucleoproteins A2/B1 | HNRNPA2B1 | Human |
| anti_5353_89 | Interleukin-1 receptor antagonist protein | IL1RN | Human |

| SOMAmer | Target Full Name | Entrez Gene Symbol | Organism |
|---|---|---|---|
| anti_5356_2 | Macrophage migration inhibitory factor | MIF | Human |
| anti_5358_3 | Osteomodulin | OMD | Human |
| anti_5360_9 | RAC-beta serine/threonine-protein kinase | AKT2 | Human |
| anti_5364_7 | Protein SET | SET | Human |
| anti_5383_14 | Tumor necrosis factor receptor superfamily member 13C | TNFRSF13C | Human |
| anti_5384_67 | Transcription factor IIIB 90 kDa subunit | BRF1 | Human |
| anti_5392_73 | Tumor necrosis factor receptor superfamily member 6 | FAS | Human |
| anti_5400_52 | Leptin receptor, soluble | LEPR | Human |
| anti_5404_53 | Tumor necrosis factor receptor superfamily member 21 | TNFRSF21 | Human |
| anti_5410_53 | Cadherin-15 | CDH15 | Human |
| anti_5424_55 | Tumor necrosis factor receptor superfamily member 11A | TNFRSF11A | Human |
| anti_5437_63 | Fatty acid-binding protein, heart | FABP3 | Human |
| anti_5439_51 | Neutrophil gelatinase-associated lipocalin_RAT | non-human | Rat |
| anti_5440_26 | Troponin I, fast skeletal muscle | TNNI2 | Human |
| anti_5451_1 | CD166 antigen | ALCAM | Human |
| anti_5452_71 | Asialoglycoprotein receptor 1 | ASGR1 | Human |
| anti_5459_33 | Cystatin-SN | CST1 | Human |
| anti_5463_22 | Growth arrest-specific protein 1 | GAS1 | Human |
| anti_5464_52 | Growth factor receptor-bound protein 2 | GRB2 | Human |
| anti_5465_32 | Heparan-sulfate 6-O-sulfotransferase 1 | HS6ST1 | Human |
| anti_5467_15 | Heat shock protein HSP 90-beta | HSP90AB1 | Human |
| anti_5470_69 | Mediator of RNA polymerase II transcription subunit 1 | MED1 | Human |
| anti_5480_49 | C-C motif chemokine 5 | CCL5 | Human |
| anti_5483_1 | Repulsive guidance molecule A | RGMA | Human |
| anti_5486_73 | Intercellular adhesion molecule 2 | ICAM2 | Human |
| anti_5487_7 | SLAM family member 7 | SLAMF7 | Human |
| anti_5489_18 | Stress-induced-phosphoprotein 1 | STIP1 | Human |
| anti_5490_53 | Testican-1 | SPOCK1 | Human |
| anti_5491_12 | Testican-2 | SPOCK2 | Human |
| anti_5493_17 | Serine/threonine-protein kinase WNK3 | WNK3 | Human |
| anti_5494_52 | Small nuclear ribonucleoprotein F | SNRPF | Human |
| anti_5496_49 | Spondin-1 | SPON1 | Human |
| anti_5497_29 | Fibroblast growth factor receptor 2_MOUSE | Fgfr2 | Mouse |
| anti_5509_7 | Epidermal growth factor | EGF | Human |
| anti_5581_28 | Fibrinogen-like protein 1 | FGL1 | Human |
| anti_5584_21 | Holo-Transcobalamin-2 | TCN2 | Human |
| anti_5596_75 | T-cell surface glycoprotein CD5 | CD5 | Human |
| anti_5599_88 | Prenylcysteine oxidase-like | PCYOX1L | Human |
| anti_5605_77 | Beta-1,3-N-acetylglucosaminyltransferase manic fringe | MFNG | Human |
| anti_5617_41 | Pro-FMRFamide-related neuropeptide FF | NPFF | Human |
| anti_5618_50 | Protein FAM3B | FAM3B | Human |
| anti_5620_13 | Peptidyl-glycine alpha-amidating monooxygenase | PAM | Human |
| anti_5628_21 | Semaphorin-3G | SEMA3G | Human |
| anti_5635_66 | Triggering receptor expressed on myeloid cells 2 | TREM2 | Human |
| anti_5636_10 | Microfibril-associated glycoprotein 4 | MFAP4 | Human |
| anti_5638_23 | Procollagen galactosyltransferase 1 | COLGALT1 | Human |
| anti_5644_60 | Ribonuclease 4 | RNASE4 | Human |
| anti_5646_20 | Ribonuclease K6 | RNASE6 | Human |
| anti_5648_28 | Chymotrypsinogen B2 | CTRB2 | Human |
| anti_5650_9 | Protein disulfide-isomerase A6 | PDIA6 | Human |
| anti_5654_70 | Protein disulfide-isomerase TMX3 | TMX3 | Human |
| anti_5660_51 | Extracellular superoxide dismutase [Cu-Zn] | SOD3 | Human |
| anti_5675_6 | Tenascin | TNC | Human |
| anti_5688_65 | Cerebellin-4 | CBLN4 | Human |
| anti_5690_49 | Tuftelin | TUFT1 | Human |
| anti_5691_2 | Cysteine-rich secretory protein LCCL domain-containing 2 | CRISPLD2 | Human |
| anti_5694_57 | SPARC-related modular calcium-binding protein 1 | SMOC1 | Human |
| anti_5698_60 | Tenascin-X | TNXB | Human |

| SOMAmer | Target Full Name | Entrez Gene Symbol | Organism |
|---|---|---|---|
| anti_5708_1 | Liver-expressed antimicrobial peptide 2 | LEAP2 | Human |
| anti_5722_78 | Lysosomal Pro-X carboxypeptidase | PRCP | Human |
| anti_5725_1 | Osteomodulin | OMD | Human |
| anti_5735_54 | C1GALT1-specific chaperone 1 | C1GALT1C1 | Human |
| anti_5736_1 | Trem-like transcript 2 protein | TREML2 | Human |
| anti_5738_25 | Tenascin | TNC | Human |
| anti_5739_75 | Keratinocyte differentiation-associated protein | KRTDAP | Human |
| anti_5741_55 | Eosinophil cationic protein | RNASE3 | Human |
| anti_5843_60 | Histone-lysine N-methyltransferase EHMT2 | EHMT2 | Human |
| anti_5846_24 | Noggin | NOG | Human |
| anti_5852_6 | Protein S100-A12 | S100A12 | Human |
| anti_5858_6 | 14-3-3 protein zeta/delta | YWHAZ | Human |
| anti_5861_78 | 3-hydroxyanthranilate 3,4-dioxygenase | HAAO | Human |
| anti_5864_10 | Fructose-bisphosphate aldolase A | ALDOA | Human |
| anti_5879_51 | Dynactin subunit 2 | DCTN2 | Human |
| anti_5885_55 | Eukaryotic translation initiation factor 4H | EIF4H | Human |
| anti_5888_29 | Eukaryotic translation initiation factor 5A-1 | EIF5A | Human |
| anti_5903_91 | Heat shock cognate 71 kDa protein | HSPA8 | Human |
| anti_5909_51 | Nucleoside diphosphate kinase A | NME1 | Human |
| anti_5918_5 | Proteasome activator complex subunit 1 | PSME1 | Human |
| anti_5934_1 | Ferritin | FTH1\|FTL | Human |
| anti_5960_49 | Fc_MOUSE | Igh | Mouse |
| anti_5963_9 | Dermokine | DMKN | Human |
| anti_5988_49 | Tumor necrosis factor ligand superfamily member 14 | TNFSF14 | Human |
| anti_6019_12 | Complement C1q and tumor necrosis factor-related protein 9A | C1QTNF9 | Human |
| anti_6022_57 | Protein kinase C-binding protein NELL2 | NELL2 | Human |
| anti_6039_24 | Corticotropin-releasing factor-binding protein | CRHBP | Human |
| anti_6042_52 | Tenascin | TNC | Human |
| anti_6049_64 | Receptor-type tyrosine-protein phosphatase S | PTPRS | Human |
| anti_6055_53 | Interferon alpha/beta receptor 1 | IFNAR1 | Human |
| anti_6064_4 | Endoplasmic reticulum resident protein 44 | ERP44 | Human |
| anti_6069_71 | Sulfatase-modifying factor 2 | SUMF2 | Human |
| anti_6077_63 | Adenosine deaminase CECR1 | ADA2 | Human |
| anti_6079_59 | 2-phosphoxylose phosphatase 1 | PXYLP1 | Human |
| anti_6081_52 | Procollagen C-endopeptidase enhancer 2 | PCOLCE2 | Human |
| anti_6086_15 | Chordin-like protein 2 | CHRDL2 | Human |
| anti_6103_70 | Fc receptor-like protein 5 | FCRL5 | Human |
| anti_6104_1 | Fibroblast growth factor receptor-like 1 | FGFRL1 | Human |
| anti_6107_3 | Chymotrypsin-like elastase family member 1 | CELA1 | Human |
| anti_6115_40 | Fc_MOUSE | Igh | Mouse |
| anti_6117_4 | Neuroendocrine convertase 2 | PCSK2 | Human |
| anti_6123_69 | Cellular tumor antigen p53 | TP53 | Human |
| anti_6152_111 | Cellular tumor antigen p53 | TP53 | Human |
| anti_6172_7 | PolyUbiquitin K63-linked | UBC | Human |
| anti_6213_10 | Choriogonadotropin subunit beta variant 2 | CGB2 | Human |
| anti_6227_1 | Kallikrein-10 | KLK10 | Human |
| anti_6229_13 | Chorionic somatomammotropin hormone | CSH1\|CSH2 | Human |
| anti_6232_54 | T-lymphocyte activation antigen CD86 | CD86 | Human |
| anti_6252_62 | Secretoglobin family 3A member 1 | SCGB3A1 | Human |
| anti_6255_74 | Probable carboxypeptidase X1 | CPXM1 | Human |
| anti_6256_9 | Lymphocyte antigen 6 complex locus protein G6c | LY6G6C | Human |
| anti_6257_56 | Cell growth regulator with EF hand domain protein 1 | CGREF1 | Human |
| anti_6260_14 | Tenascin | TNC | Human |
| anti_6273_58 | Prolyl 3-hydroxylase 1 | P3H1 | Human |
| anti_6276_16 | Furin | FURIN | Human |
| anti_6281_51 | CMP-N-acetylneuraminate-beta-galactosamide-alpha-2,3-sialyltransferase 2 | ST3GAL2 | Human |
| anti_6283_60 | Mast cell-expressed membrane protein 1 | MCEMP1 | Human |
| anti_6285_71 | Malectin | MLEC | Human |
| anti_6295_67 | Fc_MOUSE | Igh | Mouse |
| anti_6297_49 | Heparan sulfate glucosamine 3-O-sulfotransferase 1 | HS3ST1 | Human |

| SOMAmer | Target Full Name | Entrez Gene Symbol | Organism |
|---|---|---|---|
| anti_6300_14 | Fc_MOUSE | Igh | Mouse |
| anti_6304_8 | Complement C1q tumor necrosis factor-related protein 1 | C1QTNF1 | Human |
| anti_6342_10 | Nephronectin | NPNT | Human |
| anti_6353_60 | Fc_MOUSE | Igh | Mouse |
| anti_6356_3 | Carboxypeptidase B | CPB1 | Human |
| anti_6357_83 | Chymotrypsin-like elastase family member 3B | CELA3B | Human |
| anti_6362_6 | Augurin | ECRG4 | Human |
| anti_6364_7 | Tapasin-related protein | TAPBPL | Human |
| anti_6366_38 | Thioredoxin domain-containing protein 15 | TXNDC15 | Human |
| anti_6368_9 | Cystatin-like 1 | CSTL1 | Human |
| anti_6369_82 | Deformed epidermal autoregulatory factor 1 homolog | DEAF1 | Human |
| anti_6372_7 | Y-box-binding protein 2 | YBX2 | Human |
| anti_6373_54 | Protein delta homolog 1 | DLK1 | Human |
| anti_6378_2 | Protein CEI | C5orf38 | Human |
| anti_6382_17 | Beta-mannosidase | MANBA | Human |
| anti_6386_4 | Fc_MOUSE | Igh | Mouse |
| anti_6387_61 | Defensin-5 | DEFA5 | Human |
| anti_6388_21 | Coiled-coil domain-containing protein 126 | CCDC126 | Human |
| anti_6389_57 | Fc_MOUSE | Igh | Mouse |
| anti_6392_7 | WNT1-inducible-signaling pathway protein 2 | CCN5 | Human |
| anti_6393_63 | Endoplasmin | HSP90B1 | Human |
| anti_6405_74 | Pregnancy-specific beta-1-glycoprotein 2 | PSG2 | Human |
| anti_6406_3 | Normal mucosa of esophagus-specific gene 1 protein | NMES1 | Human |
| anti_6408_2 | Inhibin beta C chain | INHBC | Human |
| anti_6409_57 | Adhesion G protein-coupled receptor F5 | ADGRF5 | Human |
| anti_6414_8 | Out at first protein homolog | OAF | Human |
| anti_6417_55 | Fc_MOUSE | Igh | Mouse |
| anti_6419_75 | Fc_MOUSE | Igh | Mouse |
| anti_6420_4 | Succinate dehydrogenase assembly factor 2, mitochondrial | SDHAF2 | Human |
| anti_6423_66 | Complement C1q-like protein 2 | C1QL2 | Human |
| anti_6424_2 | Probable inactive ribonuclease-like protein 13 | RNASE13 | Human |
| anti_6425_87 | Matrix metalloproteinase-19 | MMP19 | Human |
| anti_6440_31 | Microfibrillar-associated protein 5 | MFAP5 | Human |
| anti_6451_64 | Asporin | ASPN | Human |
| anti_6453_70 | Leukocyte immunoglobulin-like receptor subfamily B member 4 | LILRB4 | Human |
| anti_6458_6 | Dolichyl-diphosphooligosaccharide--protein glycosyltransferase subunit 1 | RPN1 | Human |
| anti_6461_54 | Apolipoprotein C-III | APOC3 | Human |
| anti_6462_12 | Metalloproteinase inhibitor 4 | TIMP4 | Human |
| anti_6463_59 | Orexigenic neuropeptide QRFP | QRFP | Human |
| anti_6464_40 | Transmembrane protein PVRIG | PVRIG | Human |
| anti_6466_7 | Fc_MOUSE | Igh | Mouse |
| anti_6468_37 | Tachykinin-4 | TAC4 | Human |
| anti_6470_19 | Fibulin-1 | FBLN1 | Human |
| anti_6471_53 | Complement factor H-related protein 4 | CFHR4 | Human |
| anti_6472_40 | Melanocyte protein PMEL | PMEL | Human |
| anti_6480_1 | Soluble calcium-activated nucleotidase 1 | CANT1 | Human |
| anti_6491_59 | Kallikrein-15 | KLK15 | Human |
| anti_6496_60 | Protein delta homolog 1 | DLK1 | Human |
| anti_6506_54 | Transmembrane emp24 domain-containing protein 10 | TMED10 | Human |
| anti_6508_68 | Secretoglobin family 1D member 2 | SCGB1D2 | Human |
| anti_6510_56 | E3 ubiquitin-protein ligase RNF128 | RNF128 | Human |
| anti_6511_17 | Protein FAM19A4 | TAFA4 | Human |
| anti_6517_14 | Growth/differentiation factor 10 | GDF10 | Human |
| anti_6518_85 | Appetite-regulating hormone | GHRL | Human |
| anti_6520_87 | Matrix Gla protein | MGP | Human |
| anti_6521_35 | Neuronal pentraxin-2 | NPTX2 | Human |
| anti_6522_57 | Fc_MOUSE | Igh | Mouse |
| anti_6527_1 | TLR4 interactor with leucine rich repeats | TRIL | Human |

| SOMAmer | Target Full Name | Entrez Gene Symbol | Organism |
|---|---|---|---|
| anti_6536_54 | Putative phospholipase B-like 2 | PLBD2 | Human |
| anti_6545_58 | Major prion protein | PRNP | Human |
| anti_6550_4 | Intercellular adhesion molecule 4 | ICAM4 | Human |
| anti_6551_94 | Serpin A12 | SERPINA12 | Human |
| anti_6555_58 | Stomatin-like protein 2, mitochondrial | STOML2 | Human |
| anti_6556_5 | Ectonucleotide pyrophosphatase/ phosphodiesterase family member 5 | ENPP5 | Human |
| anti_6557_50 | Leucine-rich repeat-containing protein 15 | LRRC15 | Human |
| anti_6561_77 | Ig Kappa chain V-I region HK102- like | None | Human |
| anti_6574_11 | Fas apoptotic inhibitory molecule 3 | FCMR | Human |
| anti_6576_1 | Ecto-ADP-ribosyltransferase 4 | ART4 | Human |
| anti_6586_19 | Disintegrin and metalloproteinase domain-containing protein 11 | ADAM11 | Human |
| anti_6587_6 | Leucine-rich repeat and fibronectin type-III domain-containing protein 5 | LRFN5 | Human |
| anti_6590_54 | Neuropilin-2 | NRP2 | Human |
| anti_6601_1 | Ubiquitin-conjugating enzyme E2 J2 | UBE2J2 | Human |
| anti_6603_18 | Anosmin-1 | ANOS1 | Human |
| anti_6604_59 | Protein NDNF | NDNF | Human |
| anti_6612_90 | BRICHOS domain-containing protein 5 | BRICD5 | Human |
| anti_6617_12 | Fc receptor-like protein 6 | FCRL6 | Human |
| anti_6620_82 | Leucine-rich repeat and immunoglobulin-like domain-containing nogo receptor-interacting protein 1 | LINGO1 | Human |
| anti_6621_10 | Prorelaxin H2 | RLN2 | Human |
| anti_6626_81 | Carbohydrate sulfotransferase 12 | CHST12 | Human |
| anti_6629_3 | Beta-defensin 1 | DEFB1 | Human |
| anti_6631_17 | Collagen alpha-1(IX) chain | COL9A1 | Human |
| anti_6641_60 | PolyUbiquitin K48-linked | UBB | Human |
| anti_6645_53 | Periostin | POSTN | Human |
| anti_6647_55 | PolyUbiquitin K63-linked | UBC | Human |
| anti_6651_74 | PolyUbiquitin K48-linked | UBB | Human |
| anti_6896_3 | Transmembrane protein C1orf162 | C1orf162 | Human |
| anti_6900_30 | Ubiquitin-conjugating enzyme E2 J1 | UBE2J1 | Human |
| anti_6909_40 | Alpha-1,6-mannosyl-glycoprotein 2-beta-N-acetylglucosaminyltransferase | MGAT2 | Human |
| anti_6914_15 | Amphoterin-induced protein 2 | AMIGO2 | Human |
| anti_6919_3 | Hemoglobin subunit zeta | HBZ | Human |
| anti_6927_7 | Bifunctional heparan sulfate N-deacetylase/N-sulfotransferase 1 | NDST1 | Human |
| anti_6931_10 | Cancer/testis antigen 1 | CTAG1A | Human |
| anti_6934_8 | Fc_MOUSE | Igh | Mouse |
| anti_6936_7 | Protein TMEPAI | PMEPA1 | Human |
| anti_6940_18 | Junctophilin-1 | JPH1 | Human |
| anti_6941_11 | Sulfatase-modifying factor 1 | SUMF1 | Human |
| anti_6947_4 | Type 2 lactosamine alpha-2,3-sialyltransferase | ST3GAL6 | Human |
| anti_6948_82 | Polyphosphoinositide phosphatase | FIG4 | Human |
| anti_6955_68 | Sorting nexin-1 | SNX1 | Human |
| anti_6961_14 | Insulin growth factor-like family member 3 | IGFL3 | Human |
| anti_6962_5 | HLA class II histocompatibility antigen, DR beta 3 chain | HLA-DRB3 | Human |
| anti_6963_82 | P-selectin glycoprotein ligand 1 | SELPLG | Human |
| anti_6973_111 | Insulin-like growth factor II | IGF2 | Human |
| anti_6975_52 | SNARE-associated protein Snapin | SNAPIN | Human |
| anti_6984_6 | Immunoglobulin superfamily member 8 | IGSF8 | Human |
| anti_6991_24 | Alpha-(1,3)-fucosyltransferase 9 | FUT9 | Human |
| anti_6994_19 | Uncharacterized protein C3orf18 | C3orf18 | Human |
| anti_6998_106 | Aspartyl/asparaginyl beta-hydroxylase | ASPH | Human |
| anti_7008_13 | Protein eva-1 homolog C | EVA1C | Human |
| anti_7012_35 | Vascular non-inflammatory molecule 2 | VNN 2 | Human |
| anti_7015_8 | Leukocyte immunoglobulin-like receptor subfamily B member 5 | LILRB5 | Human |
| anti_7034_4 | Fc_MOUSE | Igh | Mouse |
| anti_7038_45 | CMP-N-acetylneuraminate-poly-alpha-2,8-sialyltransferase | ST8SIA4 | Human |

| SOMAmer | Target Full Name | Entrez Gene Symbol | Organism |
|---|---|---|---|
| anti_7049_2 | Disintegrin and metalloproteinase domain-containing protein 23 | ADAM23 | Human |
| anti_7050_5 | Neuronal growth regulator 1 | NEGR1 | Human |
| anti_7054_87 | C-type lectin domain family 2 member D | CLEC2D | Human |
| anti_7057_18 | Fc_MOUSE | Igh | Mouse |
| anti_7060_2 | ERO1-like protein alpha | ERO1A | Human |
| anti_7065_1 | Protein G6b | MPIG6B | Human |
| anti_7071_23 | Endoplasmic reticulum mannosyl-oligosaccharide 1,2-alpha-mannosidase | MAN1B1 | Human |
| anti_7082_2 | UDP-GlcNAc:betaGal beta-1,3-N-acetylglucosaminyltransferase 6 | B3GNT6 | Human |
| anti_7085_81 | Corneodesmosin | CDSN | Human |
| anti_7090_17 | Polypeptide N-acetylgalactosaminyltransferase 1 | GALNT1 | Human |
| anti_7106_37 | Oocyte-secreted protein 2 | OOSP2 | Human |
| anti_7110_2 | DnaJ homolog subfamily B member 11 | DNAJB11 | Human |
| anti_7115_5 | Semenogelin-1 | SEMG1 | Human |
| anti_7123_25 | 39S ribosomal protein L52, mitochondrial | MRPL52 | Human |
| anti_7127_3 | Apolipoprotein A-II | APOA2 | Human |
| anti_7131_207 | Tenascin | TNC | Human |
| anti_7136_7 | Fc_MOUSE | Igh | Mouse |
| anti_7139_14 | SLIT and NTRK-like protein 4 | SLITRK4 | Human |
| anti_7140_1 | Chymotrypsin-like elastase family member 2A | CELA2A | Human |
| anti_7142_5 | Carboxypeptidase N catalytic chain | CPN1 | Human |
| anti_7144_234 | Kazal-type serine protease inhibitor domain-containing protein 1 | KAZALD1 | Human |
| anti_7145_1 | Inter-alpha-trypsin inhibitor heavy chain H3 | ITIH3 | Human |
| anti_7148_42 | Protein FAM173A | ANTKMT | Human |
| anti_7152_5 | Hepatitis A virus cellular receptor 2 | HAVCR2 | Human |
| anti_7154_92 | Protein LEG1 homolog | LEG1 | Human |
| anti_7155_46 | C4b-binding protein beta chain | C4BPB | Human |
| anti_7156_2 | Alpha-(1,3)-fucosyltransferase 10 | FUT10 | Human |
| anti_7157_22 | Protocadherin alpha-4 | PCDHA4 | Human |
| anti_7161_25 | GDH/6PGL endoplasmic bifunctional protein | H6PD | Human |
| anti_7167_102 | Vesicle-associated membrane protein-associated protein A | VAPA | Human |
| anti_7173_141 | Transmembrane protein 132C | TMEM132C | Human |
| anti_7178_59 | Protein DEPP | DEPP1 | Human |
| anti_7179_69 | Neurofascin | NFASC | Human |
| anti_7182_1 | Liver carboxylesterase 1 | CES1 | Human |
| anti_7184_13 | Carcinoembryonic antigen-related cell adhesion molecule 7 | CEACAM7 | Human |
| anti_7190_50 | Ephrin type-A receptor 4 | EPHA4 | Human |
| anti_7191_32 | Sarcolemmal membrane-associated protein | SLMAP | Human |
| anti_7194_36 | Neuroplastin | NPTN | Human |
| anti_7195_119 | Ephrin type-A receptor 7 | EPHA7 | Human |
| anti_7195_12 | Ephrin type-A receptor 7 | EPHA7 | Human |
| anti_7198_197 | Glycosaminoglycan xylosylkinase | FAM20B | Human |
| anti_7203_125 | Beta-1,3-N-acetylglucosaminyltransferase radical fringe | RFNG | Human |
| anti_7206_20 | Fructose-1,6-bisphosphatase 1 | FBP1 | Human |
| anti_7210_25 | Amyloid-like protein 1 | APLP1 | Human |
| anti_7215_18 | NADH-cytochrome b5 reductase 3 | CYB5R3 | Human |
| anti_7218_87 | Sodium/potassium-transporting ATPase subunit beta-2 | ATP1B2 | Human |
| anti_7223_60 | Protein S100-A13 | S100A13 | Human |
| anti_7227_75 | Cochlin | COCH | Human |
| anti_7241_12 | T-cell surface glycoprotein CD3 zeta chain | CD247 | Human |
| anti_7249_307 | Bcl-2-like protein 10 | BCL2L10 | Human |
| anti_7251_64 | Complement C1q tumor necrosis factor-related protein 3 | C1QTNF3 | Human |
| anti_7266_4 | Serpin A9 | SERPINA9 | Human |
| anti_7625_27 | 14-3-3 protein theta | YWHAQ | Human |
| anti_7628_40 | Cysteine-rich with EGF-like domain protein 1 | CRELD1 | Human |
| anti_7648_9 | Myosin-binding protein C, slow-type | MYBPC1 | Human |
| anti_7660_21 | Tropomyosin alpha-4 chain | TPM4 | Human |

| SOMAmer | Target Full Name | Entrez Gene Symbol | Organism |
|---|---|---|---|
| anti_7661_51 | 14-3-3 protein sigma_MOUSE | Sfn | Mouse |
| anti_7669_6 | Growth-regulated alpha protein_MOUSE | Cxcl1 | Mouse |
| anti_7670_4 | Insulin-like growth factor-binding protein 3_MOUSE | Igfbp3 | Mouse |
| anti_7672_7 | Pro-interleukin-16_MOUSE | Il16 | Mouse |
| anti_7676_7 | Galectin-4_MOUSE | Lgals4 | Mouse |
| anti_7695_106 | Sequestosome-1 | SQSTM1 | Human |
| anti_7696_104 | Next to BRCA1 gene 1 protein | NBR1 | Human |
| anti_7717_95 | Cystic fibrosis transmembrane conductance regulator | CFTR | Human |
| anti_7721_81 | Fragile X mental retardation protein 1 | FMR1 | Human |
| anti_7747_47 | NADH dehydrogenase [ubiquinone] 1 beta subcomplex subunit 11, mitochondrial | NDUFB11 | Human |
| anti_7751_121 | Secreted frizzled-related protein 2 | SFRP2 | Human |
| anti_7757_5 | HLA class II histocompatibility antigen, DQ alpha 2 chain | HLA-DQA2 | Human |
| anti_7758_217 | RING finger protein 215 | RNF215 | Human |
| anti_7766_25 | Zona pellucida sperm-binding protein 4 | ZP4 | Human |
| anti_7768_10 | Carboxypeptidase M | CPM | Human |
| anti_7773_20 | Killer cell immunoglobulin-like receptor 2DL2 | KIR2DL2 | Human |
| anti_7776_20 | Netrin receptor UNC5B | UNC5B | Human |
| anti_7779_86 | Carbohydrate sulfotransferase 11 | CHST11 | Human |
| anti_7785_1 | Ephrin-B3 | EFNB3 | Human |
| anti_7787_25 | Leukocyte immunoglobulin-like receptor subfamily A member 5 | LILRA5 | Human |
| anti_7796_10 | B-cell antigen receptor complex-associated protein alpha chain | CD79A | Human |
| anti_7806_33 | Beta-1,4-galactosyltransferase 7 | B4GALT7 | Human |
| anti_7808_5 | D-glucuronyl C5-epimerase | GLCE | Human |
| anti_7809_22 | Spermatogenesis-associated protein 9 | SPATA9 | Human |
| anti_7810_20 | Complement C1q tumor necrosis factor-related protein 5 | C1QTNF5 | Human |
| anti_7818_101 | Guanylate-binding protein 6 | GBP6 | Human |
| anti_7823_22 | Alpha-N-acetylgalactosaminide alpha-2,6-sialyltransferase 2 | ST6GALNAC2 | Human |
| anti_7832_181 | Protein disulfide-isomerase A3 | PDIA3 | Human |
| anti_7839_99 | Protein FAM19A3 | TAFA3 | Human |
| anti_7846_44 | Pregnancy-specific beta-1-glycoprotein 11 | PSG11 | Human |
| anti_7850_1 | Cytochrome c oxidase subunit 4 isoform 2, mitochondrial | COX4I2 | Human |
| anti_7853_19 | Protein SCO1 homolog, mitochondrial | SCO1 | Human |
| anti_7856_51 | Protein FAM151A | FAM151A | Human |
| anti_7857_22 | Neurotensin/neuromedin N | NTS | Human |
| anti_7859_21 | Protocadherin gamma-C3 | PCDHGC3 | Human |
| anti_7860_9 | Probable E3 ubiquitin-protein ligase HERC4 | HERC4 | Human |
| anti_7861_9 | Tyrosine-protein kinase transmembrane receptor ROR2 | ROR2 | Human |
| anti_7863_50 | Membrane-associated progesterone receptor component 1 | PGRMC1 | Human |
| anti_7866_11 | DnaJ homolog subfamily C member 30 | DNAJC30 | Human |
| anti_7870_8 | Beta-Ala-His dipeptidase | CNDP1 | Human |
| anti_7871_16 | Transmembrane protein 132A | TMEM132A | Human |
| anti_7875_86 | Pleckstrin | PLEK | Human |
| anti_7880_9 | Proteasome subunit alpha type-1 | PSMA1 | Human |
| anti_7881_244 | Mono [ADP-ribose] polymerase PARP16 | PARP16 | Human |
| anti_7882_31 | SLAM family member 7 | SLAMF7 | Human |
| anti_7884_15 | Discoidin domain-containing receptor 2 | DDR2 | Human |
| anti_7886_26 | Serine palmitoyltransferase 1 | SPTLC1 | Human |
| anti_7893_19 | Oxidized low-density lipoprotein receptor 1 | OLR1 | Human |
| anti_7895_108 | Melanocortin-2 receptor accessory protein | MRAP | Human |
| anti_7910_41 | Leucine-rich repeat and fibronectin type III domain-containing protein 1 | LRFN1 | Human |
| anti_7933_75 | Disintegrin and metalloproteinase domain-containing protein 22 | ADAM22 | Human |
| anti_7934_11 | Protein NDRG4 | NDRG4 | Human |

| SOMAmer | Target Full Name | Entrez Gene Symbol | Organism |
|---|---|---|---|
| anti_7945_10 | Semaphorin-6A | SEMA6A | Human |
| anti_7948_129 | Glycolipid transfer protein domain-containing protein 2 | GLTPD2 | Human |
| anti_7956_11 | MICOS complex subunit MIC10 | MICOS10 | Human |
| anti_7957_2 | Secretogranin-3 | SCG3 | Human |
| anti_7959_34 | Cadherin-7 | CDH7 | Human |
| anti_7962_11 | Serine protease inhibitor Kazal-type 8 | SPINK8 | Human |
| anti_7965_25 | Hemoglobin subunit theta-1 | HBQ1 | Human |
| anti_7969_163 | Protein WFDC11 | WFDC11 | Human |
| anti_7970_315 | Ecto-ADP-ribosyltransferase 3 | ART3 | Human |
| anti_7980_72 | N-acetyllactosaminide beta-1,3-N-acetylglucosaminyltransferase 2 | B3GNT2 | Human |
| anti_8007_19 | Cathepsin B | CTSB | Human |
| anti_8009_121 | Surfeit locus protein 1 | SURF1 | Human |
| anti_8014_359 | Glycoprotein endo-alpha-1,2-mannosidase | MANEA | Human |
| anti_8017_23 | FAD-dependent oxidoreductase domain-containing protein 1 | FOXRED1 | Human |
| anti_8024_64 | Protein-tyrosine sulfotransferase 2 | TPST2 | Human |
| anti_8028_22 | Serine protease inhibitor Kazal-type 5 | SPINK5 | Human |
| anti_8037_53 | DnaJ homolog subfamily B member 14 | DNAJB14 | Human |
| anti_8038_41 | Sterol-4-alpha-carboxylate 3-dehydrogenase, decarboxylating | NSDHL | Human |
| anti_8039_41 | Protein FAM177A1 | FAM177A1 | Human |
| anti_8040_9 | LEM domain-containing protein 1 | LEMD1 | Human |
| anti_8044_90 | Glycoprotein Xg | XG | Human |
| anti_8046_9 | Galectin-1 | LGALS1 | Human |
| anti_8048_9 | Ferritin, mitochondrial | FTMT | Human |
| anti_8074_32 | Transmembrane protein 70, mitochondrial | TMEM70 | Human |
| anti_8076_4 | Sperm acrosome membrane-associated protein 3 | SPACA3 | Human |
| anti_8080_24 | Prostate-associated microseminoprotein | MSMP | Human |
| anti_8086_49 | Integral membrane protein 2B | ITM2B | Human |
| anti_8092_29 | Interleukin-6 receptor subunit alpha | IL6R | Human |
| anti_8097_77 | Lipase member N | LIPN | Human |
| anti_8098_37 | Thromboxane-A synthase | TBXAS1 | Human |
| anti_8099_42 | Spondin-2 | SPON2 | Human |
| anti_8219_14 | Zymogen granule membrane protein 16 | ZG16 | Human |
| anti_8220_15 | Uncharacterized protein C10orf35 | FAM241B | Human |
| anti_8225_86 | Ephrin type-B receptor 2 | EPHB2 | Human |
| anti_8229_1 | Glucoside xylosyltransferase 1 | GXYLT1 | Human |
| anti_8232_90 | Junctional adhesion molecule-like | JAML | Human |
| anti_8233_2 | Inter-alpha-trypsin inhibitor heavy chain H5 | ITIH5 | Human |
| anti_8235_48 | Secretogranin-1 | CHGB | Human |
| anti_8240_207 | Transmembrane glycoprotein NMB | GPNMB | Human |
| anti_8244_16 | Alpha-(1,6)-fucosyltransferase | FUT8 | Human |
| anti_8245_27 | Intercellular adhesion molecule 5 | ICAM5 | Human |
| anti_8246_9 | Proprotein convertase subtilisin/kexin type 9 | PCSK9 | Human |
| anti_8248_222 | Sialic acid-binding Ig-like lectin 14 | SIGLEC14 | Human |
| anti_8250_2 | Receptor-type tyrosine-protein phosphatase eta | PTPRJ | Human |
| anti_8252_2 | Palmitoleoyl-protein carboxylesterase NOTUM | NOTUM | Human |
| anti_8258_22 | UDP-glucuronic acid decarboxylase 1 | UXS1 | Human |
| anti_8264_43 | Delta-like protein 1 | DLL1 | Human |
| anti_8269_327 | Arylsulfatase K | ARSK | Human |
| anti_8273_84 | Interleukin-31 receptor subunit alpha | IL31RA | Human |
| anti_8274_64 | Syntaxin-7 | STX7 | Human |
| anti_8275_31 | Platelet endothelial aggregation receptor 1 | PEAR1 | Human |
| anti_8295_16 | Noelin-2 | OLFM2 | Human |
| anti_8299_66 | Leukocyte immunoglobulin-like receptor subfamily A member 4 | LILRA4 | Human |
| anti_8304_50 | Tumor necrosis factor receptor superfamily member 11B | TNFRSF11B | Human |
| anti_8305_18 | Extracellular sulfatase Sulf-2 | SULF2 | Human |
| anti_8309_12 | Hyaluronidase-1 | HYAL1 | Human |
| anti_8312_139 | UMP-CMP kinase | CMPK1 | Human |
| anti_8323_163 | Trefoil factor 3 | TFF3 | Human |
| anti_8325_37 | Alcohol dehydrogenase 4 | ADH4 | Human |

| SOMAmer | Target Full Name | Entrez Gene Symbol | Organism |
|---|---|---|---|
| anti_8327_26 | Dipeptidase 2 | DPEP2 | Human |
| anti_8330_1 | Low-density lipoprotein receptor-related protein 11 | LRP11 | Human |
| anti_8334_65 | T-complex protein 11 homolog | TCP11 | Human |
| anti_8352_26 | Sialic acid-binding Ig-like lectin 12 | SIGLEC12 | Human |
| anti_8368_102 | Tumor necrosis factor receptor superfamily member 1B | TNFRSF1B | Human |
| anti_8374_5 | Tumor necrosis factor receptor superfamily member 16 | NGFR | Human |
| anti_8378_3 | Protein LDOC1 | LDOC1 | Human |
| anti_8382_47 | MAP kinase-activated protein kinase 5 | MAPKAPK5 | Human |
| anti_8385_248 | Hepatocyte growth factor activator | HGFAC | Human |
| anti_8387_33 | 14-3-3 protein epsilon | YWHAE | Human |
| anti_8389_8 | Leucine-rich repeat-containing protein 74A | LRRC74A | Human |
| anti_8397_147 | Sulfhydryl oxidase 2 | QSOX2 | Human |
| anti_8398_277 | Retinoic acid receptor responder protein 1 | RARRES1 | Human |
| anti_8402_22 | Cytokine-like protein 1 | CYTL1 | Human |
| anti_8405_108 | Disintegrin and metalloproteinase domain-containing protein 10 | ADAM10 | Human |
| anti_8407_84 | Neurogenic locus notch homolog protein 2 | NOTCH2 | Human |
| anti_8428_102 | Neurotrimin | NTM | Human |
| anti_8429_16 | Axin-2 | AXIN2 | Human |
| anti_8458_111 | Alpha-synuclein | SNCA | Human |
| anti_8459_10 | Bone morphogenetic protein 6 | BMP6 | Human |
| anti_8460_2 | Interleukin-11_MOUSE | Il11 | Mouse |
| anti_8471_53 | Allophycocyanin_CYACA | apcB\|apcA | Red alga\|Red alga |
| anti_8474_6 | Inactive tyrosine-protein kinase transmembrane receptor ROR1 | ROR1 | Human |
| anti_8476_11 | Chromogranin-A | CHGA | Human |
| anti_8484_24 | Leptin | LEP | Human |
| anti_8484_8 | Leptin | LEP | Human |
| anti_8485_7 | Kelch-like ECH-associated protein 1 | KEAP1 | Human |
| anti_8518_55 | Fc_MOUSE | Igh | Mouse |
| anti_8528_74 | Immunoglobulin superfamily containing leucine-rich repeat protein 2 | ISLR2 | Human |
| anti_8535_102 | Dermokine | DMKN | Human |
| anti_8565_160 | Plasmalemma vesicle-associated protein | PLVAP | Human |
| anti_8578_45 | Fc_MOUSE | Igh | Mouse |
| anti_8589_13 | CUB domain-containing protein 1 | CDCP1 | Human |
| anti_8597_1 | Transmembrane protein 9 | TMEM9 | Human |
| anti_8620_56 | Kallikrein-14 | KLK14 | Human |
| anti_8634_187 | Fc_MOUSE | Igh | Mouse |
| anti_8644_46 | Cathepsin H | CTSH | Human |
| anti_8660_33 | Olfactomedin-like protein 3 | OLFML3 | Human |
| anti_8660_5 | Olfactomedin-like protein 3 | OLFML3 | Human |
| anti_8667_42 | Fc_MOUSE | Igh | Mouse |
| anti_8683_119 | Paired immunoglobulin-like type 2 receptor alpha | PILRA | Human |
| anti_8696_15 | Galactosylceramide sulfotransferase | GAL3ST1 | Human |
| anti_8697_38 | Glypican-1 | GPC1 | Human |
| anti_8755_202 | Fc_MOUSE | Igh | Mouse |
| anti_8758_2 | Protocadherin alpha-7 | PCDHA7 | Human |
| anti_8760_10 | DCN1-like protein 5 | DCUN1D5 | Human |
| anti_8768_4 | B-cell lymphoma/leukemia 10 | BCL10 | Human |
| anti_8780_2 | Protocadherin-10 | PCDH10 | Human |
| anti_8791_151 | Carbonic anhydrase 5A, mitochondrial | CA5A | Human |
| anti_8795_48 | Transferrin receptor protein 1 | TFRC | Human |
| anti_8814_33 | Proactivator polypeptide-like 1 | PSAPL1 | Human |
| anti_8819_3 | Insulin-like growth factor-binding protein 2 | IGFBP2 | Human |
| anti_8837_8 | Microfibrillar-associated protein 3-like | MFAP3L | Human |
| anti_8866_53 | Glutaminyl-peptide cyclotransferase-like protein | QPCTL | Human |
| anti_8868_4 | Fc_MOUSE | Igh | Mouse |
| anti_8882_1 | Disintegrin and metalloproteinase domain-containing protein 17 | ADAM17 | Human |
| anti_8887_21 | Nuclear migration protein nudC | NUDC | Human |
| anti_8888_33 | Small integral membrane protein 9 | SMIM9 | Human |

| SOMAmer | Target Full Name | Entrez Gene Symbol | Organism |
|---|---|---|---|
| anti_8890_9 | Transmembrane protein 132B | TMEM132B | Human |
| anti_8892_14 | Platelet endothelial aggregation receptor 1 | PEAR1 | Human |
| anti_8900_28 | Neogenin | NEO1 | Human |
| anti_8901_40 | 14-3-3 protein gamma | YWHAG | Human |
| anti_8903_1 | Cytochrome c oxidase subunit 6C | COX6C | Human |
| anti_8906_60 | Leucine-rich repeat and transmembrane domain-containing protein 2 | LRTM2 | Human |
| anti_8907_11 | UDP-glucuronosyltransferase 2A1 | UGT2A1 | Human |
| anti_8908_14 | Potassium voltage-gated channel subfamily E regulatory beta subunit 5 | KCNE5 | Human |
| anti_8916_32 | Stromal interaction molecule 1 | STIM1 | Human |
| anti_8918_64 | Protein-tyrosine kinase 2-beta | PTK2B | Human |
| anti_8922_4 | Transmembrane and coiled-coil domains protein 3 | TMCC3 | Human |
| anti_8932_1 | Ectonucleoside triphosphate diphosphohydrolase 6 | ENTPD6 | Human |
| anti_8942_2 | 39S ribosomal protein L21, mitochondrial | MRPL21 | Human |
| anti_8950_4 | Calcium-activated chloride channel regulator 2 | CLCA2 | Human |
| anti_8952_65 | Granulocyte colony-stimulating factor | CSF3 | Human |
| anti_8953_47 | Hepatoma-derived growth factor | HDGF | Human |
| anti_8956_96 | Scavenger receptor class F member 2 | SCARF2 | Human |
| anti_8960_3 | Endoplasmic reticulum aminopeptidase 2 | ERAP2 | Human |
| anti_8967_6 | Dual specificity protein phosphatase 26 | DUSP26 | Human |
| anti_8971_9 | Neurexin-1 | NRXN1 | Human |
| anti_8974_172 | Collagen alpha-1(XV) chain | COL15A1 | Human |
| anti_8982_65 | Thrombospondin-3 | THBS3 | Human |
| anti_8983_7 | Golgi membrane protein 1 | GOLM1 | Human |
| anti_8990_42 | Sodium/potassium-transporting ATPase subunit beta-3 | ATP1B3 | Human |
| anti_8991_115 | Fc receptor-like protein 4 | FCRL4 | Human |
| anti_9000_177 | Neutral and basic amino acid transport protein rBAT | SLC3A1 | Human |
| anti_9003_99 | Macrophage receptor MARCO | MARCO | Human |
| anti_9005_16 | Plexin-A1 | PLXNA1 | Human |
| anti_9016_12 | Oxysterols receptor LXR-beta | NR1H2 | Human |
| anti_9023_9 | Fc_MOUSE | Igh | Mouse |
| anti_9025_5 | Alpha-hemoglobin-stabilizing protein | AHSP | Human |
| anti_9028_5 | Poly(A) RNA polymerase, mitochondrial | MTPAP | Human |
| anti_9037_1 | Syntaxin-18 | STX18 | Human |
| anti_9039_47 | Torsin-1A-interacting protein 1 | TOR1AIP1 | Human |
| anti_9044_1 | Contactin-associated protein-like 5 | CNTNAP5 | Human |
| anti_9045_3 | Very long-chain acyl-CoA synthetase | SLC27A2 | Human |
| anti_9053_16 | Cysteine-rich protein 2 | CRIP2 | Human |
| anti_9061_3 | Rho guanine nucleotide exchange factor 10 | ARHGEF10 | Human |
| anti_9063_9 | Fc_MOUSE | Igh | Mouse |
| anti_9068_17 | Janus kinase and microtubule-interacting protein 3 | JAKMIP3 | Human |
| anti_9076_25 | Proenkephalin-A | PENK | Human |
| anti_9077_10 | Mannosyl-oligosaccharide 1,2-alpha-mannosidase IB | MAN1A2 | Human |
| anti_9082_25 | Protein sel-1 homolog 2 | SEL1L2 | Human |
| anti_9087_8 | NHL repeat-containing protein 3 | NHLRC3 | Human |
| anti_9088_20 | Fc_MOUSE | Igh | Mouse |
| anti_9090_9 | Synaptotagmin-like protein 4 | SYTL4 | Human |
| anti_9092_33 | Angiopoietin-related protein 1 | ANGPTL1 | Human |
| anti_9099_19 | Synaptotagmin-5 | SYT5 | Human |
| anti_9100_32 | Uncharacterized protein C5orf46 | C5orf46 | Human |
| anti_9102_28 | Protrudin | ZFYVE27 | Human |
| anti_9118_7 | PH and SEC7 domain-containing protein 2 | PSD2 | Human |
| anti_9169_14 | Small ubiquitin-related modifier 3 | SUMO3 | Human |
| anti_9172_69 | Neutrophil collagenase | MMP8 | Human |
| anti_9177_6 | Protein FAM3B | FAM3B | Human |
| anti_9183_7 | Interferon alpha/beta receptor 1 | IFNAR1 | Human |
| anti_9187_2 | Non-histone chromosomal protein HMG-14 | HMGN1 | Human |
| anti_9189_76 | Appetite-regulating hormone | GHRL | Human |
| anti_9190_7 | CD63 antigen | CD63 | Human |
| anti_9191_8 | Trefoil factor 2 | TFF2 | Human |
| anti_9196_8 | Galectin-7 | LGALS7 | Human |

| SOMAmer | Target Full Name | Entrez Gene Symbol | Organism |
|---|---|---|---|
| anti_9199_6 | Ubiquitin-conjugating enzyme E2 G2 | UBE2G2 | Human |
| anti_9201_13 | Transgelin-2 | TAGLN2 | Human |
| anti_9207_60 | Quinone oxidoreductase-like protein 1 | CRYZL1 | Human |
| anti_9212_22 | Cathepsin F | CTSF | Human |
| anti_9216_100 | Plexin-B2 | PLXNB2 | Human |
| anti_9223_11 | Mesencephalic astrocyte-derived neurotrophic factor | MANF | Human |
| anti_9229_9 | Chymotrypsin-like protease CTRL-1 | CTRL | Human |
| anti_9231_23 | Inositol monophosphatase 3 | IMPAD1 | Human |
| anti_9232_1 | Roundabout homolog 4 | ROBO4 | Human |
| anti_9234_8 | Twisted gastrulation protein homolog 1 | TWSG1 | Human |
| anti_9235_3 | Plexin domain-containing protein 1 | PLXDC1 | Human |
| anti_9243_10 | Lysozyme g-like protein 1 | LYG1 | Human |
| anti_9244_27 | Palmitoyl-protein thioesterase 1 | PPT1 | Human |
| anti_9250_87 | Neutrophil defensin 1 | DEFA1 | Human |
| anti_9251_28 | Epididymis-specific alpha-mannosidase | MAN2B2 | Human |
| anti_9253_52 | Histo-blood group ABO system transferase | ABO | Human |
| anti_9256_78 | Neuronal pentraxin-1 | NPTX1 | Human |
| anti_9267_2 | Carboxypeptidase A4 | CPA4 | Human |
| anti_9271_101 | Stromal interaction molecule 1 | STIM1 | Human |
| anti_9275_2 | Sialic acid-binding Ig-like lectin 5 | SIGLEC5 | Human |
| anti_9276_7 | Carboxypeptidase A2 | CPA2 | Human |
| anti_9278_9 | Stromal cell-derived factor 1 | CXCL12 | Human |
| anti_9282_12 | Cysteine-rich secretory protein 2 | CRISP2 | Human |
| anti_9284_25 | Secreted frizzled-related protein 4 | SFRP4 | Human |
| anti_9294_45 | Microfibrillar-associated protein 2 | MFAP2 | Human |
| anti_9296_15 | Receptor-type tyrosine-protein phosphatase delta | PTPRD | Human |
| anti_9304_27 | Follistatin-related protein 1 | FSTL1 | Human |
| anti_9313_27 | Cerebellin-1 | CBLN1 | Human |
| anti_9315_16 | Mitochondrial ubiquitin ligase activator of NFKB 1 | MUL1 | Human |
| anti_9317_4 | Leucine-rich repeat-containing protein 25 | LRRC25 | Human |
| anti_9322_15 | Reticulocalbin-1 | RCN1 | Human |
| anti_9338_2 | Fc_MOUSE | Igh | Mouse |
| anti_9339_204 | Peptidyl-prolyl cis-trans isomerase FKBP2 | FKBP2 | Human |
| anti_9341_1 | Platelet-derived growth factor D | PDGFD | Human |
| anti_9357_4 | Protein CREG1 | CREG1 | Human |
| anti_9362_11 | Fc_MOUSE | Igh | Mouse |
| anti_9370_69 | Gamma-glutamyl hydrolase | GGH | Human |
| anti_9377_25 | Kit ligand | KITLG | Human |
| anti_9380_2 | Group XIIB secretory phospholipase A2-like protein | PLA2G12B | Human |
| anti_9384_17 | Cathelicidin antimicrobial peptide | CAMP | Human |
| anti_9385_4 | Lysosomal alpha-glucosidase | GAA | Human |
| anti_9388_18 | Methylmalonyl-CoA epimerase, mitochondrial | MCEE | Human |
| anti_9391_60 | ProSAAS | PCSK1N | Human |
| anti_9392_43 | Alpha-1,3-mannosyl-glycoprotein 4-beta-N-acetylglucosaminyltransferase A | MGAT4A | Human |
| anti_9394_19 | Carboxypeptidase Q | CPQ | Human |
| anti_9400_40 | Galectin-7 | LGALS7 | Human |
| anti_9401_57 | Metalloprotease TIKI1 | TRABD2A | Human |
| anti_9409_11 | Tryptase beta-1 | TPSAB1 | Human |
| anti_9416_77 | Carboxypeptidase M | CPM | Human |
| anti_9451_20 | Uromodulin | UMOD | Human |
| anti_9456_34 | Interleukin-22 receptor subunit alpha-2 | IL22RA2 | Human |
| anti_9459_7 | Tumor necrosis factor receptor superfamily member 6 | FAS | Human |
| anti_9461_2 | Intercellular adhesion molecule 4 | ICAM4 | Human |
| anti_9468_8 | Vesicular integral-membrane protein VIP36 | LMAN2 | Human |
| anti_9482_110 | ADP-ribose pyrophosphatase, mitochondrial | NUDT9 | Human |
| anti_9484_75 | Desmoglein-2 | DSG2 | Human |
| anti_9490_3 | Uncharacterized protein C4orf32 | FAM241A | Human |
| anti_9506_10 | Apolipoprotein L1 | APOL1 | Human |
| anti_9511_61 | Neurexophilin-2 | NXPH2 | Human |
| anti_9522_3 | Apoptosis-inducing factor 1, mitochondrial | AIFM1 | Human |

| SOMAmer | Target Full Name | Entrez Gene Symbol | Organism |
|---|---|---|---|
| anti_9525_1 | Inactive tyrosine-protein kinase 7 | PTK7 | Human |
| anti_9526_3 | Colipase-like protein 1 | CLPSL1 | Human |
| anti_9532_5 | WW domain binding protein 1-like | WBP1L | Human |
| anti_9535_14 | Fc_MOUSE | Igh | Mouse |
| anti_9536_16 | Epididymal secretory protein E3-alpha | EDDM3A | Human |
| anti_9557_5 | MANSC domain-containing protein 1 | MANSC1 | Human |
| anti_9560_56 | T-cell surface protein tactile | CD96 | Human |
| anti_9574_11 | Myc box-dependent-interacting protein 1 | BIN1 | Human |
| anti_9584_105 | Fc_MOUSE | Igh | Mouse |
| anti_9590_10 | Cytochrome c oxidase subunit 7A-related protein, mitochondrial | COX7A2L | Human |
| anti_9595_11 | Beta-1,4-galactosyltransferase 2 | B4GALT2 | Human |
| anti_9598_23 | CUB and sushi domain-containing protein 1 | CSMD1 | Human |
| anti_9599_6 | PILR alpha-associated neural protein | PIANP | Human |
| anti_9609_25 | Zinc finger protein 275 | ZNF275 | Human |
| anti_9627_15 | Vitrin | VIT | Human |
| anti_9713_67 | Platelet-derived growth factor receptor-like protein | PDGFRL | Human |
| anti_9719_145 | Matrix metalloproteinase-16 | MMP16 | Human |
| anti_9735_44 | T-cell surface protein tactile | CD96 | Human |
| anti_9738_7 | FAS-associated factor 2 | FAF2 | Human |
| anti_9742_59 | Spliceosome RNA helicase DDX39B | DDX39B | Human |
| anti_9744_139 | DnaJ homolog subfamily A member 4 | DNAJA4 | Human |
| anti_9745_20 | Eukaryotic translation initiation factor 4E type 2 | EIF4E2 | Human |
| anti_9749_190 | Plastin-2 | LCP1 | Human |
| anti_9753_17 | Ezrin | EZR | Human |
| anti_9755_19 | Drebrin-like protein | DBNL | Human |
| anti_9756_6 | Transgelin | TAGLN | Human |
| anti_9757_29 | Protein enabled homolog | ENAH | Human |
| anti_9761_89 | L-lactate dehydrogenase A chain | LDHA | Human |
| anti_9762_14 | Vasodilator-stimulated phosphoprotein | VASP | Human |
| anti_9764_79 | Heterogeneous nuclear ribonucleoprotein F | HNRNPF | Human |
| anti_9777_138 | Erythroid membrane-associated protein | ERMAP | Human |
| anti_9778_45 | Transmembrane protein 52B | TMEM52B | Human |
| anti_9789_52 | Nesprin-2 | SYNE2 | Human |
| anti_9793_145 | Immunoglobulin superfamily DCC subclass member 4 | IGDCC4 | Human |
| anti_9796_4 | Bile salt-activated lipase | CEL | Human |
| anti_9799_3 | Fc_MOUSE | Igh | Mouse |
| anti_9816_37 | Isochorismatase domain-containing protein 1 | ISOC1 | Human |
| anti_9823_2 | Dihydrofolate reductase | DHFR | Human |
| anti_9830_109 | Guanine nucleotide exchange factor VAV3 | VAV3 | Human |
| anti_9835_16 | Aldehyde dehydrogenase family 1 member A3 | ALDH1A3 | Human |
| anti_9836_20 | Deoxycytidine kinase | DCK | Human |
| anti_9837_60 | NAD(P)H dehydrogenase [quinone] 1 | NQO1 | Human |
| anti_9838_4 | Mothers against decapentaplegic homolog 1 | SMAD1 | Human |
| anti_9840_2 | Cell division control protein 42 homolog | CDC42 | Human |
| anti_9843_5 | Alpha-actinin-1 | ACTN1 | Human |
| anti_9846_32 | Rho GDP-dissociation inhibitor 2 | ARHGDIB | Human |
| anti_9849_13 | DNA-(apurinic or apyrimidinic site) lyase | APEX1 | Human |
| anti_9851_9 | Fatty acid-binding protein, adipocyte | FABP4 | Human |
| anti_9853_3 | Interferon-induced protein with tetratricopeptide repeats 2 | IFIT2 | Human |
| anti_9855_10 | Transforming protein RhoA | RHOA | Human |
| anti_9869_28 | Nuclear factor NF-kappa-B p105 subunit | NFKB1 | Human |
| anti_9870_17 | Tryptophan--tRNA ligase, cytoplasmic | WARS1 | Human |
| anti_9874_28 | Cyclin-dependent kinase 4 inhibitor B | CDKN2B | Human |
| anti_9876_20 | Fructose-bisphosphate aldolase C | ALDOC | Human |
| anti_9877_28 | Crk-like protein | CRKL | Human |
| anti_9878_3 | Estrogen sulfotransferase | SULT1E1 | Human |
| anti_9880_33 | Tryptophan 2,3-dioxygenase | TDO2 | Human |
| anti_9883_29 | Lactoylglutathione lyase | GLO1 | Human |
| anti_9887_40 | E3 ubiquitin-protein ligase RBBP6 | RBBP6 | Human |
| anti_9906_21 | Testican-3 | SPOCK3 | Human |
| anti_9910_9 | S-methyl-5'-thioadenosine phosphorylase | MTAP | Human |
| anti_9916_146 | Leucine-rich repeat-containing protein 4B | LRRC4B | Human |

| SOMAmer | Target Full Name | Entrez Gene Symbol | Organism |
|---|---|---|---|
| anti_9917_16 | Fc_MOUSE | Igh | Mouse |
| anti_9918_23 | Calbindin | CALB1 | Human |
| anti_9926_4 | Protein S100-P | S100P | Human |
| anti_9927_96 | Tensin-4 | TNS4 | Human |
| anti_9928_125 | Leukocyte cell-derived chemotaxin 1 | CNMD | Human |
| anti_9931_20 | Keratin, type II cytoskeletal 1 | KRT1 | Human |
| anti_9934_29 | Phosphatidylinositol transfer protein alpha isoform | PITPNA | Human |
| anti_9945_8 | Armadillo repeat-containing protein 5 | ARMC5 | Human |
| anti_9950_229 | Lymphocyte activation gene 3 protein | LAG3 | Human |
| anti_9959_60 | Transmembrane protein 59-like | TMEM59L | Human |
| anti_9962_1 | Cadherin-related family member 5 | CDHR5 | Human |
| anti_9963_19 | Protocadherin beta-10 | PCDHB10 | Human |
| anti_9964_10 | Myocardial zonula adherens protein | MYZAP | Human |
| anti_9970_7 | UBX domain-containing protein 4 | UBXN4 | Human |
| anti_9971_5 | CUB and sushi domain-containing protein 2 | CSMD2 | Human |
| anti_9979_13 | Amphoterin-induced protein 1 | AMIGO1 | Human |
| anti_9991_112 | EMILIN-3 | EMILIN3 | Human |
| anti_9994_217 | Potassium-transporting ATPase subunit beta | ATP4B | Human |
| anti_9999_1 | Interferon regulatory factor 6 | IRF6 | Human |
| anti_10000_28 | Beta-crystallin B2 | CRYBB2 | Human |
| anti_10001_7 | RAF proto-oncogene serine/threonine-protein kinase | RAF1 | Human |
| anti_10003_15 | Zinc finger protein 41 | ZNF41 | Human |
| anti_10006_25 | ETS domain-containing protein Elk-1 | ELK1 | Human |
| anti_10008_43 | Guanylyl cyclase-activating protein 1 | GUCA1A | Human |
| anti_10009_2 | Interferon regulatory factor 1 | IRF1 | Human |
| anti_10010_10 | Beclin-1 | BECN1 | Human |
| anti_10012_5 | SAM pointed domain-containing Ets transcription factor | SPDEF | Human |
| anti_10013_34 | Fc_MOUSE | Igh | Mouse |
| anti_10014_31 | Zinc finger protein SNAI2 | SNAI2 | Human |
| anti_10015_119 | Voltage-gated potassium channel subunit beta-2 | KCNAB2 | Human |
| anti_10021_1 | Fc_MOUSE | Igh | Mouse |
| anti_10022_207 | DNA polymerase eta | POLH | Human |
| anti_10023_32 | Vitamin D3 receptor | VDR | Human |
| anti_10024_44 | 4-hydroxy-2-oxoglutarate aldolase, mitochondrial | HOGA1 | Human |
| anti_10030_8 | Adenine DNA glycosylase | MUTYH | Human |
| anti_10035_6 | Dual specificity protein phosphatase 4 | DUSP4 | Human |
| anti_10037_98 | Sialic acid-binding Ig-like lectin 12 | SIGLEC12 | Human |
| anti_10040_63 | Tumor protein 63 | TP63 | Human |
| anti_10042_8 | Serine/threonine-protein kinase Sgk3 | SGK3 | Human |
| anti_10043_31 | Bromodomain-containing protein 4 | BRD4 | Human |
| anti_10044_12 | Protein Wnt-10a | WNT10A | Human |
| anti_10045_47 | Cullin-3 | CUL3 | Human |
| anti_10049_112 | Telomeric repeat-binding factor 1 | TERF1 | Human |
| anti_10053_5 | Integrin-linked protein kinase | ILK | Human |
| anti_10054_3 | Gigaxonin | GAN | Human |
| anti_10056_5 | Forkhead box protein M1 | FOXM1 | Human |
| anti_10063_10 | E3 ubiquitin-protein ligase FANCL | FANCL | Human |
| anti_10069_2 | Peptidyl-prolyl cis-trans isomerase NIMA-interacting 1 | PIN1 | Human |
| anti_10070_22 | 1-phosphatidylinositol 4,5-bisphosphate phosphodiesterase gamma-2 | PLCG2 | Human |
| anti_10073_22 | Tyrosyl-DNA phosphodiesterase 1 | TDP1 | Human |
| anti_10074_128 | Bromodomain-containing protein 2 | BRD2 | Human |
| anti_10075_75 | Acyl-CoA-binding domain-containing protein 6 | ACBD6 | Human |
| anti_10076_1 | AP-4 complex subunit mu-1 | AP4M1 | Human |
| anti_10078_5 | BAG family molecular chaperone regulator 3 | BAG3 | Human |
| anti_10080_9 | Translation initiation factor eIF-2B subunit alpha | EIF2B1 | Human |
| anti_10081_17 | DNA-binding protein SATB2 | SATB2 | Human |
| anti_10087_10 | Alpha-crystallin A chain | CRYAA | Human |
| anti_10088_37 | Adenine phosphoribosyltransferase | APRT | Human |
| anti_10089_7 | N-acetylserotonin O-methyltransferase-like protein | ASMTL | Human |
| anti_10336_3 | E3 ubiquitin-protein ligase CHIP | STUB1 | Human |

| SOMAmer | Target Full Name | Entrez Gene Symbol | Organism |
|---|---|---|---|
| anti_10337_83 | CCAAT/enhancer-binding protein beta | CEBPB | Human |
| anti_10342_55 | E3 SUMO-protein ligase PIAS4 | PIAS4 | Human |
| anti_10356_21 | Transcription factor AP-1 | JUN | Human |
| anti_10358_33 | Induced myeloid leukemia cell differentiation protein Mcl-1 | MCL1 | Human |
| anti_10361_25 | 2'-5'-oligoadenylate synthase 1 | OAS1 | Human |
| anti_10362_35 | Myc proto-oncogene protein | MYC | Human |
| anti_10364_6 | Mothers against decapentaplegic homolog 2 | SMAD2 | Human |
| anti_10365_132 | Interleukin-23 | IL12B\|IL23A | Human |
| anti_10366_11 | Platelet-derived growth factor receptor alpha | PDGFRA | Human |
| anti_10367_26 | Interleukin-12 | IL12A\|IL12B | Human |
| anti_10367_62 | Interleukin-12 | IL12A\|IL12B | Human |
| anti_10370_21 | Signal transducer and activator of transcription 1-alpha/beta | STAT1 | Human |
| anti_10373_253 | Cerebral dopamine neurotrophic factor_MOUSE | Cdnf | Mouse |
| anti_10375_4 | Alpha-2-macroglobulin receptor-associated protein_MOUSE | Lrpap1 | Mouse |
| anti_10376_28 | T-cell surface glycoprotein CD4_RAT | Cd4 | Rat |
| anti_10390_21 | E3 ubiquitin-protein ligase ZNRF3 | ZNRF3 | Human |
| anti_10396_6 | Induced myeloid leukemia cell differentiation protein Mcl-1 | MCL1 | Human |
| anti_10398_110 | Ankyrin repeat domain-containing protein 1 | ANKRD1 | Human |
| anti_10416_79 | Fc_MOUSE | Igh | Mouse |
| anti_10420_30 | Fas apoptotic inhibitory molecule 1 | FAIM | Human |
| anti_10422_44 | Thioredoxin | TXN | Human |
| anti_10424_31 | Neural proliferation differentiation and control protein 1 | NPDC1 | Human |
| anti_10425_3 | Beta-1,4-galactosyltransferase 5 | B4GALT5 | Human |
| anti_10426_21 | Golgi SNAP receptor complex member 2 | GOSR2 | Human |
| anti_10427_2 | Fc_MOUSE | Igh | Mouse |
| anti_10430_31 | 4F2 cell-surface antigen heavy chain | SLC3A2 | Human |
| anti_10432_3 | Uncharacterized protein KIAA1644 | SHISAL1 | Human |
| anti_10435_2 | N(4)-(beta-N-acetylglucosaminyl)-L-asparaginase | AGA | Human |
| anti_10439_57 | Alpha-amylase 2B | AMY2B | Human |
| anti_10440_26 | CXADR-like membrane protein | CLMP | Human |
| anti_10442_1 | Transmembrane protein 190 | TMEM190 | Human |
| anti_10447_18 | Polyadenylate-binding protein 3 | PABPC3 | Human |
| anti_10449_31 | Protein shisa-2 homolog | SHISA2 | Human |
| anti_10450_3 | Interferon alpha-17 | IFNA17 | Human |
| anti_10452_24 | Fc_MOUSE | Igh | Mouse |
| anti_10453_7 | Carcinoembryonic antigen-related cell adhesion molecule 20 | CEACAM20 | Human |
| anti_10454_99 | DnaJ homolog subfamily C member 16 | DNAJC16 | Human |
| anti_10455_196 | Interleukin-31 | IL31 | Human |
| anti_10457_3 | Interleukin-18 receptor accessory protein | IL18RAP | Human |
| anti_10460_1 | Chitotriosidase-1 | CHIT1 | Human |
| anti_10461_57 | Fc_MOUSE | Igh | Mouse |
| anti_10462_14 | Insulin-like peptide INSL5 | INSL5 | Human |
| anti_10463_23 | Intestinal-type alkaline phosphatase | ALPI | Human |
| anti_10467_58 | Fc_MOUSE | Igh | Mouse |
| anti_10470_34 | Thioredoxin domain-containing protein 11 | TXNDC11 | Human |
| anti_10471_25 | Fc_MOUSE | Igh | Mouse |
| anti_10472_53 | Tumor necrosis factor receptor superfamily member 14 | TNFRSF14 | Human |
| anti_10473_2 | Trefoil factor 2 | TFF2 | Human |
| anti_10477_162 | Fc_MOUSE | Igh | Mouse |
| anti_10479_18 | Stromelysin-2 | MMP10 | Human |
| anti_10485_56 | Melanoma-associated antigen 4 | MAGEA4 | Human |
| anti_10489_19 | Fc_MOUSE | Igh | Mouse |
| anti_10490_3 | Dolichyl-diphosphooligosaccharide--protein glycosyltransferase subunit 1 | RPN1 | Human |
| anti_10491_21 | Fc_MOUSE | Igh | Mouse |
| anti_10494_48 | Fc_MOUSE | Igh | Mouse |
| anti_10495_10 | Fc_MOUSE | Igh | Mouse |
| anti_10497_242 | Fc_MOUSE | Igh | Mouse |

| SOMAmer | Target Full Name | Entrez Gene Symbol | Organism |
|---|---|---|---|
| anti_10499_1 | Transmembrane protein 106A | TMEM106A | Human |
| anti_10501_3 | Fc_MOUSE | Igh | Mouse |
| anti_10502_15 | Fc_MOUSE | Igh | Mouse |
| anti_10503_55 | Fc_MOUSE | Igh | Mouse |
| anti_10504_6 | Fc_MOUSE | Igh | Mouse |
| anti_10505_12 | E3 ubiquitin-protein ligase RNF43 | RNF43 | Human |
| anti_10506_53 | Membrane-associated progesterone receptor component 2 | PGRMC2 | Human |
| anti_10507_166 | Acrosomal protein SP-10 | ACRV1 | Human |
| anti_10510_62 | SLP adapter and CSK-interacting membrane protein | SCIMP | Human |
| anti_10512_13 | Cytokine receptor common subunit beta | CSF2RB | Human |
| anti_10513_13 | Calsenilin | KCNIP3 | Human |
| anti_10516_53 | Fc_MOUSE | Igh | Mouse |
| anti_10518_14 | Fc_MOUSE | Igh | Mouse |
| anti_10527_22 | ATP-dependent RNA helicase A | DHX9 | Human |
| anti_10528_2 | Dipeptidase 1 | DPEP1 | Human |
| anti_10529_19 | Fc_MOUSE | Igh | Mouse |
| anti_10530_8 | Proteasome subunit beta type-6 | PSMB6 | Human |
| anti_10533_1 | Fc_MOUSE | Igh | Mouse |
| anti_10534_40 | Poly [ADP-ribose] polymerase 1 | PARP1 | Human |
| anti_10535_25 | Cytochrome c oxidase subunit 6A2, mitochondrial | COX6A2 | Human |
| anti_10539_30 | CD99 antigen-like protein 2 | CD99L2 | Human |
| anti_10546_2 | Thrombospondin-type laminin G domain and EAR repeat-containing protein | TSPEAR | Human |
| anti_10547_42 | Fc_MOUSE | Igh | Mouse |
| anti_10548_35 | Fc_MOUSE | Igh | Mouse |
| anti_10550_37 | Bone morphogenetic protein receptor type-1B | BMPR1B | Human |
| anti_10552_88 | NKG2-F type II integral membrane protein | KLRC4 | Human |
| anti_10554_23 | Beta-galactosidase | GLB1 | Human |
| anti_10558_26 | Protocadherin-9 | PCDH9 | Human |
| anti_10560_1 | Integral membrane protein 2C | ITM2C | Human |
| anti_10561_5 | Peptidoglycan recognition protein 3 | PGLYRP3 | Human |
| anti_10562_42 | Neuropilin and tolloid-like protein 2 | NETO2 | Human |
| anti_10563_13 | LysM and putative peptidoglycan-binding domain-containing protein 3 | LYSMD3 | Human |
| anti_10571_14 | Alpha-1,3-mannosyl-glycoprotein 2-beta-N-acetylglucosaminyltransferase | MGAT1 | Human |
| anti_10574_10 | Beta-2-microglobulin | B2M | Human |
| anti_10582_36 | Protein O-mannose kinase | POMK | Human |
| anti_10583_1 | Transmembrane protein 108 | TMEM108 | Human |
| anti_10584_7 | NADH dehydrogenase [ubiquinone] iron-sulfur protein 4, mitochondrial | NDUFS4 | Human |
| anti_10588_39 | Lactosylceramide alpha-2,3-sialyltransferase | ST3GAL5 | Human |
| anti_10593_14 | Fc_MOUSE | Igh | Mouse |
| anti_10603_1 | Histatin-3 | HTN3 | Human |
| anti_10605_22 | Adipocyte plasma membrane-associated protein | APMAP | Human |
| anti_10607_4 | Fc_MOUSE | Igh | Mouse |
| anti_10608_9 | Histatin-1 | HTN1 | Human |
| anti_10610_8 | Fc_MOUSE | Igh | Mouse |
| anti_10613_33 | Protein CASC4 | CASC4 | Human |
| anti_10618_190 | Low-density lipoprotein receptor-related protein 2 | LRP2 | Human |
| anti_10621_26 | Protein Largen | PRR16 | Human |
| anti_10622_9 | Fc_MOUSE | Igh | Mouse |
| anti_10623_19 | Mucin-1 | MUC1 | Human |
| anti_10624_45 | Serine protease inhibitor Kazal-type 13 | SPINK13 | Human |
| anti_10630_5 | Oxidoreductase HTATIP2 | HTATIP2 | Human |
| anti_10631_9 | Membrane-associated progesterone receptor component 2 | PGRMC2 | Human |
| anti_10636_1 | Cell surface glycoprotein CD200 receptor 1 | CD200R1 | Human |
| anti_10638_1 | TERF1-interacting nuclear factor 2 | TINF2 | Human |
| anti_10640_9 | Fc_MOUSE | Igh | Mouse |
| anti_10655_43 | Fc_MOUSE | Igh | Mouse |
| anti_10658_28 | Fc_MOUSE | Igh | Mouse |
| anti_10659_49 | Fc_MOUSE | Igh | Mouse |

| SOMAmer | Target Full Name | Entrez Gene Symbol | Organism |
|---|---|---|---|
| anti_10660_33 | Fc_MOUSE | Igh | Mouse |
| anti_10661_4 | Fc_MOUSE | Igh | Mouse |
| anti_10663_42 | Insulin-induced gene 1 protein | INSIG1 | Human |
| anti_10665_30 | Fc_MOUSE | Igh | Mouse |
| anti_10667_78 | Uncharacterized protein C1orf185 | C1orf185 | Human |
| anti_10668_5 | Syntaxin-4 | STX4 | Human |
| anti_10670_26 | Semaphorin-3E | SEMA3E | Human |
| anti_10671_67 | Fc_MOUSE | Igh | Mouse |
| anti_10672_75 | Pulmonary surfactant-associated protein B | SFTPB | Human |
| anti_10675_223 | Thioredoxin-related transmembrane protein 2 | TMX2 | Human |
| anti_10689_5 | Beta-defensin 119 | DEFB119 | Human |
| anti_10693_43 | Killer cell lectin-like receptor subfamily G member 2 | KLRG2 | Human |
| anti_10695_12 | Protein WFDC10B | WFDC10B | Human |
| anti_10696_217 | Leucine-rich PPR motif-containing protein, mitochondrial | LRPPRC | Human |
| anti_10699_52 | Prolow-density lipoprotein receptor-related protein 1 | LRP1 | Human |
| anti_10700_10 | Immunoglobulin superfamily member 11 | IGSF11 | Human |
| anti_10703_203 | Sin3 histone deacetylase corepressor complex component SDS3 | SUDS3 | Human |
| anti_10704_91 | Lymphoid-restricted membrane protein | LRMP | Human |
| anti_10705_14 | Alpha-N-acetylgalactosaminide alpha-2,6-sialyltransferase 3 | ST6GALNAC3 | Human |
| anti_10708_3 | Progonadoliberin-2 | GNRH2 | Human |
| anti_10710_23 | Zona pellucida-binding protein 2 | ZPBP2 | Human |
| anti_10713_151 | CMRF35-like molecule 7 | CD300LB | Human |
| anti_10716_35 | 26S proteasome non-ATPase regulatory subunit 5 | PSMD5 | Human |
| anti_10724_45 | Fc_MOUSE | Igh | Mouse |
| anti_10734_339 | Galactose-3-O-sulfotransferase 2 | GAL3ST2 | Human |
| anti_10735_12 | 26S proteasome non-ATPase regulatory subunit 1 | PSMD1 | Human |
| anti_10738_11 | Fibulin-5 | FBLN5 | Human |
| anti_10741_22 | Macrophage colony-stimulating factor 1 | CSF1 | Human |
| anti_10746_24 | Dickkopf-related protein 3 | DKK 3 | Human |
| anti_10748_216 | Protocadherin beta-2 | PCDHB2 | Human |
| anti_10752_8 | Chromodomain Y-like protein 2 | CDYL2 | Human |
| anti_10754_113 | Prokineticin-2 | PROK2 | Human |
| anti_10756_34 | Urocortin-3 | UCN3 | Human |
| anti_10758_2 | Keratocan | KERA | Human |
| anti_10761_5 | Transmembrane emp24 domain-containing protein 2 | TMED2 | Human |
| anti_10762_2 | Killer cell lectin-like receptor subfamily G member 2 | KLRG2 | Human |
| anti_10772_21 | Chondroitin sulfate N-acetylgalactosaminyltransferase 2 | CSGALNACT2 | Human |
| anti_10780_10 | Ras-related protein Rab-3D | RAB3D | Human |
| anti_10785_8 | Leucine-rich repeat-containing protein 52 | LRRC52 | Human |
| anti_10791_21 | Fc_MOUSE | Igh | Mouse |
| anti_10800_15 | Serpin H1 | SERPINH1 | Human |
| anti_10803_22 | Heat shock 70 kDa protein 1A | HSPA1A | Human |
| anti_10809_14 | Killer cell lectin-like receptor subfamily B member 1 | KLRB1 | Human |
| anti_10814_7 | T-cell surface glycoprotein CD1a | CD1A | Human |
| anti_10815_2 | Intracellular hyaluronan-binding protein 4 | HABP4 | Human |
| anti_10816_150 | Paired immunoglobulin-like type 2 receptor alpha isoform FDF03-M14 | PILRA | Human |
| anti_10825_12 | V-set and immunoglobulin domain-containing protein 10 | VSIG10 | Human |
| anti_10827_67 | Leucine-rich repeat and immunoglobulin-like domain-containing nogo receptor-interacting protein 3 | LINGO3 | Human |
| anti_10830_5 | EF-hand calcium-binding domain-containing protein 14 | EFCAB14 | Human |
| anti_10832_24 | Beta-1,4-galactosyltransferase 6 | B4GALT6 | Human |
| anti_10847_1 | Sialic acid-binding Ig-like lectin 15 | SIGLEC15 | Human |

| SOMAmer | Target Full Name | Entrez Gene Symbol | Organism |
|---|---|---|---|
| anti_10848_137 | Butyrophilin-like protein 3 | BTNL3 | Human |
| anti_10852_114 | Heterogeneous nuclear ribonucleoprotein D-like | HNRNPDL | Human |
| anti_10872_103 | Scavenger receptor class A member 3 | SCARA3 | Human |
| anti_10878_1 | Tumor necrosis factor receptor superfamily member 10B | TNFRSF10B | Human |
| anti_10888_4 | Fc_MOUSE | Igh | Mouse |
| anti_10890_135 | Lactase-like protein | LCTL | Human |
| anti_10895_28 | Transmembrane protease serine 11B | TMPRSS11B | Human |
| anti_10900_272 | Stathmin-2 | STMN2 | Human |
| anti_10901_334 | Integral membrane protein 2C | ITM2C | Human |
| anti_10903_50 | Syntaxin-8 | STX8 | Human |
| anti_10908_2 | Polypeptide N-acetylgalactosaminyltransferase 13 | GALNT13 | Human |
| anti_10910_6 | Carcinoembryonic antigen-related cell adhesion molecule 4 | CEACAM4 | Human |
| anti_10914_4 | Fc_MOUSE | Igh | Mouse |
| anti_10917_40 | Guanine nucleotide-binding protein G(I)/G(S)/G(O) subunit gamma-T2 | GNGT2 | Human |
| anti_10924_258 | Neuferricin | CYB5D2 | Human |
| anti_10927_65 | Fc_MOUSE | Igh | Mouse |
| anti_10928_7 | Fc_MOUSE | Igh | Mouse |
| anti_10933_107 | Calcium-binding protein 8 | CALN1 | Human |
| anti_10938_13 | Lymphocyte function-associated antigen 3 | CD58 | Human |
| anti_10939_16 | Pleckstrin homology domain-containing family A member 4 | PLEKHA4 | Human |
| anti_10940_25 | Sarcalumenin | SRL | Human |
| anti_10943_36 | Fc_MOUSE | Igh | Mouse |
| anti_10948_14 | Phospholipase D3 | PLD3 | Human |
| anti_10949_59 | 60S acidic ribosomal protein P2 | RPLP2 | Human |
| anti_10953_14 | C-type lectin domain family 2 member A | CLEC2A | Human |
| anti_10955_4 | C-type lectin domain family 10 member A | CLEC10A | Human |
| anti_10959_125 | BET1-like protein | BET1L | Human |
| anti_10962_46 | Fc_MOUSE | Igh | Mouse |
| anti_10974_20 | Serine protease inhibitor Kazal-type 7 | SPINK7 | Human |
| anti_10975_59 | Mucin-like protein 1 | MUCL1 | Human |
| anti_10976_44 | Mucin-1 | MUC1 | Human |
| anti_10978_39 | Growth hormone variant | GH2 | Human |
| anti_10980_11 | Acetylcholinesterase | ACHE | Human |
| anti_10981_56 | Pro-neuregulin-3, membrane-bound isoform | NRG3 | Human |
| anti_11071_1 | Interleukin-5 | IL5 | Human |
| anti_11072_200 | Diacylglycerol kinase_ECOLI | dgkA | Escherichia coli (strain K12) |
| anti_11081_1 | Glycerol-3-phosphate dehydrogenase [NAD(+)], cytoplasmic | GPD1 | Human |
| anti_11083_23 | Gamma-enolase | ENO2 | Human |
| anti_11094_104 | Galectin-10 | CLC | Human |
| anti_11098_1 | Pyridoxal kinase | PDXK | Human |
| anti_11101_18 | Toll-like receptor 4 | TLR4 | Human |
| anti_11103_24 | Heat shock protein beta-1 | HSPB1 | Human |
| anti_11107_25 | Teneurin-3 | TENM3 | Human |
| anti_11108_16 | Filamin-A | FLNA | Human |
| anti_11110_4 | Transmembrane protein 119 | TMEM119 | Human |
| anti_11112_18 | Protein kish-A | TMEM167A | Human |
| anti_11117_2 | Spermatogenesis-associated protein 20 | SPATA20 | Human |
| anti_11122_97 | Maspardin | SPG21 | Human |
| anti_11124_9 | Filamin-A | FLNA | Human |
| anti_11126_102 | Triple functional domain protein | TRIO | Human |
| anti_11128_29 | Transmembrane protein 132C | TMEM132C | Human |
| anti_11129_66 | Bone morphogenetic protein 15 | BMP15 | Human |
| anti_11130_158 | Voltage-dependent L-type calcium channel subunit beta-4 | CACNB4 | Human |
| anti_11134_30 | Peptide chain release factor 1-like, mitochondrial | MTRF1L | Human |
| anti_11135_5 | 39S ribosomal protein L55, mitochondrial | MRPL55 | Human |
| anti_11137_43 | Cytokine receptor common subunit beta | CSF2RB | Human |
| anti_11138_16 | Runt-related transcription factor 3 | RUNX3 | Human |
| anti_11139_4 | Fc_MOUSE | Igh | Mouse |

| SOMAmer | Target Full Name | Entrez Gene Symbol | Organism |
|---|---|---|---|
| anti_11144_10 | Beta-defensin 116 | DEFB116 | Human |
| anti_11145_72 | UPF0606 protein KIAA1549L | KIAA1549L | Human |
| anti_11146_4 | T-box transcription factor TBX22 | TBX22 | Human |
| anti_11147_17 | Trem-like transcript 1 protein | TREML1 | Human |
| anti_11149_3 | Toll-like receptor 1 | TLR1 | Human |
| anti_11150_3 | Collagen alpha-1(VI) chain | COL6A1 | Human |
| anti_11154_3 | Nuclear factor erythroid 2-related factor 1 | NFE2L1 | Human |
| anti_11155_16 | Collagen alpha-5(VI) chain | COL6A5 | Human |
| anti_11158_40 | Bicaudal D-related protein 1 | BICDL1 | Human |
| anti_11159_14 | Mitochondrial ubiquitin ligase activator of NFKB 1 | MUL1 | Human |
| anti_11161_5 | Spartin | SPART | Human |
| anti_11162_37 | Equatorin | EQTN | Human |
| anti_11163_7 | Protein FAM162B | FAM162B | Human |
| anti_11164_7 | Teneurin-2 | TENM2 | Human |
| anti_11167_6 | Myotubularin-related protein 1 | MTMR1 | Human |
| anti_11168_3 | Multiple epidermal growth factor-like domains protein 10 | MEGF10 | Human |
| anti_11170_9 | Filamin-A | FLNA | Human |
| anti_11173_29 | Allergin-1 | MILR1 | Human |
| anti_11175_45 | Lysophosphatidylcholine acyltransferase 2 | LPCAT2 | Human |
| anti_11179_7 | Testis-specific serine/threonine-protein kinase 1 | TSSK1B | Human |
| anti_11180_17 | SWI/SNF complex subunit SMARCC1 | SMARCC1 | Human |
| anti_11185_145 | GTP cyclohydrolase 1 | GCH1 | Human |
| anti_11186_12 | Transmembrane protein 52 | TMEM52 | Human |
| anti_11187_11 | C-type lectin domain family 12 member A | CLEC12A | Human |
| anti_11190_129 | ATP-dependent zinc metalloprotease YME1L1 | YME1L1 | Human |
| anti_11192_168 | Tubulointerstitial nephritis antigen-like | TINAGL1 | Human |
| anti_11193_27 | Hepatocyte nuclear factor 1-alpha | HNF1A | Human |
| anti_11194_6 | Signaling threshold-regulating transmembrane adapter 1 | SIT 1 | Human |
| anti_11198_37 | Cyclic AMP-responsive element-binding protein 3-like protein 1 | CREB3L1 | Human |
| anti_11201_19 | tRNA pseudouridine synthase A, mitochondrial | PUS1 | Human |
| anti_11202_70 | T-box transcription factor TBX5 | TBX5 | Human |
| anti_11204_80 | Coxsackievirus and adenovirus receptor | CXADR | Human |
| anti_11207_3 | Macrophage scavenger receptor types I and II | MSR1 | Human |
| anti_11211_7 | Tubulin-specific chaperone E | TBCE | Human |
| anti_11217_16 | C-terminal-binding protein 1 | CTBP1 | Human |
| anti_11219_95 | Fibroblast growth factor-binding protein 3 | FGFBP3 | Human |
| anti_11220_53 | Uncharacterized protein C10orf105 | C10orf105 | Human |
| anti_11222_62 | Fc_MOUSE | Igh | Mouse |
| anti_11223_1 | Transmembrane protein 154 | TMEM154 | Human |
| anti_11226_16 | Ubiquitin-protein ligase E3A | UBE3A | Human |
| anti_11227_31 | Transducin beta-like protein 2 | TBL2 | Human |
| anti_11228_37 | Protein RIC-3 | RIC3 | Human |
| anti_11229_16 | Serine/threonine-protein kinase/endoribonuclease IRE1 | ERN 1 | Human |
| anti_11230_12 | Fc_MOUSE | Igh | Mouse |
| anti_11231_12 | Adrenodoxin-like protein, mitochondrial | FDX2 | Human |
| anti_11232_46 | Transcobalamin-1 | TCN1 | Human |
| anti_11239_49 | Transmembrane protease serine 6 | TMPRSS6 | Human |
| anti_11242_33 | Protein-glutamine gamma-glutamyltransferase K | TGM1 | Human |
| anti_11244_63 | Latent-transforming growth factor beta-binding protein 4 | LTBP4 | Human |
| anti_11247_20 | N-acetylglutamate synthase, mitochondrial | NAGS | Human |
| anti_11248_43 | Uroporphyrinogen-III synthase | UROS | Human |
| anti_11252_30 | Leucine-rich repeat and calponin homology domain-containing protein 4 | LRCH4 | Human |
| anti_11254_13 | Papilin | PAPLN | Human |
| anti_11259_71 | Protocadherin gamma-A8 | PCDHGA8 | Human |
| anti_11260_47 | SUN domain-containing protein 5 | SUN5 | Human |
| anti_11262_39 | TraB domain-containing protein | TRABD | Human |
| anti_11264_33 | Xanthine dehydrogenase/oxidase | XDH | Human |
| anti_11266_8 | P-selectin glycoprotein ligand 1 | SELPLG | Human |

| SOMAmer | Target Full Name | Entrez Gene Symbol | Organism |
|---|---|---|---|
| anti_11267_11 | Mucin-1 | MUC1 | Human |
| anti_11270_17 | Coiled-coil-helix-coiled-coil-helix domain-containing protein 10, mitochondrial | CHCHD10 | Human |
| anti_11273_176 | Glutathione S-transferase theta-2B | GSTT2B | Human |
| anti_11275_94 | Low-density lipoprotein receptor-related protein 1B | LRP1B | Human |
| anti_11276_1 | Centrosomal protein of 57 kDa | CEP57 | Human |
| anti_11279_42 | Gamma-aminobutyric acid type B receptor subunit 1 | GABBR1 | Human |
| anti_11280_6 | Glutamate decarboxylase 1 | GAD1 | Human |
| anti_11281_6 | Growth factor receptor-bound protein 7 | GRB7 | Human |
| anti_11282_16 | Macrophage scavenger receptor types I and II | MSR1 | Human |
| anti_11283_13 | DDB1- and CUL4-associated factor 5 | DCAF5 | Human |
| anti_11284_24 | Leukocyte-associated immunoglobulin-like receptor 1 | LAIR1 | Human |
| anti_11285_8 | Hematopoietic progenitor cell antigen CD34 | CD34 | Human |
| anti_11287_14 | Cytochrome b5 | CYB5A | Human |
| anti_11288_26 | Cytosolic purine 5'-nucleotidase | NT5C2 | Human |
| anti_11289_31 | Casein kinase I isoform delta | CSNK1D | Human |
| anti_11291_73 | Fc_MOUSE | Igh | Mouse |
| anti_11292_13 | Scavenger receptor class A member 3 | SCARA3 | Human |
| anti_11293_14 | Leucine-rich repeat neuronal protein 1 | LRRN1 | Human |
| anti_11297_54 | Neurogenic locus notch homolog protein 2 | NOTCH2 | Human |
| anti_11300_32 | Sortilin | SORT1 | Human |
| anti_11302_237 | Tenascin-R | TNR | Human |
| anti_11303_7 | Deoxynucleoside triphosphate triphosphohydrolase SAMHD1 | SAMHD1 | Human |
| anti_11307_33 | NEDD4-like E3 ubiquitin-protein ligase WWP1 | WWP1 | Human |
| anti_11308_8 | Cyclic AMP-responsive element-binding protein 3-like protein 4 | CREB3L4 | Human |
| anti_11310_8 | Desmoglein-3 | DSG3 | Human |
| anti_11311_79 | V(D)J recombination-activating protein 1 | RAG1 | Human |
| anti_11312_40 | Mismatch repair endonuclease PMS2 | PMS2 | Human |
| anti_11313_100 | Pterin-4-alpha-carbinolamine dehydratase | PCBD1 | Human |
| anti_11315_148 | Protein phosphatase 1D | PPM1D | Human |
| anti_11318_20 | Apolipoprotein A-V | APOA5 | Human |
| anti_11319_106 | Double-strand break repair protein MRE11 | MRE11 | Human |
| anti_11324_3 | Peroxidasin-like protein | PXDNL | Human |
| anti_11325_8 | Protein BTG2 | BTG2 | Human |
| anti_11327_56 | Dual specificity protein kinase CLK2 | CLK2 | Human |
| anti_11328_9 | U6 snRNA phosphodiesterase | USB1 | Human |
| anti_11334_7 | Leukocyte immunoglobulin-like receptor subfamily B member 3 | LILRB3 | Human |
| anti_11336_9 | Fanconi anemia group F protein | FANCF | Human |
| anti_11338_49 | Tyrosine-protein kinase BLK | BLK | Human |
| anti_11342_59 | Plexin domain-containing protein 2 | PLXDC2 | Human |
| anti_11350_30 | E3 ubiquitin-protein ligase CHIP | STUB1 | Human |
| anti_11351_233 | Non-homologous end-joining factor 1 | NHEJ1 | Human |
| anti_11352_42 | Titin | TTN | Human |
| anti_11353_143 | Mothers against decapentaplegic homolog 2 | SMAD2 | Human |
| anti_11356_19 | Protein DGCR14 | ESS2 | Human |
| anti_11358_15 | Growth factor receptor-bound protein 10 | GRB10 | Human |
| anti_11360_39 | Ribonucleoside-diphosphate reductase large subunit | RRM1 | Human |
| anti_11363_58 | Aquaporin-4 | AQP4 | Human |
| anti_11365_17 | Teneurin-4 | TENM4 | Human |
| anti_11368_32 | Adenylate kinase 2, mitochondrial | AK2 | Human |
| anti_11370_20 | 72 kDa inositol polyphosphate 5-phosphatase | INPP5E | Human |
| anti_11371_1 | Probable G-protein coupled receptor 101 | GPR101 | Human |
| anti_11375_49 | Forkhead box protein L2 | FOXL2 | Human |
| anti_11377_19 | Alcohol dehydrogenase class 4 mu/sigma chain | ADH7 | Human |
| anti_11378_37 | Tyrosine-protein kinase SYK | SYK | Human |
| anti_11380_84 | RNA-binding protein 24 | RBM24 | Human |
| anti_11381_56 | Ribose-phosphate pyrophosphokinase 1 | PRPS1 | Human |
| anti_11383_41 | Keratin, type II cytoskeletal 7 | KRT7 | Human |

| SOMAmer | Target Full Name | Entrez Gene Symbol | Organism |
|---|---|---|---|
| anti_11388_75 | WAP four-disulfide core domain protein 2 | WFDC2 | Human |
| anti_11390_24 | Carbonic anhydrase-related protein | CA8 | Human |
| anti_11395_5 | Protein-tyrosine kinase 2-beta | PTK2B | Human |
| anti_11396_39 | Dynein intermediate chain 1, axonemal | DNAI1 | Human |
| anti_11402_17 | Histone-lysine N-methyltransferase 2C | KMT2C | Human |
| anti_11405_150 | Caspase recruitment domain-containing protein 9 | CARD9 | Human |
| anti_11406_82 | Isobutyryl-CoA dehydrogenase, mitochondrial | ACAD8 | Human |
| anti_11407_57 | Phospholipid scramblase 3 | PLSCR3 | Human |
| anti_11422_2 | Homeobox protein DLX-3 | DLX3 | Human |
| anti_11428_31 | PDZ and LIM domain protein 1 | PDLIM1 | Human |
| anti_11429_80 | Heterogeneous nuclear ribonucleoproteins C1/C2 | HNRNPC | Human |
| anti_11431_235 | ATP-dependent DNA helicase Q1 | RECQL | Human |
| anti_11432_11 | Polyglutamine-binding protein 1 | PQBP1 | Human |
| anti_11433_11 | Tectonic-2 | TCTN2 | Human |
| anti_11436_6 | Alpha-internexin | INA | Human |
| anti_11438_6 | DnaJ homolog subfamily B member 2 | DNAJB2 | Human |
| anti_11439_88 | Rhophilin-2 | RHPN2 | Human |
| anti_11442_1 | Delta and Notch-like epidermal growth factor-related receptor | DNER | Human |
| anti_11444_49 | DNA-directed RNA polymerase III subunit RPC6 | POLR3F | Human |
| anti_11449_22 | F-actin-capping protein subunit alpha-1 | CAPZA1 | Human |
| anti_11456_2 | Melanoma-associated antigen B10 | MAGEB10 | Human |
| anti_11459_81 | RNA polymerase II elongation factor ELL | ELL | Human |
| anti_11462_8 | RNA binding protein fox-1 homolog 2 | RBFOX2 | Human |
| anti_11464_9 | Transcription factor RelB | RELB | Human |
| anti_11465_4 | Probable G-protein coupled receptor 135 | GPR135 | Human |
| anti_11468_15 | Probable RNA-binding protein 19 | RBM19 | Human |
| anti_11480_1 | Aldehyde dehydrogenase, dimeric NADP-preferring | ALDH3A1 | Human |
| anti_11487_4 | Testican-1 | SPOCK1 | Human |
| anti_11490_42 | Ubiquitin-like protein 4A | UBL4A | Human |
| anti_11505_1 | ADP-ribosyl cyclase/cyclic ADP-ribose hydrolase 1 | CD38 | Human |
| anti_11513_92 | ADP-ribosyl cyclase/cyclic ADP-ribose hydrolase 1 | CD38 | Human |
| anti_11516_7 | Fatty acid-binding protein, liver | FABP1 | Human |
| anti_11531_24 | V-type immunoglobulin domain-containing suppressor of T-cell activation | VSIR | Human |
| anti_11534_6 | Leucine-rich repeat, immunoglobulin-like domain and transmembrane domain-containing protein 3 | LRIT3 | Human |
| anti_11537_12 | Transferrin receptor protein 2 | TFR2 | Human |
| anti_11538_216 | Malonyl-CoA decarboxylase, mitochondrial | MLYCD | Human |
| anti_11540_37 | Forkhead box protein O3 | FOXO3 | Human |
| anti_11543_84 | LIM domain and actin-binding protein 1 | LIMA1 | Human |
| anti_11544_39 | PHD finger protein 3 | PHF3 | Human |
| anti_11546_7 | Cytoglobin | CYGB | Human |
| anti_11548_84 | M-phase inducer phosphatase 1 | CDC25A | Human |
| anti_11549_6 | Insulin gene enhancer protein ISL-1 | ISL1 | Human |
| anti_11551_16 | SWI/SNF-related matrix-associated actin-dependent regulator of chromatin subfamily E member 1-related | HMG20B | Human |
| anti_11557_3 | E3 ubiquitin-protein ligase SMURF1 | SMURF1 | Human |
| anti_11560_76 | Nuclear factor of activated T-cells, cytoplasmic 4 | NFATC4 | Human |
| anti_11561_32 | Fc_MOUSE | Igh | Mouse |
| anti_11562_9 | DNA polymerase epsilon subunit 2 | POLE2 | Human |
| anti_11565_58 | Zinc finger protein 23 | ZNF23 | Human |
| anti_11566_48 | Keratin, type II cytoskeletal 72 | KRT72 | Human |
| anti_11568_2 | Peptidyl-prolyl cis-trans isomerase FKBP1B | FKBP1B | Human |
| anti_11570_94 | B-cell receptor-associated protein 29 | BCAP29 | Human |
| anti_11572_4 | Dynamin-2 | DNM2 | Human |
| anti_11573_3 | Serine/arginine-rich splicing factor 6 | SRSF6 | Human |
| anti_11582_63 | DnaJ homolog subfamily A member 2 | DNAJA2 | Human |
| anti_11587_5 | MAX gene-associated protein | MGA | Human |
| anti_11590_5 | Probable RNA-binding protein 23 | RBM23 | Human |
| anti_11591_43 | Protein regulator of cytokinesis 1 | PRC1 | Human |

| SOMAmer | Target Full Name | Entrez Gene Symbol | Organism |
|---|---|---|---|
| anti_11592_1 | ELAV-like protein 1 | ELAVL1 | Human |
| anti_11593_21 | C-X-C motif chemokine 9 | CXCL9 | Human |
| anti_11596_47 | Zinc finger protein 75D | ZNF75D | Human |
| anti_11601_26 | ATP-dependent RNA helicase DHX8 | DHX8 | Human |
| anti_11602_12 | Copine-1 | CPNE1 | Human |
| anti_11606_22 | DnaJ homolog subfamily B member 6 | DNAJB6 | Human |
| anti_11607_15 | Bromodomain-containing protein 1 | BRD1 | Human |
| anti_11616_9 | Heat shock factor protein 1 | HSF1 | Human |
| anti_11617_1 | Integrin alpha-L | ITGAL | Human |
| anti_11618_83 | Transcriptional activator Myb | MYB | Human |
| anti_11629_36 | TNF receptor-associated factor 4 | TRAF4 | Human |
| anti_11638_42 | Beta-1,3-galactosyltransferase 2 | B3GALT2 | Human |
| anti_11643_73 | E3 ubiquitin-protein ligase DTX3L | DTX3L | Human |
| anti_11645_9 | Prolyl 4-hydroxylase subunit alpha-1 | P4HA1 | Human |
| anti_11646_4 | Carbohydrate sulfotransferase 9 | CHST9 | Human |
| anti_11647_6 | Frizzled-10 | FZD10 | Human |
| anti_11651_24 | Protein argonaute-1 | AGO1 | Human |
| anti_11653_69 | Sodium- and chloride-dependent glycine transporter 1 | SLC6A9 | Human |
| anti_11654_77 | Neurensin-1 | NRSN1 | Human |
| anti_11656_110 | Ena/VASP-like protein | EVL | Human |
| anti_11657_86 | Suppressor of cytokine signaling 7 | SOCS7 | Human |
| anti_11661_11 | NACHT, LRR and PYD domains-containing protein 1 | NLRP1 | Human |
| anti_11664_32 | ADP-ribosylation factor GTPase-activating protein 2 | ARFGAP2 | Human |
| anti_11667_29 | Tensin-2 | TNS2 | Human |
| anti_11669_39 | Solute carrier organic anion transporter family member 5A1 | SLCO5A1 | Human |
| anti_11670_18 | Probable G-protein coupled receptor 101 | GPR101 | Human |
| anti_11671_19 | Peregrin | BRPF1 | Human |
| anti_11677_17 | Probable palmitoyltransferase ZDHHC14 | ZDHHC14 | Human |
| anti_11678_105 | Gap junction delta-2 protein | GJD2 | Human |
| anti_11681_8 | Arf-GAP domain and FG repeat-containing protein 1 | AGFG1 | Human |
| anti_11683_19 | ADP-ribosylation factor-binding protein GGA3 | GGA3 | Human |
| anti_11690_47 | Anaphase-promoting complex subunit 7 | ANAPC7 | Human |
| anti_11692_21 | SHC-transforming protein 4 | SHC4 | Human |
| anti_11699_16 | Protein tyrosine phosphatase type IVA 2 | PTP4A2 | Human |
| anti_11708_2 | Lipocalin-1 | LCN1 | Human |
| anti_11709_29 | Carnitine O-palmitoyltransferase 1, muscle isoform | CPT1B | Human |
| anti_11712_207 | Protein unc-45 homolog A | UNC45A | Human |
| anti_11715_1 | POU domain, class 2, transcription factor 1 | POU2F1 | Human |
| anti_11716_28 | Leucine-rich repeat, immunoglobulin-like domain and transmembrane domain-containing protein 2 | LRIT2 | Human |
| anti_11817_1 | Ribosomal protein S6 kinase beta-1 | RPS6KB1 | Human |
| anti_11825_27 | Peroxisome proliferator-activated receptor gamma coactivator 1-alpha | PPARGC1A | Human |
| anti_11827_7 | Nuclear receptor ROR-gamma | RORC | Human |
| anti_11833_83 | Frataxin, mitochondrial | FXN | Human |
| anti_11835_8 | Triggering receptor expressed on myeloid cells 2 | TREM2 | Human |
| anti_11837_7 | Tumor necrosis factor receptor superfamily member 18 | TNFRSF18 | Human |
| anti_11838_130 | Piezo-type mechanosensitive ion channel component 1 | PIEZO1 | Human |
| anti_11860_3 | Fc_MOUSE | Igh | Mouse |
| anti_11872_9 | Protocadherin gamma-B1 | PCDHGB1 | Human |
| anti_11890_2 | Fc_MOUSE | Igh | Mouse |
| anti_11910_27 | Homeobox protein DLX-4 | DLX4 | Human |
| anti_11911_13 | Leucine-rich repeat-containing protein 4B | LRRC4B | Human |
| anti_11912_45 | Fc_MOUSE | Igh | Mouse |
| anti_11917_8 | Fc_MOUSE | Igh | Mouse |
| anti_11919_84 | Fc_MOUSE | Igh | Mouse |
| anti_11927_3 | Fc_MOUSE | Igh | Mouse |

| SOMAmer | Target Full Name | Entrez Gene Symbol | Organism |
|---|---|---|---|
| anti_11934_9 | Integrator complex subunit 3 | INTS3 | Human |
| anti_11939_11 | Fc_MOUSE | Igh | Mouse |
| anti_11949_25 | Epidermal growth factor | EGF | Human |
| anti_11952_1 | Immunoglobulin superfamily DCC subclass member 3 | IGDCC3 | Human |
| anti_11955_1 | Rho GTPase-activating protein 1 | ARHGAP1 | Human |
| anti_11967_23 | Cellular retinoic acid-binding protein 1 | CRABP1 | Human |
| anti_11969_5 | Protein S100-A2 | S100A2 | Human |
| anti_11986_30 | Fc_MOUSE | Igh | Mouse |
| anti_11988_24 | Receptor-type tyrosine-protein phosphatase H | PTPRH | Human |
| anti_11989_35 | ER membrane protein complex subunit 1 | EMC1 | Human |
| anti_11993_227 | Fc_MOUSE | Igh | Mouse |
| anti_12001_7 | Tight junction protein ZO-1 | TJP1 | Human |
| anti_12008_3 | T-cell antigen CD7 | CD7 | Human |
| anti_12012_33 | Fc_MOUSE | Igh | Mouse |
| anti_12014_19 | 6-pyruvoyl tetrahydrobiopterin synthase | PTS | Human |
| anti_12016_60 | E3 ubiquitin-protein ligase CBL | CBL | Human |
| anti_12018_84 | Thiamin pyrophosphokinase 1 | TPK1 | Human |
| anti_12022_12 | Mothers against decapentaplegic homolog 4 | SMAD4 | Human |
| anti_12030_82 | Desmin | DES | Human |
| anti_12033_3 | Mitotic checkpoint serine/threonine-protein kinase BUB1 | BUB1 | Human |
| anti_12041_33 | Heat shock 70 kDa protein 1-like | HSPA1L | Human |
| anti_12046_51 | TAR DNA-binding protein 43 | TARDBP | Human |
| anti_12060_28 | Growth/differentiation factor 11 | GDF11 | Human |
| anti_12067_1 | 10 kDa chaperonin_MYCTU | groS | Mycobacterium tuberculosis |
| anti_12073_32 | Antigen 85-A_MYCTU | FCN1 FCNM | Mycobacterium tuberculosis |
| anti_12073_8 | Antigen 85-A_MYCTU | FCN1 FCNM | Mycobacterium tuberculosis |
| anti_12074_11 | Antigen 85-B_MYCTU | fbpB Rv1886c MTCY180.32 | Mycobacterium tuberculosis |
| anti_12074_5 | Antigen 85-B_MYCTU | fbpB Rv1886c MTCY180.32 | Mycobacterium tuberculosis |
| anti_12075_16 | Antigen 85-C_MYCTU | fbpC mpt45 Rv0129c MTCI5.03c | Mycobacterium tuberculosis |
| anti_12075_40 | Antigen 85-C_MYCTU | fbpC mpt45 Rv0129c MTCI5.03c | Mycobacterium tuberculosis |
| anti_12087_24 | 50S ribosomal protein L7/L12_MYCTU | rplL | Mycobacterium tuberculosis |
| anti_12089_13 | Putative glyoxylase CFP32_MYCTU | cfp30B | Mycobacterium tuberculosis |
| anti_12090_3 | Adenylate kinase_MYCTU | adk | Mycobacterium tuberculosis |
| anti_12092_7 | Antigen 85-A_MYCTU | FCN1 FCNM | Mycobacterium tuberculosis |
| anti_12093_26 | Antigen 85-B_MYCTU | fbpB Rv1886c MTCY180.32 | Mycobacterium tuberculosis |
| anti_12329_21 | Ribosomal protein S6 kinase alpha-1 | RPS6KA1 | Human |
| anti_12334_25 | Serine hydroxymethyltransferase, cytosolic | SHMT1 | Human |
| anti_12338_27 | Pikachurin | EGFLAM | Human |
| anti_12340_17 | Alanine--tRNA ligase, cytoplasmic | AARS1 | Human |
| anti_12343_14 | Arf-GAP with coiled-coil, ANK repeat and PH domain-containing protein 2 | ACAP2 | Human |
| anti_12345_4 | Anaphase-promoting complex subunit 10 | ANAPC10 | Human |
| anti_12348_46 | Serine--tRNA ligase, mitochondrial | SARS2 | Human |
| anti_12350_86 | Protein C-ets-2 | ETS2 | Human |
| anti_12351_25 | Signal transducer and activator of transcription 1-alpha/beta | STAT1 | Human |
| anti_12352_70 | Arrestin domain-containing protein 3 | ARRDC3 | Human |
| anti_12356_65 | Sorcin | SRI | Human |
| anti_12357_41 | Synaptosomal-associated protein 29 | SNAP29 | Human |
| anti_12358_6 | Immunoglobulin-binding protein 1 | IGBP1 | Human |
| anti_12361_102 | Ras-related protein R-Ras2 | RRAS2 | Human |

| SOMAmer | Target Full Name | Entrez Gene Symbol | Organism |
|---|---|---|---|
| anti_12363_70 | Tribbles homolog 2 | TRIB2 | Human |
| anti_12366_16 | Gamma-crystallin D | CRYGD | Human |
| anti_12367_52 | Macoilin | MACO1 | Human |
| anti_12368_18 | Histone acetyltransferase KAT2B | KAT2B | Human |
| anti_12372_50 | Tropomyosin alpha-3 chain | TPM3 | Human |
| anti_12373_73 | Transformer-2 protein homolog beta | TRA2B | Human |
| anti_12374_8 | Platelet-activating factor acetylhydrolase IB subunit gamma | PAFAH1B3 | Human |
| anti_12376_85 | Cyclin-dependent kinase 4 inhibitor D | CDKN2D | Human |
| anti_12378_71 | Tapasin | TAPBP | Human |
| anti_12381_26 | Carbonyl reductase [NADPH] 1 | CBR1 | Human |
| anti_12382_2 | Probable ATP-dependent RNA helicase DDX58 | DDX58 | Human |
| anti_12387_7 | PDZ and LIM domain protein 4 | PDLIM4 | Human |
| anti_12391_27 | PDZ domain-containing protein 7 | PDZD7 | Human |
| anti_12394_53 | Transmembrane protein C16orf54 | C16orf54 | Human |
| anti_12395_86 | Aspartate--tRNA ligase, mitochondrial | DARS2 | Human |
| anti_12396_19 | 3-hydroxyisobutyryl-CoA hydrolase, mitochondrial | HIBCH | Human |
| anti_12398_15 | Paired box protein Pax-4 | PAX4 | Human |
| anti_12399_194 | Coiled-coil domain-containing protein 50 | CCDC50 | Human |
| anti_12400_25 | Ubiquitin-conjugating enzyme E2 T | UBE2T | Human |
| anti_12401_3 | AMSH-like protease | STAMBPL1 | Human |
| anti_12403_30 | Ras-related protein Rab-39B | RAB39B | Human |
| anti_12408_333 | Ras-related protein Rab-22A | RAB22A | Human |
| anti_12409_90 | Ras-related protein Rab-7b | RAB7B | Human |
| anti_12411_60 | Protein max | MAX | Human |
| anti_12422_143 | Arachidonate 15-lipoxygenase B | ALOX15B | Human |
| anti_12423_38 | APOBEC1 complementation factor | A1CF | Human |
| anti_12424_107 | Thymocyte nuclear protein 1 | THYN1 | Human |
| anti_12425_104 | ADP-ribosylation factor 6 | ARF6 | Human |
| anti_12428_2 | Lysophospholipase-like protein 1 | LYPLAL1 | Human |
| anti_12430_78 | Polyadenylate-binding protein-interacting protein 1 | PAIP1 | Human |
| anti_12432_23 | Calcyclin-binding protein | CACYBP | Human |
| anti_12434_25 | IST1 homolog | IST1 | Human |
| anti_12437_18 | Serine/threonine-protein kinase ULK3 | ULK3 | Human |
| anti_12438_127 | DNA-3-methyladenine glycosylase | MPG | Human |
| anti_12439_67 | Interferon regulatory factor 9 | IRF9 | Human |
| anti_12442_4 | Rho-related GTP-binding protein RhoD | RHOD | Human |
| anti_12444_39 | Nuclear receptor subfamily 5 group A member 2 | NR5A2 | Human |
| anti_12445_50 | Ankyrin repeat domain-containing protein 27 | ANKRD27 | Human |
| anti_12448_246 | Glycylpeptide N-tetradecanoyltransferase 1 | NMT1 | Human |
| anti_12449_16 | Peptidyl-prolyl cis-trans isomerase H | PPIH | Human |
| anti_12450_42 | Phosphomevalonate kinase | PMVK | Human |
| anti_12451_62 | Transcription regulator protein BACH1 | BACH1 | Human |
| anti_12452_32 | Histone-lysine N-methyltransferase SUV420H2 | KMT5C | Human |
| anti_12453_161 | Nuclear RNA export factor 1 | NXF1 | Human |
| anti_12454_105 | N-terminal Xaa-Pro-Lys N-methyltransferase 1 | NTMT1 | Human |
| anti_12455_48 | Myoneurin | MYNN | Human |
| anti_12459_13 | Pleckstrin homology domain-containing family A member 1 | PLEKHA1 | Human |
| anti_12460_18 | Proteasome subunit alpha type-7 | PSMA7 | Human |
| anti_12462_20 | Histone-lysine N-methyltransferase SETMAR | SETMAR | Human |
| anti_12471_47 | Double-stranded RNA-binding protein Staufen homolog 1 | STAU1 | Human |
| anti_12473_48 | BTB/POZ domain-containing protein KCTD5 | KCTD5 | Human |
| anti_12475_48 | Chloride intracellular channel protein 5 | CLIC5 | Human |
| anti_12477_42 | Translin | TSN | Human |
| anti_12478_15 | 60S ribosomal protein L30 | RPL30 | Human |
| anti_12479_50 | cAMP-dependent protein kinase type I-beta regulatory subunit | PRKAR1B | Human |
| anti_12482_5 | Nucleosome-remodeling factor subunit BPTF | BPTF | Human |
| anti_12483_62 | Nuclear receptor ROR-beta | RORB | Human |
| anti_12486_8 | Glutaredoxin-2, mitochondrial | GLRX2 | Human |
| anti_12493_42 | Ubiquitin thioesterase OTUB2 | OTUB2 | Human |

| SOMAmer | Target Full Name | Entrez Gene Symbol | Organism |
|---|---|---|---|
| anti_12497_29 | Tudor-interacting repair regulator protein | NUDT16L1 | Human |
| anti_12498_12 | Tax1-binding protein 3 | TAX1BP3 | Human |
| anti_12499_108 | Endophilin-A1 | SH3GL2 | Human |
| anti_12504_26 | Leiomodin-1 | LMOD1 | Human |
| anti_12507_16 | Inositol-trisphosphate 3-kinase C | ITPKC | Human |
| anti_12508_9 | Charged multivesicular body protein 3 | CHMP3 | Human |
| anti_12509_115 | COMM domain-containing protein 1 | COMMD1 | Human |
| anti_12511_83 | Solute carrier family 41 member 2 | SLC41A2 | Human |
| anti_12513_8 | Glycolipid transfer protein | GLTP | Human |
| anti_12514_16 | tRNA (guanine-N(7)-)-methyltransferase | METTL1 | Human |
| anti_12515_45 | Uridine-cytidine kinase 2 | UCK2 | Human |
| anti_12516_13 | Transcriptional enhancer factor TEF-3 | TEAD4 | Human |
| anti_12518_289 | Protein polybromo-1 | PBRM1 | Human |
| anti_12521_3 | Cyclin-dependent kinase 4 inhibitor C | CDKN2C | Human |
| anti_12522_6 | UV excision repair protein RAD23 homolog B | RAD23B | Human |
| anti_12527_50 | Thyroid hormone receptor alpha | THRA | Human |
| anti_12528_40 | ATPase WRNIP1 | WRNIP1 | Human |
| anti_12531_5 | Endogenous retrovirus group V member 1 Env polyprotein | ERVV-1 | Human |
| anti_12533_135 | Cytohesin-2 | CYTH2 | Human |
| anti_12534_10 | Calcium-binding and coiled-coil domain-containing protein 2 | CALCOCO2 | Human |
| anti_12535_2 | DNA repair protein XRCC1 | XRCC1 | Human |
| anti_12536_46 | Myotubularin-related protein 6 | MTMR6 | Human |
| anti_12537_88 | Transcriptional activator protein Pur-alpha | PURA | Human |
| anti_12538_19 | Regulator of G-protein signaling 7 | RGS7 | Human |
| anti_12543_76 | Zinc finger protein 560 | ZNF560 | Human |
| anti_12546_1 | Smoothelin | SMTN | Human |
| anti_12548_75 | Ras-related GTP-binding protein C | RRAGC | Human |
| anti_12553_5 | Serine/threonine-protein kinase VRK1 | VRK1 | Human |
| anti_12556_7 | Ubiquitin-conjugating enzyme E2 C | UBE2C | Human |
| anti_12557_18 | RNA-binding protein Nova-1 | NOVA1 | Human |
| anti_12558_3 | Ubiquitin-associated and SH3 domain-containing protein B | UBASH3B | Human |
| anti_12560_9 | 5'(3')-deoxyribonucleotidase, cytosolic type | NT5C | Human |
| anti_12568_14 | Kelch-like ECH-associated protein 1 | KEAP1 | Human |
| anti_12569_25 | T-complex protein 1 subunit epsilon | CCT5 | Human |
| anti_12571_14 | ADP-ribosylation factor-like protein 3 | ARL 3 | Human |
| anti_12573_80 | Tripartite motif-containing protein 3 | TRIM3 | Human |
| anti_12576_21 | Melanoma-associated antigen 3 | MAGEA3 | Human |
| anti_12577_100 | Flap endonuclease 1 | FEN1 | Human |
| anti_12578_13 | ADP-ribosylation factor 3 | ARF3 | Human |
| anti_12580_7 | Proteasome subunit beta type-5 | PSMB5 | Human |
| anti_12581_39 | Inositol monophosphatase 2 | IMPA2 | Human |
| anti_12583_77 | Serine/threonine-protein kinase A-Raf | ARAF | Human |
| anti_12585_39 | DNA excision repair protein ERCC-1 | ERCC1 | Human |
| anti_12587_65 | ADP-ribosylation factor-like protein 2 | ARL 2 | Human |
| anti_12590_67 | Fc_MOUSE | Igh | Mouse |
| anti_12591_27 | Oligophrenin-1 | OPHN1 | Human |
| anti_12593_33 | p53 and DNA damage-regulated protein 1 | PDRG1 | Human |
| anti_12594_5 | Grancalcin | GCA | Human |
| anti_12595_11 | Tropomodulin-1 | TMOD1 | Human |
| anti_12597_68 | Autophagy protein 5 | ATG5 | Human |
| anti_12605_1 | Exosome complex component RRP40 | EXOSC3 | Human |
| anti_12612_37 | Proteasome subunit beta type-1 | PSMB1 | Human |
| anti_12616_45 | Nuclear receptor-binding protein | NRBP1 | Human |
| anti_12617_2 | Serine/threonine-protein kinase 24 | STK24 | Human |
| anti_12618_50 | Aldo-keto reductase family 1 member C1 | AKR1C1 | Human |
| anti_12622_96 | Histone-lysine N-methyltransferase ASH1L | ASH1L | Human |
| anti_12623_84 | Protein Jumonji | JARID2 | Human |
| anti_12625_138 | Kelch-like protein 7 | KLHL7 | Human |
| anti_12626_6 | Sentrin-specific protease 7 | SENP7 | Human |
| anti_12628_31 | LanC-like protein 2 | LANCL2 | Human |
| anti_12633_3 | Tyrosine-protein phosphatase non-receptor type 9 | PTPN9 | Human |
| anti_12634_79 | Breast cancer anti-estrogen resistance protein 3 | BCAR3 | Human |

| SOMAmer | Target Full Name | Entrez Gene Symbol | Organism |
|---|---|---|---|
| anti_12635_9 | tRNA (cytosine(38)-C(5))-methyltransferase | TRDMT1 | Human |
| anti_12636_113 | N-lysine methyltransferase SMYD2 | SMYD2 | Human |
| anti_12637_7 | Carnitine O-acetyltransferase | CRAT | Human |
| anti_12641_3 | Type II inositol 1,4,5-trisphosphate 5-phosphatase | INPP5B | Human |
| anti_12644_63 | Adenylosuccinate synthetase isozyme 2 | ADSS2 | Human |
| anti_12646_2 | Ribulose-phosphate 3-epimerase | RPE | Human |
| anti_12647_52 | Histone-lysine N-methyltransferase SETD2 | SETD2 | Human |
| anti_12651_21 | [Pyruvate dehydrogenase (acetyl-transferring)] kinase isozyme 2, mitochondrial | PDK2 | Human |
| anti_12652_37 | Adenosylhomocysteinase 2 | AHCYL1 | Human |
| anti_12653_13 | Casein kinase I isoform gamma-2 | CSNK1G2 | Human |
| anti_12656_1 | Kinesin light chain 1 | KLC1 | Human |
| anti_12657_2 | GDP-L-fucose synthase | TSTA3 | Human |
| anti_12662_82 | Delta(3,5)-Delta(2,4)-dienoyl-CoA isomerase, mitochondrial | ECH1 | Human |
| anti_12663_1 | Thiosulfate sulfurtransferase | TST | Human |
| anti_12664_19 | Tyrosine-protein phosphatase non-receptor type 13 | PTPN13 | Human |
| anti_12669_30 | E3 ubiquitin-protein ligase HECW1 | HECW1 | Human |
| anti_12670_15 | Cell cycle checkpoint protein RAD1 | RAD1 | Human |
| anti_12671_35 | Sulfotransferase family cytosolic 1B member 1 | SULT1B1 | Human |
| anti_12676_1 | Protein kinase C and casein kinase substrate in neurons protein 1 | PACSIN1 | Human |
| anti_12677_164 | Protein flightless-1 homolog | FLII | Human |
| anti_12678_66 | U1 small nuclear ribonucleoprotein A | SNRPA | Human |
| anti_12682_5 | Kynurenine--oxoglutarate transaminase 3 | KYAT3 | Human |
| anti_12683_156 | Dihydropyrimidinase-related protein 5 | DPYSL5 | Human |
| anti_12685_57 | Homer protein homolog 2 | HOMER2 | Human |
| anti_12686_15 | 3-mercaptopyruvate sulfurtransferase | MPST | Human |
| anti_12687_2 | 2,4-dienoyl-CoA reductase, mitochondrial | DECR1 | Human |
| anti_12688_115 | Ribosomal protein S6 kinase alpha-6 | RPS6KA6 | Human |
| anti_12691_44 | Serine hydroxymethyltransferase, mitochondrial | SHMT2 | Human |
| anti_12692_56 | Histone-lysine N-methyltransferase, H3 lysine-79 specific | DOT1L | Human |
| anti_12693_2 | SH3 domain-binding glutamic acid-rich-like protein | SH3BGRL | Human |
| anti_12694_28 | Probable dimethyladenosine transferase | DIMT1 | Human |
| anti_12695_62 | Kelch-like protein 12 | KLHL12 | Human |
| anti_12696_166 | Protein arginine N-methyltransferase 3 | PRMT3 | Human |
| anti_12697_30 | Phosphatidylinositol 5-phosphate 4-kinase type-2 alpha | PIP4K2A | Human |
| anti_12698_72 | Importin subunit alpha-3 | KPNA4 | Human |
| anti_12699_28 | T-complex protein 1 subunit theta | CCT8 | Human |
| anti_12700_9 | ATP-citrate synthase | ACLY | Human |
| anti_12701_1 | Eukaryotic translation initiation factor 1b | EIF1B | Human |
| anti_12702_13 | E3 ubiquitin-protein ligase pellino homolog 2 | PELI2 | Human |
| anti_12703_6 | Serine/threonine-protein kinase Nek7 | NEK7 | Human |
| anti_12705_9 | Probable E3 ubiquitin-protein ligase HERC1 | HERC1 | Human |
| anti_12707_26 | Dihydropyrimidinase-related protein 3 | DPYSL3 | Human |
| anti_12708_91 | Lethal(3)malignant brain tumor-like protein 2 | L3MBTL2 | Human |
| anti_12709_63 | Histone H1x | H1FX | Human |
| anti_12711_19 | Gap junction alpha-8 protein | GJA8 | Human |
| anti_12712_9 | High mobility group protein 20A | HMG20A | Human |
| anti_12714_38 | AP-1 complex subunit gamma-like 2 | AP1G2 | Human |
| anti_12715_30 | mRNA-decapping enzyme 1B | DCP1B | Human |
| anti_12716_3 | Zinc finger protein 175 | ZNF175 | Human |
| anti_12717_65 | TOX high mobility group box family member 3 | TOX3 | Human |
| anti_12718_43 | Peptidyl-prolyl cis-trans isomerase NIMA-interacting 4 | PIN4 | Human |
| anti_12721_4 | Tax1-binding protein 1 | TAX1BP1 | Human |
| anti_12724_81 | Cold-inducible RNA-binding protein | CIRBP | Human |
| anti_12726_3 | Regulatory factor X-associated protein | RFXAP | Human |
| anti_12729_12 | Proteasome assembly chaperone 3 | PSMG3 | Human |
| anti_12731_12 | Pleckstrin homology domain-containing family A member 7 | PLEKHA7 | Human |
| anti_12734_112 | Kinesin-like protein KIF22 | KIF22 | Human |

| SOMAmer | Target Full Name | Entrez Gene Symbol | Organism |
|---|---|---|---|
| anti_12735_39 | Cold shock domain-containing protein E1 | CSDE1 | Human |
| anti_12738_43 | Nischarin | NISCH | Human |
| anti_12740_55 | Protein FEV | FEV | Human |
| anti_12743_18 | BAG family molecular chaperone regulator 5 | BAG5 | Human |
| anti_12748_6 | Bromodomain testis-specific protein | BRDT | Human |
| anti_12750_9 | Integrin beta-2 | ITGB2 | Human |
| anti_12751_26 | Zinc fingers and homeoboxes protein 1 | ZHX1 | Human |
| anti_12753_6 | Amyloid beta A4 precursor protein-binding family B member 2 | APBB2 | Human |
| anti_12759_47 | Interleukin enhancer-binding factor 3 | ILF3 | Human |
| anti_12760_34 | Zinc finger protein 774 | ZNF774 | Human |
| anti_12761_12 | Amyloid beta A4 precursor protein-binding family B member 2 | APBB2 | Human |
| anti_12763_69 | Zinc finger protein 334 | ZNF334 | Human |
| anti_12764_3 | Engulfment and cell motility protein 1 | ELMO1 | Human |
| anti_12766_33 | Probable G-protein coupled receptor 142 | GPR142 | Human |
| anti_12771_19 | Zinc finger protein 180 | ZNF180 | Human |
| anti_12772_8 | Nuclear pore complex protein Nup98-Nup96 | NUP98 | Human |
| anti_12774_12 | Potassium/sodium hyperpolarization-activated cyclic nucleotide-gated channel 1 | HCN1 | Human |
| anti_12781_2 | RISC-loading complex subunit TARBP2 | TARBP2 | Human |
| anti_12785_49 | Transcriptional regulator Kaiso | ZBTB33 | Human |
| anti_12786_61 | Glycerophosphocholine phosphodiesterase GPCPD1 | GPCPD1 | Human |
| anti_12787_47 | Zinc finger protein 134 | ZNF134 | Human |
| anti_12788_6 | SAGA-associated factor 29 homolog | SGF29 | Human |
| anti_12790_10 | Coiled-coil domain-containing protein 51 | CCDC51 | Human |
| anti_12793_4 | Piwi-like protein 1 | PIWIL1 | Human |
| anti_12794_6 | NACHT, LRR and PYD domains-containing protein 4 | NLRP4 | Human |
| anti_12795_2 | Zinc finger protein 566 | ZNF566 | Human |
| anti_12796_44 | Diphosphoinositol polyphosphate phosphohydrolase 1 | NUDT3 | Human |
| anti_12798_46 | Electroneutral sodium bicarbonate exchanger 1 | SLC4A8 | Human |
| anti_12799_65 | DnaJ homolog subfamily C member 27 | DNAJC27 | Human |
| anti_12801_33 | Interferon regulatory factor 2 | IRF2 | Human |
| anti_12803_9 | Zinc finger protein 329 | ZNF329 | Human |
| anti_12804_5 | Glucocorticoid modulatory element-binding protein 2 | GMEB2 | Human |
| anti_12808_103 | Calcium-regulated heat-stable protein 1 | CARHSP1 | Human |
| anti_12811_55 | Zinc finger protein 415 | ZNF415 | Human |
| anti_12812_25 | Acylphosphatase-2 | ACYP2 | Human |
| anti_12813_18 | EH domain-binding protein 1 | EHBP1 | Human |
| anti_12814_17 | RNA-binding protein 40 | RNPC3 | Human |
| anti_12815_9 | Isoleucine--tRNA ligase, cytoplasmic | IARS1 | Human |
| anti_12818_159 | Urea transporter 2 | SLC14A2 | Human |
| anti_12820_1 | GRB2-related adapter protein | GRAP | Human |
| anti_12821_6 | NACHT, LRR and PYD domains-containing protein 10 | NLRP10 | Human |
| anti_12825_18 | DCC-interacting protein 13-alpha | APPL1 | Human |
| anti_12827_37 | Regulator of G-protein signaling 3 | RGS3 | Human |
| anti_12830_4 | Histone deacetylase complex subunit SAP18 | SAP18 | Human |
| anti_12832_10 | Set1/Ash2 histone methyltransferase complex subunit ASH2 | ASH2L | Human |
| anti_12834_3 | Potassium voltage-gated channel subfamily F member 1 | KCNF1 | Human |
| anti_12835_101 | Pyrin domain-containing protein 1 | PYDC1 | Human |
| anti_12838_28 | Dual specificity protein phosphatase 15 | DUSP15 | Human |
| anti_12843_6 | Zinc finger protein 410 | ZNF410 | Human |
| anti_12844_10 | BAG family molecular chaperone regulator 4 | BAG4 | Human |
| anti_12846_3 | F-box/LRR-repeat protein 5 | FBXL5 | Human |
| anti_12847_27 | mRNA-capping enzyme | RNGTT | Human |
| anti_12848_9 | Rho guanine nucleotide exchange factor 2 | ARHGEF2 | Human |
| anti_12849_25 | GSK3-beta interaction protein | GSKIP | Human |
| anti_12851_5 | DNA-directed DNA/RNA polymerase mu | POLM | Human |

| SOMAmer | Target Full Name | Entrez Gene Symbol | Organism |
|---|---|---|---|
| anti_12853_112 | Tropomodulin-2 | TMOD2 | Human |
| anti_12854_3 | Transcriptional enhancer factor TEF-5 | TEAD3 | Human |
| anti_12855_16 | Cas scaffolding protein family member 4 | CASS4 | Human |
| anti_12856_14 | Transmembrane protein 237 | TMEM237 | Human |
| anti_12859_33 | Enoyl-CoA delta isomerase 2, mitochondrial | ECI2 | Human |
| anti_12860_7 | cAMP-regulated phosphoprotein 21 | ARPP21 | Human |
| anti_12862_14 | Enhancer of filamentation 1 | NEDD9 | Human |
| anti_12864_9 | Magnesium transporter NIPA4 | NIPAL4 | Human |
| anti_12869_68 | Probable ATP-dependent RNA helicase DDX6 | DDX6 | Human |
| anti_12871_10 | Twinfilin-1 | TWF1 | Human |
| anti_12872_35 | Cyclic nucleotide-gated olfactory channel | CNGA2 | Human |
| anti_12873_11 | Leukosialin | SPN | Human |
| anti_12875_28 | Ubiquitin domain-containing protein 2 | UBTD2 | Human |
| anti_12876_39 | Sperm flagellar protein 1 | SPEF1 | Human |
| anti_12878_60 | Oxysterol-binding protein-related protein 11 | OSBPL11 | Human |
| anti_12879_5 | Retinoblastoma-like protein 1 | RBL1 | Human |
| anti_12880_1 | Synaptic vesicle glycoprotein 2A | SV2A | Human |
| anti_12882_7 | Poly [ADP-ribose] polymerase 11 | PARP11 | Human |
| anti_12885_42 | Nuclear receptor subfamily 1 group D member 2 | NR1D2 | Human |
| anti_12888_18 | Histone deacetylase complex subunit SAP30 | SAP30 | Human |
| anti_12892_10 | Synaptotagmin-like protein 1 | SYTL1 | Human |
| anti_12898_5 | Fc_MOUSE | Igh | Mouse |
| anti_12900_29 | Fc_MOUSE | Igh | Mouse |
| anti_12904_180 | cubilin | CUBN | Human |
| anti_12908_15 | Apolipoprotein A-V | APOA5 | Human |
| anti_12915_26 | Fc_MOUSE | Igh | Mouse |
| anti_12916_3 | Protein p13 MTCP-1 | MTCP1 | Human |
| anti_12921_51 | Fc_MOUSE | Igh | Mouse |
| anti_12925_105 | Axin-2 | AXIN2 | Human |
| anti_12926_118 | Collectrin | TMEM27 | Human |
| anti_12931_16 | Mineralocorticoid receptor | NR3C2 | Human |
| anti_12933_17 | Ecto-NOX disulfide-thiol exchanger 1 | ENOX1 | Human |
| anti_12939_1 | DNA-directed RNA polymerases I and III subunit RPAC1 | POLR1C | Human |
| anti_12940_35 | Aldehyde dehydrogenase family 3 member B1 | ALDH3B1 | Human |
| anti_12941_24 | Fc_MOUSE | Igh | Mouse |
| anti_12956_40 | KIF1-binding protein | KIFBP | Human |
| anti_12957_62 | Cytochrome b-c1 complex subunit 7 | UQCRB | Human |
| anti_12958_8 | Fc_MOUSE | Igh | Mouse |
| anti_12963_1 | Protein GPR107 | GPR107 | Human |
| anti_12968_2 | Cysteine and glycine-rich protein 2 | CSRP2 | Human |
| anti_12970_35 | Double-stranded RNA-binding protein Staufen homolog 2 | STAU2 | Human |
| anti_12975_11 | Keratin, type I cytoskeletal 20 | KRT20 | Human |
| anti_12976_49 | Vinexin b | SORBS3 | Human |
| anti_12980_31 | Pre-mRNA-splicing factor RBM22 | RBM22 | Human |
| anti_12986_12 | Protein lin-7 homolog C | LIN7C | Human |
| anti_12987_12 | Serine/arginine-rich splicing factor 7 | SRSF7 | Human |
| anti_12988_49 | RNA-binding protein EWS | EWSR1 | Human |
| anti_12990_39 | Mitogen-activated protein kinase kinase kinase 3 | MAP3K3 | Human |
| anti_12991_49 | Cullin-9 | CUL9 | Human |
| anti_12993_21 | Nuclear protein localization protein 4 homolog | NPLOC4 | Human |
| anti_13007_66 | Epididymal-specific lipocalin-10 | LCN10 | Human |
| anti_13011_20 | 40S ribosomal protein S10 | RPS10 | Human |
| anti_13013_41 | E3 ubiquitin-protein ligase parkin | PRKN | Human |
| anti_13021_12 | Fc_MOUSE | Igh | Mouse |
| anti_13023_8 | Fc_MOUSE | Igh | Mouse |
| anti_13025_4 | Fibroblast growth factor receptor substrate 2 | FRS2 | Human |
| anti_13027_20 | Chromobox protein homolog 7 | CBX7 | Human |
| anti_13032_1 | Beclin-1 | BECN1 | Human |
| anti_13039_1 | Nuclear inhibitor of protein phosphatase 1 | PPP1R8 | Human |
| anti_13041_47 | TNF receptor-associated factor 4 | TRAF4 | Human |
| anti_13043_157 | ATPase family AAA domain-containing protein 2 | ATAD2 | Human |
| anti_13044_5 | Tumor susceptibility gene 101 protein | TSG101 | Human |
| anti_13055_53 | PH and SEC7 domain-containing protein 1 | PSD | Human |

| SOMAmer | Target Full Name | Entrez Gene Symbol | Organism |
|---|---|---|---|
| anti_13056_18 | Orphan sodium- and chloride-dependent neurotransmitter transporter NTT5 | SLC6A16 | Human |
| anti_13059_33 | Riboflavin kinase | RFK | Human |
| anti_13062_4 | Glia maturation factor gamma | GMFG | Human |
| anti_13068_139 | Copper chaperone for superoxide dismutase | CCS | Human |
| anti_13076_4 | Fragile X mental retardation syndrome-related protein 1 | FXR1 | Human |
| anti_13082_9 | Protein unc-45 homolog A | UNC45A | Human |
| anti_13083_18 | Valine--tRNA ligase | VARS1 | Human |
| anti_13085_18 | Glucagon-like peptide 1 receptor | GLP1R | Human |
| anti_13088_397 | Betacellulin | BTC | Human |
| anti_13094_75 | R-spondin-3 | RSPO3 | Human |
| anti_13097_11 | Bcl-2-like protein 2 | BCL2L2 | Human |
| anti_13101_60 | Sclerostin | SOST | Human |
| anti_13122_19 | Leucine-rich repeat transmembrane protein FLRT2 | FLRT2 | Human |
| anti_13123_3 | Leucine-rich repeat transmembrane protein FLRT3 | FLRT3 | Human |
| anti_13129_40 | Low-density lipoprotein receptor | LDLR | Human |
| anti_13130_150 | Hexokinase-2 | HK2 | Human |
| anti_13228_75 | E3 ubiquitin-protein ligase Mdm2 | MDM2 | Human |
| anti_13229_20 | Protein Mdm4 | MDM4 | Human |
| anti_13230_174 | Ig gamma-2, Kappa | IGHG2 | Human |
| anti_13240_170 | SH3 and multiple ankyrin repeat domains protein 1 | SHANK1 | Human |
| anti_13242_134 | SH3 and multiple ankyrin repeat domains protein 3 | SHANK3 | Human |
| anti_13256_21 | SH3 and multiple ankyrin repeat domains protein 1 | SHANK1 | Human |
| anti_13268_45 | Protein Wnt-5a | WNT5A | Human |
| anti_13375_48 | Synaptogyrin-3 | SYNGR3 | Human |
| anti_13384_110 | Fumarate hydratase, mitochondrial | FH | Human |
| anti_13386_248 | E3 ubiquitin-protein ligase RNF34 | RNF34 | Human |
| anti_13387_55 | ETS homologous factor | EHF | Human |
| anti_13388_57 | Neuroendocrine convertase 1 | PCSK1 | Human |
| anti_13399_33 | RELT-like protein 1 | RELL1 | Human |
| anti_13403_5 | ETS translocation variant 5 | ETV5 | Human |
| anti_13405_61 | Serine protease inhibitor Kazal-type 2 | SPINK2 | Human |
| anti_13406_161 | CMRF35-like molecule 6 | CD300C | Human |
| anti_13409_9 | Fc_MOUSE | Igh | Mouse |
| anti_13411_21 | E3 ubiquitin-protein ligase NRDP1 | RNF41 | Human |
| anti_13412_5 | WAP four-disulfide core domain protein 6 | WFDC6 | Human |
| anti_13421_17 | Protein kish-B | TMEM167B | Human |
| anti_13423_94 | Redox-regulatory protein FAM213A | PRXL2A | Human |
| anti_13428_52 | E3 ubiquitin-protein ligase ZNRF3 | ZNRF3 | Human |
| anti_13429_3 | WAP four-disulfide core domain protein 10A | WFDC10A | Human |
| anti_13431_74 | Membrane protein FAM159A | SHISAL2A | Human |
| anti_13432_9 | Protrudin | ZFYVE27 | Human |
| anti_13434_172 | Alpha-parvin | PARVA | Human |
| anti_13435_31 | Interleukin-20 receptor subunit beta | IL20RB | Human |
| anti_13436_54 | Myc-associated zinc finger protein | MAZ | Human |
| anti_13439_6 | IQ domain-containing protein F3 | IQCF3 | Human |
| anti_13441_30 | Dixin | DIXDC1 | Human |
| anti_13449_25 | Splicing factor 3B subunit 4 | SF3B4 | Human |
| anti_13450_49 | Ubiquitin carboxyl-terminal hydrolase 8 | USP8 | Human |
| anti_13451_2 | Fibronectin type III domain-containing protein 4 | FNDC4 | Human |
| anti_13452_113 | Small integral membrane protein 13 | SMIM13 | Human |
| anti_13453_2 | 39S ribosomal protein L33, mitochondrial | MRPL33 | Human |
| anti_13457_33 | ETS-related transcription factor Elf-5 | ELF5 | Human |
| anti_13459_30 | Torsin-4A | TOR4A | Human |
| anti_13463_1 | Peroxidasin homolog | PXDN | Human |
| anti_13470_43 | Parathyroid hormone/parathyroid hormone-related peptide receptor | PTH1R | Human |
| anti_13473_55 | Inositol-trisphosphate 3-kinase A | ITPKA | Human |
| anti_13476_16 | DNA/RNA-binding protein KIN17 | KIN | Human |
| anti_13477_65 | Leukotriene B4 receptor 1 | LTB4R | Human |
| anti_13481_24 | Transcription elongation factor A protein 2 | TCEA2 | Human |

| SOMAmer | Target Full Name | Entrez Gene Symbol | Organism |
|---|---|---|---|
| anti_13482_14 | CCR4-NOT transcription complex subunit 1 | CNOT1 | Human |
| anti_13484_69 | Collagen alpha-1(I) chain | COL1A1 | Human |
| anti_13487_24 | Protein unc-93 homolog B1 | UNC93B1 | Human |
| anti_13490_1 | MAGUK p55 subfamily member 6 | MPP6 | Human |
| anti_13492_44 | Stress-70 protein, mitochondrial | HSPA9 | Human |
| anti_13493_5 | Glutamate receptor ionotropic, delta-1 | GRID1 | Human |
| anti_13494_6 | Ceramide synthase 5 | CERS5 | Human |
| anti_13497_34 | Eukaryotic translation initiation factor 3 subunit J | EIF3J | Human |
| anti_13502_2 | Sodium-independent sulfate anion transporter | SLC26A11 | Human |
| anti_13503_19 | PRA1 family protein 3 | ARL6IP5 | Human |
| anti_13504_147 | Heterogeneous nuclear ribonucleoprotein R | HNRNPR | Human |
| anti_13506_10 | tRNA-dihydrouridine(20) synthase [NAD(P)+]-like | DUS2 | Human |
| anti_13509_5 | Secretory carrier-associated membrane protein 5 | SCAMP5 | Human |
| anti_13511_29 | DNA-binding protein SATB1 | SATB1 | Human |
| anti_13512_28 | Heterogeneous nuclear ribonucleoprotein R | HNRNPR | Human |
| anti_13513_174 | E3 SUMO-protein ligase PIAS3 | PIAS3 | Human |
| anti_13514_121 | Ras-related protein Rab-35 | RAB35 | Human |
| anti_13515_8 | Regulation of nuclear pre-mRNA domain-containing protein 1A | RPRD1A | Human |
| anti_13517_3 | Patched domain-containing protein 3 | PTCHD3 | Human |
| anti_13519_112 | Rho guanine nucleotide exchange factor 25 | ARHGEF25 | Human |
| anti_13524_25 | Heparan-sulfate 6-O-sulfotransferase 2 | HS6ST2 | Human |
| anti_13526_5 | Lupus La protein | SSB | Human |
| anti_13530_5 | Urotensin-2 receptor | UTS2R | Human |
| anti_13532_25 | Nuclear envelope phosphatase-regulatory subunit 1 | CNEP1R1 | Human |
| anti_13534_20 | Myomesin-2 | MYOM2 | Human |
| anti_13535_2 | Collagen type IV alpha-3-binding protein | CERT1 | Human |
| anti_13536_56 | DNA polymerase iota | POLI | Human |
| anti_13539_131 | Small conductance calcium-activated potassium channel protein 1 | KCNN1 | Human |
| anti_13541_1 | Myc target protein 1 | MYCT1 | Human |
| anti_13543_7 | Prostaglandin reductase 1 | PTGR1 | Human |
| anti_13544_9 | Rho GTPase-activating protein 45 | ARHGAP45 | Human |
| anti_13545_97 | Probable RNA-binding protein EIF1AD | EIF1AD | Human |
| anti_13548_53 | Phosphatidate phosphatase PPAPDC1A | PLPP4 | Human |
| anti_13549_15 | Disintegrin and metalloproteinase domain-containing protein 29 | ADAM29 | Human |
| anti_13552_7 | Switch-associated protein 70 | SWAP70 | Human |
| anti_13553_4 | DCN1-like protein 3 | DCUN1D3 | Human |
| anti_13554_78 | Replication initiator 1 | REPIN1 | Human |
| anti_13556_28 | 5-hydroxytryptamine receptor 2A | HTR2A | Human |
| anti_13557_3 | Nectin-3 | NECTIN3 | Human |
| anti_13563_259 | Acyl-CoA-binding domain-containing protein 7 | ACBD7 | Human |
| anti_13566_2 | RNA polymerase II subunit A C-terminal domain phosphatase SSU72 | SSU72 | Human |
| anti_13568_30 | 26S proteasome non-ATPase regulatory subunit 4 | PSMD4 | Human |
| anti_13570_43 | Vigilin | HDLBP | Human |
| anti_13574_50 | Rho-related GTP-binding protein Rho6 | RND1 | Human |
| anti_13577_25 | Splicing factor U2AF 65 kDa subunit | U2AF2 | Human |
| anti_13578_98 | Actin-binding LIM protein 3 | ABLIM3 | Human |
| anti_13580_2 | UDP-N-acetylhexosamine pyrophosphorylase | UAP1 | Human |
| anti_13587_10 | Rac GTPase-activating protein 1 | RACGAP1 | Human |
| anti_13588_11 | Annexin A9 | ANXA9 | Human |
| anti_13591_31 | DNA primase small subunit | PRIM1 | Human |
| anti_13592_22 | Nicotinamide N-methyltransferase | NNMT | Human |
| anti_13594_158 | ADP-ribosylation factor-binding protein GGA1 | GGA1 | Human |
| anti_13595_20 | Thiamine-triphosphatase | THTPA | Human |
| anti_13597_20 | Ras-related protein Rab-31 | RAB31 | Human |
| anti_13599_15 | Rab9 effector protein with kelch motifs | RABEPK | Human |
| anti_13602_6 | NHP2-like protein 1 | SNU13 | Human |
| anti_13604_27 | E3 ubiquitin-protein ligase NEURL1 | NEURL1 | Human |
| anti_13609_11 | General transcription factor II-I | GTF2I | Human |
| anti_13612_7 | Cytohesin-interacting protein | CYTIP | Human |

| SOMAmer | Target Full Name | Entrez Gene Symbol | Organism |
|---|---|---|---|
| anti_13613_23 | Adenylate kinase isoenzyme 5 | AK5 | Human |
| anti_13618_15 | Mitotic spindle assembly checkpoint protein MAD1 | MAD1L1 | Human |
| anti_13620_10 | ATPase ASNA1 | ASNA1 | Human |
| anti_13621_31 | AP-2 complex subunit alpha-2 | AP2A2 | Human |
| anti_13623_4 | Histone-lysine N-methyltransferase 2D | KMT2D | Human |
| anti_13624_17 | NAD kinase | NADK | Human |
| anti_13625_19 | Lupus La protein | SSB | Human |
| anti_13631_1 | Retinoblastoma-binding protein 5 | RBBP5 | Human |
| anti_13634_209 | Pirin | PIR | Human |
| anti_13639_101 | General vesicular transport factor p115 | USO1 | Human |
| anti_13640_5 | B-cell lymphoma 6 protein | BCL6 | Human |
| anti_13642_90 | Interferon-induced protein with tetratricopeptide repeats 3 | IFIT3 | Human |
| anti_13644_30 | Hepatocyte growth factor-regulated tyrosine kinase substrate | HGS | Human |
| anti_13645_14 | Pre-mRNA-splicing factor ATP-dependent RNA helicase PRP16 | DHX38 | Human |
| anti_13650_11 | Kv channel-interacting protein 1 | KCNIP1 | Human |
| anti_13651_54 | E3 ubiquitin-protein ligase ZFP91 | ZFP91 | Human |
| anti_13653_335 | O-acetyl-ADP-ribose deacetylase MACROD1 | MACROD1 | Human |
| anti_13654_1 | Rho-associated protein kinase 2 | ROCK2 | Human |
| anti_13655_34 | Nucleolin | NCL | Human |
| anti_13657_2 | Bifunctional polynucleotide phosphatase/kinase | PNKP | Human |
| anti_13658_31 | Platelet-derived growth factor C | PDGFC | Human |
| anti_13660_76 | Angiopoietin-2 | ANGPT2 | Human |
| anti_13661_193 | Cystatin-D | CST5 | Human |
| anti_13663_2 | Interleukin-4 | IL4 | Human |
| anti_13665_35 | Serine/threonine-protein phosphatase 2A regulatory subunit B'' subunit alpha | PPP2R3A | Human |
| anti_13666_222 | Carbonic anhydrase-related protein 10 | CA10 | Human |
| anti_13668_44 | Tyrosine-protein kinase receptor TYRO3 | TYRO3 | Human |
| anti_13680_3 | NGFI-A-binding protein 2 | NAB2 | Human |
| anti_13681_173 | Casein kinase II subunit alpha' | CSNK2A2 | Human |
| anti_13686_2 | Interleukin-5 receptor subunit alpha | IL5RA | Human |
| anti_13687_5 | C-C motif chemokine 1 | CCL1 | Human |
| anti_13688_2 | Calcyphosin-like protein | CAPSL | Human |
| anti_13689_2 | Ornithine decarboxylase | ODC1 | Human |
| anti_13690_26 | Biglycan | BGN | Human |
| anti_13692_154 | WNT1-inducible-signaling pathway protein 1 | CCN4 | Human |
| anti_13693_5 | Cerebral dopamine neurotrophic factor | CDNF | Human |
| anti_13694_24 | Cytokine receptor-like factor 2 | CRLF2 | Human |
| anti_13698_28 | Dyslexia-associated protein KIAA0319-like protein | KIAA0319L | Human |
| anti_13699_6 | Prostate-specific antigen | KLK3 | Human |
| anti_13701_2 | C-X-C motif chemokine 13 | CXCL13 | Human |
| anti_13704_5 | Hydroxymethylglutaryl-CoA synthase, mitochondrial | HMGCS2 | Human |
| anti_13706_12 | Interleukin-12 receptor subunit beta-1 | IL12RB1 | Human |
| anti_13708_56 | Kallikrein-8 | KLK8 | Human |
| anti_13711_10 | Clathrin heavy chain 1 | CLTC | Human |
| anti_13713_164 | Vesicle-trafficking protein SEC22a | SEC22A | Human |
| anti_13718_1 | Interleukin-17A | IL17A | Human |
| anti_13719_19 | Serine/threonine-protein kinase PAK 4 | PAK4 | Human |
| anti_13723_6 | Interleukin-10 | IL10 | Human |
| anti_13724_27 | Fibroblast growth factor 19 | FGF19 | Human |
| anti_13725_3 | Fibroblast growth factor 16 | FGF16 | Human |
| anti_13726_4 | T-lymphocyte activation antigen CD80 | CD80 | Human |
| anti_13727_44 | Speckle-type POZ protein | SPOP | Human |
| anti_13729_26 | Sideroflexin-5 | SFXN5 | Human |
| anti_13733_5 | Interleukin-12 subunit beta | IL12B | Human |
| anti_13734_22 | Interferon lambda-1 | IFNL1 | Human |
| anti_13735_1 | Rap1 GTPase-activating protein 1 | RAP1GAP | Human |
| anti_13738_8 | Inhibin beta A chain | INHBA | Human |
| anti_13739_3 | Leucine-rich repeat transmembrane protein FLRT1 | FLRT1 | Human |

| SOMAmer | Target Full Name | Entrez Gene Symbol | Organism |
|---|---|---|---|
| anti_13743_56 | Cullin-4B | CUL4B | Human |
| anti_13745_10 | Polycystin-2 | PKD2 | Human |
| anti_13748_4 | C-C motif chemokine 8 | CCL8 | Human |
| anti_13924_13 | Myocardial zonula adherens protein | MYZAP | Human |
| anti_13926_1 | RecQ-mediated genome instability protein 1 | RMI1 | Human |
| anti_13929_27 | Peroxisomal carnitine O-octanoyltransferase | CROT | Human |
| anti_13930_3 | DNA dC->dU-editing enzyme APOBEC-3G | APOBEC3G | Human |
| anti_13934_3 | Guanine nucleotide exchange factor DBS | MCF2L | Human |
| anti_13936_24 | Phosphoglycerate kinase 2 | PGK 2 | Human |
| anti_13939_14 | UTP--glucose-1-phosphate uridylyltransferase | UGP2 | Human |
| anti_13940_19 | Gamma-interferon-inducible protein 16 | IFI16 | Human |
| anti_13942_140 | SPRY domain-containing SOCS box protein 1 | SPSB1 | Human |
| anti_13943_38 | Protein dpy-30 homolog | DPY30 | Human |
| anti_13944_3 | Sulfotransferase 1A3 | SULT1A3 | Human |
| anti_13947_371 | Peroxisomal NADH pyrophosphatase NUDT12 | NUDT12 | Human |
| anti_13950_9 | Collagen type IV alpha-3-binding protein | CERT1 | Human |
| anti_13954_9 | Glucosamine 6-phosphate N-acetyltransferase | GNPNAT1 | Human |
| anti_13955_33 | Death-associated protein kinase 1 | DAPK1 | Human |
| anti_13958_5 | Probable RNA-binding protein 46 | RBM46 | Human |
| anti_13960_15 | Arf-GAP with GTPase, ANK repeat and PH domain-containing protein 3 | AGAP3 | Human |
| anti_13969_24 | Importin subunit alpha-7 | KPNA6 | Human |
| anti_13972_4 | 17-beta-hydroxysteroid dehydrogenase 14 | HSD17B14 | Human |
| anti_13973_62 | Tubulin--tyrosine ligase | TTL | Human |
| anti_13975_56 | Liprin-alpha-1 | PPFIA1 | Human |
| anti_13976_9 | Rho guanine nucleotide exchange factor 1 | ARHGEF1 | Human |
| anti_13977_28 | BRCA1-associated RING domain protein 1 | BARD1 | Human |
| anti_13978_122 | T-box transcription factor TBX3 | TBX3 | Human |
| anti_13979_3 | Anion exchange transporter | SLC26A7 | Human |
| anti_13982_33 | Regulator of G-protein signaling 18 | RGS18 | Human |
| anti_13984_23 | ATP-dependent RNA helicase DDX25 | DDX25 | Human |
| anti_13985_12 | E3 ubiquitin-protein ligase SMURF2 | SMURF2 | Human |
| anti_13986_6 | LanC-like protein 1 | LANCL1 | Human |
| anti_13990_1 | Pyruvate carboxylase, mitochondrial | PC | Human |
| anti_13996_16 | Phosphopantothenoylcysteine decarboxylase | PPCDC | Human |
| anti_13998_26 | Adenylosuccinate synthetase isozyme 1 | ADSS1 | Human |
| anti_14002_18 | Dedicator of cytokinesis protein 9 | DOCK9 | Human |
| anti_14005_2 | Chromodomain-helicase-DNA-binding protein 7 | CHD7 | Human |
| anti_14006_36 | Glycine N-methyltransferase | GNMT | Human |
| anti_14007_22 | Bifunctional 3'-phosphoadenosine 5'-phosphosulfate synthase 1 | PAPSS1 | Human |
| anti_14012_17 | Probable ATP-dependent RNA helicase DHX58 | DHX58 | Human |
| anti_14013_11 | TRAF family member-associated NF-kappa-B activator | TANK | Human |
| anti_14019_73 | Envoplakin | EVPL | Human |
| anti_14021_81 | Mediator of RNA polymerase II transcription subunit 4 | MED4 | Human |
| anti_14022_17 | Interleukin-17B | IL17B | Human |
| anti_14024_196 | Ectodysplasin-A, secreted form | EDA | Human |
| anti_14025_18 | Tumor necrosis factor receptor superfamily member 9 | TNFRSF9 | Human |
| anti_14026_24 | Interleukin-17F | IL17F | Human |
| anti_14030_21 | Tumor necrosis factor ligand superfamily member 4 | TNFSF4 | Human |
| anti_14031_18 | Fibroblast growth factor 7 | FGF7 | Human |
| anti_14032_2 | Vascular endothelial growth factor A, isoform 121 | VEGFA | Human |
| anti_14034_22 | Tumor-associated calcium signal transducer 2 | TACSTD2 | Human |
| anti_14035_13 | T-lymphoma invasion and metastasis-inducing protein 1 | TIAM1 | Human |
| anti_14036_116 | Multiple PDZ domain protein | MPDZ | Human |
| anti_14037_18 | Ran-binding protein 3 | RANBP3 | Human |
| anti_14039_33 | Kallikrein-5 | KLK5 | Human |
| anti_14041_13 | Granzyme B | GZMB | Human |
| anti_14042_11 | Secreted frizzled-related protein 1 | SFRP1 | Human |
| anti_14043_12 | Early endosome antigen 1 | EEA1 | Human |

| SOMAmer | Target Full Name | Entrez Gene Symbol | Organism |
|---|---|---|---|
| anti_14045_12 | Nuclear receptor coactivator 2 | NCOA2 | Human |
| anti_14049_17 | Interleukin-7 | IL7 | Human |
| anti_14050_61 | Ephrin-A4 | EFNA4 | Human |
| anti_14051_54 | Forkhead box protein C2 | FOXC2 | Human |
| anti_14052_26 | Protein unc-13 homolog A | UNC13A | Human |
| anti_14056_4 | Glycoprotein hormones alpha chain | CGA | Human |
| anti_14060_67 | Neutrophil cytosol factor 4 | NCF4 | Human |
| anti_14061_48 | Tumor necrosis factor ligand superfamily member 11 | TNFSF11 | Human |
| anti_14063_17 | Oncostatin-M | OSM | Human |
| anti_14064_21 | Peptidoglycan recognition protein 1 | PGLYRP1 | Human |
| anti_14065_11 | T-cell surface glycoprotein CD5 | CD5 | Human |
| anti_14066_49 | Membrane-associated guanylate kinase, WW and PDZ domain-containing protein 2 | MAGI2 | Human |
| anti_14067_6 | Plakophilin-2 | PKP2 | Human |
| anti_14069_61 | Carbonic anhydrase 4 | CA4 | Human |
| anti_14070_56 | Intersectin-1 | ITSN1 | Human |
| anti_14074_2 | SHC-transforming protein 2 | SHC2 | Human |
| anti_14076_74 | Cystatin-S | CST4 | Human |
| anti_14079_14 | Interleukin-18 receptor 1 | IL18R1 | Human |
| anti_14081_5 | NKG2D ligand 1 | ULBP1 | Human |
| anti_14082_56 | Talin-2 | TLN2 | Human |
| anti_14083_25 | Selenide, water dikinase 1 | SEPHS1 | Human |
| anti_14090_23 | Differentially expressed in FDCP 6 homolog | DEF6 | Human |
| anti_14091_42 | Carbonyl reductase [NADPH] 3 | CBR3 | Human |
| anti_14093_10 | Fms-related tyrosine kinase 3 ligand | FLT3LG | Human |
| anti_14095_1 | NKG2-D type II integral membrane protein | KLRK1 | Human |
| anti_14098_28 | Cysteine--tRNA ligase, cytoplasmic | CARS1 | Human |
| anti_14099_20 | Proteasome subunit alpha type-4 | PSMA4 | Human |
| anti_14100_63 | Complement C1q subcomponent subunit C | C1QC | Human |
| anti_14103_12 | Tryptase gamma | TPSG1 | Human |
| anti_14104_1 | TLR4 interactor with leucine rich repeats | TRIL | Human |
| anti_14106_46 | Rap1 GTPase-GDP dissociation stimulator 1 | RAP1GDS1 | Human |
| anti_14108_15 | Transforming growth factor beta-1 | TGFB1 | Human |
| anti_14111_15 | Thrombospondin-2 | THBS2 | Human |
| anti_14112_40 | Tumor necrosis factor receptor superfamily member 19L | RELT | Human |
| anti_14114_18 | PILR alpha-associated neural protein | PIANP | Human |
| anti_14121_24 | Tumor necrosis factor receptor superfamily member 10D | TNFRSF10D | Human |
| anti_14122_132 | E3 ubiquitin-protein ligase ZNRF3 | ZNRF3 | Human |
| anti_14125_5 | Apolipoprotein M | APOM | Human |
| anti_14127_240 | Interferon beta | IFNB1 | Human |
| anti_14128_121 | Interferon alpha-10 | IFNA10 | Human |
| anti_14129_1 | Interferon alpha-7 | IFNA7 | Human |
| anti_14131_37 | Ephrin-B2 | EFNB2 | Human |
| anti_14134_49 | Amphoterin-induced protein 2 | AMIGO2 | Human |
| anti_14135_3 | Relaxin receptor 1 | RXFP1 | Human |
| anti_14139_16 | Neuregulin-4 | NRG4 | Human |
| anti_14143_8 | Histone H2B type 2-E | HIST2H2BE | Human |
| anti_14146_92 | Histone H3.1 | H3C1 | Human |
| anti_14147_50 | Interferon gamma | IFNG | Human |
| anti_14149_9 | Interleukin-36 beta | IL36B | Human |
| anti_14151_4 | Ubiquitin-like protein ISG15 | ISG15 | Human |
| anti_14153_8 | Ephrin-A3 | EFNA3 | Human |
| anti_14157_21 | 14-3-3 protein epsilon | YWHAE | Human |
| anti_14158_17 | Annexin A5 | ANXA5 | Human |
| anti_14169_66 | Thioredoxin-like protein 4B | TXNL4B | Human |
| anti_14175_78 | SCP2 sterol-binding domain-containing protein 1 | SCP2D1 | Human |
| anti_14178_18 | Cyclin-dependent kinase inhibitor 3 | CDKN3 | Human |
| anti_14197_2 | PR domain zinc finger protein 1 | PRDM1 | Human |
| anti_14203_3 | Annexin A7 | ANXA7 | Human |
| anti_14205_6 | Protein HEXIM2 | HEXIM2 | Human |
| anti_14206_28 | Amyloid beta A4 precursor protein-binding family B member 1 | APBB1 | Human |

| SOMAmer | Target Full Name | Entrez Gene Symbol | Organism |
|---|---|---|---|
| anti_14216_35 | Probable ATP-dependent RNA helicase DDX46 | DDX46 | Human |
| anti_14227_21 | Myosin light chain 6B | MYL6B | Human |
| anti_14228_1 | Fc_MOUSE | Igh | Mouse |
| anti_14245_195 | Sorting nexin-7 | SNX7 | Human |
| anti_14246_50 | Docking protein 2 | DOK2 | Human |
| anti_14249_68 | Probable E3 ubiquitin-protein ligase MID2 | MID2 | Human |
| anti_14250_115 | Bleomycin hydrolase | BLMH | Human |
| anti_14256_4 | Fc_MOUSE | Igh | Mouse |
| anti_14260_112 | Neuroepithelial cell-transforming gene 1 protein | NET1 | Human |
| anti_14271_23 | Ras-related protein Rab-6B | RAB6B | Human |
| anti_14272_43 | F-box/LRR-repeat protein 4 | FBXL4 | Human |
| anti_14273_19 | Prolyl endopeptidase | PREP | Human |
| anti_14284_23 | eIF-2-alpha kinase GCN2 | EIF2AK4 | Human |
| anti_14286_2 | Nuclear factor of activated T-cells, cytoplasmic 1 | NFATC1 | Human |
| anti_14291_53 | Arf-GAP with GTPase, ANK repeat and PH domain-containing protein 2 | AGAP2 | Human |
| anti_14308_192 | Fc_MOUSE | Igh | Mouse |
| anti_14314_6 | Peptidyl-prolyl cis-trans isomerase-like 2 | PPIL2 | Human |
| anti_14324_52 | Structural maintenance of chromosomes protein 3 | SMC3 | Human |
| anti_14326_4 | Ubiquitin-conjugating enzyme E2 E1 | UBE2E1 | Human |
| anti_14329_4 | Death-inducer obliterator 1 | DIDO1 | Human |
| anti_14331_262 | Dynein light chain Tctex-type 1 | DYNLT1 | Human |
| anti_14334_3 | Recoverin | RCVRN | Human |
| anti_14483_8 | Alpha-crystallin_MYCTU | hspX | Mycobacterium tuberculosis |
| anti_14484_33 | 60 kDa chaperonin 2_MYCTU | groL2 | Mycobacterium tuberculosis |
| anti_14484_4 | 60 kDa chaperonin 2_MYCTU | groL2 | Mycobacterium tuberculosis |
| anti_14485_44 | Phosphate-binding protein pstS 1_MYCTU | pstS1 | Mycobacterium tuberculosis |
| anti_14485_59 | Phosphate-binding protein pstS 1_MYCTU | pstS1 | Mycobacterium tuberculosis |
| anti_14486_2 | Chaperone protein DnaK_MYCTU | dnaK | Mycobacterium tuberculosis |
| anti_14487_46 | Malate synthase G_MYCTU | glcB | Mycobacterium tuberculosis |
| anti_14488_1 | 10 kDa chaperonin_MYCTU | groS | Mycobacterium tuberculosis |
| anti_14488_3 | 10 kDa chaperonin_MYCTU | groS | Mycobacterium tuberculosis |
| anti_14489_14 | 50S ribosomal protein L7/L12_MYCTU | rplL | Mycobacterium tuberculosis |
| anti_14489_18 | 50S ribosomal protein L7/L12_MYCTU | rplL | Mycobacterium tuberculosis |
| anti_14490_127 | Probable thiol peroxidase_MYCTU | tpx | Mycobacterium tuberculosis |
| anti_14490_6 | Probable thiol peroxidase_MYCTU | tpx | Mycobacterium tuberculosis |
| anti_14491_10 | Adenylate kinase_MYCTU | adk | Mycobacterium tuberculosis |
| anti_14491_43 | Adenylate kinase_MYCTU | adk | Mycobacterium tuberculosis |
| anti_14492_11 | Antigen 85-A_MYCTU | FCN1 FCNM | Mycobacterium tuberculosis |
| anti_14492_7 | Antigen 85-A_MYCTU | FCN1 FCNM | Mycobacterium tuberculosis |
| anti_14493_16 | Antigen 85-B_MYCTU | fbpB Rv1886c MTCY180.32 | Mycobacterium tuberculosis |
| anti_14493_5 | Antigen 85-B_MYCTU | fbpB Rv1886c MTCY180.32 | Mycobacterium tuberculosis |
| anti_14494_124 | Antigen 85-C_MYCTU | fbpC mpt45 Rv0129c MTCI5.03c | Mycobacterium tuberculosis |
| anti_14494_53 | Antigen 85-C_MYCTU | fbpC mpt45 Rv0129c MTCI5.03c | Mycobacterium tuberculosis |

| SOMAmer | Target Full Name | Entrez Gene Symbol | Organism |
|---|---|---|---|
| anti_14495_161 | Growth/differentiation factor 11 | GDF11 | Human |
| anti_14496_43 | Immunogenic protein MPT64_MYCTU | mpt64 | Mycobacterium tuberculosis |
| anti_14498_14 | 60 kDa chaperonin 2_MYCTU | groL2 | Mycobacterium tuberculosis |
| anti_14499_2 | Malate synthase G_MYCTU | glcB | Mycobacterium tuberculosis |
| anti_14502_11 | Adenylate kinase_MYCTU | adk | Mycobacterium tuberculosis |
| anti_14502_14 | Adenylate kinase_MYCTU | adk | Mycobacterium tuberculosis |
| anti_14503_5 | MPT51/MPB51 antigen_MYCTU | mpt51 | Mycobacterium tuberculosis |
| anti_14504_6 | Antigen 85-A_MYCTU | FCN1 FCNM | Mycobacterium tuberculosis |
| anti_14505_57 | Antigen 85-B_MYCTU | fbpB Rv1886c MTCY180.32 | Mycobacterium tuberculosis |
| anti_14506_48 | Antigen 85-C_MYCTU | fbpC mpt45 Rv0129c MTCI5.03c | Mycobacterium tuberculosis |
| anti_14506_76 | Antigen 85-C_MYCTU | fbpC mpt45 Rv0129c MTCI5.03c | Mycobacterium tuberculosis |
| anti_14558_169 | Molybdopterin biosynthesis protein MoeX_MYCTU | moeX | strain ATCC 25618 / H37Rv |
| anti_14560_195 | Oxidoreductase_MYCTU | Rv3368c | strain ATCC 25618 / H37Rv |
| anti_14560_260 | Oxidoreductase_MYCTU | Rv3368c | strain ATCC 25618 / H37Rv |
| anti_14561_28 | Penicillin-binding protein 1A | ponA1 | strain ATCC 25618 / H37Rv |
| anti_14561_42 | Penicillin-binding protein 1A | ponA1 | strain ATCC 25618 / H37Rv |
| anti_14561_65 | Penicillin-binding protein 1A | ponA1 | strain ATCC 25618 / H37Rv |
| anti_14562_23 | Molybdopterin biosynthesis protein MoeX_MYCTU | moeX | strain ATCC 25618 / H37Rv |
| anti_14566_153 | Oxidoreductase_MYCTU | Rv3368c | strain ATCC 25618 / H37Rv |
| anti_14566_23 | Oxidoreductase_MYCTU | Rv3368c | strain ATCC 25618 / H37Rv |
| anti_14567_159 | N-acetylmuramoyl-L-alanine amidase_MYCTU | Rv3915 | strain ATCC 25618 / H37Rv |
| anti_14567_54 | N-acetylmuramoyl-L-alanine amidase_MYCTU | Rv3915 | strain ATCC 25618 / H37Rv |
| anti_14568_50 | Penicillin-binding protein 1A | ponA1 | strain ATCC 25618 / H37Rv |
| anti_14569_1 | Molybdopterin biosynthesis protein MoeX_MYCTU | moeX | strain ATCC 25618 / H37Rv |
| anti_14571_34 | Oxidoreductase_MYCTU | Rv3368c | strain ATCC 25618 / H37Rv |
| anti_14571_46 | Oxidoreductase_MYCTU | Rv3368c | strain ATCC 25618 / H37Rv |
| anti_14572_288 | Putative uncharacterized protein_MYCTX | ORFNames=TBCG_03312; | Mycobacterium tuberculosis C |
| anti_14573_74 | Penicillin-binding protein 1A | ponA1 | strain ATCC 25618 / H37Rv |
| anti_14574_16 | Molybdopterin biosynthesis protein MoeX_MYCTU | moeX | strain ATCC 25618 / H37Rv |
| anti_14577_124 | Oxidoreductase_MYCTU | Rv3368c | strain ATCC 25618 / H37Rv |
| anti_14577_18 | Oxidoreductase_MYCTU | Rv3368c | strain ATCC 25618 / H37Rv |
| anti_14578_44 | Putative uncharacterized protein_MYCTX | ORFNames=TBCG_03312; | Mycobacterium tuberculosis C |
| anti_14579_263 | Long-chain-fatty-acid--AMP ligase FadD32_MYCTU | fadD32 | strain ATCC 25618 / H37Rv |

| SOMAmer | Target Full Name | Entrez Gene Symbol | Organism |
|---|---|---|---|
| anti_14579_80 | Long-chain-fatty-acid--AMP ligase FadD32_MYCTU | fadD32 | strain ATCC 25618 / H37Rv |
| anti_14580_14 | Probable propionyl-CoA carboxylase beta chain 6_MYCTU | accD6 | strain ATCC 25618 / H37Rv |
| anti_14580_66 | Probable propionyl-CoA carboxylase beta chain 6_MYCTU | accD6 | strain ATCC 25618 / H37Rv |
| anti_14581_134 | Oxidoreductase_MYTCT | Rv2857c | strain ATCC 25618 / H37Rv |
| anti_14581_8 | Oxidoreductase_MYTCT | Rv2857c | strain ATCC 25618 / H37Rv |
| anti_14584_31 | Long-chain-fatty-acid--AMP ligase FadD32_MYCTU | fadD32 | strain ATCC 25618 / H37Rv |
| anti_14584_60 | Long-chain-fatty-acid--AMP ligase FadD32_MYCTU | fadD32 | strain ATCC 25618 / H37Rv |
| anti_14585_53 | Probable propionyl-CoA carboxylase beta chain 6_MYCTU | accD6 | strain ATCC 25618 / H37Rv |
| anti_14585_66 | Probable propionyl-CoA carboxylase beta chain 6_MYCTU | accD6 | strain ATCC 25618 / H37Rv |
| anti_14586_102 | Oxidoreductase_MYTCT | Rv2857c | strain ATCC 25618 / H37Rv |
| anti_14586_33 | Oxidoreductase_MYTCT | Rv2857c | strain ATCC 25618 / H37Rv |
| anti_14586_69 | Oxidoreductase_MYTCT | Rv2857c | strain ATCC 25618 / H37Rv |
| anti_14587_16 | Growth/differentiation factor 11 | GDF11 | Human |
| anti_14597_5 | Semaphorin-6C, cytoplasmic | SEMA6C | Human |
| anti_14599_18 | Stabilin-1 | STAB1 | Human |
| anti_14603_51 | Uncharacterized protein KIAA0040 | KIAA0040 | Human |
| anti_14605_2 | Fc_MOUSE | Igh | Mouse |
| anti_14615_46 | Keratin-associated protein 2-4 | KRTAP2-4 | Human |
| anti_14616_16 | Zinc finger protein 382 | ZNF382 | Human |
| anti_14617_7 | Fc_MOUSE | Igh | Mouse |
| anti_14618_26 | Vesicular, overexpressed in cancer, prosurvival protein 1 | VOPP1 | Human |
| anti_14619_8 | Zinc finger protein 526 | ZNF526 | Human |
| anti_14623_26 | Small ubiquitin-related modifier 3 | SUMO3 | Human |
| anti_14624_51 | Transcriptional repressor CTCF | CTCF | Human |
| anti_14628_72 | F-box only protein 3 | FBXO3 | Human |
| anti_14631_22 | Dual specificity protein phosphatase 16 | DUSP16 | Human |
| anti_14634_13 | Lutropin-choriogonadotropic hormone receptor | LHCGR | Human |
| anti_14635_28 | Fc_MOUSE | Igh | Mouse |
| anti_14636_25 | Ribonuclease UK114 | RIDA | Human |
| anti_14645_253 | Glutathione S-transferase A4 | GSTA4 | Human |
| anti_14655_1 | DnaJ homolog subfamily C member 17 | DNAJC17 | Human |
| anti_14662_6 | Myeloid zinc finger 1 | MZF1 | Human |
| anti_14674_63 | Protein kinase C and casein kinase substrate in neurons protein 3 | PACSIN3 | Human |
| anti_14687_6 | Perilipin-3 | PLIN3 | Human |
| anti_14688_6 | Tyrosine-protein phosphatase non-receptor type 7 | PTPN7 | Human |
| anti_14696_45 | Tryptase beta-2 | TPSB2 | Human |
| anti_14703_6 | Prokineticin-1 | PROK1 | Human |
| anti_14704_10 | Epithelial discoidin domain-containing receptor 1 | DDR1 | Human |
| anti_14713_46 | Azurocidin | AZU1 | Human |
| anti_14747_9 | Cytokine receptor-like factor 1 | CRLF1 | Human |
| anti_14748_31 | Rho GTPase-activating protein 5 | ARHGAP5 | Human |
| anti_14755_4 | Lactotransferrin | LTF | Human |
| anti_14757_144 | Fibroblast growth factor 8 isoform B | FGF8 | Human |
| anti_14999_10 | Malate synthase G_MYCTU | glcB | Mycobacterium tuberculosis |
| anti_14999_49 | Malate synthase G_MYCTU | glcB | Mycobacterium tuberculosis |
| anti_15000_1 | Beta-lactamase | blaA | strain ATCC 25618 / H37Rv |
| anti_15000_8 | Beta-lactamase | blaA | strain ATCC 25618 / H37Rv |

| SOMAmer | Target Full Name | Entrez Gene Symbol | Organism |
|---|---|---|---|
| anti_15001_182 | Low molecular weight antigen MTB12 | mtb12 | strain ATCC 25618 / H37Rv |
| anti_15001_2 | Low molecular weight antigen MTB12 | mtb12 | strain ATCC 25618 / H37Rv |
| anti_15001_29 | Low molecular weight antigen MTB12 | mtb12 | strain ATCC 25618 / H37Rv |
| anti_15002_16 | Ornithine carbamoyltransferase_MYCTU | argF | strain ATCC 25618 / H37Rv |
| anti_15002_17 | Ornithine carbamoyltransferase_MYCTU | argF | strain ATCC 25618 / H37Rv |
| anti_15003_10 | Probable phosphoadenosine phosphosulfate reductase_MYCTU | cysH | strain ATCC 25618 / H37Rv |
| anti_15003_24 | Probable phosphoadenosine phosphosulfate reductase_MYCTU | cysH | strain ATCC 25618 / H37Rv |
| anti_15004_52 | Putative S-adenosyl-L-methionine-dependent methyltransferase Rv1729c_MYCTU | Rv1729c | strain ATCC 25618 / H37Rv |
| anti_15005_35 | Malate synthase G_MYCTU | glcB | Mycobacterium tuberculosis |
| anti_15005_42 | Malate synthase G_MYCTU | glcB | Mycobacterium tuberculosis |
| anti_15006_1 | Beta-lactamase | blaA | strain ATCC 25618 / H37Rv |
| anti_15006_113 | Beta-lactamase | blaA | strain ATCC 25618 / H37Rv |
| anti_15007_1 | Low molecular weight antigen MTB12 | mtb12 | strain ATCC 25618 / H37Rv |
| anti_15007_43 | Low molecular weight antigen MTB12 | mtb12 | strain ATCC 25618 / H37Rv |
| anti_15007_48 | Low molecular weight antigen MTB12 | mtb12 | strain ATCC 25618 / H37Rv |
| anti_15008_14 | Ornithine carbamoyltransferase_MYCTU | argF | strain ATCC 25618 / H37Rv |
| anti_15008_28 | Ornithine carbamoyltransferase_MYCTU | argF | strain ATCC 25618 / H37Rv |
| anti_15009_68 | Probable phosphoadenosine phosphosulfate reductase_MYCTU | cysH | strain ATCC 25618 / H37Rv |
| anti_15010_1 | Putative S-adenosyl-L-methionine-dependent methyltransferase Rv1729c_MYCTU | Rv1729c | strain ATCC 25618 / H37Rv |
| anti_15010_4 | Putative S-adenosyl-L-methionine-dependent methyltransferase Rv1729c_MYCTU | Rv1729c | strain ATCC 25618 / H37Rv |
| anti_15011_186 | Complex of EsxA and EsxB_MYCTU | EsxA\|EsxB | M. tuberculosis |
| anti_15011_45 | Complex of EsxA and EsxB_MYCTU | EsxA\|EsxB | M. tuberculosis |
| anti_15012_10 | Complex of EsxG and EsxH_MYCTU | EsxG\|EsxH | M. tuberculosis |
| anti_15012_33 | Complex of EsxG and EsxH_MYCTU | EsxG\|EsxH | M. tuberculosis |
| anti_15013_1 | Malate synthase G_MYCTU | glcB | Mycobacterium tuberculosis |
| anti_15013_72 | Malate synthase G_MYCTU | glcB | Mycobacterium tuberculosis |
| anti_2192_63 | C-C motif chemokine 27 | CCL27 | Human |
| anti_2247_20 | Prokineticin-1 | PROK1 | Human |
| anti_2330_2 | Stromal cell-derived factor 1 | CXCL12 | Human |
| anti_2333_72 | Transforming growth factor beta-1 | TGFB1 | Human |
| anti_2431_17 | Cathepsin G | CTSG | Human |
| anti_2441_2 | Fibroblast growth factor 10 | FGF10 | Human |
| anti_2443_10 | Fibroblast growth factor 8 isoform B | FGF8 | Human |
| anti_2447_7 | Group IIE secretory phospholipase A2 | PLA2G2E | Human |
| anti_2449_1 | Calcium-dependent phospholipase A2 | PLA2G5 | Human |
| anti_2455_17 | Interleukin-12 | IL12A\|IL12B | Human |
| anti_2468_62 | C-C motif chemokine 20 | CCL20 | Human |
| anti_2500_2 | Angiopoietin-4 | ANGPT4 | Human |
| anti_2505_49 | GDNF family receptor alpha-3 | GFRA3 | Human |
| anti_2514_65 | Ephrin-B3 | EFNB3 | Human |
| anti_2526_53 | Tumor necrosis factor receptor superfamily member 11B | TNFRSF11B | Human |
| anti_2558_51 | Beta-endorphin | POMC | Human |

| SOMAmer | Target Full Name | Entrez Gene Symbol | Organism |
|---|---|---|---|
| anti_2573_20 | Interleukin-6 | IL6 | Human |
| anti_2578_67 | C-C motif chemokine 2 | CCL2 | Human |
| anti_2598_9 | Tumor necrosis factor receptor superfamily member 9 | TNFRSF9 | Human |
| anti_2599_51 | Tumor necrosis factor ligand superfamily member 9 | TNFSF9 | Human |
| anti_2603_61 | T-lymphocyte activation antigen CD80 | CD80 | Human |
| anti_2607_54 | Cytokine receptor-like factor 1:Cardiotrophin-like cytokine factor 1 Complex | CRLF1\|CLCF1 | Human |
| anti_2611_72 | Tyrosine-protein kinase receptor TYRO3 | TYRO3 | Human |
| anti_2612_5 | Eukaryotic translation initiation factor 5 | EIF5 | Human |
| anti_2614_28 | Ephrin-A4 | EFNA4 | Human |
| anti_2615_60 | Ephrin-A5 | EFNA5 | Human |
| anti_2616_23 | Receptor tyrosine-protein kinase erbB-2 | ERBB2 | Human |
| anti_2618_10 | Receptor tyrosine-protein kinase erbB-4 | ERBB4 | Human |
| anti_2619_72 | Tumor-associated calcium signal transducer 2 | TACSTD2 | Human |
| anti_2622_18 | Heme oxygenase 2 | HMOX2 | Human |
| anti_2623_54 | Protein E7_HPV16 | Human-virus | Human papillomavirus type 16 |
| anti_2624_31 | Protein E7_HPV18 | Human-virus | Human papillomavirus type 18 |
| anti_2626_3 | Heat shock protein HSP 90-beta | HSP90AB1 | Human |
| anti_2631_50 | Interleukin-10 receptor subunit beta | IL10RB | Human |
| anti_2632_5 | Interleukin-12 receptor subunit beta-1 | IL12RB1 | Human |
| anti_2634_2 | Cytokine receptor common subunit gamma | IL2RG | Human |
| anti_2635_61 | Layilin | LAYN | Human |
| anti_2640_3 | Macrophage-stimulating protein receptor | MST1R | Human |
| anti_2642_4 | Platelet-activating factor acetylhydrolase IB subunit beta | PAFAH1B2 | Human |
| anti_2644_11 | Protein kinase C alpha type | PRKCA | Human |
| anti_2645_54 | Protein kinase C zeta type | PRKCZ | Human |
| anti_2649_77 | Intercellular adhesion molecule 3 | ICAM3 | Human |
| anti_2658_27 | NT-3 growth factor receptor | NTRK3 | Human |
| anti_2666_53 | Decorin | DCN | Human |
| anti_2681_23 | Hepatocyte growth factor | HGF | Human |
| anti_2690_51 | Serum albumin_Bovine | ALB | Bovine |
| anti_2695_25 | Platelet endothelial cell adhesion molecule | PECAM1 | Human |
| anti_2696_87 | Persephin | PSPN | Human |
| anti_2704_74 | Tumor necrosis factor receptor superfamily member 13B | TNFRSF13B | Human |
| anti_2705_5 | C-C motif chemokine 25 | CCL25 | Human |
| anti_2708_54 | Tumor necrosis factor ligand superfamily member 18 | TNFSF18 | Human |
| anti_2711_6 | Ciliary neurotrophic factor receptor subunit alpha | CNTFR | Human |
| anti_2715_25 | Erythropoietin receptor | EPOR | Human |
| anti_2719_3 | Granulocyte colony-stimulating factor receptor | CSF3R | Human |
| anti_2723_9 | Interleukin-37 | IL37 | Human |
| anti_2728_62 | Tenascin | TNC | Human |
| anti_2730_58 | MHC class I polypeptide-related sequence A | MICA | Human |
| anti_2731_29 | NADPH--cytochrome P450 reductase | POR | Human |
| anti_2732_58 | Homeobox protein NANOG | NANOG | Human |
| anti_2734_49 | Natural cytotoxicity triggering receptor 2 | NCR2 | Human |
| anti_2737_22 | Protein NOV homolog | CCN3 | Human |
| anti_2741_22 | Sialic acid-binding Ig-like lectin 6 | SIGLEC6 | Human |
| anti_2743_5 | Sonic hedgehog protein | SHH | Human |
| anti_2746_56 | Cytokine receptor-like factor 2 | CRLF2 | Human |
| anti_2747_3 | NKG2D ligand 3 | ULBP3 | Human |
| anti_2748_3 | Activin A | INHBA | Human |
| anti_2751_16 | Azurocidin | AZU1 | Human |
| anti_2752_62 | Growth/differentiation factor 5 | GDF5 | Human |
| anti_2760_2 | Protein FAM107A | FAM107A | Human |
| anti_2761_49 | Fibroblast growth factor 18 | FGF18 | Human |
| anti_2763_66 | Fibroblast growth factor 20 | FGF20 | Human |

| SOMAmer | Target Full Name | Entrez Gene Symbol | Organism |
|---|---|---|---|
| anti_2764_20 | Fibroblast growth factor 9 | FGF9 | Human |
| anti_2769_3 | Protein Rev_HV2BE | rev | HIV-2 |
| anti_2773_50 | Interleukin-10 | IL10 | Human |
| anti_2775_54 | Interleukin-17F | IL17F | Human |
| anti_2781_63 | C-C motif chemokine 4-like | CCL4L1 | Human |
| anti_2785_15 | C-C motif chemokine 8 | CCL8 | Human |
| anti_2794_60 | Superoxide dismutase [Cu-Zn] | SOD1 | Human |
| anti_2797_56 | Apolipoprotein B | APOB | Human |
| anti_2805_6 | Angiotensin-converting enzyme 2 | ACE2 | Human |
| anti_2806_49 | Activin receptor type-1B | ACVR1B | Human |
| anti_2809_25 | A disintegrin and metalloproteinase with thrombospondin motifs 4 | ADAMTS4 | Human |
| anti_2813_11 | Agouti-related protein | AGRP | Human |
| anti_2816_50 | Basal Cell Adhesion Molecule | BCAM | Human |
| anti_2823_7 | COMM domain-containing protein 7 | COMMD7 | Human |
| anti_2826_53 | Ectodysplasin-A, secreted form | EDA | Human |
| anti_2827_23 | Fractalkine | CX3CL1 | Human |
| anti_2828_82 | Kunitz-type protease inhibitor 1 | SPINT1 | Human |
| anti_2829_19 | Interleukin-27 | IL27\|EBI3 | Human |
| anti_2831_29 | Kallikrein-11 | KLK11 | Human |
| anti_2833_20 | Kallikrein-4 | KLK4 | Human |
| anti_2834_54 | Kallikrein-8 | KLK8 | Human |
| anti_2835_1 | X-ray repair cross-complementing protein 6 | XRCC6 | Human |
| anti_2838_53 | Matrix metalloproteinase-17 | MMP17 | Human |
| anti_2839_2 | Tumor necrosis factor ligand superfamily member 4 | TNFSF4 | Human |
| anti_2843_13 | Kunitz-type protease inhibitor 2 | SPINT2 | Human |
| anti_2848_2 | Wnt inhibitory factor 1 | WIF1 | Human |
| anti_2853_68 | Serine/threonine-protein kinase Chk1 | CHEK1 | Human |
| anti_2858_29 | Histone acetyltransferase type B catalytic subunit | HAT1 | Human |
| anti_2864_2 | Dual specificity mitogen-activated protein kinase kinase 1 | MAP2K1 | Human |
| anti_2865_77 | Histone acetyltransferase KAT6A | KAT6A | Human |
| anti_2867_52 | RAC-alpha serine/threonine-protein kinase | AKT1 | Human |
| anti_2875_15 | TATA-box-binding protein | TBP | Human |
| anti_2876_74 | DNA topoisomerase 1 | TOP 1 | Human |
| anti_2878_66 | Tyrosine-protein kinase Yes | YES1 | Human |
| anti_2889_37 | Cardiotrophin-1 | CTF1 | Human |
| anti_2890_59 | C-C motif chemokine 28 | CCL28 | Human |
| anti_2891_1 | B-cell receptor CD22 | CD22 | Human |
| anti_2906_55 | Interleukin-4 | IL4 | Human |
| anti_2911_27 | Midkine | MDK | Human |
| anti_2915_6 | Proliferating cell nuclear antigen | PCNA | Human |
| anti_2939_10 | Artemin | ARTN | Human |
| anti_2944_66 | Neuroblastoma suppressor of tumorigenicity 1 | NBL1 | Human |
| anti_2945_25 | Estrogen receptor | ESR1 | Human |
| anti_2949_6 | Group 10 secretory phospholipase A2 | PLA2G10 | Human |
| anti_2962_50 | Parathyroid hormone-related protein | PTHLH | Human |
| anti_2968_61 | Tumor necrosis factor ligand superfamily member 15 | TNFSF15 | Human |
| anti_2969_11 | Serine/threonine-protein kinase receptor R3 | ACVRL1 | Human |
| anti_2970_60 | Amphiregulin | AREG | Human |
| anti_2972_57 | Bone morphogenetic protein 7 | BMP7 | Human |
| anti_2973_15 | Platelet glycoprotein 4 | CD36 | Human |
| anti_2975_19 | Connective tissue growth factor | CCN2 | Human |
| anti_2977_7 | Tumor necrosis factor receptor superfamily member EDAR | EDAR | Human |
| anti_2979_8 | C-X-C motif chemokine 5 | CXCL5 | Human |
| anti_2982_82 | Galectin-4 | LGALS4 | Human |
| anti_2987_37 | Histone H1.2 | H1-2 | Human |
| anti_2988_57 | Inducible T-cell costimulator | ICOS | Human |
| anti_2989_17 | Interferon gamma | IFNG | Human |
| anti_2991_9 | Interleukin-1 receptor type 1 | IL1R1 | Human |
| anti_2993_1 | Interleukin-18 receptor accessory protein | IL18RAP | Human |
| anti_2994_71 | Interleukin-1 receptor-like 2 | IL1RL2 | Human |

| SOMAmer | Target Full Name | Entrez Gene Symbol | Organism |
|---|---|---|---|
| anti_2998_53 | Junctional adhesion molecule C | JAM3 | Human |
| anti_3003_29 | Natural cytotoxicity triggering receptor 3 | NCR3 | Human |
| anti_3004_67 | Programmed cell death 1 ligand 2 | PDCD1LG2 | Human |
| anti_3010_53 | Thymic stromal lymphopoietin | TSLP | Human |
| anti_3022_4 | Cytotoxic T-lymphocyte protein 4 | CTLA4 | Human |
| anti_3025_50 | Fibroblast growth factor 2 | FGF2 | Human |
| anti_3028_36 | Ck-beta-8-1 | CCL23 | Human |
| anti_3033_57 | Galectin-2 | LGALS2 | Human |
| anti_3034_1 | Glial fibrillary acidic protein | GFAP | Human |
| anti_3038_9 | C-X-C motif chemokine 11 | CXCL11 | Human |
| anti_3040_59 | C-C motif chemokine 3 | CCL3 | Human |
| anti_3047_95 | Diacylglycerol kinase_ECOLI | dgkA | Escherichia coli (strain K12) |
| anti_3052_8 | Tumor necrosis factor ligand superfamily member 6, soluble form | FASLG | Human |
| anti_3053_49 | Fms-related tyrosine kinase 3 ligand | FLT3LG | Human |
| anti_3055_54 | Interleukin-4 receptor subunit alpha | IL4R | Human |
| anti_3056_11 | NKG2-D type II integral membrane protein | KLRK1 | Human |
| anti_3059_50 | Tumor necrosis factor ligand superfamily member 13B | TNFSF13B | Human |
| anti_3062_6 | N-acylneuraminate cytidylyltransferase_NEIME | neuA | Neisseria meningitidis |
| anti_3065_65 | Fibroblast growth factor 5 | FGF5 | Human |
| anti_3070_1 | Interleukin-2 | IL2 | Human |
| anti_3072_4 | Interleukin-13 | IL13 | Human |
| anti_3073_51 | Interleukin-18-binding protein | IL18BP | Human |
| anti_3078_1 | Placenta growth factor | PGF | Human |
| anti_3081_70 | NKG2D ligand 1 | ULBP1 | Human |
| anti_3082_9 | NKG2D ligand 2 | ULBP2 | Human |
| anti_3083_71 | Tumor necrosis factor receptor superfamily member 27 | EDA2R | Human |
| anti_3091_70 | Aurora kinase A | AURKA | Human |
| anti_3115_64 | Mitogen-activated protein kinase 1 | MAPK1 | Human |
| anti_3132_1 | Vascular endothelial growth factor C | VEGFC | Human |
| anti_3143_3 | T-cell surface glycoprotein CD4 | CD4 | Human |
| anti_3151_6 | Interleukin-2 receptor subunit alpha | IL2RA | Human |
| anti_3166_92 | Myeloid cell surface antigen CD33 | CD33 | Human |
| anti_3168_8 | A disintegrin and metalloproteinase with thrombospondin motifs 5 | ADAMTS5 | Human |
| anti_3170_6 | Methionine aminopeptidase 2 | METAP2 | Human |
| anti_3173_49 | N-acylethanolamine-hydrolyzing acid amidase | NAAA | Human |
| anti_3174_2 | A disintegrin and metalloproteinase with thrombospondin motifs 1 | ADAMTS1 | Human |
| anti_3177_49 | Carbonic anhydrase 4 | CA4 | Human |
| anti_3178_5 | Dipeptidyl peptidase 1 | CTSC | Human |
| anti_3187_52 | Cysteine-rich secretory protein 3 | CRISP3 | Human |
| anti_3189_61 | Enteropeptidase | TMPRSS15 | Human |
| anti_3191_50 | WAP, kazal, immunoglobulin, kunitz and NTR domain-containing protein 1 | WFIKKN1 | Human |
| anti_3192_3 | Cytosolic non-specific dipeptidase | CNDP2 | Human |
| anti_3196_6 | Hyaluronan and proteoglycan link protein 1 | HAPLN1 | Human |
| anti_3200_49 | Kallikrein-13 | KLK13 | Human |
| anti_3201_49 | Kallikrein-5 | KLK5 | Human |
| anti_3202_28 | Kremen protein 2 | KREMEN2 | Human |
| anti_3204_2 | Leukotriene A-4 hydrolase | LTA4H | Human |
| anti_3208_2 | Matrilin-3 | MATN3 | Human |
| anti_3209_69 | Matrix extracellular phosphoglycoprotein | MEPE | Human |
| anti_3210_1 | Methionine aminopeptidase 1 | METAP1 | Human |
| anti_3212_30 | Neutral ceramidase | ASAH2 | Human |
| anti_3221_54 | Secreted frizzled-related protein 1 | SFRP1 | Human |
| anti_3222_11 | Semaphorin-3A | SEMA3A | Human |
| anti_3235_50 | WAP, Kazal, immunoglobulin, Kunitz and NTR domain-containing protein 2 | WFIKKN2 | Human |
| anti_3236_12 | Glycogen synthase kinase-3 beta | GSK3B | Human |
| anti_3280_49 | Aggrecan core protein | ACAN | Human |

| SOMAmer | Target Full Name | Entrez Gene Symbol | Organism |
|---|---|---|---|
| anti_3281_19 | Angiopoietin-related protein 3 | ANGPTL3 | Human |
| anti_3284_75 | Biglycan | BGN | Human |
| anti_3289_19 | Carbonic anhydrase-related protein 10 | CA10 | Human |
| anti_3290_50 | CD109 antigen | CD109 | Human |
| anti_3294_55 | Cryptic protein | CFC1 | Human |
| anti_3296_92 | Contactin-2 | CNTN2 | Human |
| anti_3299_29 | Contactin-5 | CNTN5 | Human |
| anti_3312_64 | High affinity immunoglobulin gamma Fc receptor I | FCGR1A | Human |
| anti_3314_74 | GDNF family receptor alpha-1 | GFRA1 | Human |
| anti_3315_15 | Glypican-2 | GPC2 | Human |
| anti_3317_33 | Serine protease HTRA2, mitochondrial | HTRA2 | Human |
| anti_3321_2 | Interleukin-24 | IL24 | Human |
| anti_3322_52 | Leucine-rich repeats and immunoglobulin-like domains protein 3 | LRIG3 | Human |
| anti_3323_37 | Low-density lipoprotein receptor-related protein 8 | LRP8 | Human |
| anti_3324_51 | T-lymphocyte surface antigen Ly-9 | LY9 | Human |
| anti_3326_58 | Cell adhesion molecule 1 | CADM1 | Human |
| anti_3341_33 | Tyrosine-protein kinase ABL1 | ABL1 | Human |
| anti_3342_76 | Abelson tyrosine-protein kinase 2 | ABL2 | Human |
| anti_3346_72 | Aurora kinase B | AURKB | Human |
| anti_3347_9 | beta-adrenergic receptor kinase 1 | GRK2 | Human |
| anti_3350_53 | Calcium/calmodulin-dependent protein kinase type II subunit alpha | CAMK2A | Human |
| anti_3351_1 | Calcium/calmodulin-dependent protein kinase type II subunit beta | CAMK2B | Human |
| anti_3356_50 | Carbonic anhydrase 7 | CA7 | Human |
| anti_3357_67 | Cyclin-dependent kinase 2:Cyclin-A2 complex | CDK2\|CCNA2 | Human |
| anti_3358_51 | Cyclin-dependent kinase 5:Cyclin-dependent kinase 5 activator 1 complex | CDK5\|CDK5R1 | Human |
| anti_3359_11 | Cyclin-dependent kinase 8:Cyclin-C complex | CDK8\|CCNC | Human |
| anti_3360_50 | Serine/threonine-protein kinase Chk2 | CHEK2 | Human |
| anti_3361_26 | C-type lectin domain family 4 member K | CD207 | Human |
| anti_3364_76 | Cathepsin L2 | CTSV | Human |
| anti_3373_5 | Granzyme H | GZMH | Human |
| anti_3374_49 | Tyrosine-protein kinase HCK | HCK | Human |
| anti_3376_49 | Interleukin-17 receptor D | IL17RD | Human |
| anti_3379_29 | Protein kinase C iota type | PRKCI | Human |
| anti_3387_1 | Serine/threonine-protein kinase PAK 3 | PAK3 | Human |
| anti_3388_58 | Serine/threonine-protein kinase PAK 5 | PAK5 | Human |
| anti_3390_72 | PIK3CA/PIK3R1 | PIK3CA\|PIK3R1 | Human |
| anti_3394_81 | Serine/threonine-protein kinase PLK1 | PLK1 | Human |
| anti_3397_7 | Tyrosine-protein phosphatase non-receptor type 11 | PTPN11 | Human |
| anti_3400_49 | Serine/threonine-protein kinase TBK1 | TBK1 | Human |
| anti_3401_8 | Tyrosine-protein phosphatase non-receptor type 2 | PTPN2 | Human |
| anti_3404_51 | Tryptase gamma | TPSG1 | Human |
| anti_3405_6 | Ubiquitin-fold modifier-conjugating enzyme 1 | UFC1 | Human |
| anti_3412_7 | Apoptosis regulator Bcl-2 | BCL2 | Human |
| anti_3414_40 | Cytoplasmic tyrosine-protein kinase BMX | BMX | Human |
| anti_3415_61 | Bone sialoprotein 2 | IBSP | Human |
| anti_3420_21 | Carbonic anhydrase 13 | CA13 | Human |
| anti_3421_54 | Tumor necrosis factor ligand superfamily member 8 | TNFSF8 | Human |
| anti_3422_4 | Cyclin-dependent kinase 1:G2/mitotic-specific cyclin-B1 complex | CDK1\|CCNB1 | Human |
| anti_3423_59 | Chymase | CMA1 | Human |
| anti_3427_63 | Casein kinase II subunit alpha | CSNK2A1 | Human |
| anti_3432_21 | Ephrin type-A receptor 3 | EPHA3 | Human |
| anti_3440_7 | Granzyme A | GZMA | Human |
| anti_3441_64 | Glycogen synthase kinase-3 alpha | GSK3A | Human |
| anti_3443_61 | Homeodomain-interacting protein kinase 3 | HIPK3 | Human |
| anti_3445_53 | Interleukin-15 receptor subunit alpha | IL15RA | Human |
| anti_3447_64 | Interleukin-8 | CXCL8 | Human |
| anti_3448_13 | Insulin receptor | INSR | Human |
| anti_3452_17 | Tyrosine-protein kinase Lck | LCK | Human |

| SOMAmer | Target Full Name | Entrez Gene Symbol | Organism |
|---|---|---|---|
| anti_3453_87 | Tyrosine-protein kinase Lyn | LYN | Human |
| anti_3457_57 | Periostin | POSTN | Human |
| anti_3459_49 | Platelet-derived growth factor receptor beta | PDGFRB | Human |
| anti_3461_58 | Brevican core protein | BCAN | Human |
| anti_3466_8 | cAMP-dependent protein kinase catalytic subunit alpha | PRKACA | Human |
| anti_3469_74 | Ribosomal protein S6 kinase alpha-3 | RPS6KA3 | Human |
| anti_3472_40 | Baculoviral IAP repeat-containing protein 5 | BIRC5 | Human |
| anti_3473_78 | Thrombopoietin Receptor | MPL | Human |
| anti_3479_71 | Trypsin-3 | PRSS3 | Human |
| anti_3480_7 | Dual specificity protein phosphatase 3 | DUSP3 | Human |
| anti_3481_87 | Xaa-Pro aminopeptidase 1 | XPNPEP1 | Human |
| anti_3486_58 | Fibroblast growth factor 1 | FGF1 | Human |
| anti_3487_32 | C-X-C motif chemokine 13 | CXCL13 | Human |
| anti_3489_9 | Ciliary neurotrophic factor | CNTF | Human |
| anti_3494_71 | Fibroblast growth factor 17 | FGF17 | Human |
| anti_3495_15 | C-X-C motif chemokine 6 | CXCL6 | Human |
| anti_3496_68 | Glutaredoxin-1_ECOLI | grxA | Escherichia coli (strain K12) |
| anti_3497_13 | Interferon alpha-2 | IFNA2 | Human |
| anti_3498_53 | Interleukin-17A | IL17A | Human |
| anti_3499_77 | Interleukin-17B | IL17B | Human |
| anti_3505_6 | Lymphotoxin alpha1:beta2 | LTA\|LTB | Human |
| anti_3507_1 | Malic dehydrogenase_THETH | mdh | Thermus thermophilus |
| anti_3512_72 | Adenylate kinase_GEOSE | adk | Bacillus stearothermophilus |
| anti_3513_13 | Tyrosine-protein phosphatase YopH_YEREN | yopH | Yersinia enterocolitica |
| anti_3517_1 | Alpha-hemolysin_STAAU | hly | Staphylococcus aureus |
| anti_3520_58 | Transforming growth factor beta-3 | TGFB3 | Human |
| anti_3521_16 | Thyroid Stimulating Hormone | CGA\|TSHB | Human |
| anti_3522_57 | Vasoactive Intestinal Peptide | VIP | Human |
| anti_3534_14 | CD40 ligand | CD40LG | Human |
| anti_3538_26 | Aromatic-L-amino-acid decarboxylase | DDC | Human |
| anti_3593_72 | Caspase-3 | CASP3 | Human |
| anti_3594_6 | Cathepsin E | CTSE | Human |
| anti_3616_3 | N-acetylglucosamine-6-sulfatase | GNS | Human |
| anti_3620_67 | Interleukin-22 receptor subunit alpha-1 | IL22RA1 | Human |
| anti_3654_27 | BMP-binding endothelial regulator protein | BMPER | Human |
| anti_3656_9 | Cadherin-12 | CDH12 | Human |
| anti_3684_78 | Macrophage scavenger receptor types I and II | MSR1 | Human |
| anti_3717_23 | Luciferin 4-monooxygenase_PHOPY | | Common eastern firefly |
| anti_3719_2 | Cyclin-dependent kinase inhibitor 1B | CDKN1B | Human |
| anti_3721_5 | Nodulation protein H_NOH4 | nodH | Ensifer meliloti |
| anti_3723_1 | Brain natriuretic peptide 32 | NPPB | Human |
| anti_3724_64 | Exendin-4_HELSU | | Gila monster |
| anti_3727_35 | Peptide YY | PYY | Human |
| anti_3728_52 | Secretin | SCT | Human |
| anti_3730_81 | Tumor necrosis factor receptor superfamily member 4 | TNFRSF4 | Human |
| anti_3738_54 | Macrophage colony-stimulating factor 1 | CSF1 | Human |
| anti_3742_78 | Nigrin b_SAMNI | | European elder |
| anti_3743_1 | Stromelysin-2 | MMP10 | Human |
| anti_3761_4 | Prostaglandin G/H synthase 2 | PTGS2 | Human |
| anti_3766_51 | Syntaxin-1A | STX1A | Human |
| anti_3773_15 | Angiopoietin-1 receptor, soluble | TEK | Human |
| anti_3795_6 | Disintegrin and metalloproteinase domain-containing protein 9 | ADAM9 | Human |
| anti_3797_1 | Cadherin-2 | CDH2 | Human |
| anti_3798_71 | Carbonic anhydrase 9 | CA9 | Human |
| anti_3800_71 | Creatine kinase B-type | CKB | Human |

| SOMAmer | Target Full Name | Entrez Gene Symbol | Organism |
|---|---|---|---|
| anti_3802_50 | Cystatin-S | CST4 | Human |
| anti_3803_10 | Cystatin-D | CST5 | Human |
| anti_3806_55 | Ephrin type-A receptor 5 | EPHA5 | Human |
| anti_3807_1 | Fibroblast growth factor 23 | FGF23 | Human |
| anti_3808_76 | Fibroblast growth factor receptor 2 | FGFR2 | Human |
| anti_3809_1 | Fibroblast growth factor receptor 3 | FGFR3 | Human |
| anti_3810_50 | Tyrosine-protein kinase Fgr | FGR | Human |
| anti_3813_3 | Tyrosine-protein kinase Fyn | FYN | Human |
| anti_3814_63 | Interleukin-11 receptor subunit alpha | IL11RA | Human |
| anti_3815_14 | Interleukin-12 receptor subunit beta-2 | IL12RB2 | Human |
| anti_3817_18 | Protein kinase C theta type | PRKCQ | Human |
| anti_3821_28 | MAP kinase-activated protein kinase 5 | MAPKAPK5 | Human |
| anti_3823_9 | Megakaryocyte-associated tyrosine-protein kinase | MATK | Human |
| anti_3825_18 | Mitogen-activated protein kinase 8 | MAPK8 | Human |
| anti_3827_22 | Serine/threonine-protein kinase PAK 6 | PAK6 | Human |
| anti_3828_54 | Platelet-derived growth factor C | PDGFC | Human |
| anti_3831_21 | Phosphatidylinositol 3,4,5-trisphosphate 3-phosphatase and dual-specificity protein phosphatase PTEN | PTEN | Human |
| anti_3832_51 | Protein-tyrosine kinase 6 | PTK6 | Human |
| anti_3835_11 | Toll-like receptor 2 | TLR2 | Human |
| anti_3836_51 | Ubiquitin-fold modifier 1 | UFM1 | Human |
| anti_3837_6 | Tyrosine-protein kinase ZAP-70 | ZAP70 | Human |
| anti_3839_60 | AH receptor-interacting protein | AIP | Human |
| anti_3845_51 | Dynein light chain roadblock-type 1 | DYNLRB1 | Human |
| anti_3849_56 | Green fluorescent protein_AEQVI | GFP | Jellyfish |
| anti_3852_19 | DnaJ homolog subfamily B member 1 | DNAJB1 | Human |
| anti_3854_24 | Nascent polypeptide-associated complex subunit alpha | NACA | Human |
| anti_3858_5 | Low molecular weight phosphotyrosine protein phosphatase | ACP1 | Human |
| anti_3859_50 | Proteasome subunit alpha type-1 | PSMA1 | Human |
| anti_3860_7 | Proteasome subunit alpha type-6 | PSMA6 | Human |
| anti_3864_5 | 40S ribosomal protein S7 | RPS7 | Human |
| anti_3865_53 | Ribosomal protein S6 kinase alpha-5 | RPS6KA5 | Human |
| anti_3866_7 | Ribosome maturation protein SBDS | SBDS | Human |
| anti_3867_49 | Seizure 6-like protein 2 | SEZ6L2 | Human |
| anti_3868_8 | Small glutamine-rich tetratricopeptide repeat-containing protein alpha | SGTA | Human |
| anti_3873_51 | Thyroid peroxidase | TPO | Human |
| anti_3874_8 | Ubiquitin-conjugating enzyme E2 L3 | UBE2L3 | Human |
| anti_3875_62 | AT-rich interactive domain-containing protein 3A | ARID3A | Human |
| anti_3879_50 | Hsp90 co-chaperone Cdc37 | CDC37 | Human |
| anti_3881_49 | Dynein light chain 1, cytoplasmic | DYNLL1 | Human |
| anti_3887_90 | Importin subunit beta-1 | KPNB1 | Human |
| anti_3889_64 | Lamin-B1 | LMNB1 | Human |
| anti_3891_56 | Methyl-CpG-binding domain protein 4 | MBD4 | Human |
| anti_3892_21 | Mediator of RNA polymerase II transcription subunit 1 | MED1 | Human |
| anti_3893_55 | Mesothelin | MSLN | Human |
| anti_3893_64 | Mesothelin | MSLN | Human |
| anti_3896_5 | Phosphoglycerate mutase 1 | PGAM1 | Human |
| anti_3897_61 | Pyridoxal phosphate phosphatase | PDXP | Human |
| anti_3898_5 | 26S proteasome non-ATPase regulatory subunit 7 | PSMD7 | Human |
| anti_3903_49 | Sorting nexin-4 | SNX4 | Human |
| anti_4122_12 | Epithelial discoidin domain-containing receptor 1 | DDR1 | Human |
| anti_4123_60 | Fibroblast growth factor 4 | FGF4 | Human |
| anti_4124_24 | Heat shock 70 kDa protein 1A | HSPA1A | Human |
| anti_4128_27 | C-C motif chemokine 24 | CCL24 | Human |
| anti_4130_71 | Fibroblast growth factor 6 | FGF6 | Human |
| anti_4132_27 | Follistatin | FST | Human |
| anti_4133_54 | Granzyme B | GZMB | Human |
| anti_4134_4 | Heparin-binding EGF-like growth factor | HBEGF | Human |
| anti_4136_40 | Interleukin-17D | IL17D | Human |
| anti_4137_57 | Interleukin-25 | IL25 | Human |

| SOMAmer | Target Full Name | Entrez Gene Symbol | Organism |
|---|---|---|---|
| anti_4138_25 | Interleukin-20 | IL20 | Human |
| anti_4140_3 | Interleukin-7 | IL7 | Human |
| anti_4141_79 | C-X-C motif chemokine 10 | CXCL10 | Human |
| anti_4143_74 | Lymphotactin | XCL1 | Human |
| anti_4144_13 | C-C motif chemokine 13 | CCL13 | Human |
| anti_4145_58 | Neurotrophin-3 | NTF3 | Human |
| anti_4146_58 | Neurotrophin-4 | NTF4 | Human |
| anti_4150_75 | Plasmin | PLG | Human |
| anti_4156_74 | Transforming growth factor beta-2 | TGFB2 | Human |
| anti_4158_54 | Urokinase-type plasminogen activator | PLAU | Human |
| anti_4163_5 | Histone H2A.z | H2AZ1 | Human |
| anti_4165_2 | Thyroglobulin | TG | Human |
| anti_4184_43 | Eukaryotic translation initiation factor 4E-binding protein 2 | EIF4EBP2 | Human |
| anti_4188_1 | Aflatoxin B1 aldehyde reductase member 2 | AKR7A2 | Human |
| anti_4212_5 | Epidermal growth factor receptor substrate 15-like 1 | EPS15L1 | Human |
| anti_4217_49 | 3-hydroxyacyl-CoA dehydrogenase type-2 | HSD17B10 | Human |
| anti_4220_39 | Tyrosine-protein kinase Fer | FER | Human |
| anti_4224_7 | Heterogeneous nuclear ribonucleoprotein Q | SYNCRIP | Human |
| anti_4230_1 | Eukaryotic translation initiation factor 4 gamma 2 | EIF4G2 | Human |
| anti_4232_19 | Insulin-like growth factor 1 receptor | IGF1R | Human |
| anti_4238_4 | Protein lin-7 homolog B | LIN7B | Human |
| anti_4245_80 | E3 ubiquitin-protein ligase Mdm2 | MDM2 | Human |
| anti_4249_64 | Nucleoside diphosphate kinase B | NME2 | Human |
| anti_4250_23 | NSFL1 cofactor p47 | NSFL1C | Human |
| anti_4254_6 | NudC domain-containing protein 3 | NUDCD3 | Human |
| anti_4261_55 | Serum paraoxonase/arylesterase 1 | PON1 | Human |
| anti_4267_81 | Pescadillo homolog | PES 1 | Human |
| anti_4271_75 | Prefoldin subunit 5 | PFDN5 | Human |
| anti_4278_14 | Protein disulfide-isomerase | P4HB | Human |
| anti_4280_47 | Proteasome subunit alpha type-2 | PSMA2 | Human |
| anti_4284_18 | RNA-binding protein 39 | RBM39 | Human |
| anti_4294_16 | Sphingosine kinase 1 | SPHK1 | Human |
| anti_4301_58 | Thymidine kinase, cytosolic | TK1 | Human |
| anti_4304_18 | Ligand-dependent nuclear receptor corepressor-like protein | LCORL | Human |
| anti_4318_12 | Tyrosine-protein phosphatase non-receptor type 6 | PTPN6 | Human |
| anti_4322_28 | Protein amnionless | AMN | Human |
| anti_4324_33 | Cystatin-SA | CST2 | Human |
| anti_4328_2 | Brother of CDO | BOC | Human |
| anti_4330_4 | Prostate-specific antigen | KLK3 | Human |
| anti_4383_97 | Inhibin beta A chain:Inhibin beta B chain heterodimer | INHBA\|INHBB | Human |
| anti_4389_2 | Desert hedgehog protein N-product | DHH | Human |
| anti_4392_54 | Fibroblast growth factor 12 | FGF12 | Human |
| anti_4393_3 | Fibroblast growth factor 16 | FGF16 | Human |
| anti_4394_71 | Fibroblast growth factor 8 isoform A | FGF8 | Human |
| anti_4396_54 | Interferon lambda-1 | IFNL1 | Human |
| anti_4397_26 | Interferon lambda-2 | IFNL2 | Human |
| anti_4420_7 | Disintegrin and metalloproteinase domain-containing protein 12 | ADAM12 | Human |
| anti_4423_77 | Bcl-2-like protein 1 | BCL2L1 | Human |
| anti_4428_1 | Carbohydrate sulfotransferase 2 | CHST2 | Human |
| anti_4429_51 | Carbohydrate sulfotransferase 6 | CHST6 | Human |
| anti_4435_66 | Ectonucleotide pyrophosphatase/ phosphodiesterase family member 7 | ENPP7 | Human |
| anti_4436_1 | Ectonucleoside triphosphate diphosphohydrolase 3 | ENTPD3 | Human |
| anti_4440_15 | Fc receptor-like protein 3 | FCRL3 | Human |
| anti_4449_67 | Gremlin-1 | GREM1 | Human |
| anti_4452_9 | Leucine-rich repeat transmembrane neuronal protein 1 | LRRTM1 | Human |
| anti_4453_83 | Leucine-rich repeat transmembrane neuronal protein 3 | LRRTM3 | Human |
| anti_4455_89 | Lactadherin | MFGE8 | Human |

| SOMAmer | Target Full Name | Entrez Gene Symbol | Organism |
|---|---|---|---|
| anti_4459_68 | Proprotein convertase subtilisin/kexin type 7 | PCSK7 | Human |
| anti_4460_8 | 3-phosphoinositide-dependent protein kinase 1 | PDPK1 | Human |
| anti_4464_10 | Sialoadhesin | SIGLEC1 | Human |
| anti_4468_21 | Sphingosine kinase 2 | SPHK2 | Human |
| anti_4471_50 | Protein-glutamine gamma-glutamyltransferase E | TGM3 | Human |
| anti_4472_5 | Tropomyosin beta chain | TPM2 | Human |
| anti_4475_60 | Tyrosine-protein kinase ZAP-70 | ZAP70 | Human |
| anti_4480_59 | Complement C3b | C3 | Human |
| anti_4487_1 | Fibroblast growth factor 7 | FGF7 | Human |
| anti_4490_65 | Interleukin-3 receptor subunit alpha | IL3RA | Human |
| anti_4491_4 | Interleukin-5 receptor subunit alpha | IL5RA | Human |
| anti_4493_92 | Interleukin-11 | IL11 | Human |
| anti_4494_63 | Interleukin-23 | IL12B\|IL23A | Human |
| anti_4496_60 | Macrophage metalloelastase | MMP12 | Human |
| anti_4503_73 | Clumping factor A_STAAW | clfA MW0764 | strain MW2 |
| anti_4504_43 | Clumping factor B_STAAE | clfB NWMN_2529 | strain Newman |
| anti_4508_14 | Beta-lactam-inducible penicillin-binding protein_STAAU | pbp | Staphylococcus aureus |
| anti_4509_52 | Beta-lactamase_STAAU | blaZ | Staphylococcus aureus |
| anti_4510_7 | Clumping factor A_STAAW | clfA MW0764 | strain MW2 |
| anti_4511_72 | Clumping factor B_STAAE | clfB NWMN_2529 | strain Newman |
| anti_4520_8 | Immunoglobulin G-binding protein A_STAAU | spa | Staphylococcus aureus |
| anti_4521_49 | Beta-lactamase_STAAU | blaZ | Staphylococcus aureus |
| anti_4522_5 | Clumping factor A_STAAW | clfA MW0764 | strain MW2 |
| anti_4523_59 | Clumping factor B_STAAE | clfB NWMN_2529 | strain Newman |
| anti_4530_2 | Beta-lactam-inducible penicillin-binding protein_STAAU | pbp | Staphylococcus aureus |
| anti_4531_56 | Immunoglobulin G-binding protein A_STAAU | spa | Staphylococcus aureus |
| anti_4533_76 | A disintegrin and metalloproteinase with thrombospondin motifs 15 | ADAMTS15 | Human |
| anti_4534_10 | Brain-specific serine protease 4 | PRSS22 | Human |
| anti_4535_50 | ADP-ribosyl cyclase/cyclic ADP-ribose hydrolase 2 | BST1 | Human |
| anti_4540_11 | Chromobox protein homolog 5 | CBX5 | Human |
| anti_4543_65 | Collagen alpha-1(XXIII) chain | COL23A1 | Human |
| anti_4545_53 | Mitochondrial import inner membrane translocase subunit TIM14 | DNAJC19 | Human |
| anti_4547_59 | Leucine-rich repeat transmembrane protein FLRT1 | FLRT1 | Human |
| anti_4548_4 | Galactoside 3(4)-L-fucosyltransferase | FUT3 | Human |
| anti_4549_78 | Alpha-(1,3)-fucosyltransferase 5 | FUT5 | Human |
| anti_4550_3 | Tyrosine-protein kinase Fyn | FYN | Human |
| anti_4551_72 | Adhesion G-protein coupled receptor G5 | ADGRG5 | Human |
| anti_4553_65 | Hepatoma-derived growth factor-related protein 2 | HDGFL2 | Human |
| anti_4556_10 | Interleukin-34 | IL34 | Human |
| anti_4557_61 | Kin of IRRE-like protein 3 | KIRREL3 | Human |
| anti_4559_64 | Kynureninase | KYNU | Human |
| anti_4560_34 | Tyrosine-protein kinase Lck | LCK | Human |
| anti_4561_65 | Baculoviral IAP repeat-containing protein 7 Isoform beta | BIRC7 | Human |
| anti_4563_61 | 1-phosphatidylinositol 4,5-bisphosphate phosphodiesterase gamma-1 | PLCG1 | Human |
| anti_4566_24 | R-spondin-2 | RSPO2 | Human |
| anti_4568_17 | SLIT and NTRK-like protein 5 | SLITRK5 | Human |
| anti_4569_52 | VPS10 domain-containing receptor SorCS2 | SORCS2 | Human |
| anti_4584_5 | Melittin_VESMG | MELT | Hornet |
| anti_4588_1 | Pancreatic hormone | PPY | Human |
| anti_4594_22 | Pituitary adenylate cyclase-activating polypeptide 38 | ADCYAP1 | Human |
| anti_4673_13 | Interleukin-6 | IL6 | Human |
| anti_4693_72 | 3-hydroxyisobutyrate dehydrogenase, mitochondrial | HIBADH | Human |

| SOMAmer | Target Full Name | Entrez Gene Symbol | Organism |
|---|---|---|---|
| anti_4695_49 | alpha-S1-casein | CSN1S1 | Human |
| anti_4696_2 | Fatty acid-binding protein, heart | FABP3 | Human |
| anti_4697_59 | Granulocyte-macrophage colony-stimulating factor | CSF2 | Human |
| anti_4703_87 | Lymphotoxin-alpha | LTA | Human |
| anti_4717_55 | Interleukin-3 | IL3 | Human |
| anti_4726_3 | Fibronectin-binding protein A_STAA3 | fnbA | Staphylococcus aureus |
| anti_4727_62 | Iron-regulated surface determinant protein A_STAAR | isdA frpA stbA SAR1103 | strain MRSA252 |
| anti_4734_16 | Gamma-hemolysin component C_STAAU | hlgC | Staphylococcus aureus |
| anti_4735_54 | Leukocidin-S subunit_STAAU | lukS | Staphylococcus aureus |
| anti_4736_37 | Leukocidin-F subunit_STAAU | lukF | Staphylococcus aureus |
| anti_4745_51 | Fibronectin-binding protein A_STAA3 | fnbA | Staphylococcus aureus |
| anti_4746_3 | Iron-regulated surface determinant protein A_STAAR | isdA frpA stbA SAR1103 | strain MRSA252 |
| anti_4752_53 | Gamma-hemolysin component C_STAAU | hlgC | Staphylococcus aureus |
| anti_4753_16 | Leukocidin-S subunit_STAAU | lukS | Staphylococcus aureus |
| anti_4754_96 | Leukocidin-F subunit_STAAU | lukF | Staphylococcus aureus |
| anti_4758_6 | ADP-ribosyltransferase enzymatic component_CLODI | cdtA | Clostridium difficile |
| anti_4769_10 | Olfactomedin-4 | OLFM4 | Human |
| anti_4774_62 | Protein FAM107B | FAM107B | Human |
| anti_4785_30 | Corticosteroid-binding globulin | SERPINA6 | Human |
| anti_4801_13 | Lactoperoxidase | LPO | Human |
| anti_4807_13 | Collagen alpha-1(VIII) chain | COL8A1 | Human |
| anti_4832_75 | Tumor necrosis factor receptor superfamily member 10A | TNFRSF10A | Human |
| anti_4834_61 | Ephrin type-A receptor 2 | EPHA2 | Human |
| anti_4840_73 | Granulocyte colony-stimulating factor | CSF3 | Human |
| anti_4842_62 | Glypican-3 | GPC3 | Human |
| anti_4859_6 | Bone morphogenetic protein receptor type-1A | BMPR1A | Human |
| anti_4862_63 | Bone morphogenetic protein receptor type-2 | BMPR2 | Human |
| anti_4867_15 | Vascular endothelial growth factor A, isoform 121 | VEGFA | Human |
| anti_4880_21 | Growth/differentiation factor 2 | GDF2 | Human |
| anti_4883_56 | Insulin | INS | Human |
| anti_4886_3 | C-C motif chemokine 7 | CCL7 | Human |
| anti_4889_82 | Protein Wnt-7a | WNT7A | Human |
| anti_4890_10 | Corticotropin | POMC | Human |
| anti_4891_50 | Glucagon | GCG | Human |
| anti_4904_7 | Caspase-2 | CASP2 | Human |
| anti_4908_6 | Endoglin | ENG | Human |
| anti_4909_68 | Galectin-8 | LGALS8 | Human |
| anti_4910_21 | Phospholipase A2 | PLA2G1B | Human |
| anti_4914_10 | Human Chorionic Gonadotropin | CGA\|CGB3\|CGB7 | Human |
| anti_4917_62 | Integrin alpha-V: beta-5 complex | ITGAV\|ITGB5 | Human |
| anti_4922_13 | C-C motif chemokine 19 | CCL19 | Human |
| anti_4925_54 | Collagenase 3 | MMP13 | Human |
| anti_4931_59 | Tissue Factor | F3 | Human |
| anti_4937_55 | Toxin B_CLODI | tcdB toxB | Clostridium difficile |
| anti_4939_280 | Toxin A_CLODI | tcdA toxA | Clostridium difficile |
| anti_4940_23 | Toxin B_CLODI | tcdB toxB | Clostridium difficile |
| anti_4942_207 | Heat-labile enterotoxin A chain_ECOLX | non-human | Escherichia coli |
| anti_4943_51 | Toxin A_CLODI | tcdA toxA | Clostridium difficile |

| SOMAmer | Target Full Name | Entrez Gene Symbol | Organism |
|---|---|---|---|
| anti_4948_1 | Antigen 85-A_MYCTU | FCN1 FCNM | Mycobacterium tuberculosis |
| anti_4949_52 | Antigen 85-B_MYCTU | fbpB Rv1886c MTCY180.32 | Mycobacterium tuberculosis |
| anti_4950_27 | Antigen 85-C_MYCTU | fbpC mpt45 Rv0129c MTCI5.03c | Mycobacterium tuberculosis |
| anti_4951_88 | Shiga-like toxin 1 subunit B_BPH19 | non-human | Bacteriophage H19B |
| anti_4953_64 | Antigen 85-A_MYCTU | FCN1 FCNM | Mycobacterium tuberculosis |
| anti_4954_5 | Antigen 85-B_MYCTU | fbpB Rv1886c MTCY180.32 | Mycobacterium tuberculosis |
| anti_4955_49 | Antigen 85-C_MYCTU | fbpC mpt45 Rv0129c MTCI5.03c | Mycobacterium tuberculosis |
| anti_4956_2 | Epiregulin | EREG | Human |
| anti_4961_17 | Annexin A2 | ANXA2 | Human |
| anti_4962_52 | Cerebral dopamine neurotrophic factor | CDNF | Human |
| anti_4963_19 | cAMP-regulated phosphoprotein 19 | ARPP19 | Human |
| anti_4968_50 | Macrophage-capping protein | CAPG | Human |
| anti_4970_55 | Carbonic anhydrase 2 | CA2 | Human |
| anti_4973_18 | Baculoviral IAP repeat-containing protein 3 | BIRC3 | Human |
| anti_4976_57 | Adapter molecule crk | CRK | Human |
| anti_4981_6 | Desmocollin-3 | DSC3 | Human |
| anti_4986_59 | Focal adhesion kinase 1 | PTK2 | Human |
| anti_4987_17 | Immunoglobulin alpha Fc receptor | FCAR | Human |
| anti_4991_12 | Glypican-5 | GPC5 | Human |
| anti_4993_16 | Glutathione S-transferase A3 | GSTA3 | Human |
| anti_4994_178 | Heterogeneous nuclear ribonucleoprotein K | HNRNPK | Human |
| anti_4997_19 | Eukaryotic initiation factor 4A-III | EIF4A3 | Human |
| anti_5002_76 | Matrix metalloproteinase-14 | MMP14 | Human |
| anti_5004_69 | Mitogen-activated protein kinase 11 | MAPK11 | Human |
| anti_5005_4 | Mitogen-activated protein kinase 12 | MAPK12 | Human |
| anti_5006_71 | Mitogen-activated protein kinase 13 | MAPK13 | Human |
| anti_5009_11 | Moesin | MSN | Human |
| anti_5011_11 | Nicotinamide phosphoribosyltransferase | NAMPT | Human |
| anti_5014_49 | Cytoplasmic protein NCK1 | NCK1 | Human |
| anti_5016_61 | Protein DJ-1 | PARK7 | Human |
| anti_5017_19 | Peroxiredoxin-5, mitochondrial | PRDX5 | Human |
| anti_5024_67 | Retinoblastoma-associated protein | RB1 | Human |
| anti_5026_66 | 40S ribosomal protein S3 | RPS3 | Human |
| anti_5031_10 | Spectrin alpha chain, non-erythrocytic 1 | SPTAN1 | Human |
| anti_5032_64 | FACT complex subunit SSRP1 | SSRP1 | Human |
| anti_5033_27 | Tropomyosin alpha-1 chain | TPM1 | Human |
| anti_5035_7 | Thymidylate synthase | TYMS | Human |
| anti_5036_50 | Tumor necrosis factor-inducible gene 6 protein | TNFAIP6 | Human |
| anti_5060_62 | Programmed cell death 1 ligand 1 | CD274 | Human |
| anti_5062_60 | CD226 antigen | CD226 | Human |
| anti_5063_12 | Natural killer cell receptor 2B4 | CD244 | Human |
| anti_5065_8 | CD83 antigen | CD83 | Human |
| anti_5068_54 | Cytotoxic and regulatory T-cell molecule | CRTAM | Human |
| anti_5070_76 | Tumor necrosis factor receptor superfamily member 6B | TNFRSF6B | Human |
| anti_5076_53 | Ephrin type-A receptor 10 | EPHA10 | Human |
| anti_5085_18 | Interleukin-20 receptor subunit alpha | IL20RA | Human |
| anti_5087_5 | Interleukin-22 receptor subunit alpha-2 | IL22RA2 | Human |
| anti_5088_175 | Interleukin-23 receptor | IL23R | Human |
| anti_5089_11 | Interleukin-7 receptor subunit alpha | IL7R | Human |
| anti_5093_47 | Protein jagged-2 | JAG2 | Human |
| anti_5094_62 | Junctional adhesion molecule-like | JAML | Human |
| anti_5095_7 | Killer cell immunoglobulin-like receptor 2DL4 | KIR2DL4 | Human |
| anti_5096_51 | Killer cell immunoglobulin-like receptor 3DL2 | KIR3DL2 | Human |
| anti_5097_14 | Killer cell immunoglobulin-like receptor 3DS1 | KIR3DS1 | Human |
| anti_5098_79 | Killer cell lectin-like receptor subfamily F member 1 | KLRF1 | Human |
| anti_5099_14 | Lymphocyte activation gene 3 protein | LAG3 | Human |

| SOMAmer | Target Full Name | Entrez Gene Symbol | Organism |
|---|---|---|---|
| anti_5100_53 | Lysosome membrane protein 2 | SCARB2 | Human |
| anti_5102_55 | MHC class I polypeptide-related sequence B | MICB | Human |
| anti_5103_30 | Cell surface glycoprotein CD200 receptor 1 | CD200R1 | Human |
| anti_5106_52 | Neurogenic locus notch homolog protein 2 | NOTCH2 | Human |
| anti_5109_24 | Neuronal cell adhesion molecule | NRCAM | Human |
| anti_5110_84 | Neurexin-1-beta | NRXN1 | Human |
| anti_5112_73 | OX-2 membrane glycoprotein | CD200 | Human |
| anti_5115_31 | Tumor necrosis factor receptor superfamily member 19L | RELT | Human |
| anti_5117_14 | Roundabout homolog 3 | ROBO3 | Human |
| anti_5121_3 | Semaphorin-6B | SEMA6B | Human |
| anti_5122_92 | Semaphorin-6A | SEMA6A | Human |
| anti_5124_62 | Intercellular adhesion molecule 5 | ICAM5 | Human |
| anti_5128_53 | SLAM family member 6 | SLAMF6 | Human |
| anti_5130_67 | Scavenger receptor class F member 2 | SCARF2 | Human |
| anti_5131_15 | Tumor necrosis factor receptor superfamily member 19 | TNFRSF19 | Human |
| anti_5133_17 | TGF-beta receptor type-2 | TGFBR2 | Human |
| anti_5134_52 | Hepatitis A virus cellular receptor 2 | HAVCR2 | Human |
| anti_5138_50 | Tumor necrosis factor receptor superfamily member 12A | TNFRSF12A | Human |
| anti_5140_56 | Netrin receptor UNC5D | UNC5D | Human |
| anti_5178_5 | High affinity cAMP-specific 3',5'-cyclic phosphodiesterase 7A | PDE7A | Human |
| anti_5183_53 | AMP Kinase (alpha1beta1gamma1) | PRKAA1\|PRKAB1\|PRKAG1 | Human |
| anti_5193_51 | GTPase KRas | KRAS | Human |
| anti_5196_7 | Glycylpeptide N-tetradecanoyltransferase 1 | NMT1 | Human |
| anti_5201_50 | High affinity cGMP-specific 3',5'-cyclic phosphodiesterase 9A | PDE9A | Human |
| anti_5202_4 | Peptidyl-prolyl cis-trans isomerase D | PPID | Human |
| anti_5204_13 | Proteasome activator complex subunit 3 | PSME3 | Human |
| anti_5215_53 | Glucokinase Regulatory Protein_MACFA | Unknown | Cynomolgus monkey |
| anti_5216_2 | Proprotein convertase subtilisin/kexin type 9_MOUSE | non-human | Mouse |
| anti_5223_59 | Glucokinase regulatory protein | GCKR | Human |
| anti_5224_20 | Casein kinase II subunit alpha | CSNK2A1 | Human |
| anti_5225_50 | Casein kinase II 2-alpha:2-beta heterotetramer | CSNK2A1\|CSNK2B | Human |
| anti_5226_36 | Casein kinase II 2-alpha':2-beta heterotetramer | CSNK2A2\|CSNK2B | Human |
| anti_5228_25 | Kinesin-like protein KIF23 | KIF23 | Human |
| anti_5229_90 | Inosine-5'-monophosphate dehydrogenase 1 | IMPDH1 | Human |
| anti_5230_99 | 3-hydroxy-3-methylglutaryl-coenzyme A reductase | HMGCR | Human |
| anti_5236_2 | Nuclear receptor subfamily 1 group D member 1 | NR1D1 | Human |
| anti_5237_55 | Peptidyl-prolyl cis-trans isomerase F, mitochondrial_MOUSE | non-human | Mouse |
| anti_5238_26 | Peptidyl-prolyl cis-trans isomerase E | PPIE | Human |
| anti_5244_12 | Mitogen-activated protein kinase 9 | MAPK9 | Human |
| anti_5245_40 | AMP Kinase (alpha2beta2gamma1) | PRKAA2\|PRKAB2\|PRKAG1 | Human |
| anti_5248_68 | Peptidyl-prolyl cis-trans isomerase F, mitochondrial | PPIF | Human |
| anti_5253_1 | Calcium/calmodulin-dependent 3',5'-cyclic nucleotide phosphodiesterase 1A | PDE1A | Human |
| anti_5265_12 | GRB2-related adapter protein 2 | GRAP2 | Human |
| anti_5268_49 | Matrix metalloproteinase-16 | MMP16 | Human |
| anti_5271_5 | Ras-related C3 botulinum toxin substrate 3 | RAC3 | Human |
| anti_5275_28 | Proto-oncogene vav | VAV1 | Human |
| anti_5280_68 | Mitochondrial glutamate carrier 2 | SLC25A18 | Human |
| anti_5328_33 | Epidermal growth factor receptor variant III | EGFR | Human |
| anti_5337_64 | T-lymphocyte activation antigen CD86 | CD86 | Human |
| anti_5339_49 | Protein S100-A9 | S100A9 | Human |
| anti_5343_74 | Carboxypeptidase E | CPE | Human |
| anti_5347_59 | G2/mitotic-specific cyclin-B1 | CCNB1 | Human |

| SOMAmer | Target Full Name | Entrez Gene Symbol | Organism |
|---|---|---|---|
| anti_5352_11 | Tumor necrosis factor receptor superfamily member 14 | TNFRSF14 | Human |
| anti_5354_11 | Keratin, type I cytoskeletal 18 | KRT18 | Human |
| anti_5355_69 | Tumor necrosis factor ligand superfamily member 14 | TNFSF14 | Human |
| anti_5357_60 | Neuroligin-4, X-linked | NLGN4X | Human |
| anti_5359_65 | Serine/threonine-protein kinase pim-1 | PIM1 | Human |
| anti_5363_51 | Semaphorin-3E | SEMA3E | Human |
| anti_5430_66 | Tyrosine-protein phosphatase non-receptor type substrate 1 | SIRPA | Human |
| anti_5441_67 | Troponin I, cardiac muscle | TNNI3 | Human |
| anti_5443_62 | Atrial natriuretic factor | NPPA | Human |
| anti_5457_5 | Collectin-12 | COLEC12 | Human |
| anti_5460_60 | ATP-dependent RNA helicase DDX19B | DDX19B | Human |
| anti_5468_67 | Interleukin-17 receptor C | IL17RC | Human |
| anti_5475_10 | Protein kinase C beta type (splice variant beta-II) | PRKCB | Human |
| anti_5476_66 | Protein kinase C gamma type | PRKCG | Human |
| anti_5478_50 | Glutamate carboxypeptidase 2 | FOLH1 | Human |
| anti_5481_16 | Ras GTPase-activating protein 1 | RASA1 | Human |
| anti_5484_63 | 40S ribosomal protein S3a | RPS3A | Human |
| anti_5488_74 | Proto-oncogene tyrosine-protein kinase Src isoform 2 | SRC | Human |
| anti_5526_53 | Tumor necrosis factor receptor superfamily member 18 | TNFRSF18 | Human |
| anti_5534_49 | Tumor necrosis factor receptor superfamily member 10B | TNFRSF10B | Human |
| anti_5557_2 | ESAT-6-like protein esxB_MYCTU | clpC1 Rv3596c MTCY07H7B.26 | Mycobacterium tuberculosis |
| anti_5558_86 | Phosphate-binding protein pstS 1_MYCTU | pstS1 | Mycobacterium tuberculosis |
| anti_5560_59 | MPT51/MPB51 antigen_MYCTU | mpt51 | Mycobacterium tuberculosis |
| anti_5562_95 | ESAT-6-like protein esxB_MYCTU | clpC1 Rv3596c MTCY07H7B.26 | Mycobacterium tuberculosis |
| anti_5564_89 | Toxin A_CLODI | tcdA toxA | Clostridium difficile |
| anti_5569_2 | Antigen 85-C_MYCTU | fbpC mpt45 Rv0129c MTCI5.03c | Mycobacterium tuberculosis |
| anti_5573_4 | Toxin B_CLODI | tcdB toxB | Clostridium difficile |
| anti_5575_1 | Antigen 85-C_MYCTU | fbpC mpt45 Rv0129c MTCI5.03c | Mycobacterium tuberculosis |
| anti_5579_11 | ADP-ribosyltransferase enzymatic component_CLODI | cdtA | Clostridium difficile |
| anti_5583_62 | Tyrosine-protein kinase RYK | RYK | Human |
| anti_5587_3 | Coiled-coil domain-containing protein 134 | CCDC134 | Human |
| anti_5590_11 | Zona pellucida-like domain-containing protein 1 | ZPLD1 | Human |
| anti_5593_11 | Protein disulfide-isomerase A5 | PDIA5 | Human |
| anti_5594_87 | LDLR chaperone MESD | MESD | Human |
| anti_5598_3 | Gremlin-2 | GREM2 | Human |
| anti_5602_62 | Inactive ribonuclease-like protein 10 | RNASE10 | Human |
| anti_5604_30 | Heparanase | HPSE | Human |
| anti_5606_24 | Microfibrillar-associated protein 1 | MFAP1 | Human |
| anti_5609_92 | Protein FAM19A5 | TAFA5 | Human |
| anti_5610_32 | Protein FAM171A1 | FAM171A1 | Human |
| anti_5611_56 | Beta-defensin 108B | DEFB108B | Human |
| anti_5612_16 | Histidine triad nucleotide-binding protein 2, mitochondrial | HINT2 | Human |
| anti_5613_75 | Linker for activation of T-cells family member 2 | LAT2 | Human |
| anti_5614_44 | Corticoliberin | CRH | Human |
| anti_5615_62 | Protein FAM172A | FAM172A | Human |
| anti_5621_64 | Thrombospondin type-1 domain-containing protein 1 | THSD1 | Human |
| anti_5623_11 | CMRF35-like molecule 1 | CD300LF | Human |
| anti_5624_66 | Folate receptor gamma | FOLR3 | Human |

| SOMAmer | Target Full Name | Entrez Gene Symbol | Organism |
|---|---|---|---|
| anti_5626_20 | Chymotrypsin-C | CTRC | Human |
| anti_5627_53 | Progonadoliberin-1 | GNRH1 | Human |
| anti_5629_58 | NKG2-A/NKG2-B type II integral membrane protein | KLRC1 | Human |
| anti_5630_48 | CMRF35-like molecule 8 | CD300A | Human |
| anti_5631_83 | Promotilin | MLN | Human |
| anti_5633_65 | Leukocyte immunoglobulin-like receptor subfamily B member 2 | LILRB2 | Human |
| anti_5634_39 | GDP-fucose protein O-fucosyltransferase 1 | POFUT1 | Human |
| anti_5637_81 | Netrin-G1 | NTNG1 | Human |
| anti_5639_49 | Neuropilin and tolloid-like protein 1 | NETO1 | Human |
| anti_5643_2 | Epididymal-specific lipocalin-8 | LCN8 | Human |
| anti_5647_51 | Islet amyloid polypeptide | IAPP | Human |
| anti_5649_83 | Pregnancy-specific beta-1-glycoprotein 4 | PSG4 | Human |
| anti_5651_50 | Putative pregnancy-specific beta-1-glycoprotein 7 | PSG7 | Human |
| anti_5653_23 | Probable inactive serine protease 37 | PRSS37 | Human |
| anti_5656_53 | Poly(U)-specific endoribonuclease | ENDOU | Human |
| anti_5657_28 | CMP-N-acetylneuraminate-beta-galactosamide-alpha-2,3-sialyltransferase 1 | ST3GAL1 | Human |
| anti_5659_11 | Pro-Thyrotropin-releasing hormone | TRH | Human |
| anti_5661_15 | Interleukin-18 | IL18 | Human |
| anti_5663_18 | Platelet factor 4 variant | PF4V1 | Human |
| anti_5664_57 | Beta-defensin 106 | DEFB106A | Human |
| anti_5666_64 | Interleukin-1 receptor type 2 | IL1R2 | Human |
| anti_5667_3 | Semaphorin-3B | SEMA3B | Human |
| anti_5668_49 | Anterior gradient protein 3 | AGR3 | Human |
| anti_5669_26 | Signal-regulatory protein beta-2 | SIRPB2 | Human |
| anti_5671_1 | Chymotrypsinogen B | CTRB1 | Human |
| anti_5676_54 | Agouti-signaling protein | ASIP | Human |
| anti_5677_15 | Uncharacterized protein C2orf66 | C2orf66 | Human |
| anti_5679_16 | Beta-defensin 103 | DEFB103A | Human |
| anti_5680_54 | Odorant-binding protein 2b | OBP2B | Human |
| anti_5687_5 | Glucosidase 2 subunit beta | PRKCSH | Human |
| anti_5689_1 | Beta-defensin 112 | DEFB112 | Human |
| anti_5692_79 | Tumor necrosis factor | TNF | Human |
| anti_5693_6 | Lithostathine-1-beta | REG1B | Human |
| anti_5695_5 | BPI fold-containing family A member 2 | BPIFA2 | Human |
| anti_5699_19 | Protein FAM189A2 | FAM189A2 | Human |
| anti_5703_26 | Noelin | OLFM1 | Human |
| anti_5704_74 | Granzyme M | GZMM | Human |
| anti_5707_55 | ETS domain-containing protein Elk-3 | ELK3 | Human |
| anti_5713_9 | Interferon lambda-3 | IFNL3 | Human |
| anti_5714_88 | Interferon alpha-6 | IFNA6 | Human |
| anti_5715_4 | Protein ABHD14A | ABHD14A | Human |
| anti_5716_49 | Putative chondrosarcoma-associated gene 1 protein | CSAG1 | Human |
| anti_5717_2 | Cartilage intermediate layer protein 1 | CILP | Human |
| anti_5719_66 | Protein FAM189A2 | FAM189A2 | Human |
| anti_5721_1 | CD226 antigen | CD226 | Human |
| anti_5723_4 | Insulin-like 3 | INSL3 | Human |
| anti_5724_58 | Suprabasin | SBSN | Human |
| anti_5726_49 | Sperm-associated antigen 11A | SPAG11A | Human |
| anti_5727_35 | Beta-1,3-glucosyltransferase | B3GLCT | Human |
| anti_5728_60 | Fc receptor-like protein 1 | FCRL1 | Human |
| anti_5729_27 | Sclerostin | SOST | Human |
| anti_5730_60 | C-X-C motif chemokine 14 | CXCL14 | Human |
| anti_5731_1 | Serine protease inhibitor Kazal-type 6 | SPINK6 | Human |
| anti_5733_61 | Interferon alpha-10 | IFNA10 | Human |
| anti_5734_13 | Nectin-4 | NECTIN4 | Human |
| anti_5737_61 | Semaphorin-4D | SEMA4D | Human |
| anti_5740_17 | Roundabout homolog 1 | ROBO1 | Human |
| anti_5742_14 | Lysophosphatidic acid phosphatase type 6 | ACP6 | Human |
| anti_5743_82 | Cocaine- and amphetamine-regulated transcript protein | CARTPT | Human |
| anti_5744_12 | Protein MENT | MENT | Human |

| SOMAmer | Target Full Name | Entrez Gene Symbol | Organism |
|---|---|---|---|
| anti_5745_64 | Peptidase inhibitor 15 | PI15 | Human |
| anti_5746_37 | Ephrin-B1 | EFNB1 | Human |
| anti_5747_67 | Osteocalcin | BGLAP | Human |
| anti_5748_20 | Acid ceramidase | ASAH1 | Human |
| anti_5749_53 | Colipase | CLPS | Human |
| anti_5751_14 | LETM1 domain-containing protein 1 | LETMD1 | Human |
| anti_5752_63 | Sushi domain-containing protein 3 | SUSD3 | Human |
| anti_5753_21 | Protein FAM19A1 | TAFA1 | Human |
| anti_5754_76 | Insulin-like peptide INSL6 | INSL6 | Human |
| anti_5755_29 | Gastric inhibitory polypeptide | GIP | Human |
| anti_5756_66 | Pappalysin-2 | PAPPA2 | Human |
| anti_5757_45 | Neuroendocrine secretory protein 55 | GNAS | Human |
| anti_5758_49 | Kallikrein-9 | KLK9 | Human |
| anti_5759_10 | Ephrin-A3 | EFNA3 | Human |
| anti_5762_35 | Sperm-associated antigen 11B | SPAG11B | Human |
| anti_5763_67 | Beta-defensin 104 | DEFB104A | Human |
| anti_5764_4 | Polypeptide N-acetylgalactosaminyltransferase 2 | GALNT2 | Human |
| anti_5765_53 | Beta-defensin 121 | DEFB121 | Human |
| anti_5766_6 | R-spondin-4 | RSPO4 | Human |
| anti_5792_8 | alpha-Fetoprotein | AFP | Human |
| anti_5798_3 | BH3-interacting domain death agonist | BID | Human |
| anti_5801_72 | beta-nerve growth factor | NGF | Human |
| anti_5807_77 | CD70 antigen | CD70 | Human |
| anti_5810_25 | Teratocarcinoma-derived growth factor 1 | TDGF1 | Human |
| anti_5813_58 | Erythropoietin | EPO | Human |
| anti_5822_22 | Glial cell line-derived neurotrophic factor | GDNF | Human |
| anti_5825_49 | Interferon gamma receptor 1 | IFNGR1 | Human |
| anti_5834_18 | Interleukin-9 | IL9 | Human |
| anti_5837_49 | Leukemia inhibitory factor receptor | LIFR | Human |
| anti_5854_60 | Microtubule-associated protein tau | MAPT | Human |
| anti_5867_60 | Arginase-1 | ARG1 | Human |
| anti_5870_23 | Bcl2-associated agonist of cell death | BAD | Human |
| anti_5882_34 | Elongation factor 1-beta | EEF1B2 | Human |
| anti_5897_58 | Gastrin-releasing peptide | GRP | Human |
| anti_5900_11 | Histidine triad nucleotide-binding protein 1 | HINT1 | Human |
| anti_5915_58 | Peroxisomal targeting signal 1 receptor | PEX5 | Human |
| anti_5921_58 | Protein S100-A7 | S100A7 | Human |
| anti_5927_4 | WNT1-inducible-signaling pathway protein 3 | CCN6 | Human |
| anti_5930_54 | Troponin I, cardiac muscle | TNNI3 | Human |
| anti_5936_53 | Tumor necrosis factor | TNF | Human |
| anti_5939_42 | Tumor necrosis factor ligand superfamily member 12 | TNFSF12 | Human |
| anti_5947_90 | Thrombopoietin | THPO | Human |
| anti_5954_62 | Parathyroid hormone | PTH | Human |
| anti_5957_30 | Somatostatin-28 | SST | Human |
| anti_5970_61 | Leukocyte cell-derived chemotaxin-2 | LECT2 | Human |
| anti_5980_55 | BolA-like protein 3 | BOLA3 | Human |
| anti_5981_6 | ES1 protein homolog, mitochondrial | GATD3 C21orf33 GATD3A | Human |
| anti_5992_50 | T-cell surface glycoprotein CD8 alpha chain | CD8A | Human |
| anti_6003_28 | Sialidase-1 | NEU1 | Human |
| anti_6020_52 | Urotensin-2 | UTS2 | Human |
| anti_6024_68 | Carboxypeptidase E | CPE | Human |
| anti_6027_31 | Relaxin-3 | RLN3 | Human |
| anti_6035_2 | Beta-galactoside alpha-2,6-sialyltransferase 1 | ST6GAL1 | Human |
| anti_6036_78 | Ephrin type-A receptor 10 | EPHA10 | Human |
| anti_6037_6 | Fc_MOUSE | Igh | Mouse |
| anti_6048_4 | Fc_MOUSE | Igh | Mouse |
| anti_6054_6 | Neurexophilin-3 | NXPH3 | Human |
| anti_6060_2 | Prolactin-inducible protein | PIP | Human |
| anti_6070_11 | Fc_MOUSE | Igh | Mouse |
| anti_6096_23 | Collagen alpha-2(VI) chain | COL6A2 | Human |
| anti_6102_2 | Fc_MOUSE | Igh | Mouse |
| anti_6113_31 | Endothelin-3 | EDN3 | Human |
| anti_6119_14 | MAM domain-containing protein 2 | MAMDC2 | Human |

| SOMAmer | Target Full Name | Entrez Gene Symbol | Organism |
|---|---|---|---|
| anti_6121_5 | Fc_MOUSE | Igh | Mouse |
| anti_6151_18 | Dual specificity mitogen-activated protein kinase kinase 3 | MAP2K3 | Human |
| anti_6168_11 | Cellular tumor antigen p53 R175H mutant | TP53 | Human |
| anti_6207_10 | Prosaposin | PSAP | Human |
| anti_6209_2 | Amiloride-sensitive amine oxidase [copper-containing] | AOC1 | Human |
| anti_6210_100 | Interferon alpha-5 | IFNA5 | Human |
| anti_6214_84 | Interferon alpha-8 | IFNA8 | Human |
| anti_6219_14 | PDZ domain-containing protein 11 | PDZD11 | Human |
| anti_6221_1 | ADP-dependent glucokinase | ADPGK | Human |
| anti_6223_5 | Guanylate cyclase activator 2B | GUCA2B | Human |
| anti_6226_69 | Calcium uptake protein 2, mitochondrial | MICU2 | Human |
| anti_6228_58 | Sperm acrosome membrane-associated protein 1 | SPACA1 | Human |
| anti_6230_56 | Cerberus | CER1 | Human |
| anti_6231_46 | Stanniocalcin-2 | STC2 | Human |
| anti_6234_74 | Vitrin | VIT | Human |
| anti_6236_51 | Collagen triple helix repeat-containing protein 1 | CTHRC1 | Human |
| anti_6237_11 | Fibroblast growth factor receptor-like 1 | FGFRL1 | Human |
| anti_6238_55 | Pregnancy-specific beta-1-glycoprotein 8 | PSG8 | Human |
| anti_6240_55 | R-spondin-1 | RSPO1 | Human |
| anti_6240_70 | R-spondin-1 | RSPO1 | Human |
| anti_6245_4 | Nectin-2 | NECTIN2 | Human |
| anti_6247_9 | Signal-regulatory protein beta-1 | SIRPB1 | Human |
| anti_6248_68 | Tumor necrosis factor receptor superfamily member 6 | FAS | Human |
| anti_6262_14 | Interleukin-17 receptor B | IL17RB | Human |
| anti_6263_77 | Phospholipid transfer protein | PLTP | Human |
| anti_6264_9 | Protein CYR61 | CCN1 | Human |
| anti_6267_51 | Stathmin-4 | STMN4 | Human |
| anti_6274_15 | WSC domain-containing protein 2 | WSCD2 | Human |
| anti_6280_11 | Protocadherin-8 | PCDH8 | Human |
| anti_6284_7 | ALK and LTK ligand 2 | ALKAL2 | Human |
| anti_6290_3 | Urotensin-2B | UTS2B | Human |
| anti_6291_55 | Calsyntenin-3 | CLSTN3 | Human |
| anti_6294_11 | B melanoma antigen 2 | BAGE2 | Human |
| anti_6296_36 | Oncoprotein-induced transcript 3 protein | OIT3 | Human |
| anti_6315_58 | Phospholipase B-like 1 | PLBD1 | Human |
| anti_6321_65 | Protocadherin gamma-A10 | PCDHGA10 | Human |
| anti_6324_11 | Deoxyribonuclease-1-like 2 | DNASE1L2 | Human |
| anti_6326_20 | Bone morphogenetic protein 4 | BMP4 | Human |
| anti_6350_43 | Apolipoprotein C-II | APOC2 | Human |
| anti_6351_55 | B-cell antigen receptor complex-associated protein beta chain | CD79B | Human |
| anti_6352_8 | Insulin receptor-related protein | INSRR | Human |
| anti_6354_13 | Hephaestin-like protein 1 | HEPHL1 | Human |
| anti_6359_50 | Protein O-linked-mannose beta-1,4-N-acetylglucosaminyltransferase 2 | POMGNT2 | Human |
| anti_6360_7 | Beta-defensin 128 | DEFB128 | Human |
| anti_6361_49 | Receptor-type tyrosine-protein phosphatase R | PTPRR | Human |
| anti_6363_55 | Fc_MOUSE | Igh | Mouse |
| anti_6365_62 | Fc_MOUSE | Igh | Mouse |
| anti_6367_66 | fibromodulin | FMOD | Human |
| anti_6371_50 | Angiopoietin-related protein 7 | ANGPTL7 | Human |
| anti_6374_7 | Matrix metalloproteinase-20 | MMP20 | Human |
| anti_6375_75 | Xyloside xylosyltransferase 1 | XXYLT1 | Human |
| anti_6377_54 | Lysozyme-like protein 2 | LYZL2 | Human |
| anti_6383_90 | Tolloid-like protein 1 | TLL1 | Human |
| anti_6384_19 | WAP four-disulfide core domain protein 3 | WFDC3 | Human |
| anti_6385_63 | von Willebrand factor A domain-containing protein 1 | VWA1 | Human |
| anti_6395_58 | Glycoprotein hormone alpha-2 | GPHA2 | Human |
| anti_6398_12 | Lon protease homolog, mitochondrial | LONP1 | Human |
| anti_6399_52 | Beta-defensin 107 | DEFB107A | Human |
| anti_6400_33 | Tumor protein p53-inducible protein 13 | TP53I13 | Human |

| SOMAmer | Target Full Name | Entrez Gene Symbol | Organism |
|---|---|---|---|
| anti_6401_73 | Fc_MOUSE | Igh | Mouse |
| anti_6402_8 | Paired immunoglobulin-like type 2 receptor alpha isoform FDF03-deltaTM | PILRA | Human |
| anti_6404_20 | C1q-related factor | C1QL1 | Human |
| anti_6407_63 | Ladinin-1 | LAD1 | Human |
| anti_6410_26 | Early placenta insulin-like peptide | INSL4 | Human |
| anti_6411_58 | Beta-defensin 135 | DEFB135 | Human |
| anti_6412_26 | Fc_MOUSE | Igh | Mouse |
| anti_6413_79 | Lipase member K | LIPK | Human |
| anti_6416_8 | Gastrokine-2 | GKN2 | Human |
| anti_6421_52 | Interferon alpha-16 | IFNA16 | Human |
| anti_6430_36 | Protein FAM19A2 | TAFA2 | Human |
| anti_6433_57 | Pseudokinase FAM20A | FAM20A | Human |
| anti_6434_18 | Retina-specific copper amine oxidase | AOC2 | Human |
| anti_6441_62 | A disintegrin and metalloproteinase with thrombospondin motifs 6 | ADAMTS6 | Human |
| anti_6443_68 | Fc_MOUSE | Igh | Mouse |
| anti_6444_15 | Pregnancy-specific beta-1-glycoprotein 3 | PSG3 | Human |
| anti_6447_73 | Pentraxin-related protein PTX3 | PTX3 | Human |
| anti_6448_36 | Semaphorin-3C | SEMA3C | Human |
| anti_6450_8 | Gastrotropin | FABP6 | Human |
| anti_6454_38 | Rho GDP-dissociation inhibitor 1 | ARHGDIA | Human |
| anti_6455_52 | Hyaluronan and proteoglycan link protein 4 | HAPLN4 | Human |
| anti_6456_17 | Pregnancy-specific beta-1-glycoprotein 6 | PSG6 | Human |
| anti_6457_50 | Protocadherin gamma-A1 | PCDHGA1 | Human |
| anti_6467_65 | Protein O-glucosyltransferase 1 | POGLUT1 | Human |
| anti_6469_62 | Lymphocyte antigen 6 complex locus protein G6d | LY6G6D | Human |
| anti_6473_55 | BPI fold-containing family A member 1 | BPIFA1 | Human |
| anti_6478_2 | IgLON family member 5 | IGLON5 | Human |
| anti_6484_11 | Periostin | POSTN | Human |
| anti_6485_59 | Immunoglobulin lambda-like polypeptide 1 | IGLL1 | Human |
| anti_6493_9 | Carboxypeptidase Z | CPZ | Human |
| anti_6494_60 | Sperm acrosome-associated protein 5 | SPACA5 | Human |
| anti_6495_14 | Endothelin-1 | EDN1 | Human |
| anti_6497_10 | Retinoschisin | RS1 | Human |
| anti_6502_50 | Cysteine-rich motor neuron 1 protein | CRIM1 | Human |
| anti_6504_65 | Lysyl oxidase homolog 2 | LOXL2 | Human |
| anti_6507_16 | Neural cell adhesion molecule 2 | NCAM2 | Human |
| anti_6512_68 | Kremen protein 1 | KREMEN1 | Human |
| anti_6525_17 | Dual specificity protein phosphatase 13 isoform A | DUSP13 | Human |
| anti_6526_77 | Odorant-binding protein 2a | OBP2A | Human |
| anti_6528_95 | Exostosin-like 2 | EXTL2 | Human |
| anti_6530_63 | Opiorphin prepropeptide | OPRPN | Human |
| anti_6531_29 | Protein FAM162A | FAM162A | Human |
| anti_6533_20 | Deoxyribonuclease-2-beta | DNASE2B | Human |
| anti_6538_90 | Uncharacterized protein KIAA2013 | KIAA2013 | Human |
| anti_6543_182 | Prolactin-releasing peptide | PRLH | Human |
| anti_6544_33 | Protein kinase C-binding protein NELL1 | NELL1 | Human |
| anti_6546_41 | Dickkopf-related protein 2 | DKK 2 | Human |
| anti_6547_83 | Transmembrane protease serine 11D | TMPRSS11D | Human |
| anti_6549_60 | V-set and transmembrane domain-containing protein 2-like protein | VSTM2L | Human |
| anti_6553_68 | Cilia- and flagella-associated protein 45 | CFAP45 | Human |
| anti_6558_5 | Collectin-10 | COLEC10 | Human |
| anti_6563_78 | Heat shock 70 kDa protein 1A | HSPA1A | Human |
| anti_6565_68 | CUB domain-containing protein 1 | CDCP1 | Human |
| anti_6568_18 | HEPACAM family member 2 | HEPACAM2 | Human |
| anti_6570_1 | Collagen alpha-1(XIII) chain | COL13A1 | Human |
| anti_6571_75 | Uncharacterized protein C17orf78 | C17orf78 | Human |
| anti_6572_10 | Leucine-rich repeat transmembrane neuronal protein 4 | LRRTM4 | Human |
| anti_6575_79 | ADAMTS-like protein 1 | ADAMTSL1 | Human |
| anti_6577_64 | Laminin subunit alpha-4 | LAMA4 | Human |
| anti_6578_29 | Tenomodulin | TNMD | Human |
| anti_6581_50 | Fc_MOUSE | Igh | Mouse |

| SOMAmer | Target Full Name | Entrez Gene Symbol | Organism |
|---|---|---|---|
| anti_6584_1 | Sarcoplasmic reticulum histidine-rich calcium-binding protein | HRC | Human |
| anti_6593_5 | Polypeptide N-acetylgalactosaminyltransferase 3 | GALNT3 | Human |
| anti_6594_64 | SAYSvFN domain-containing protein 1 | SAYSD1 | Human |
| anti_6597_24 | Membrane protein FAM174A | FAM174A | Human |
| anti_6599_5 | Protein APCDD1 | APCDD1 | Human |
| anti_6600_70 | RELT-like protein 2 | RELL2 | Human |
| anti_6609_22 | 2',3'-cyclic-nucleotide 3'-phosphodiesterase | CNP | Human |
| anti_6611_8 | KDEL motif-containing protein 1 | POGLUT2 | Human |
| anti_6622_90 | Apelin | APLN | Human |
| anti_6624_94 | Protein Wnt-11 | WNT11 | Human |
| anti_6625_31 | ATPase family AAA domain-containing protein 1 | ATAD1 | Human |
| anti_6627_25 | Inactive pancreatic lipase-related protein 1 | PNLIPRP1 | Human |
| anti_6633_43 | Uroplakin-3b-like protein | UPK3BL1 UPLP | Human |
| anti_6634_4 | Netrin-1 | NTN1 | Human |
| anti_6643_62 | Leukocyte surface antigen CD47 | CD47 | Human |
| anti_6649_51 | Netrin-1 | NTN1 | Human |
| anti_6650_20 | Periostin | POSTN | Human |
| anti_6653_58 | Leukocyte surface antigen CD47 | CD47 | Human |
| anti_6706_18 | Alkaline phosphatase, placental-like | ALPG | Human |
| anti_6713_4 | Low-density lipoprotein receptor-related protein 11 | LRP11 | Human |
| anti_6715_63 | Alkaline phosphatase, placental-like | ALPG | Human |
| anti_6895_1 | Transferrin receptor protein 1 | TFRC | Human |
| anti_6897_38 | Galactosylgalactosylxylosylprotein 3-beta-glucuronosyltransferase 3 | B3GAT3 | Human |
| anti_6899_37 | Pterin-4-alpha-carbinolamine dehydratase 2 | PCBD2 | Human |
| anti_6904_14 | Leucine-rich repeat transmembrane neuronal protein 2 | LRRTM2 | Human |
| anti_6907_17 | Transmembrane and coiled-coil domain-containing protein 5A | TMCO5A | Human |
| anti_6911_103 | C-type lectin domain family 6 member A | CLEC6A | Human |
| anti_6912_6 | Calcium uptake protein 3, mitochondrial | MICU3 | Human |
| anti_6913_189 | High mobility group protein B2 | HMGB2 | Human |
| anti_6915_2 | CD164 sialomucin-like 2 protein | CD164L2 | Human |
| anti_6917_49 | Leucine-rich repeat-containing protein 3 | LRRC3 | Human |
| anti_6918_183 | Cholecystokinin | CCK | Human |
| anti_6920_1 | GDNF family receptor alpha-like | GFRAL | Human |
| anti_6921_24 | Beta-1,4-galactosyltransferase 3 | B4GALT3 | Human |
| anti_6923_1 | Procollagen-lysine,2-oxoglutarate 5-dioxygenase 2 | PLOD2 | Human |
| anti_6925_26 | Sorting nexin-8 | SNX8 | Human |
| anti_6929_10 | Sia-alpha-2,3-Gal-beta-1,4-GlcNAc-R:alpha 2,8-sialyltransferase | ST8SIA3 | Human |
| anti_6930_95 | Alpha-2,8-sialyltransferase 8F | ST8SIA6 | Human |
| anti_6932_42 | Integrin alpha-5 | ITGA5 | Human |
| anti_6933_20 | 39S ribosomal protein L34, mitochondrial | MRPL34 | Human |
| anti_6935_123 | BMP and activin membrane-bound inhibitor homolog | BAMBI | Human |
| anti_6937_251 | Uncharacterized protein C1orf43 | C1orf43 | Human |
| anti_6938_21 | Protocadherin gamma-A12 | PCDHGA12 | Human |
| anti_6951_26 | Acid-sensing ion channel 4 | ASIC4 | Human |
| anti_6956_37 | FAS-associated factor 2 | FAF2 | Human |
| anti_6957_14 | Pancreatic lipase-related protein 2 | PNLIPRP2 | Human |
| anti_6965_19 | Contactin-associated protein-like 2 | CNTNAP2 | Human |
| anti_6966_144 | Syntaxin-1B | STX1B | Human |
| anti_6967_5 | Fc_MOUSE | Igh | Mouse |
| anti_6969_14 | WAP four-disulfide core domain protein 5 | WFDC5 | Human |
| anti_6973_2 | Insulin-like growth factor II | IGF2 | Human |
| anti_6974_6 | HLA class II histocompatibility antigen gamma chain | CD74 | Human |
| anti_6986_17 | Heparan sulfate glucosamine 3-O-sulfotransferase 3B1 | HS3ST3B1 | Human |
| anti_6990_44 | Stromal cell-derived factor 2-like protein 1 | SDF2L1 | Human |
| anti_6992_67 | Hemoglobin subunit delta | HBD | Human |
| anti_6993_8 | 5'-Nucleotidase | NT5E | Human |

| SOMAmer | Target Full Name | Entrez Gene Symbol | Organism |
|---|---|---|---|
| anti_6997_32 | Ras-related protein Rab-26 | RAB26 | Human |
| anti_7002_1 | Matrix-remodeling-associated protein 8 | MXRA8 | Human |
| anti_7003_4 | Polypeptide N-acetylgalactosaminyltransferase 10 | GALNT10 | Human |
| anti_7006_4 | Collagen alpha-1(XXV) chain | COL25A1 | Human |
| anti_7007_24 | Trafficking protein particle complex subunit 4 | TRAPPC4 | Human |
| anti_7009_6 | B-cell differentiation antigen CD72 | CD72 | Human |
| anti_7009_8 | B-cell differentiation antigen CD72 | CD72 | Human |
| anti_7011_8 | Kv channel-interacting protein 4 | KCNIP4 | Human |
| anti_7014_27 | Fc_MOUSE | Igh | Mouse |
| anti_7016_12 | Beta-1,3-galactosyl-O-glycosyl-glycoprotein beta-1,6-N-acetylglucosaminyltransferase | GCNT1 | Human |
| anti_7018_10 | Ribosome biogenesis protein TSR3 homolog | TSR3 | Human |
| anti_7019_13 | Semaphorin-7A | SEMA7A | Human |
| anti_7020_13 | Carbohydrate sulfotransferase 5 | CHST5 | Human |
| anti_7045_4 | BCL2/adenovirus E1B 19 kDa protein-interacting protein 3 | BNIP3 | Human |
| anti_7046_6 | Uncharacterized protein C20orf173 | C20orf173 | Human |
| anti_7048_4 | Carcinoembryonic antigen-related cell adhesion molecule 19 | CEACAM19 | Human |
| anti_7056_16 | 4F2 cell-surface antigen heavy chain | SLC3A2 | Human |
| anti_7059_14 | Leukocyte immunoglobulin-like receptor subfamily A member 6 | LILRA6 | Human |
| anti_7064_2 | Vesicle-associated membrane protein 8 | VAMP8 | Human |
| anti_7066_199 | Protein FAM209B | FAM209B | Human |
| anti_7069_9 | t-SNARE domain-containing protein 1 | TSNARE1 | Human |
| anti_7070_25 | Kell blood group glycoprotein | KEL | Human |
| anti_7073_69 | Uncharacterized protein C22orf15 | C22orf15 | Human |
| anti_7076_17 | Endothelin-converting enzyme-like 1 | ECEL1 | Human |
| anti_7077_9 | Fc_MOUSE | Igh | Mouse |
| anti_7081_2 | Butyrophilin subfamily 3 member A1 | BTN3A1 | Human |
| anti_7083_74 | Matrilin-4 | MATN4 | Human |
| anti_7084_1 | Draxin | DRAXIN | Human |
| anti_7089_42 | Synaptotagmin-11 | SYT11 | Human |
| anti_7092_7 | Protein disulfide-isomerase-like protein of the testis | PDILT | Human |
| anti_7093_20 | Neurexin-3-beta | NRXN3 | Human |
| anti_7096_30 | Regulator of microtubule dynamics protein 1 | RMDN1 | Human |
| anti_7097_8 | Testis-specific chromodomain protein Y 1 | CDY1 | Human |
| anti_7099_33 | Follistatin-related protein 5 | FSTL5 | Human |
| anti_7100_31 | T-cell surface antigen CD2 | CD2 | Human |
| anti_7104_71 | Calcium-binding protein 7 | CABP7 | Human |
| anti_7105_7 | CDK5 and ABL1 enzyme substrate 2 | CABLES2 | Human |
| anti_7108_7 | Small integral membrane protein 24 | SMIM24 | Human |
| anti_7113_1 | GrpE protein homolog 1, mitochondrial | GRPEL1 | Human |
| anti_7116_31 | Osteoclast-associated immunoglobulin-like receptor | OSCAR | Human |
| anti_7117_21 | Serpin I2 | SERPINI2 | Human |
| anti_7118_24 | Immunoglobulin superfamily DCC subclass member 3 | IGDCC3 | Human |
| anti_7121_2 | Synaptotagmin-7 | SYT7 | Human |
| anti_7122_31 | Leucine-rich repeat-containing G-protein coupled receptor 5 | LGR5 | Human |
| anti_7124_18 | Interleukin-21 | IL21 | Human |
| anti_7125_4 | Fc_MOUSE | Igh | Mouse |
| anti_7128_9 | von Willebrand factor A domain-containing protein 2 | VWA2 | Human |
| anti_7130_4 | Leukocyte-associated immunoglobulin-like receptor 2 | LAIR2 | Human |
| anti_7131_8 | Cholesteryl ester transfer protein | CETP | Human |
| anti_7132_55 | Complement C1q-like protein 4 | C1QL4 | Human |
| anti_7134_14 | Bcl-2-like protein 2 | BCL2L2 | Human |
| anti_7136_107 | Hemoglobin subunit epsilon | HBE1 | Human |
| anti_7137_8 | Mitochondrial peptide methionine sulfoxide reductase | MSRA | Human |

| SOMAmer | Target Full Name | Entrez Gene Symbol | Organism |
|---|---|---|---|
| anti_7141_21 | Alpha-1,3-mannosyl-glycoprotein 4-beta-N-acetylglucosaminyltransferase B | MGAT4B | Human |
| anti_7143_9 | N-acetyllactosaminide beta-1,6-N-acetylglucosaminyl-transferase, isoform C | GCNT2 GCNT5 II NACGT1 | Human |
| anti_7146_16 | Protein disulfide-isomerase A4 | PDIA4 | Human |
| anti_7146_5 | Protein disulfide-isomerase A4 | PDIA4 | Human |
| anti_7147_35 | WAP four-disulfide core domain protein 8 | WFDC8 | Human |
| anti_7153_66 | Neugrin | NGRN | Human |
| anti_7174_15 | ADP-ribosyl cyclase/cyclic ADP-ribose hydrolase 1 | CD38 | Human |
| anti_7175_4 | Protocadherin gamma-A2 | PCDHGA2 | Human |
| anti_7180_114 | Interferon alpha-14 | IFNA14 | Human |
| anti_7181_17 | Vesicle-associated membrane protein-associated protein B/C | VAPB | Human |
| anti_7183_102 | Angiopoietin-like protein 8 | ANGPTL8 | Human |
| anti_7186_111 | Syntaxin-3 | STX3 | Human |
| anti_7187_3 | Protein phosphatase 1L | PPM1L | Human |
| anti_7189_55 | Carbohydrate sulfotransferase 3 | CHST3 | Human |
| anti_7192_37 | Interferon lambda receptor 1 | IFNLR1 | Human |
| anti_7193_98 | Protocadherin alpha-C1 | PCDHAC1 | Human |
| anti_7196_21 | Interferon omega-1 | IFNW1 | Human |
| anti_7197_2 | DnaJ homolog subfamily C member 15 | DNAJC15 | Human |
| anti_7199_3 | Synapsin-3 | SYN3 | Human |
| anti_7200_4 | Leucine-rich repeat and fibronectin type-III domain-containing protein 2 | LRFN2 | Human |
| anti_7201_5 | Iron-sulfur cluster assembly enzyme ISCU, mitochondrial | ISCU | Human |
| anti_7202_107 | Semaphorin-6C | SEMA6C | Human |
| anti_7204_1 | Carcinoembryonic antigen-related cell adhesion molecule 21 | CEACAM21 | Human |
| anti_7207_4 | Protein Red | IK | Human |
| anti_7208_60 | Alpha-1,3-mannosyl-glycoprotein 4-beta-N-acetylglucosaminyltransferase C | MGAT4C | Human |
| anti_7211_2 | Ribonuclease pancreatic | RNASE1 | Human |
| anti_7219_152 | Heat shock 70 kDa protein 1A | HSPA1A | Human |
| anti_7220_20 | Interleukin-32 | IL32 | Human |
| anti_7221_56 | Cytoskeleton-associated protein 4 | CKAP4 | Human |
| anti_7224_11 | T-cell receptor-associated transmembrane adapter 1 | TRAT1 | Human |
| anti_7225_51 | Membrane magnesium transporter 1 | MMGT1 | Human |
| anti_7228_2 | Alpha-N-acetylgalactosaminide alpha-2,6-sialyltransferase 6 | ST6GALNAC6 | Human |
| anti_7231_37 | Transmembrane protease serine 11A | TMPRSS11A | Human |
| anti_7233_73 | P2X purinoceptor 6 | P2RX6 | Human |
| anti_7234_12 | Procollagen galactosyltransferase 2 | COLGALT2 | Human |
| anti_7239_9 | Glutathione S-transferase Mu 1 | GSTM1 | Human |
| anti_7240_2 | Membrane-bound transcription factor site-1 protease | MBTPS1 | Human |
| anti_7242_14 | V-set and transmembrane domain-containing protein 4 | VSTM4 | Human |
| anti_7243_8 | Interferon beta | IFNB1 | Human |
| anti_7244_16 | IGF-like family receptor 1 | IGFLR1 | Human |
| anti_7245_2 | CUGBP Elav-like family member 2 | CELF2 | Human |
| anti_7246_4 | FRAS1-related extracellular matrix protein 2 | FREM2 | Human |
| anti_7247_1 | RAB6-interacting golgin | GORAB | Human |
| anti_7249_37 | Bcl-2-like protein 10 | BCL2L10 | Human |
| anti_7253_6 | Protein quaking | QKI | Human |
| anti_7257_18 | Tuberoinfundibular peptide of 39 residues | PTH2 | Human |
| anti_7258_5 | Bone marrow proteoglycan | PRG2 | Human |
| anti_7262_191 | Carbohydrate sulfotransferase 14 | CHST14 | Human |
| anti_7265_32 | Osteocrin | OSTN | Human |
| anti_7267_2 | Serine palmitoyltransferase 2 | SPTLC2 | Human |
| anti_7268_12 | Interferon alpha-4 | IFNA4 | Human |
| anti_7551_33 | Leucine-rich repeat-containing protein 32 | LRRC32 | Human |

| SOMAmer | Target Full Name | Entrez Gene Symbol | Organism |
|---|---|---|---|
| anti_7587_49 | 50S ribosomal protein L7/L12_MYCTU | rplL | Mycobacterium tuberculosis |
| anti_7592_57 | 60 kDa chaperonin 2_MYCTU | groL2 | Mycobacterium tuberculosis |
| anti_7595_51 | 10 kDa chaperonin_MYCTU | groS | Mycobacterium tuberculosis |
| anti_7596_2 | 50S ribosomal protein L7/L12_MYCTU | rplL | Mycobacterium tuberculosis |
| anti_7598_15 | Adenylate kinase_MYCTU | adk | Mycobacterium tuberculosis |
| anti_7600_67 | 10 kDa chaperonin_MYCTU | groS | Mycobacterium tuberculosis |
| anti_7604_59 | Alpha-crystallin_MYCTU | hspX | Mycobacterium tuberculosis |
| anti_7605_11 | 60 kDa chaperonin 2_MYCTU | groL2 | Mycobacterium tuberculosis |
| anti_7606_49 | Chaperone protein DnaK_MYCTU | dnaK | Mycobacterium tuberculosis |
| anti_7608_61 | 10 kDa chaperonin_MYCTU | groS | Mycobacterium tuberculosis |
| anti_7609_13 | Probable thiol peroxidase_MYCTU | tpx | Mycobacterium tuberculosis |
| anti_7610_49 | Putative glyoxylase CFP32_MYCTU | cfp30B | Mycobacterium tuberculosis |
| anti_7612_56 | 6 kDa early secretory antigenic target_MYCTU | esxB cfp10 lhp mtsA10 Rv3874 MTV027.09 | Mycobacterium tuberculosis |
| anti_7615_18 | Immunogenic protein MPT64_MYCTU | mpt64 | Mycobacterium tuberculosis |
| anti_7616_43 | Alpha-crystallin_MYCTU | hspX | Mycobacterium tuberculosis |
| anti_7618_50 | Putative glyoxylase CFP32_MYCTU | cfp30B | Mycobacterium tuberculosis |
| anti_7619_25 | MPT51/MPB51 antigen_MYCTU | mpt51 | Mycobacterium tuberculosis |
| anti_7620_5 | 6 kDa early secretory antigenic target_MYCTU | esxB cfp10 lhp mtsA10 Rv3874 MTV027.09 | Mycobacterium tuberculosis |
| anti_7622_15 | Phosphate-binding protein pstS 1_MYCTU | pstS1 | Mycobacterium tuberculosis |
| anti_7624_19 | Ankyrin-2 | ANK2 | Human |
| anti_7655_11 | N-terminal pro-BNP | NPPB | Human |
| anti_7662_46 | C-C motif chemokine 21a_MOUSE | Ccl21a | Mouse |
| anti_7663_6 | C-C motif chemokine 2_MOUSE | Ccl2 | Mouse |
| anti_7664_173 | C-C motif chemokine 22_MOUSE | Ccl22 | Mouse |
| anti_7665_13 | C-C motif chemokine 5_MOUSE | Ccl5 | Mouse |
| anti_7667_15 | C-X-C motif chemokine 2_MOUSE | Cxcl2 | Mouse |
| anti_7668_20 | C-X-C motif chemokine 5_MOUSE | Cxcl5 | Mouse |
| anti_7670_68 | Insulin-like growth factor-binding protein 3_MOUSE | Igfbp3 | Mouse |
| anti_7671_58 | Interleukin-13_MOUSE | Il13 | Mouse |
| anti_7673_46 | Interleukin-19_MOUSE | Il19 | Mouse |
| anti_7674_25 | Interleukin-3_MOUSE | Il3 | Mouse |
| anti_7675_16 | Interleukin-4_MOUSE | Il4 | Mouse |
| anti_7676_90 | Galectin-4_MOUSE | Lgals4 | Mouse |
| anti_7678_84 | Mannose-binding protein C_MOUSE | Mbl2 | Mouse |
| anti_7679_4 | Midkine_MOUSE | Mdk | Mouse |
| anti_7680_205 | Ovostatin homolog_MOUSE | Ovos | Mouse |
| anti_7681_3 | Resistin_MOUSE | Retn | Mouse |
| anti_7684_24 | Fms-related tyrosine kinase 3 ligand_MOUSE | Flt3lg | Mouse |
| anti_7686_226 | Tissue-type plasminogen activator_MOUSE | Plat | Mouse |
| anti_7691_11 | Phosphoadenosine phosphosulfate reductase_ECOLI | cysH | strain K12 |
| anti_7693_13 | Tumor necrosis factor receptor superfamily member 10B | TNFRSF10B | Human |
| anti_7696_3 | Next to BRCA1 gene 1 protein | NBR1 | Human |
| anti_7713_102 | Fragile X mental retardation protein 1 | FMR1 | Human |

| SOMAmer | Target Full Name | Entrez Gene Symbol | Organism |
|---|---|---|---|
| anti_7713_50 | Fragile X mental retardation protein 1 | FMR1 | Human |
| anti_7732_45 | Vesicle-associated membrane protein 4 | VAMP4 | Human |
| anti_7736_28 | Opalin | OPALIN | Human |
| anti_7737_76 | Integrin beta-6 | ITGB6 | Human |
| anti_7738_299 | Syntaxin-2 | STX2 | Human |
| anti_7740_33 | Fetal and adult testis-expressed transcript protein | FATE1 | Human |
| anti_7741_111 | RNA polymerase-associated protein RTF1 homolog | RTF1 | Human |
| anti_7742_11 | RING finger protein 148 | RNF148 | Human |
| anti_7743_5 | Mitochondrial import inner membrane translocase subunit TIM50 | TIMM50 | Human |
| anti_7744_10 | Low-density lipoprotein receptor-related protein 12 | LRP12 | Human |
| anti_7745_3 | CDGSH iron-sulfur domain-containing protein 1 | CISD1 | Human |
| anti_7746_230 | Fc_MOUSE | Igh | Mouse |
| anti_7748_11 | NADH dehydrogenase [ubiquinone] flavoprotein 2, mitochondrial | NDUFV2 | Human |
| anti_7752_31 | C-type lectin domain family 4 member D | CLEC4D | Human |
| anti_7753_21 | Oxidoreductase HTATIP2 | HTATIP2 | Human |
| anti_7754_11 | Protein jagged-1 | JAG1 | Human |
| anti_7755_37 | Integrin beta-5 | ITGB5 | Human |
| anti_7756_37 | Killer cell lectin-like receptor subfamily F member 1 | KLRF1 | Human |
| anti_7761_125 | Choline/ethanolamine kinase | CHKB | Human |
| anti_7762_30 | Stannin | SNN | Human |
| anti_7763_25 | Cadherin-11 | CDH11 | Human |
| anti_7765_15 | Integral membrane protein 2A | ITM2A | Human |
| anti_7767_1 | Colipase-like protein 2 | CLPSL2 | Human |
| anti_7769_29 | SH3 domain-binding protein 2 | SH3BP2 | Human |
| anti_7770_25 | NFU1 iron-sulfur cluster scaffold homolog, mitochondrial | NFU1 | Human |
| anti_7775_15 | Kallikrein-11 | KLK11 | Human |
| anti_7778_104 | Caspase recruitment domain-containing protein 19 | CARD19 | Human |
| anti_7780_34 | Marginal zone B- and B1-cell-specific protein | MZB1 | Human |
| anti_7782_34 | Epididymal secretory protein E3-beta | EDDM3B | Human |
| anti_7786_83 | C-type lectin domain family 2 member B | CLEC2B | Human |
| anti_7788_1 | ATP synthase-coupling factor 6, mitochondrial | ATP5PF | Human |
| anti_7789_182 | Peroxiredoxin-4 | PRDX4 | Human |
| anti_7790_21 | Leukemia inhibitory factor | LIF | Human |
| anti_7792_58 | Coiled-coil domain-containing protein 90B, mitochondrial | CCDC90B | Human |
| anti_7795_14 | NKG2-E type II integral membrane protein | KLRC3 | Human |
| anti_7797_11 | Coiled-coil domain-containing protein 167 | CCDC167 | Human |
| anti_7799_3 | Killer cell immunoglobulin-like receptor 2DL5A | KIR2DL5A | Human |
| anti_7800_85 | Fc_MOUSE | Igh | Mouse |
| anti_7801_30 | Fc_MOUSE | Igh | Mouse |
| anti_7802_53 | Fc_MOUSE | Igh | Mouse |
| anti_7803_4 | Carbohydrate sulfotransferase 1 | CHST1 | Human |
| anti_7805_52 | Golgi SNAP receptor complex member 1 | GOSR1 | Human |
| anti_7812_11 | Protein S100-A9 | S100A9 | Human |
| anti_7813_6 | Alkaline phosphatase, placental type | ALPP | Human |
| anti_7815_49 | Insulin-like growth factor-binding protein-like 1 | IGFBPL1 | Human |
| anti_7816_23 | Fc_MOUSE | Igh | Mouse |
| anti_7817_36 | Fc_MOUSE | Igh | Mouse |
| anti_7821_6 | Nuclear nucleic acid-binding protein C1D | C1D | Human |
| anti_7822_11 | HRAS-like suppressor 2 | PLAAT2 | Human |
| anti_7824_88 | Methionine-R-sulfoxide reductase B3 | MSRB3 | Human |
| anti_7825_7 | Monoacylglycerol lipase ABHD12 | ABHD12 | Human |
| anti_7826_1 | Serine/threonine-protein kinase DCLK3 | DCLK3 | Human |
| anti_7827_20 | Disintegrin and metalloproteinase domain-containing protein 32 | ADAM32 | Human |
| anti_7835_2 | BCL2/adenovirus E1B 19 kDa protein-interacting protein 3-like | BNIP3L | Human |
| anti_7838_27 | Submaxillary gland androgen-regulated protein 3A | SMR3A | Human |
| anti_7840_64 | Legumain | LGMN | Human |

| SOMAmer | Target Full Name | Entrez Gene Symbol | Organism |
|---|---|---|---|
| anti_7841_84 | Endothelial cell-selective adhesion molecule | ESAM | Human |
| anti_7842_52 | Small EDRK-rich factor 1 | SERF1A | Human |
| anti_7843_152 | Fc_MOUSE | Igh | Mouse |
| anti_7847_66 | Fc_MOUSE | Igh | Mouse |
| anti_7849_3 | Glutaminyl-peptide cyclotransferase | QPCT | Human |
| anti_7851_30 | Ankyrin repeat domain-containing protein 46 | ANKRD46 | Human |
| anti_7852_9 | Translation initiation factor IF-3, mitochondrial | MTIF3 | Human |
| anti_7854_38 | Natural cytotoxicity triggering receptor 3 ligand 1 | NCR3LG1 | Human |
| anti_7864_3 | Sialic acid-binding Ig-like lectin 8 | SIGLEC8 | Human |
| anti_7865_126 | HRAS-like suppressor 3 | PLAAT3 | Human |
| anti_7867_154 | Alpha-N-acetylgalactosaminide alpha-2,6-sialyltransferase 1 | ST6GALNAC1 | Human |
| anti_7872_5 | Fc_MOUSE | Igh | Mouse |
| anti_7873_32 | Testis-specific serine/threonine-protein kinase 2 | TSSK2 | Human |
| anti_7878_2 | Protein transport protein Sec61 subunit beta | SEC61B | Human |
| anti_7879_12 | Cytochrome P450 3A4 | CYP3A4 | Human |
| anti_7887_57 | Cytochrome c oxidase subunit 5B, mitochondrial | COX5B | Human |
| anti_7888_58 | Cytochrome c oxidase assembly factor 3 homolog, mitochondrial | COA3 | Human |
| anti_7890_68 | Inactive dipeptidyl peptidase 10 | DPP10 | Human |
| anti_7891_45 | UDP-glucuronosyltransferase 1-6 | UGT1A6 | Human |
| anti_7892_132 | Protein FAM234B | FAM234B | Human |
| anti_7894_155 | Fibroblast growth factor 3 | FGF3 | Human |
| anti_7897_75 | Disks large homolog 3 | DLG3 | Human |
| anti_7898_29 | Nuclear protein MDM1 | MDM1 | Human |
| anti_7903_18 | Vesicle-associated membrane protein 3 | VAMP3 | Human |
| anti_7911_29 | Sodium channel subunit beta-4 | SCN4B | Human |
| anti_7915_31 | Amyloid-like protein 2 | APLP2 | Human |
| anti_7916_10 | Protein S100-A7 | S100A7 | Human |
| anti_7917_17 | Carboxypeptidase D | CPD | Human |
| anti_7919_278 | Protein FAM3D | FAM3D | Human |
| anti_7920_30 | Alpha-2,8-sialyltransferase 8B | ST8SIA2 | Human |
| anti_7921_65 | Four-jointed box protein 1 | FJX1 | Human |
| anti_7922_5 | Adrenomedullin | ADM | Human |
| anti_7923_41 | Semaphorin-4C | SEMA4C | Human |
| anti_7924_7 | T-cell surface glycoprotein CD3 gamma chain | CD3G | Human |
| anti_7925_18 | Single-pass membrane and coiled-coil domain-containing protein 2 | SMCO2 | Human |
| anti_7926_13 | Kunitz-type protease inhibitor 3 | SPINT3 | Human |
| anti_7927_16 | Alpha-N-acetylgalactosaminide alpha-2,6-sialyltransferase 5 | ST6GALNAC5 | Human |
| anti_7928_183 | Protein-tyrosine sulfotransferase 1 | TPST1 | Human |
| anti_7930_3 | Exosome complex component CSL4 | EXOSC1 | Human |
| anti_7932_23 | Synaptotagmin-8 | SYT8 | Human |
| anti_7935_26 | Glycosyltransferase-like protein LARGE1 | LARGE1 | Human |
| anti_7939_1 | Putative uncharacterized protein PQLC2L | SLC66A1L | Human |
| anti_7943_16 | CUB and zona pellucida-like domain-containing protein 1 | CUZD1 | Human |
| anti_7944_1 | Killer cell immunoglobulin-like receptor 3DL3 | KIR3DL3 | Human |
| anti_7947_19 | AP-4 complex accessory subunit tepsin | TEPSIN | Human |
| anti_7950_142 | Butyrophilin-like protein 9 | BTNL9 | Human |
| anti_7951_146 | Fc receptor-like protein 2 | FCRL2 | Human |
| anti_7952_2 | Vesicle transport through interaction with t-SNAREs homolog 1A | VTI1A | Human |
| anti_7953_20 | Signaling lymphocytic activation molecule | SLAMF1 | Human |
| anti_7958_15 | Kin of IRRE-like protein 2 | KIRREL2 | Human |
| anti_7960_53 | POTE ankyrin domain family member G | POTEG | Human |
| anti_7963_36 | Fc_MOUSE | Igh | Mouse |
| anti_7967_38 | Putative POTE ankyrin domain family member M | POTEM | Human |
| anti_7968_15 | Cytotoxic and regulatory T-cell molecule | CRTAM | Human |
| anti_7975_97 | Netrin receptor UNC5A | UNC5A | Human |
| anti_7976_19 | DOMON domain-containing protein FRRS1L | FRRS1L | Human |
| anti_7981_230 | Beta-1,3-galactosyltransferase 6 | B3GALT6 | Human |
| anti_7982_10 | 39S ribosomal protein L32, mitochondrial | MRPL32 | Human |
| anti_7983_1 | Protocadherin gamma-C5 | PCDHGC5 | Human |

| SOMAmer | Target Full Name | Entrez Gene Symbol | Organism |
|---|---|---|---|
| anti_7986_98 | N-acetylated-alpha-linked acidic dipeptidase 2 | NAALAD2 | Human |
| anti_7989_5 | Uncharacterized protein C1orf226 | C1orf226 | Human |
| anti_7991_54 | IQ domain-containing protein F1 | IQCF1 | Human |
| anti_7992_3 | Fc_MOUSE | Igh | Mouse |
| anti_7993_23 | Astacin-like metalloendopeptidase | ASTL | Human |
| anti_7994_41 | ERO1-like protein beta | ERO1B | Human |
| anti_7995_16 | von Willebrand factor C domain-containing protein 2-like | RIPK4 ANKRD3 DIK | Human |
| anti_7997_118 | Double C2-like domain-containing protein beta | DOC2B | Human |
| anti_7999_23 | Ectonucleoside triphosphate diphosphohydrolase 1 | ENTPD1 | Human |
| anti_8000_17 | Killer cell immunoglobulin-like receptor 2DL5A | KIR2DL5A | Human |
| anti_8002_27 | Transmembrane protease serine 5 | TMPRSS5 | Human |
| anti_8003_57 | Leucine-rich repeat LGI family member 3 | LGI3 | Human |
| anti_8004_15 | Glyceraldehyde-3-phosphate dehydrogenase, testis-specific | GAPDHS | Human |
| anti_8005_1 | Matrix-remodeling-associated protein 7 | MXRA7 | Human |
| anti_8006_12 | DnaJ homolog subfamily B member 12 | DNAJB12 | Human |
| anti_8008_28 | Activator of apoptosis harakiri | HRK | Human |
| anti_8011_96 | Iron/zinc purple acid phosphatase-like protein | ACP7 | Human |
| anti_8013_9 | VIP36-like protein | LMAN2L | Human |
| anti_8015_144 | Fc_MOUSE | Igh | Mouse |
| anti_8016_19 | DnaJ homolog subfamily C member 4 | DNAJC4 | Human |
| anti_8018_43 | V-set and immunoglobulin domain-containing protein 2 | VSIG2 | Human |
| anti_8019_73 | Stathmin-3 | STMN3 | Human |
| anti_8021_59 | 39S ribosomal protein L14, mitochondrial | MRPL14 | Human |
| anti_8023_23 | Fc_MOUSE | Igh | Mouse |
| anti_8029_35 | Fc_MOUSE | Igh | Mouse |
| anti_8031_11 | Fc_MOUSE | Igh | Mouse |
| anti_8032_23 | Mitochondrial coiled-coil domain protein 1 | MCCD1 | Human |
| anti_8033_1 | DnaJ homolog subfamily C member 18 | DNAJC18 | Human |
| anti_8034_6 | Dorsal root ganglia homeobox protein | DRGX | Human |
| anti_8035_6 | Uncharacterized protein C1orf198 | C1orf198 | Human |
| anti_8036_75 | Follitropin subunit beta | FSHB | Human |
| anti_8041_5 | Fc_MOUSE | Igh | Mouse |
| anti_8042_88 | Serine protease inhibitor Kazal-type 9 | SPINK9 | Human |
| anti_8045_3 | Killer cell immunoglobulin-like receptor 3DL3 | KIR3DL3 | Human |
| anti_8051_10 | Angiogenic factor with G patch and FHA domains 1 | AGGF1 | Human |
| anti_8052_115 | Neuroligin-1 | NLGN1 | Human |
| anti_8053_16 | DnaJ homolog subfamily B member 14 | DNAJB14 | Human |
| anti_8055_33 | Integral membrane protein DGCR2/IDD | DGCR2 | Human |
| anti_8057_78 | Fc_MOUSE | Igh | Mouse |
| anti_8059_1 | Thrombopoietin | THPO | Human |
| anti_8060_7 | Pro-neuregulin-2, membrane-bound isoform | NRG2 | Human |
| anti_8061_102 | Protein FAM171B | FAM171B | Human |
| anti_8062_15 | Protein eva-1 homolog B | EVA1B | Human |
| anti_8064_125 | Poliovirus receptor | PVR | Human |
| anti_8065_245 | Prostate and testis expressed protein 4 | PATE4 | Human |
| anti_8066_38 | Synaptotagmin-9 | SYT9 | Human |
| anti_8067_21 | Potassium voltage-gated channel subfamily E member 3 | KCNE3 | Human |
| anti_8068_43 | Uncharacterized family 31 glucosidase KIAA1161 | MYORG | Human |
| anti_8069_85 | T-cell surface glycoprotein CD3 epsilon chain | CD3E | Human |
| anti_8070_88 | Disks large homolog 4 | DLG4 | Human |
| anti_8071_114 | Protocadherin beta-7 | PCDHB7 | Human |
| anti_8072_19 | Fc_MOUSE | Igh | Mouse |
| anti_8073_3 | SLAM family member 5 | CD84 | Human |
| anti_8078_15 | Fc_MOUSE | Igh | Mouse |
| anti_8079_39 | EP300-interacting inhibitor of differentiation 3 | EID3 | Human |
| anti_8081_55 | Inactive phospholipase D5 | PLD5 | Human |
| anti_8085_10 | Fc_MOUSE | Igh | Mouse |
| anti_8087_250 | E3 ubiquitin-protein ligase RNF13 | RNF13 | Human |
| anti_8088_56 | Type III endosome membrane protein TEMP | C1orf210 | Human |
| anti_8089_173 | Nuclear receptor subfamily 4 group A member 1 | NR4A1 | Human |

| SOMAmer | Target Full Name | Entrez Gene Symbol | Organism |
|---|---|---|---|
| anti_8091_16 | Mannan-binding lectin serine protease 1 | MASP1 | Human |
| anti_8093_13 | Uncharacterized family 31 glucosidase KIAA1161 | MYORG | Human |
| anti_8094_20 | CDGSH iron-sulfur domain-containing protein 2 | CISD2 | Human |
| anti_8095_213 | Peptidyl-prolyl cis-trans isomerase F, mitochondrial | PPIF | Human |
| anti_8100_15 | ADM2 | ADM2 | Human |
| anti_8102_239 | Uncharacterized protein C1orf186 | RHEX | Human |
| anti_8104_21 | Interleukin-10 receptor subunit alpha | IL10RA | Human |
| anti_8106_15 | Translocon-associated protein subunit alpha | SSR1 | Human |
| anti_8107_12 | Transmembrane protein 234 | TMEM234 | Human |
| anti_8221_19 | Macrophage migration inhibitory factor | MIF | Human |
| anti_8222_49 | Cadherin-related family member 3 | CDHR3 | Human |
| anti_8228_10 | Fc_MOUSE | Igh | Mouse |
| anti_8231_122 | Vascular endothelial growth factor receptor 1 | FLT1 | Human |
| anti_8236_8 | UPF0729 protein C18orf32 | C18orf32 | Human |
| anti_8237_56 | Sclerostin domain-containing protein 1 | SOSTDC1 | Human |
| anti_8242_9 | C-type lectin domain family 2 member L | CLEC2L | Human |
| anti_8243_55 | Serine protease inhibitor Kazal-type 1 | SPINK1 | Human |
| anti_8249_124 | ALK tyrosine kinase receptor | ALK | Human |
| anti_8253_2 | Protein O-linked-mannose beta-1,2-N-acetylglucosaminyltransferase 1 | POMGNT1 | Human |
| anti_8255_34 | Protein MRVI1 | MRVI1 | Human |
| anti_8256_57 | Tumor necrosis factor receptor superfamily member 11A | TNFRSF11A | Human |
| anti_8257_71 | Oxytocin-neurophysin 1 | OXT | Human |
| anti_8259_25 | Fc_MOUSE | Igh | Mouse |
| anti_8260_13 | Fc_MOUSE | Igh | Mouse |
| anti_8261_51 | Erythrocyte band 7 integral membrane protein | STOM | Human |
| anti_8263_64 | TYMS opposite strand protein | TYMSOS | Human |
| anti_8265_225 | Lamina-associated polypeptide 2, isoforms beta/gamma | TMPO | Human |
| anti_8268_98 | Heparan sulfate glucosamine 3-O-sulfotransferase 3A1 | HS3ST3A1 | Human |
| anti_8271_24 | Fc_MOUSE | Igh | Mouse |
| anti_8272_22 | 3-keto-steroid reductase | HSD17B7 | Human |
| anti_8282_15 | Fc_MOUSE | Igh | Mouse |
| anti_8285_64 | Pituitary adenylate cyclase-activating polypeptide | ADCYAP1 | Human |
| anti_8286_44 | Fc_MOUSE | Igh | Mouse |
| anti_8287_17 | CMRF35-like molecule 2 | CD300E | Human |
| anti_8290_1 | Fc_MOUSE | Igh | Mouse |
| anti_8296_117 | KDEL motif-containing protein 2 | POGLUT3 | Human |
| anti_8297_8 | DnaJ homolog subfamily C member 10 | DNAJC10 | Human |
| anti_8298_8 | Serine-rich single-pass membrane protein 1 | SSMEM1 | Human |
| anti_8300_82 | Peroxisomal membrane protein PEX14 | PEX14 | Human |
| anti_8303_102 | UPF0160 protein MYG1, mitochondrial | C12orf10 | Human |
| anti_8306_54 | Transmembrane gamma-carboxyglutamic acid protein 1 | PRRG1 | Human |
| anti_8307_47 | UPF0696 protein C11orf68 | C11orf68 | Human |
| anti_8314_71 | N-acetylglucosamine-6-sulfatase | GNS | Human |
| anti_8315_5 | Beta-defensin 119 | DEFB119 | Human |
| anti_8316_36 | Fc_MOUSE | Igh | Mouse |
| anti_8318_13 | Sprouty-related, EVH1 domain-containing protein 1 | SPRED1 | Human |
| anti_8320_5 | SAFB-like transcription modulator | SLTM | Human |
| anti_8321_27 | Zinc finger protein 843 | ZNF843 | Human |
| anti_8326_63 | Single Ig IL-1-related receptor | SIGIRR | Human |
| anti_8328_9 | Ethanolamine kinase 1 | ETNK1 | Human |
| anti_8329_166 | Cadherin-12 | CDH12 | Human |
| anti_8336_267 | GRAM domain-containing protein 1C | GRAMD1C | Human |
| anti_8337_65 | Receptor-type tyrosine-protein phosphatase U | PTPRU | Human |
| anti_8339_72 | Gastrin-releasing peptide | GRP | Human |
| anti_8340_9 | Beta-defensin 110 | DEFB110 | Human |
| anti_8343_224 | Poly(A) polymerase gamma | PAPOLG | Human |
| anti_8345_27 | Glutathione peroxidase 7 | GPX7 | Human |
| anti_8346_9 | Dipeptidyl peptidase 2 | DPP7 | Human |

| SOMAmer | Target Full Name | Entrez Gene Symbol | Organism |
|---|---|---|---|
| anti_8347_222 | Beta-defensin 129 | DEFB129 | Human |
| anti_8348_4 | Ephrin type-B receptor 2 | EPHB2 | Human |
| anti_8351_17 | Serine protease 57 | PRSS57 | Human |
| anti_8353_15 | Sodium channel subunit beta-2 | SCN2B | Human |
| anti_8355_80 | Isthmin-1 | ISM1 C20orf82 ISM | Human |
| anti_8356_88 | Oxytocin-neurophysin 1 | OXT | Human |
| anti_8357_43 | Sulfotransferase 4A1 | SULT4A1 | Human |
| anti_8358_30 | Thioredoxin-dependent peroxide reductase, mitochondrial | PRDX3 | Human |
| anti_8359_149 | Serine/threonine-protein kinase 17B | STK17B | Human |
| anti_8360_169 | Natural cytotoxicity triggering receptor 1 | NCR1 | Human |
| anti_8362_102 | Immunoglobulin J chain | JCHAIN | Human |
| anti_8363_18 | UPF0577 protein KIAA1324-like | KIAA1324L | Human |
| anti_8364_74 | Uronyl 2-sulfotransferase | UST | Human |
| anti_8366_19 | Uncharacterized protein C1orf115 | C1orf115 | Human |
| anti_8367_142 | V-set and immunoglobulin domain-containing protein 1 | VSIG1 | Human |
| anti_8369_102 | Dystroglycan | DAG1 | Human |
| anti_8370_102 | Beta-defensin 119 | DEFB119 | Human |
| anti_8372_29 | Cadherin-related family member 1 | CDHR1 | Human |
| anti_8376_25 | Lutropin subunit beta | LHB | Human |
| anti_8377_87 | Fc_MOUSE | Igh | Mouse |
| anti_8379_35 | Interleukin-9 | IL9 | Human |
| anti_8380_244 | Protein delta homolog 1 | DLK1 | Human |
| anti_8381_18 | Fc_MOUSE | Igh | Mouse |
| anti_8383_20 | Uncharacterized protein C11orf94 | C11orf94 | Human |
| anti_8386_11 | Prostate and testis expressed protein 1 | PATE1 | Human |
| anti_8388_24 | Spastin | SPAST | Human |
| anti_8390_25 | Cytochrome c oxidase subunit 7A1, mitochondrial | COX7A1 | Human |
| anti_8391_12 | Beta-defensin 115 | DEFB115 | Human |
| anti_8393_121 | Cathepsin F | CTSF | Human |
| anti_8394_56 | Non-secretory ribonuclease | RNASE2 | Human |
| anti_8396_42 | Fatty-acid amide hydrolase 2 | FAAH2 | Human |
| anti_8399_6 | Serine/threonine-protein kinase 17B | STK17B | Human |
| anti_8400_74 | Gastrin-releasing peptide | GRP | Human |
| anti_8403_18 | Fatty acid synthase | FASN | Human |
| anti_8404_102 | BolA-like protein 2 | BOLA2 | Human |
| anti_8406_17 | Insulin-like growth factor I | IGF1 | Human |
| anti_8409_3 | R-spondin-2 | RSPO2 | Human |
| anti_8418_30 | Catenin beta-1 | CTNNB1 | Human |
| anti_8424_269 | Catenin beta-1 | CTNNB1 | Human |
| anti_8427_118 | R-spondin-3 | RSPO3 | Human |
| anti_8443_9 | Magainin-1, Xenopus laevis | magainins | African clawed frog |
| anti_8444_3 | Magainin-2_XENLA | magainins | African clawed frog |
| anti_8444_46 | Magainin-2_XENLA | magainins | African clawed frog |
| anti_8445_184 | Melittin_VESMG | MELT | Hornet |
| anti_8445_54 | Melittin_VESMG | MELT | Hornet |
| anti_8446_4 | Pituitary adenylate cyclase-activating polypeptide 27 | ADCYAP1 | Human |
| anti_8447_11 | Appetite-regulating hormone | GHRL | Human |
| anti_8449_103 | Exendin-4_HELSU | | Gila monster |
| anti_8449_124 | Exendin-4_HELSU | | Gila monster |
| anti_8450_36 | Pituitary adenylate cyclase-activating polypeptide 38 | ADCYAP1 | Human |
| anti_8457_4 | Lysozyme C_CHICK | LYZ_CHICK | Chicken |
| anti_8458_16 | Alpha-synuclein | SNCA | Human |
| anti_8462_18 | Somatotropin | GH1 | Human |
| anti_8464_31 | R-spondin-4 | RSPO4 | Human |
| anti_8465_52 | Cathepsin H | CTSH | Human |
| anti_8467_9 | Inhibin beta A chain:Inhibin beta B chain heterodimer | INHBA\|INHBB | Human |
| anti_8468_19 | Prostate-specific antigen | KLK3 | Human |

| SOMAmer | Target Full Name | Entrez Gene Symbol | Organism |
|---|---|---|---|
| anti_8470_213 | Ribonuclease H1 | RNASEH1 | Human |
| anti_8475_15 | Matrilysin | MMP7 | Human |
| anti_8478_2 | Tyrosine-protein phosphatase YopH_YEREN | yopH | Yersinia enterocolitica |
| anti_8478_22 | Tyrosine-protein phosphatase YopH_YEREN | yopH | Yersinia enterocolitica |
| anti_8479_4 | Stromelysin-2 | MMP10 | Human |
| anti_8481_26 | Malic dehydrogenase_THETH | mdh | Thermus thermophilus |
| anti_8481_44 | Malic dehydrogenase_THETH | mdh | Thermus thermophilus |
| anti_8482_39 | Nigrin b_SAMNI | | European elder |
| anti_8483_5 | Luciferin 4-monooxygenase_PHOPY | | Common eastern firefly |
| anti_8488_33 | Integrin alpha-IIb: beta-3 complex | ITGA2B\|ITGB3 | Human |
| anti_8520_8 | Disintegrin and metalloproteinase domain-containing protein 30 | ADAM30 | Human |
| anti_8529_1 | Tumor necrosis factor receptor superfamily member 10B | TNFRSF10B | Human |
| anti_8545_14 | Uncharacterized protein C17orf78 | C17orf78 | Human |
| anti_8556_5 | Protein FAM134B | RETREG1 | Human |
| anti_8569_147 | ADP-ribosylation factor-like protein 8B | ARL8B | Human |
| anti_8587_21 | Serine protease inhibitor Kazal-type 14 | SPINK14 | Human |
| anti_8595_75 | Fc_MOUSE | Igh | Mouse |
| anti_8601_167 | Low-density lipoprotein receptor-related protein 1, soluble | LRP1 | Human |
| anti_8606_39 | Transmembrane glycoprotein NMB | GPNMB | Human |
| anti_8613_97 | Fc_MOUSE | Igh | Mouse |
| anti_8619_12 | Fc_MOUSE | Igh | Mouse |
| anti_8624_16 | Netrin receptor UNC5D | UNC5D | Human |
| anti_8631_13 | Erythroid membrane-associated protein | ERMAP | Human |
| anti_8633_18 | E3 ubiquitin-protein ligase RNF128 | RNF128 | Human |
| anti_8635_283 | Zona pellucida-binding protein 1 | ZPBP | Human |
| anti_8644_101 | Cathepsin H | CTSH | Human |
| anti_8645_257 | Peroxisomal membrane protein PEX14 | PEX14 | Human |
| anti_8646_61 | Leucine-rich repeat transmembrane neuronal protein 4 | LRRTM4 | Human |
| anti_8653_132 | DnaJ homolog subfamily C member 4 | DNAJC4 | Human |
| anti_8654_13 | Disintegrin and metalloproteinase domain-containing protein 10 | ADAM10 | Human |
| anti_8659_68 | Protein G6b | MPIG6B | Human |
| anti_8664_36 | Extracellular tyrosine-protein kinase PKDCC | PKDCC | Human |
| anti_8671_378 | Trafficking protein particle complex subunit 5 | TRAPPC5 | Human |
| anti_8675_79 | Fc_MOUSE | Igh | Mouse |
| anti_8681_93 | Membrane-associated progesterone receptor component 2 | PGRMC2 | Human |
| anti_8686_342 | Protein Hikeshi | HIKESHI | Human |
| anti_8687_26 | Transmembrane protein 106B | TMEM106B | Human |
| anti_8690_25 | Caveolin-3 | CAV3 | Human |
| anti_8699_43 | Cysteine-rich motor neuron 1 protein | CRIM1 | Human |
| anti_8700_325 | Polypeptide N-acetylgalactosaminyltransferase 11 | GALNT11 | Human |
| anti_8702_42 | Osteoclast-associated immunoglobulin-like receptor | OSCAR | Human |
| anti_8748_45 | HLA class II histocompatibility antigen gamma chain | CD74 | Human |
| anti_8749_194 | Antigen-presenting glycoprotein CD1d | CD1D | Human |
| anti_8750_46 | Vinculin | VCL | Human |
| anti_8752_6 | Fc_MOUSE | Igh | Mouse |
| anti_8754_5 | Neuromedin-U | NMU | Human |
| anti_8756_41 | Potassium voltage-gated channel subfamily E regulatory beta subunit 5 | KCNE5 | Human |
| anti_8759_29 | Lactosylceramide 4-alpha-galactosyltransferase | A4GALT | Human |
| anti_8761_7 | Fc_MOUSE | Igh | Mouse |
| anti_8762_38 | CD70 antigen | CD70 | Human |
| anti_8765_23 | Ephrin-B2 | EFNB2 | Human |

| SOMAmer | Target Full Name | Entrez Gene Symbol | Organism |
|---|---|---|---|
| anti_8766_29 | Leukocyte immunoglobulin-like receptor subfamily A member 5 | LILRA5 | Human |
| anti_8767_44 | Endothelin-converting enzyme 1 | ECE1 | Human |
| anti_8769_30 | Fc_MOUSE | Igh | Mouse |
| anti_8770_136 | Fc_MOUSE | Igh | Mouse |
| anti_8772_5 | Ephrin-B2 | EFNB2 | Human |
| anti_8773_172 | EMILIN-3 | EMILIN3 | Human |
| anti_8775_61 | Protein FAM24B | FAM24B | Human |
| anti_8776_10 | Erlin-1 | ERLIN1 | Human |
| anti_8777_5 | Fc_MOUSE | Igh | Mouse |
| anti_8778_3 | Noggin | NOG | Human |
| anti_8783_216 | Fc_MOUSE | Igh | Mouse |
| anti_8784_7 | BRCA1-A complex subunit Abraxas | ABRAXAS1 | Human |
| anti_8785_1 | Armadillo repeat-containing protein 5 | ARMC5 | Human |
| anti_8786_6 | Protein FAM171B | FAM171B | Human |
| anti_8787_21 | Fc_MOUSE | Igh | Mouse |
| anti_8790_6 | Arylsulfatase A | ARSA | Human |
| anti_8792_17 | Fc_MOUSE | Igh | Mouse |
| anti_8793_13 | Fc_MOUSE | Igh | Mouse |
| anti_8794_13 | Dipeptidase 1 | DPEP1 | Human |
| anti_8798_29 | Golgi membrane protein 1 | GOLM1 | Human |
| anti_8800_14 | Tetratricopeptide repeat protein 17 | TTC17 | Human |
| anti_8802_24 | Fc_MOUSE | Igh | Mouse |
| anti_8803_61 | Nectin-1, isoform gamma | NECTIN1 | Human |
| anti_8804_39 | Collagen alpha-1(XX) chain | COL20A1 | Human |
| anti_8806_18 | Fc_MOUSE | Igh | Mouse |
| anti_8807_13 | Sorting nexin-1 | SNX1 | Human |
| anti_8808_90 | Fc_MOUSE | Igh | Mouse |
| anti_8810_26 | Hepatocyte cell adhesion molecule | HEPACAM | Human |
| anti_8811_24 | BMP and activin membrane-bound inhibitor homolog | BAMBI | Human |
| anti_8813_160 | Thioredoxin | TXN | Human |
| anti_8815_1 | Olfactomedin-like protein 1 | OLFML1 | Human |
| anti_8816_44 | Armadillo repeat-containing protein 5 | ARMC5 | Human |
| anti_8817_29 | Centromere protein V | CENPV | Human |
| anti_8818_13 | Interferon gamma receptor 2 | IFNGR2 | Human |
| anti_8820_2 | Fc_MOUSE | Igh | Mouse |
| anti_8822_163 | Fc_MOUSE | Igh | Mouse |
| anti_8824_2 | Fc_MOUSE | Igh | Mouse |
| anti_8825_4 | Paired immunoglobulin-like type 2 receptor alpha | PILRA | Human |
| anti_8827_1 | Integral membrane protein 2A | ITM2A | Human |
| anti_8828_21 | Epithelial splicing regulatory protein 1 | ESRP1 | Human |
| anti_8829_4 | alpha-2-macroglobulin receptor-associated protein | LRPAP1 | Human |
| anti_8830_29 | BRISC complex subunit Abro1 | ABRAXAS2 | Human |
| anti_8832_55 | Bone marrow stromal antigen 2 | BST2 | Human |
| anti_8833_20 | Tumor necrosis factor ligand superfamily member 10 | TNFSF10 | Human |
| anti_8834_58 | Calnexin | CANX | Human |
| anti_8838_10 | Protein CASC4 | CASC4 | Human |
| anti_8839_4 | Metaxin-2 | MTX2 | Human |
| anti_8841_65 | Cartilage intermediate layer protein 2 | CILP2 | Human |
| anti_8842_16 | GRAM domain-containing protein 1C | GRAMD1C | Human |
| anti_8843_34 | Myelin regulatory factor | MYRF | Human |
| anti_8845_2 | A disintegrin and metalloproteinase with thrombospondin motifs 3 | ADAMTS3 | Human |
| anti_8850_5 | Putative inactive group IIC secretory phospholipase A2 | PLA2G2C | Human |
| anti_8851_42 | Protein FAM171B | FAM171B | Human |
| anti_8852_10 | SUN domain-containing protein 3 | SUN3 | Human |
| anti_8853_2 | C-type lectin domain family 4 member A | CLEC4A | Human |
| anti_8854_59 | Ribonucleoside-diphosphate reductase subunit M2 B | RRM2B | Human |
| anti_8858_21 | Basic leucine zipper transcriptional factor ATF-like 3 | BATF3 | Human |
| anti_8859_51 | Carbonic anhydrase-related protein 11 | CA11 | Human |

| SOMAmer | Target Full Name | Entrez Gene Symbol | Organism |
|---|---|---|---|
| anti_8863_3 | Phosphatidylinositol 3-kinase regulatory subunit alpha | PIK3R1 | Human |
| anti_8864_59 | Centromere protein W | CENPW | Human |
| anti_8867_18 | Glycodelin | PAEP | Human |
| anti_8869_5 | Butyrophilin subfamily 2 member A1 | BTN2A1 | Human |
| anti_8870_38 | Nucleophosmin | NPM1 | Human |
| anti_8871_14 | Nuclear migration protein nudC | NUDC | Human |
| anti_8872_1 | Protein transport protein Sec61 subunit gamma | SEC61G | Human |
| anti_8874_53 | Ceroid-lipofuscinosis neuronal protein 5 | CLN5 | Human |
| anti_8876_51 | Neurexin-2-beta | NRXN2 | Human |
| anti_8877_22 | Protein eva-1 homolog C | EVA1C | Human |
| anti_8878_48 | YTH domain-containing protein 1 | YTHDC1 | Human |
| anti_8885_6 | Voltage-dependent calcium channel subunit alpha-2/delta-3 | CACNA2D3 | Human |
| anti_8889_5 | Cell cycle progression protein 1 | CCPG1 | Human |
| anti_8891_7 | Probable serine carboxypeptidase CPVL | CPVL | Human |
| anti_8893_29 | Poly [ADP-ribose] polymerase 1 | PARP1 | Human |
| anti_8894_80 | Heterogeneous nuclear ribonucleoprotein A/B | HNRNPAB | Human |
| anti_8897_3 | Leucine-rich repeat-containing protein 37A2 | LRRC37A2 | Human |
| anti_8898_19 | Fc_MOUSE | Igh | Mouse |
| anti_8899_75 | UDP-glucuronosyltransferase 1-8 | UGT1A8 | Human |
| anti_8909_77 | Glucosamine-6-phosphate isomerase 1 | GNPDA1 | Human |
| anti_8913_22 | Sialic acid-binding Ig-like lectin 11 | SIGLEC11 | Human |
| anti_8921_139 | Polyadenylate-binding protein 2 | PABPN1 | Human |
| anti_8923_94 | Polypeptide N-acetylgalactosaminyltransferase 16 | GALNT16 | Human |
| anti_8924_55 | Neuron-specific vesicular protein calcyon | CALY | Human |
| anti_8925_25 | Ribonucleoside-diphosphate reductase subunit M2 B | RRM2B | Human |
| anti_8927_6 | Uncharacterized protein C7orf73 | SGTA SGT SGT1 | Human |
| anti_8929_7 | Transcription factor TFIIIB component B" homolog | BDP1 | Human |
| anti_8931_124 | G0/G1 switch protein 2 | G0S2 | Human |
| anti_8933_84 | Myocyte-specific enhancer factor 2C | MEF2C | Human |
| anti_8935_22 | Multidrug resistance-associated protein 6 | ABCC6 | Human |
| anti_8941_4 | Neuroligin-3 | NLGN3 | Human |
| anti_8944_42 | Fc_MOUSE | Igh | Mouse |
| anti_8945_7 | BRISC complex subunit Abro1 | ABRAXAS2 | Human |
| anti_8946_38 | Bile acid receptor | NR1H4 | Human |
| anti_8947_268 | Fc_MOUSE | Igh | Mouse |
| anti_8948_13 | Disintegrin and metalloproteinase domain-containing protein 19 | ADAM19 | Human |
| anti_8949_3 | Fc_MOUSE | Igh | Mouse |
| anti_8951_162 | Chondroitin sulfate proteoglycan 4 | CSPG4 | Human |
| anti_8954_30 | Mitogen-activated protein kinase kinase kinase kinase 1 | MAP4K1 | Human |
| anti_8955_60 | Glycosyltransferase 8 domain-containing protein 1 | GLT8D1 | Human |
| anti_8957_72 | Endoplasmic reticulum lectin 1 | ERLEC1 | Human |
| anti_8959_61 | Disintegrin and metalloproteinase domain-containing protein 17 | ADAM17 | Human |
| anti_8962_48 | Ribosomal protein S6 kinase beta-1 | RPS6KB1 | Human |
| anti_8963_8 | Vesicle transport through interaction with t-SNAREs homolog 1B | VTI1B | Human |
| anti_8964_14 | Fc_MOUSE | Igh | Mouse |
| anti_8970_9 | Receptor-interacting serine/threonine-protein kinase 2 | RIPK2 | Human |
| anti_8973_23 | Fc receptor-like protein 4 | FCRL4 | Human |
| anti_8975_26 | Cell differentiation protein RCD1 homolog | CNOT9 | Human |
| anti_8976_13 | Beta-1,4 N-acetylgalactosaminyltransferase 1 | B4GALNT1 | Human |
| anti_8978_30 | Mitogen-activated protein kinase kinase kinase kinase 3 | MAP4K3 | Human |
| anti_8979_1 | Fc_MOUSE | Igh | Mouse |
| anti_8980_19 | Cell surface glycoprotein CD200 receptor 2 | CD200R1L | Human |
| anti_8984_28 | Leucine-rich repeat and calponin homology domain-containing protein 4 | LRCH4 | Human |
| anti_8985_13 | Protein SERAC1 | SERAC1 | Human |

| SOMAmer | Target Full Name | Entrez Gene Symbol | Organism |
|---|---|---|---|
| anti_8986_2 | D-3-phosphoglycerate dehydrogenase | PHGDH | Human |
| anti_8989_40 | Signal peptide, CUB and EGF-like domain-containing protein 1 | SCUBE1 | Human |
| anti_8992_1 | Transmembrane protein 2 | CEMIP2 | Human |
| anti_8993_151 | Receptor-interacting serine/threonine-protein kinase 2 | RIPK2 | Human |
| anti_8994_65 | SLAM family member 8 | SLAMF8 | Human |
| anti_8997_4 | Neuronal pentraxin receptor | NPTXR | Human |
| anti_8998_15 | Heparan sulfate glucosamine 3-O-sulfotransferase 4 | HS3ST4 | Human |
| anti_8999_19 | Carcinoembryonic antigen-related cell adhesion molecule 3 | CEACAM3 | Human |
| anti_9004_24 | Low-density lipoprotein receptor class A domain-containing protein 4 | LDLRAD4 | Human |
| anti_9008_6 | Transmembrane gamma-carboxyglutamic acid protein 1 | PRRG1 | Human |
| anti_9010_3 | Fc_MOUSE | Igh | Mouse |
| anti_9012_1 | Pre-mRNA-processing factor 6 | PRPF6 | Human |
| anti_9013_60 | Netrin-1 | NTN1 | Human |
| anti_9014_18 | Transmembrane protein 25 | TMEM25 | Human |
| anti_9015_1 | Proteoglycan 3 | PRG3 | Human |
| anti_9017_58 | Lactase-phlorizin hydrolase | LCT | Human |
| anti_9018_38 | Protocadherin-10 | PCDH10 | Human |
| anti_9021_1 | Hepatitis A virus cellular receptor 1 | HAVCR1 | Human |
| anti_9022_49 | Beta-defensin 132 | DEFB132 | Human |
| anti_9026_40 | Butyrophilin-like protein 8 | BTNL8 | Human |
| anti_9027_10 | Neuroligin-2 | NLGN2 | Human |
| anti_9030_56 | Uncharacterized protein C17orf89 | NDUFAF8 | Human |
| anti_9032_30 | Proto-oncogene tyrosine-protein kinase ROS | ROS1 | Human |
| anti_9035_2 | MAX-interacting protein 1 | MXI1 | Human |
| anti_9038_12 | Protein JTB | JTB | Human |
| anti_9040_144 | Mammalian ependymin-related protein 1 | EPDR1 | Human |
| anti_9043_7 | Tuftelin-interacting protein 11 | TFIP11 | Human |
| anti_9046_46 | Fc_MOUSE | Igh | Mouse |
| anti_9049_2 | Monocarboxylate transporter 4 | SLC16A3 | Human |
| anti_9050_170 | Arginine/serine-rich protein 1 | RSRP1 | Human |
| anti_9051_13 | Interleukin-32 | IL32 | Human |
| anti_9054_8 | Fc_MOUSE | Igh | Mouse |
| anti_9055_81 | Myocardial zonula adherens protein | MYZAP | Human |
| anti_9057_19 | Proteasomal ubiquitin receptor ADRM1 | ADRM1 | Human |
| anti_9058_1 | Mitochondrial import inner membrane translocase subunit Tim21 | TIMM21 | Human |
| anti_9059_14 | Rap guanine nucleotide exchange factor 5 | RAPGEF5 | Human |
| anti_9060_27 | Protein turtle homolog B | IGSF9B | Human |
| anti_9064_12 | RING finger protein 215 | RNF215 | Human |
| anti_9065_28 | Alpha-taxilin | TXLNA | Human |
| anti_9067_152 | Fc_MOUSE | Igh | Mouse |
| anti_9070_1 | Uncharacterized protein C17orf67 | C17orf67 | Human |
| anti_9073_35 | Fc_MOUSE | Igh | Mouse |
| anti_9074_6 | Fatty acid hydroxylase domain-containing protein 2 | FAXDC2 | Human |
| anti_9075_121 | UPF0258 protein KIAA1024 | MINAR1 | Human |
| anti_9078_207 | Beta-1-syntrophin | SNTB1 | Human |
| anti_9080_1 | Fc_MOUSE | Igh | Mouse |
| anti_9081_39 | Zinc transporter 3 | SLC30A3 | Human |
| anti_9083_35 | Tyrosine-protein phosphatase non-receptor type substrate 1 | SIRPA | Human |
| anti_9086_95 | Mammaglobin-B | SCGB2A1 | Human |
| anti_9089_77 | Junctophilin-3 | JPH3 | Human |
| anti_9094_5 | C-type lectin domain family 4 member C | CLEC4C | Human |
| anti_9095_5 | Mitochondrial fission regulator 1 | MTFR1 | Human |
| anti_9096_80 | Lymphoid-restricted membrane protein | LRMP | Human |
| anti_9097_5 | UPF0577 protein KIAA1324 | KIAA1324 | Human |
| anti_9106_87 | LysM and putative peptidoglycan-binding domain-containing protein 4 | LYSMD4 | Human |

| SOMAmer | Target Full Name | Entrez Gene Symbol | Organism |
|---|---|---|---|
| anti_9107_59 | Armadillo repeat-containing protein 10 | ARMC10 | Human |
| anti_9108_48 | Fc_MOUSE | Igh | Mouse |
| anti_9110_2 | Synaptotagmin-17 | SYT17 | Human |
| anti_9111_40 | Junctophilin-4 | JPH4 | Human |
| anti_9114_84 | Uromodulin-like 1 | UMODL1 | Human |
| anti_9115_78 | ADAM DEC1 | ADAMDEC1 | Human |
| anti_9116_28 | HEPACAM family member 2 | HEPACAM2 | Human |
| anti_9117_4 | Interleukin-36 gamma | IL36G | Human |
| anti_9121_28 | Galactoside 2-alpha-L-fucosyltransferase 2 | FUT2 | Human |
| anti_9122_8 | Fc_MOUSE | Igh | Mouse |
| anti_9123_18 | Protein Dos | CBARP | Human |
| anti_9125_23 | Mannan-binding lectin serine protease 1 | MASP1 | Human |
| anti_9126_171 | 5'-nucleotidase domain-containing protein 3 | NT5DC3 | Human |
| anti_9127_4 | Prestin | SLC26A5 | Human |
| anti_9128_34 | Leucine-rich repeat transmembrane protein FLRT3 | FLRT3 | Human |
| anti_9168_31 | C-C motif chemokine 26 | CCL26 | Human |
| anti_9170_24 | Interleukin-17A | IL17A | Human |
| anti_9171_11 | Cysteine and glycine-rich protein 3 | CSRP3 | Human |
| anti_9173_21 | Phosphoglucomutase-1 | PGM1 | Human |
| anti_9175_48 | Down syndrome cell adhesion molecule | DSCAM | Human |
| anti_9176_3 | Mucin-1 | MUC1 | Human |
| anti_9178_30 | Neuregulin-1 | NRG1 | Human |
| anti_9180_6 | Interferon gamma receptor 2 | IFNGR2 | Human |
| anti_9182_3 | Low-density lipoprotein receptor-related protein 1, soluble | LRP1 | Human |
| anti_9185_15 | Trefoil factor 1 | TFF1 | Human |
| anti_9188_119 | C-X-C motif chemokine 9 | CXCL9 | Human |
| anti_9197_4 | Galectin-9 | LGALS9 | Human |
| anti_9202_309 | ATP synthase subunit O, mitochondrial | ATP5PO | Human |
| anti_9204_33 | Pro-opiomelanocortin | POMC | Human |
| anti_9213_24 | Formimidoyltransferase-cyclodeaminase | FTCD | Human |
| anti_9215_117 | Ubiquitin carboxyl-terminal hydrolase 25 | USP25 | Human |
| anti_9218_7 | Tumor necrosis factor receptor superfamily member 6 | FAS | Human |
| anti_9219_70 | Neudesin | NENF | Human |
| anti_9220_7 | Ephrin type-B receptor 3 | EPHB3 | Human |
| anti_9221_6 | Inositol 1,4,5-trisphosphate receptor-interacting protein-like 1 | ITPRIPL1 | Human |
| anti_9224_20 | Prolactin receptor | PRLR | Human |
| anti_9226_6 | Transmembrane and ubiquitin-like domain-containing protein 2 | TMUB2 | Human |
| anti_9227_15 | Programmed cell death protein 1 | PDCD1 | Human |
| anti_9233_71 | Tissue factor pathway inhibitor 2 | TFPI2 | Human |
| anti_9237_54 | Lysosomal acid phosphatase | ACP2 | Human |
| anti_9241_40 | Signal-regulatory protein gamma | SIRPG | Human |
| anti_9242_11 | Betacellulin | BTC | Human |
| anti_9245_1 | Killer cell immunoglobulin-like receptor 2DL4 | KIR2DL4 | Human |
| anti_9246_1 | Fc_MOUSE | Igh | Mouse |
| anti_9248_36 | Myeloid-derived growth factor | MYDGF | Human |
| anti_9249_17 | Transmembrane protein 9 | TMEM9 | Human |
| anti_9254_18 | High affinity immunoglobulin epsilon receptor subunit alpha | FCER1A | Human |
| anti_9255_5 | Interleukin-17C | IL17C | Human |
| anti_9257_14 | Translational activator of cytochrome c oxidase 1 | TACO1 | Human |
| anti_9258_14 | Fc_MOUSE | Igh | Mouse |
| anti_9259_35 | Tumor necrosis factor receptor superfamily member 21 | TNFRSF21 | Human |
| anti_9261_14 | Ephrin type-B receptor 6 | EPHB6 | Human |
| anti_9263_57 | Sialate O-acetylesterase | SIAE | Human |
| anti_9264_11 | Cathepsin O | CTSO | Human |
| anti_9265_10 | Glioma pathogenesis-related protein 1 | GLIPR1 | Human |
| anti_9266_1 | Triggering receptor expressed on myeloid cells 1 | TREM1 | Human |
| anti_9277_16 | Regenerating islet-derived protein 3-alpha | REG3A | Human |
| anti_9279_7 | Immunoglobulin superfamily member 11 | IGSF11 | Human |
| anti_9281_50 | Fc_MOUSE | Igh | Mouse |

| SOMAmer | Target Full Name | Entrez Gene Symbol | Organism |
|---|---|---|---|
| anti_9283_8 | CD44 antigen | CD44 | Human |
| anti_9287_6 | Sepiapterin reductase | SPR | Human |
| anti_9288_7 | Peptidyl-prolyl cis-trans isomerase FKBP7 | FKBP7 | Human |
| anti_9290_8 | Regulator of microtubule dynamics protein 3 | RMDN3 | Human |
| anti_9297_12 | UDP-GlcNAc:betaGal beta-1,3-N-acetylglucosaminyltransferase 8 | B3GNT8 | Human |
| anti_9300_13 | Nectin-1, isoform gamma | NECTIN1 | Human |
| anti_9302_90 | Growth arrest and DNA damage-inducible proteins-interacting protein 1 | GADD45GIP1 | Human |
| anti_9303_9 | ICOS ligand | ICOSLG | Human |
| anti_9305_89 | Interferon gamma receptor 2 | IFNGR2 | Human |
| anti_9306_7 | Beta-defensin 118 | DEFB118 | Human |
| anti_9307_3 | Ly6/PLAUR domain-containing protein 3 | LYPD3 | Human |
| anti_9310_2 | T-cell surface glycoprotein CD8 beta chain | CD8B | Human |
| anti_9314_9 | Pregnancy-specific beta-1-glycoprotein 5 | PSG5 | Human |
| anti_9316_67 | WAP four-disulfide core domain protein 1 | WFDC1 | Human |
| anti_9319_59 | Transmembrane emp24 domain-containing protein 4 | TMED4 | Human |
| anti_9321_400 | Neuromedin-B | NMB | Human |
| anti_9327_3 | Ly6/PLAUR domain-containing protein 1 | LYPD1 | Human |
| anti_9328_55 | MHC class I polypeptide-related sequence B | MICB | Human |
| anti_9329_28 | Trem-like transcript 1 protein | TREML1 | Human |
| anti_9332_6 | Beta-defensin 136 | DEFB136 | Human |
| anti_9333_59 | Endoplasmic reticulum resident protein 27 | ERP27 | Human |
| anti_9335_28 | Pregnancy-specific beta-1-glycoprotein 9 | PSG9 | Human |
| anti_9337_43 | Protachykinin-1 | TAC1 | Human |
| anti_9340_17 | Peptidyl-prolyl cis-trans isomerase FKBP14 | FKBP14 | Human |
| anti_9343_16 | Interleukin-2 receptor subunit beta | IL2RB | Human |
| anti_9345_436 | WAP four-disulfide core domain protein 13 | WFDC13 | Human |
| anti_9347_13 | Fc_MOUSE | Igh | Mouse |
| anti_9350_3 | Follistatin-related protein 4 | FSTL4 | Human |
| anti_9352_86 | Regenerating islet-derived protein 3-gamma | REG3G | Human |
| anti_9353_8 | Fc_MOUSE | Igh | Mouse |
| anti_9355_26 | Fc_MOUSE | Igh | Mouse |
| anti_9356_20 | Tumor necrosis factor receptor superfamily member 5 | CD40 | Human |
| anti_9358_3 | Fc_MOUSE | Igh | Mouse |
| anti_9359_9 | Protein delta homolog 2 | DLK2 | Human |
| anti_9360_33 | EGF-like repeat and discoidin I-like domain-containing protein 3 | EDIL3 | Human |
| anti_9361_7 | Protocadherin alpha-C2 | PCDHAC2 | Human |
| anti_9363_11 | Cancer/testis antigen 55 | CT55 | Human |
| anti_9365_91 | Protein eyes shut homolog | EYS | Human |
| anti_9366_54 | Interleukin-21 receptor | IL21R | Human |
| anti_9368_64 | Leucine-rich repeat and transmembrane domain-containing protein 1 | LRTM1 | Human |
| anti_9369_174 | Leucine-rich repeat-containing protein 4C | LRRC4C | Human |
| anti_9372_157 | Myelin protein zero-like protein 2 | MPZL2 | Human |
| anti_9373_405 | Fc_MOUSE | Igh | Mouse |
| anti_9374_24 | Neuronatin | NNAT | Human |
| anti_9378_6 | Fibrinogen C domain-containing protein 1 | FIBCD1 | Human |
| anti_9379_248 | Small integral membrane protein 10 | SMIM10 | Human |
| anti_9382_110 | Cell surface glycoprotein CD200 receptor 1 | CD200R1 | Human |
| anti_9383_24 | Chitinase-3-like protein 2 | CHI3L2 | Human |
| anti_9386_42 | Tetratricopeptide repeat protein 9B | TTC9B | Human |
| anti_9387_13 | Fc_MOUSE | Igh | Mouse |
| anti_9389_12 | Fc_MOUSE | Igh | Mouse |
| anti_9398_30 | Galanin-like peptide | GALP | Human |
| anti_9402_18 | Discoidin, CUB and LCCL domain-containing protein 1 | DCBLD1 | Human |
| anti_9412_52 | Activin receptor type-2B | ACVR2B | Human |
| anti_9426_73 | DNA-binding protein inhibitor ID-2 | ID2 | Human |
| anti_9436_2 | DNA-binding protein inhibitor ID-1 | ID1 | Human |
| anti_9439_454 | Fc_MOUSE | Igh | Mouse |
| anti_9443_137 | Cathepsin K | CTSK | Human |

| SOMAmer | Target Full Name | Entrez Gene Symbol | Organism |
|---|---|---|---|
| anti_9444_70 | Protein DGCR6 | DGCR6 | Human |
| anti_9445_44 | Uncharacterized protein C19orf18 | C19orf18 | Human |
| anti_9450_18 | Cyclin-dependent kinase 2-associated protein 1 | CDK2AP1 | Human |
| anti_9453_12 | Vascular endothelial growth factor B | VEGFB | Human |
| anti_9457_3 | Caveolin-2 | CAV2 | Human |
| anti_9463_26 | Fc_MOUSE | Igh | Mouse |
| anti_9466_43 | Protein G6b | MPIG6B | Human |
| anti_9467_24 | Fc_MOUSE | Igh | Mouse |
| anti_9470_15 | Methyltransferase-like protein 24 | METTL24 | Human |
| anti_9474_22 | Asialoglycoprotein receptor 2 | ASGR2 | Human |
| anti_9475_22 | Epididymal secretory glutathione peroxidase | GPX5 | Human |
| anti_9478_69 | Phosphoribosyl pyrophosphate synthase-associated protein 1 | PRPSAP1 | Human |
| anti_9486_13 | Beta-defensin 125 | DEFB125 | Human |
| anti_9487_60 | Somatoliberin | GHRH | Human |
| anti_9491_13 | Fc_MOUSE | Igh | Mouse |
| anti_9492_5 | Fc_MOUSE | Igh | Mouse |
| anti_9493_56 | Fc_MOUSE | Igh | Mouse |
| anti_9495_10 | C-X-C motif chemokine 17 | CXCL17 | Human |
| anti_9497_3 | Fc_MOUSE | Igh | Mouse |
| anti_9500_5 | Fc_MOUSE | Igh | Mouse |
| anti_9503_1 | Neuromedin-S | NMS | Human |
| anti_9504_19 | Ras-related protein Rab-27A | RAB27A | Human |
| anti_9507_55 | Fibroblast growth factor-binding protein 1 | FGFBP1 | Human |
| anti_9508_104 | Fc_MOUSE | Igh | Mouse |
| anti_9509_4 | Trimeric intracellular cation channel type B | TMEM38B | Human |
| anti_9512_24 | Fc_MOUSE | Igh | Mouse |
| anti_9513_9 | Fibroblast growth factor 22 | FGF22 | Human |
| anti_9514_46 | EF-hand calcium-binding domain-containing protein 14 | EFCAB14 | Human |
| anti_9518_95 | Phospholipase A1 member A | PLA1A | Human |
| anti_9523_34 | Integral membrane protein 2C | ITM2C | Human |
| anti_9524_46 | Putative spermatogenesis-associated protein 31D4 | SPATA31D4 | Human |
| anti_9530_6 | Fc_MOUSE | Igh | Mouse |
| anti_9531_24 | Leukocyte-specific transcript 1 protein | LST1 | Human |
| anti_9539_25 | Fc_MOUSE | Igh | Mouse |
| anti_9541_15 | Beta-1,3-galactosyltransferase 1 | B3GALT1 | Human |
| anti_9543_131 | Conserved oligomeric Golgi complex subunit 8 | COG8 | Human |
| anti_9544_24 | Uncharacterized protein C7orf69 | C7orf69 | Human |
| anti_9545_156 | Granzyme K | GZMK | Human |
| anti_9547_29 | EMI domain-containing protein 1 | EMID1 | Human |
| anti_9550_153 | Tetratricopeptide repeat protein 17 | TTC17 | Human |
| anti_9561_21 | Versican core protein | VCAN | Human |
| anti_9565_6 | PRKC apoptosis WT1 regulator protein | PAWR | Human |
| anti_9566_103 | Fc_MOUSE | Igh | Mouse |
| anti_9568_289 | High affinity immunoglobulin alpha and immunoglobulin mu Fc receptor | FCAMR | Human |
| anti_9569_14 | Scavenger receptor class A member 3 | SCARA3 | Human |
| anti_9573_108 | Thioredoxin domain-containing protein 11 | TXNDC11 | Human |
| anti_9576_58 | Motor neuron and pancreas homeobox protein 1 | MNX1 | Human |
| anti_9577_26 | Synaptotagmin-2 | SYT2 | Human |
| anti_9578_263 | MANSC domain-containing protein 4 | MANSC4 | Human |
| anti_9579_59 | Transmembrane gamma-carboxyglutamic acid protein 4 | PRRG4 | Human |
| anti_9580_5 | Laminin subunit gamma-2 | LAMC2 | Human |
| anti_9581_4 | Pre-mRNA-processing factor 6 | PRPF6 | Human |
| anti_9582_93 | Sushi domain-containing protein 1 | SUSD1 | Human |
| anti_9583_17 | RING finger protein 24 | RNF24 | Human |
| anti_9585_80 | CXADR-like membrane protein | CLMP | Human |
| anti_9594_30 | Zinc transporter 5 | SLC30A5 | Human |
| anti_9600_55 | Thrombospondin type-1 domain-containing protein 7A | THSD7A | Human |
| anti_9603_9 | Interleukin-22 receptor subunit alpha-1 | IL22RA1 | Human |
| anti_9604_14 | Cadherin-11 | CDH11 | Human |
| anti_9606_4 | Nuclear pore membrane glycoprotein 210-like | NUP210L | Human |

| SOMAmer | Target Full Name | Entrez Gene Symbol | Organism |
|---|---|---|---|
| anti_9607_39 | Proline-rich protein 27 | PRR27 | Human |
| anti_9613_16 | Fc_MOUSE | Igh | Mouse |
| anti_9638_2 | T-cell immunoreceptor with Ig and ITIM domains | TIGIT | Human |
| anti_9715_15 | Immunoglobulin superfamily member 3 | IGSF3 | Human |
| anti_9723_105 | Multimerin-2 | MMRN2 | Human |
| anti_9725_46 | RING finger protein 219_MOUSE | Obi1 | Mouse |
| anti_9728_4 | E3 ubiquitin-protein ligase CCNB1IP1 | CCNB1IP1 | Human |
| anti_9730_22 | GH3 domain-containing protein | GHDC | Human |
| anti_9731_29 | Scavenger receptor class A member 3 | SCARA3 | Human |
| anti_9739_4 | DNA mismatch repair protein Msh2 | MSH2 | Human |
| anti_9747_48 | MAP/microtubule affinity-regulating kinase 3 | MARK3 | Human |
| anti_9748_31 | Glutathione S-transferase Mu 3 | GSTM3 | Human |
| anti_9750_7 | Protein S100-A4 | S100A4 | Human |
| anti_9751_72 | Nuclease-sensitive element-binding protein 1 | YBX1 | Human |
| anti_9754_33 | Ribosyldihydronicotinamide dehydrogenase [quinone] | NQO2 | Human |
| anti_9758_17 | 40S ribosomal protein S4, X isoform | RPS4X | Human |
| anti_9759_13 | Indoleamine 2,3-dioxygenase 1 | IDO1 | Human |
| anti_9760_13 | Mitogen-activated protein kinase 9 | MAPK9 | Human |
| anti_9765_4 | Nuclear distribution protein nudE homolog 1 | NDE1 | Human |
| anti_9767_22 | Protein ripply1 | RIPPLY1 | Human |
| anti_9768_5 | Interleukin-10 receptor subunit alpha | IL10RA | Human |
| anti_9769_48 | Delta and Notch-like epidermal growth factor-related receptor | DNER | Human |
| anti_9772_153 | Neuroligin-2 | NLGN2 | Human |
| anti_9773_15 | E3 ubiquitin-protein ligase RNF149 | RNF149 | Human |
| anti_9774_59 | RING finger protein 150 | RNF150 | Human |
| anti_9779_63 | Homocysteine-responsive endoplasmic reticulum-resident ubiquitin-like domain member 2 protein | HERPUD2 | Human |
| anti_9783_75 | DnaJ homolog subfamily C member 11 | DNAJC11 | Human |
| anti_9786_310 | Radiation-inducible immediate-early gene IEX-1 | IER3 | Human |
| anti_9787_23 | Collectin-12 | COLEC12 | Human |
| anti_9790_28 | Serine/threonine-protein kinase BRSK2 | BRSK2 | Human |
| anti_9794_17 | AarF domain-containing protein kinase 4 | COQ8B | Human |
| anti_9795_9 | Ceroid-lipofuscinosis neuronal protein 5 | CLN5 | Human |
| anti_9800_20 | NADH dehydrogenase [ubiquinone] 1 beta subcomplex subunit 8, mitochondrial | NDUFB8 | Human |
| anti_9802_27 | Patatin-like phospholipase domain-containing protein 2 | PNPLA2 | Human |
| anti_9804_11 | Fc_MOUSE | Igh | Mouse |
| anti_9805_51 | Semaphorin-4B | SEMA4B | Human |
| anti_9808_41 | Trinucleotide repeat-containing gene 6B protein | TNRC6B | Human |
| anti_9815_5 | Rho GTPase-activating protein 1 | ARHGAP1 | Human |
| anti_9819_110 | Induced myeloid leukemia cell differentiation protein Mcl-1 | MCL1 | Human |
| anti_9826_135 | Bis(5'-adenosyl)-triphosphatase | FHIT | Human |
| anti_9828_86 | L-lactate dehydrogenase C chain | LDHC | Human |
| anti_9829_91 | Bile salt sulfotransferase | SULT2A1 | Human |
| anti_9831_12 | Ras GTPase-activating protein-binding protein 2 | G3BP2 | Human |
| anti_9832_33 | Homogentisate 1,2-dioxygenase | HGD | Human |
| anti_9834_62 | Alcohol dehydrogenase 1B | ADH1B | Human |
| anti_9839_148 | Toll/interleukin-1 receptor domain-containing adapter protein | TIRAP | Human |
| anti_9841_197 | Multifunctional protein ADE2 | PAICS | Human |
| anti_9842_2 | Catenin beta-1 | CTNNB1 | Human |
| anti_9844_138 | Alpha-actinin-2 | ACTN2 | Human |
| anti_9845_33 | Protein DJ-1 | PARK7 | Human |
| anti_9847_21 | Homeobox protein TGIF2 | TGIF2 | Human |
| anti_9848_22 | Cyclin-H | CCNH | Human |
| anti_9850_38 | Eukaryotic translation initiation factor 1A, X-chromosomal | EIF1AX | Human |
| anti_9854_36 | Aldose reductase | AKR1B1 | Human |
| anti_9856_22 | Deoxyhypusine synthase | DHPS | Human |
| anti_9857_38 | Interferon regulatory factor 4 | IRF4 | Human |
| anti_9859_180 | Diamine acetyltransferase 1 | SAT1 | Human |

| SOMAmer | Target Full Name | Entrez Gene Symbol | Organism |
|---|---|---|---|
| anti_9863_1 | Tropomyosin alpha-4 chain | TPM4 | Human |
| anti_9864_38 | Tetratricopeptide repeat protein 1 | TTC1 | Human |
| anti_9865_40 | Ubiquitin-conjugating enzyme E2 B | UBE2B | Human |
| anti_9867_23 | Fructose-1,6-bisphosphatase isozyme 2 | FBP2 | Human |
| anti_9872_23 | Catenin alpha-2 | CTNNA2 | Human |
| anti_9873_17 | Steroid hormone receptor ERR1 | ESRRA | Human |
| anti_9875_107 | THO complex subunit 1 | THOC1 | Human |
| anti_9884_8 | Peptidyl-prolyl cis-trans isomerase-like 1 | PPIL1 | Human |
| anti_9885_41 | Ras-related protein Rap-2a | RAP2A | Human |
| anti_9886_28 | DNA repair protein XRCC4 | XRCC4 | Human |
| anti_9889_42 | Actin filament-associated protein 1-like 1 | AFAP1L1 | Human |
| anti_9890_8 | Tropomyosin alpha-3 chain | TPM3 | Human |
| anti_9893_27 | Ornithine decarboxylase antizyme 1 | OAZ1 | Human |
| anti_9894_13 | Unconventional myosin-VI | MYO6 | Human |
| anti_9895_77 | DNA repair endonuclease XPF | ERCC4 | Human |
| anti_9896_21 | Forkhead box protein G1 | FOXG1 | Human |
| anti_9897_9 | DNA replication licensing factor MCM6 | MCM6 | Human |
| anti_9898_161 | AT-rich interactive domain-containing protein 1A | ARID1A | Human |
| anti_9899_28 | Kinesin-like protein KIF1C | KIF1C | Human |
| anti_9900_36 | Neurofilament heavy polypeptide | NEFH | Human |
| anti_9901_28 | Egl nine homolog 1 | EGLN1 | Human |
| anti_9905_8 | Fc_MOUSE | Igh | Mouse |
| anti_9907_216 | Tankyrase-1 | TNKS | Human |
| anti_9909_4 | Fc_MOUSE | Igh | Mouse |
| anti_9913_4 | Ribosome-binding protein 1 | RRBP1 | Human |
| anti_9921_14 | F-box/LRR-repeat protein 4 | FBXL4 | Human |
| anti_9923_6 | Son of sevenless homolog 1 | SOS 1 | Human |
| anti_9925_56 | Scavenger receptor class F member 2 | SCARF2 | Human |
| anti_9929_16 | 3-keto-steroid reductase | HSD17B7 | Human |
| anti_9930_48 | Gamma-aminobutyric acid type B receptor subunit 2 | GABBR2 | Human |
| anti_9932_49 | Semaphorin-4F | SEMA4F | Human |
| anti_9933_49 | Protein ATP1B4 | ATP1B4 | Human |
| anti_9936_27 | E3 ubiquitin-protein ligase LNX | LNX1 | Human |
| anti_9937_7 | Gap junction alpha-1 protein | GJA1 | Human |
| anti_9940_35 | Dual specificity phosphatase 28 | DUSP28 | Human |
| anti_9941_70 | Protocadherin beta-1 | PCDHB1 | Human |
| anti_9942_2 | NTF2-related export protein 1 | NXT1 | Human |
| anti_9947_22 | DNA topoisomerase 2-binding protein 1 | TOPBP1 | Human |
| anti_9951_36 | F-box/LRR-repeat protein 4 | FBXL4 | Human |
| anti_9952_57 | Transcription factor A, mitochondrial | TFAM | Human |
| anti_9954_2 | Nuclear pore complex-interacting protein family member B3 | NPIPB3 | Human |
| anti_9955_40 | RING finger protein 148 | RNF148 | Human |
| anti_9956_7 | Disintegrin and metalloproteinase domain-containing protein 15 | ADAM15 | Human |
| anti_9957_9 | E3 ubiquitin-protein ligase RNF114 | RNF114 | Human |
| anti_9960_2 | TBC1 domain family member 5 | TBC1D5 | Human |
| anti_9974_8 | Delta-like protein 3 | DLL3 | Human |
| anti_9981_18 | Protein FAM234B | FAM234B | Human |
| anti_9983_97 | Inactive serine protease 35 | PRSS35 | Human |
| anti_9984_12 | Protein YIPF6 | YIPF6 | Human |
| anti_9986_14 | Neuropeptide W | NPW | Human |
| anti_9987_30 | Leucine-rich repeat-containing protein 25 | LRRC25 | Human |
| anti_9989_12 | Leucine-rich repeat-containing protein 24 | LRRC24 | Human |
| anti_9993_11 | Zinc finger protein 264 | ZNF264 | Human |
| anti_9995_6 | Deoxyuridine 5'-triphosphate nucleotidohydrolase, mitochondrial | DUT | Human |
| anti_9997_12 | UBX domain-containing protein 4 | UBXN4 | Human |
